# Path to drugging functional clones of luminal breast cancers using in-depth proteomics with spatially resolved mass spectrometry guided by MALDI imaging

**DOI:** 10.1101/2021.02.16.21251694

**Authors:** N. Hajjaji, S. Aboulouard, T. Cardon, D. Bertin, YM. Robin, I. Fournier, M. Salzet

## Abstract

Integrating tumor heterogeneity in the drug discovery process is a key challenge to tackle breast cancer resistance. Identifying protein targets for functionally distinct tumor clones is particularly important to tailor therapy to the heterogeneous tumor subpopulations. For this purpose, we performed an unsupervised, label-free, spatially resolved shotgun proteogenomic guided by MALDI mass spectrometry imaging (MSI) on 124 selected tumor clonal areas from early luminal breast cancers, tumor stroma, and breast cancer metastases. 2868 proteins were identified. The main protein classes found in the clonal proteome dataset were enzymes, cytoskeletal proteins, membrane-traffic, translational or scaffold proteins, or transporters. As a comparison, gene-specific transcriptional regulators, chromatin related proteins or transmembrane signal receptor were more abundant in the TCGA dataset. Moreover, 26 mutated proteins have been identified. Similarly, expanding the search to alternative proteins databases retrieved 126 alternative proteins in the clonal proteome dataset. The majority of these alternative proteins were coded mainly from non-coding RNA. To fully understand the molecular information brought by our approach and its relevance to drug target discovery, the clonal proteomic dataset was further compared to the TCGA breast cancer database and two transcriptomic panels, BC360 (nanoString®) and CDx (Foundation One®). We retrieved 139 pathways in the clonal proteome dataset. Only 55% of these pathways were also present in the TCGA dataset, 68% in BC360 and 50% in CDx. Seven of these pathways have been suggested as candidate for drug targeting, 22 have been associated with breast cancer in experimental or clinical reports, the remaining 19 pathways have been understudied in breast cancer. Among the anticancer drugs, 35 drugs matched uniquely with the clonal proteome dataset, with only 7 of them already approved in breast cancer. The number of target and drug interactions with non-anticancer drugs (such as agents targeting the cardiovascular system, metabolism, the musculoskeletal or the nervous systems) was higher in the clonal proteome dataset (540 interactions) compared to TCGA (83 interactions), BC360 (419 interactions), or CDx (172 interactions). Thus, we described the non-redundant knowledge brought by this approach compared to TCGA or transcriptomic panels, the targetable proteins identified in the clonal proteome dataset, and the potential of this approach for drug discovery and repurposing through drug interactions with antineoplastic agents and non-anticancer drugs.

**Significance:** Spatially resolved mass spectrometry guided by MALDI MS imaging is a precision oncology tool to map and profile breast cancer proteomic clones with the aim of integrating tumor heterogeneity in the target discovery process to develop clone-tailored therapeutic strategies in breast cancer.

**Highlights:**

- Spatially resolved mass spectrometry guided by MALDI mass spectrometry imaging allows an in-depth proteomic profiling of breast cancer functional clones.
- This unsupervised and unlabeled technology performed on intact tumors provides a multidimensional analysis of the clonal proteome including conventional proteins, mutated proteins, and alternative proteins.
- The rich clonal proteomic information generated was not redundant with TCGA or transcriptomic panels, and showed pathways exclusively found in the proteomic analysis.
- A large proportion of the proteins in the clonal proteome dataset were druggable with both antineoplastic agents and non-anticancer drugs, showing the potential application to drug repurposing.
- A significant number of the proteins detected had partially or not yet known drug interactions, showing the potential for discovery.

## Introduction

Breast cancer remains the most frequent cancer and the leading cause of cancer-related death among women in Europe (globocan iarc). The rational development of targeted drugs based on molecular knowledge of cancer is a major therapeutic progress that brought substantial hope to improving patients’ outcome. However, the complex biologic features of this disease, especially the existence of multiple heterogeneous tumor subclones (Yates et al. 2015), have prevented its eradication, driven drug resistance, including to targeted therapies (Garraway et al. 2012), and has been identified as a marker of poor prognosis in breast cancer patients (McDonald et al. 2019; Pereira et al. 2016). Integrating tumor heterogeneity in the target discovery process could improve and expand precision oncology strategies in breast cancer. However, this implies the ability to isolate and analyze tumor clones separately to understand their biology, find vulnerabilities and identify potential druggable targets.

Historically, sequencing methods revealed genomic alterations driving the emergence of clonal cancer cell subpopulations (Greaves et al. 2012; Williams et al. 2019). Beside this genomic heterogeneity, non-genetic mechanisms, such as dynamic transcriptional, translational and metabolic adaptations also contribute to tumor heterogeneity and drug resistance or tolerance (Pogrebniak et al. 2018; Marine et al. 2020). Thus, beside the technologies used to detect gene mutations or single nucleotide polymorphisms, technics exploring transcript expression (Gyanchandani et al. 2016), proteins (Hennessy et al. 2010), or metabolites (Kim et al. 2019) also showed significant tumor heterogeneity, demonstrating that heterogeneity is constantly present from the structural to the functional levels of the tumor. Therefore, approaches complementary to genomics are necessary to comprehensively analyze tumor heterogeneity.

Yielding large molecular information on tumor clones from small samples for biomarker or drug target discovery represents a technical challenge despite the advent of single cell technologies (Jackson et al. 2020). Current single-cell sequencing methods require suspensions of cells for isolation, whereas in routine clinical practice the majority of tumors after surgery or biopsy are fixed in formalin and embedded in paraffin blocks. Moreover, analyzing isolated cells does not capture cell-cell interaction in the microenvironment. Spatial transcriptomics represent a powerful tool to access in situ functional information about tumor subpopulations (Stahl et al. 2016), and offers the possibility to be multiplexed to fluorescence in situ hybridization (Xia et al. 2019; Lee et al. 2015). However, some limitations include the poor prediction of protein expression from RNA expression (Liu et al. 2016) or transcriptional errors (Gout et al. 2017) that may hamper drug target inference. Moreover, transcriptome measurements may not necessarily capture adaptive responses that involve post-transcription mechanisms such as translation or metabolic reprogramming (Rapino et al. 2018; Garcia et al. 2019; Jewer et al. 2020). Focusing on tumor proteomic landscape has the advantage of recapitulating both the expressed genomic landscape and the non-genetic processes. This could be of particular interest in tumors with a relatively low mutational burden such as breast cancers (Chalmers et al. 2017). Besides, given that the vast majority of drug targets are proteins (Santos et al. 2017), a proteomic approach allows direct target detection. Technologies specifically dedicated to study the spatial proteomic heterogeneity of tumors, combined or not with transcriptomics are scarce. Most rely on selected and labeled markers, limited in number, for instance with multiplexed pathology methods (Giesen et al. 2014; Schulz et al. 2018; Beechem et al. 2020), which is not suited for discovery.

We asked whether matrix-assisted laser desorption/ionization (MALDI) mass spectrometry imaging (MSI) combined with microproteomics could inform libraries with druggable protein targets from breast cancer clones to expand the drug discovery process. MALDI MSI enables the spatially resolved label-free imaging of different molecular classes, including proteins, in their histological context (Fournier et al. 2008; Lemaire et al. 2006; Lemaire et al. 2007), thus revealing functionally heterogeneous tumor subpopulations in solid tumors (Delcourt et al. 2017; Le Rhun et al. 2017). The selected subclones are further extracted in situ using a semi-automated standardized microproteomic technology to perform a full proteomic profiling with LC-MS/MS (Quanico et al. 2017) comprising identification of referenced proteins but also proteins presenting mutations or alternative proteins issued from the non-coding parts of RNA or non-coding RNA. This approach constitutes a unique tool to characterize the proteomic profile of functionally distinct tumor subpopulations, which we denoted the clonal proteome. Our aims were (i) to map and characterize luminal breast cancers’ functional clones using MALDI MSI combined with microproteomics, and (ii) determine the potential of this approach to identify druggable protein targets in luminal tumors.

## Methods

### Patient samples and consent

We carried out a retrospective single center study at Centre Oscar Lambret (Lille, France) to analyze the spatial heterogeneity of primary breast tumors and breast cancer metastases. Eligible patients were women with early breast cancer or metastatic luminal breast cancer with available FFPE tumor tissue after a surgical procedure or a fine needle biopsy. Our pathologists selected 52 primary tumors and 24 metastases from 51 and 12 patients respectively. All patients still alive gave their informed consent. This retrospective study was approved by the local institutional clinical research committee. The clinico-pathological data of both patient series were listed in **Table 1 (Supplementary Table 1)**.

**Table 1:**
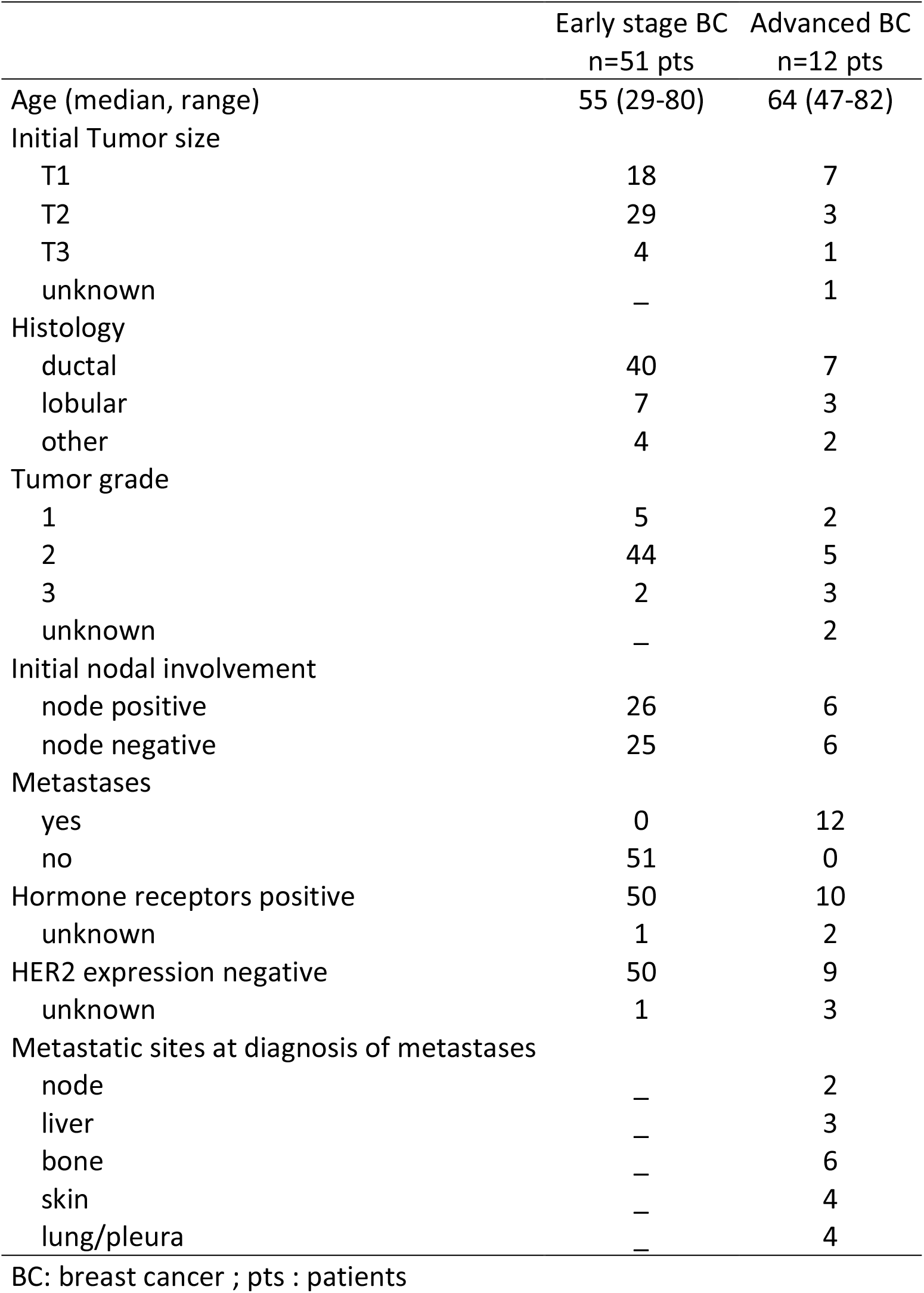
Clinico-pathological parameters for the breast cancer patients’ series

|  | Early stage BC<br>n=51 pts | Advanced BC<br>n=12 pts |
| --- | --- | --- |
| Age (median, range) | 55 (29-80) | 64 (47-82) |
| Initial Tumor size |  |  |
| T1 | 18 | 7 |
| T2 | 29 | 3 |
| T3 | 4 | 1 |
| unknown | – | 1 |
| Histology |  |  |
| ductal | 40 | 7 |
| lobular | 7 | 3 |
| other | 4 | 2 |
| Tumor grade |  |  |
| 1 | 5 | 2 |
| 2 | 44 | 5 |
| 3 | 2 | 3 |
| unknown | – | 2 |
| Initial nodal involvement |  |  |
| node positive | 26 | 6 |
| node negative | 25 | 6 |
| Metastases |  |  |
| yes | 0 | 12 |
| no | 51 | 0 |
| Hormone receptors positive | 50 | 10 |
| unknown | 1 | 2 |
| HER2 expression negative | 50 | 9 |
| unknown | 1 | 3 |
| Metastatic sites at diagnosis of metastases |  |  |
| node | – | 2 |
| liver | – | 3 |
| bone | – | 6 |
| skin | – | 4 |
| lung/pleura | – | 4 |

BC: breast cancer ; pts : patients

### MALDI mass spectrometry imaging (MALDI MSI)

For each tumor sample, 2 consecutive sections of 8 micrometers were cut of the block. The first section was used to perform the MALDI MSI analysis (Fournier et al., 2003; Lemaire et al., 2007; Lemaire et al., 2006a; Lemaire et al., 2006b; Wisztorski et al., 2007). The tumor tissue section was deposited on ITO-coated glass slides (LaserBio Labs, Valbonne, France) and vacuum-dried during 15 min. Protein demasking was performed with washing with NH4HCO3 10mM for 5 min twice, then TRIS HCl 20mM pH9 for 30 min at 95°C. Tryptic digestion was performed (40μg/mL, dissolved in NH4HCO3 50mM) by micro-spraying trypsin on the section surface using an HTX TM sprayer (HTX technologies, LLC), and incubation overnight at 56°C. The slide was dried in a dessicator prior to deposition of a solid ionic matrix HCCA-aniline using an HTX TM sprayer (HTX technologies, LLC) (Lemaire et al., 2006). Briefly, 36 μL of aniline were added to 5 mL of a solution of 10 mg/mL HCCA dissolved in ACN/0.1% TFA aqueous (7:3, v/v). A real-time control of the deposition was performed by monitoring scattered light to obtain a uniform layer of matrix. The MALDI mass spectrometry images were performed on a RapifleX Tissuetyper MALDI TOF/TOF instrument (Bruker Daltonics, Germany) equipped with a smartbeam 3D laser. The MSI mass spectra were acquired in the positive delayed extraction reflectron mode using the 500-3000 m/z range, and averaged from 200 laser shots per pixel, using a 70μm spatial resolution raster.

### MALDI MSI data processing and analysis

The MALDI-MSI data were analyzed using SCiLS Lab software (SCiLS Lab 2019, SCiLS GmbH). Common processing methods for MALDI MSI were applied with a baseline removal using a convolution method and data were normalized using Total Ion Count (TIC) method (Klein et al., 2014; Trede et al., 2012). Then, the resulting pre-processing data were clustered to obtain a spatial segmentation using the bisecting k means algorithm (Alexandrov et al., 2010). Different spatial segmentations were performed. First, an individual segmentation was applied to each tissue separately. Then, the data from all tissues were clustered together to obtain a global segmentation. Briefly, the spatial segmentation consists of grouping all spectra according to their similarity using a clustering algorithm that apply a color code to all pixels of a same cluster. Colors are arbitrarily assigned to clusters; several disconnected regions can have the same color if they share the same molecular content. To limit the pixel-to-pixel variability, edge-preserving image denoising was applied. The segmentation results were represented on a dendrogram resulting from a hierarchical clustering. The branches of the dendrogram were defined based on a distance calculation between each cluster. The manual selection of different branches of the dendrogram allows further segmentation of selected clusters to visualize more regions with distinct molecular composition. Each color-coded region identified a proteomic tumor clone. The regions/clones of interest were then subjected to on-tissue microproteomics, i.e. microdigestion and microextraction, to perform nanoLC-MS & MS/MS analysis of the extract for in-depth protein identification.

### Microproteomic analysis

Superimposing the molecular image with the immunochemistry image allowed selection of the subclonal areas to be submitted to microproteomics using the second consecutive 8 μm tumor sections. The tissue sections were deposited on polylysine glass slides, and microdigested with a trypsin solution deposited with a microspotter. On-tissue trypsin digestion was performed using a Chemical Inkjet Printer (CHIP-100, Shimadzu, Kyoto, Japan). The trypsin solution (40µg/mL, 50mM NH4HCO3 buffer) was deposited on a region defined to 1mm^2^ for 2h. During this time, the trypsin was changed every half-hour. With 350 cycles and 450pl per spot, a total of 6.3µg was deposited. After microdigestion, the spot content was micropextracted by liquid microjunction using the TriVersa Nanomate (Advion Biosciences Inc., Ithaca, NY, USA) using Liquid Extraction and Surface Analysis (LESA) settings. With 3 different solvent mixtures composed of 0.1% TFA, ACN/0.1% TFA (8:2, v/v), and MeOH/0.1% TFA (7:3, v/v). A complete LESA sequence run 2 cycles for each mixture composed of an aspiration (2µL), a mixing onto the tissue, and a dispensing into low-binding tubes. For each tumor area of interest, 2 microextraction sequences were run and pooled (Quanico et al., 2013).

### NanoLC-MS & MS/MS analysis

After liquid extraction, samples were freeze-dried in a SpeedVac concentrator (SPD131DPA, ThermoScientific, Waltham, Massachusetts, USA), reconstituted with 10µL 0.1% TFA and subjected to solid-phase extraction to remove salts and concentrate the peptides. This was done using a C-18 Ziptip (Millipore, Saint-Quentin-en-Yvelines, France), eluted with ACN/0.1% TFA (8:2, v/v) and then the samples were dried for storage. Before analysis, samples were suspended in 20µL ACN/0.1% FA (2:98, v/v), deposited in vials and 10µL were injected for analysis. The separation prior to the MS used online reversed-phase chromatography coupled with a Proxeon Easy-nLC-1000 system (Thermo Scientific) equipped with an Acclaim PepMap trap column (75 μm ID x 2 cm, Thermo Scientific) and C18 packed tip Acclaim PepMap RSLC column (75 μm ID x 50 cm, Thermo Scientific). Peptides were separated using an increasing amount of acetonitrile (5%-40% over 140 minutes) and a flow rate of 300 nL/min. The LC eluent was electrosprayed directly from the analytical column and a voltage of 2 kV was applied via the liquid junction of the nanospray source. The chromatography system was coupled to a Thermo Scientific Q-Exactive mass spectrometer. The mass spectrometer was programmed to acquire in a data-dependent mode. The survey scans were acquired in the Orbitrap mass analyzer operated at 70,000 (FWHM) resolving power. A mass range of 200 to 2000 m/z and a target of 3E6 ions were used for the survey scans. Precursors observed with an intensity over 500 counts were selected “on the fly” for ion trap collision-induced dissociation (CID) fragmentation with an isolation window of 4 amu and a normalized collision energy of 30%. A target of 5000 ions and a maximum injection time of 120 ms were used for CID MS2 spectra. The method was set to analyze the top 10 most intense ions from the survey scan and a dynamic exclusion was enabled for 20 s. Extracts were sequenced randomly to avoid batch effect.

### Data analysis

All MS data were processed with MaxQuant (Cox and Mann, 2008; Tyanova et al., 2015) (Version 1.5.6.5) using the Andromeda (Cox et al., 2011) search engine. The proteins were identified by searching MS and MS/MS data against the Decoy version of the complete proteome for Homo sapiens in the UniProt database (Release March 2017, 70941 entries) combined with 262 commonly detected contaminants. Trypsin specificity was used for digestion mode, with N-terminal acetylation and methionine oxidation selected as a variable. We allowed up to two missed cleavages. Initial mass accuracy of 6 ppm was selected for MS spectra, and the MS/MS tolerance was set to 20 ppm for the HCD data. False discovery rate (FDR) at the peptide spectrum matches (PSM) and protein level was set to 1%. Relative, label-free quantification of the proteins was conducted into MaxQuant using the MaxLFQ algorithm (Cox et al., 2014) with default parameters. Analysis of the identified proteins was performed using Perseus software (http://www.perseus-framework.org/) (version 1.6.12.0). The file containing the information from the identification was filtered to remove hits from the reverse database, proteins with only modified peptides and potential contaminants. The LFQ intensity was logarithmized (log2[x]). Categorical annotation of the rows was used to define the different groups. Principal component analysis (PCA) was done to compare the protein content of each sample. Multiple-sample tests were performed using ANOVA with a p-value of 1%. Normalization was achieved using a Z-score with matrix access by rows. Only proteins that were significant by ANOVA were retained. The hierarchical clustering and profile plots of the statistically significant proteins were performed and visualized with Perseus. Functional annotation and characterization of the identified proteins were performed using FunRich software (version 3) and STRING (version 9.1, http://stringdb.org) (Szklarczyk et al., 2011). Pearson’s correlation coefficient and matrix representation were generated in R software using corrplot package. Gene Set Enrichment Analysis (GSEA) and Cytoscape software (version 3.6.1) were used for the biological process analysis of the clusters selected from the heatmap. The data sets were deposited at the ProteomeXchange Consortium (http://proteomecentral.proteomexchange.org) via the PRIDE partner repository (Vizcaino et al. 2014) with Data are available via ProteomeXchange with identifier PXD024134

### Subnetwork Enrichment Pathway Analyses and statistical Testing

The Elsevier’s Pathway Studio version 10.0 (Ariadne Genomics/Elsevier) was used to deduce relationships among differentially expressed proteomics protein candidates using the Ariadne ResNet database (Bonnet et al. 2009; Yuryev et al. 2009). “Subnetwork Enrichment Analysis” (SNEA) algorithm was selected to extract statistically significant altered biological and functional pathways pertaining to each identified set of protein hits among the different groups. SNEA utilizes Fisher’s statistical test set to determine if there are nonrandom associations between two categorical variables organized by specific relationships. Integrated Venn diagram analysis was performed using “the InteractiVenn”: a web-based tool for the analysis of complex data sets (Heberle et al., 2015). Annotation analysis of gene ontology terms for the identified proteins was performed using PANTHER Classification System (version 15.0, http://www.pantherdb.org) (Mi et al. 2019). Interaction network analyses were performed with Cytoscape (version 3.7.2) and the Cluego application (version 2.5.5) to interpret the lists of genes and proteins by selecting representative Gene Ontology terms and pathways from multiple ontologies and visualize them into functionally organized networks (Bindea et al. 2009). The ontologies used included GO_BiologicalProcess-EBI-UniProt-GOA_27.02.2019,DGO_CellularComponent-EBI-UniProt-GOA_27.02.2019, GO_ImmuneSystemProcess-EBI-UniProt-GOA_27.02.2019, GO_MolecularFunction-EBI-UniProt-GOA_27.02.2019, KEGG_27.02.2019, REACTOME_Pathways_27.02.2019, REACTOME_Reactions_27.02.2019, and WikiPathways_27.02.2019. The GO level range was 3 to 8, and groups with more than 50% overlap were merged. The statistical test used was enrichment/depletion (two-sided hypergeometric test) with a Bonferroni step down correction method.

### Mutated protein identification

Protein identification was also performed using the mutation-specific database (Flores and Lazar, 2020). XMan v2 database contains 2 539 031 mutated peptide sequences from 17 599 Homo sapiens proteins (2 377 103 are missense and 161 928 are nonsense mutations). The interrogation was performed by Proteome Discoverer 2.3 software and Sequest HT package, using an iterative method. The precursor mass tolerance was set to 15 ppm and the fragment mass tolerance was set to 0.02 Da. For high confidence result, the FDR values were specified to 1%. A filter with a minimum Xcorr of 2 was applied. The generated result file was filtered using a Python script to remove unmutated peptides. All mutations were then manually checked based on MS/MS spectra profile.

### Alternative Proteins identification

RAW data obtained by nanoLC-MS/MS analysis were processed using Proteome Discoverer V2.3 (Thermo Scientific) with the following parameters: Trypsin as an enzyme, 2 missed cleavages, methionine oxidation as a variable modification, Precursor Mass Tolerance: 10 ppm and Fragment mass tolerance: 0.6 Da. The validation was performed using Percolator with an FDR set to 0.001%. A consensus workflow was then applied for the statistical arrangement, using the high confidence protein identification. The protein database was uploaded from Openprot (https://openprot.org/) and included reference proteins, novel isoforms, and alternative proteins predicted from both Ensembl and RefSeq annotations (GRCh38.83, GRCh38.p7) (Cardon et al., 2019).

### TCGA, BC360, CDx datasets

To compare our proteomic data with genomic and transcriptomic datasets used in breast cancer, the reference genes of the proteins were contrasted to publically available TCGA, BC360 (nanoString®) and CDx (Foundation One®) datasets. Breast cancer genomic alterations were collected from the TCGA web portal using “breast cancer” as keyword in the search engine. The TCGA gene list is in **Supplementary material 1**. The gene list of the BC360 and CDx panels were obtained from nanoString and Foundation One websites. The gene lists are in **Supplementary material 2 and 3**.

### Druggable genome database

DrugCentral (http://drugcentral.org) is an online drug information resource created and maintained by the Division of Translational Informatics at University of New Mexico in collaboration with the IDG Illuminating the Druggable Genome (IDG) (https://druggablegenome.net/index) (Sheils et al. 2020). DrugCentral provides information on active ingredients, chemical entities, pharmaceutical products, the mode of action of drugs, indications, and pharmacologic action. Data is monitored on FDA, EMA, and PMDA for new drug approval on regular basis. Supported target search terms are HUGO gene symbols, Uniprot accessions and target names, and Swissprot identifiers. We used the WHO anatomical therapeutic chemical (ATC) classification to categorize drugs.

### Druggability level of the targets

The druggability level of the targets was classified using the definition of the Illuminating the Druggable Genome Knowledge Management Center (IDG-KMC) based on four target development levels (TDLs) categorized as follows: (i) Tclin: targets with activities in DrugCentral (ie. approved drugs) and known mechanism of action, (ii) Tchem: targets with activities in ChEMBL or DrugCentral that satisfy the activity thresholds detailed in https://druggablegenome.net/ProteinFam, (iii) Tbio: targets with no known drug or small molecule activities that satisfy the activity thresholds and criteria (detailed in https://druggablegenome.net/ProteinFam), (iv) Tdark: targets with virtually no known drug or small molecule activities that satisfy the criteria defined by IDG-KMC.

### Statistical analyses

Descriptive analyses used frequency of distribution, median, quartiles and extremes. Survival analyses were performed using the breast cancer Kaplan-Meier plotter online tool (https://kmplot.com; n=3955). The database sources included GEO, EGA, and TCGA (Gyorffy et al 2010). A multiple gene testing was run using available cohorts of patients with estrogen receptor positive and HER2 negative disease to analyze the reference genes association with distant metastases free survival (DMFS) and overall survival (OS). A logrank p<0.05 was considered significant.

## Results

### Workflow for breast cancer clonal proteome analysis

The workflow described in **Figure 1A** was applied to two FFPE tumor slides to provide a spatially resolved unsupervised and unlabeled visualization of breast cancer spatial heterogeneity, and an in-depth proteomic profiling. The MALDI MSI on-tissue spatial analysis mapped high molecular weight peptide composition on the first tumor slide. The spectral data obtained were clustered by the bisecting k-means method, which attributed color-coded groups to tumors areas according to the similarity of their proteomic signature. Manual group splitting (group segmentation) was limited to 3 in order to map only main functional differences between tumor subpopulations. Imaging revealed distinct proteomic clones, as illustrated in **Figure 1B** showing representative MALDI MS images of a primary tumor and a metastasis sample among the 76 luminal tumors analyzed (52 primary breast tumors and 24 metastases in (**Supplementary material 4 and 5)**. From the MSI data, 124 MSI clonal areas were retrieved corresponding to 52 areas of primary tumors, 48 areas of primary tumor stroma and 24 areas of metastases. Each of these 124 MSI clones were individually analyzed by spatially resolved shotgun proteomic. MaxLFQ algorithm was used to perform label-free quantification of proteins and resulted in a total of 2868 proteins from the 124 clonal areas (**Figure 2A**). The number of proteins identified did not significantly differ between tissue types **(Supplementary material 6A)**, or according to the sampling method, *i*.*e*. mastectomy, surgical biopsy or fine needle core biopsy **(Supplementary material 6B)**. Panther analysis showed that the main protein classes found in the clonal proteome dataset were enzymes, cytoskeletal proteins, membrane-traffic, translational or scaffold proteins, or transporters (**Figure 2B**). As a comparison, gene-specific transcriptional regulators, chromatin related proteins or transmembrane signal receptor were more abundant in the TCGA dataset (**Figure 2B**). Differences between primary tumors and stroma, or between primary tumors and metastases were mild among the main protein classes (**Supplementary material material 7**). We also detected modified proteins, specifically mutated proteins and alternative proteins. Mutations-missense at a protein-level were identified by expanding the search of the raw mass spectrometry files of our proteomic dataset against a mutated peptide database. The search identified 26 mutated proteins, 18 in primary tumors, 20 in stroma and 12 in metastases, with various frequencies (**Figure 2C**). Similarly, expanding the search to alternative proteins databases retrieved 126 alternative proteins in the clonal proteome dataset (**Figure 2D)**: 79 were identified in primary tumors, 69 in stroma and 50 in metastases. The majority of these alternative proteins had a length ranging from 29 to 150 amino acids (**Figures 2E, 2F, 2G)**. They were coded mainly from non-coding RNA (**Figures 2H, 2I, 2J**). These proteins were infrequent and found mostly in less than 25% of the patients (**Figures 2K, 2L, 2M**).

**Figure 1:**
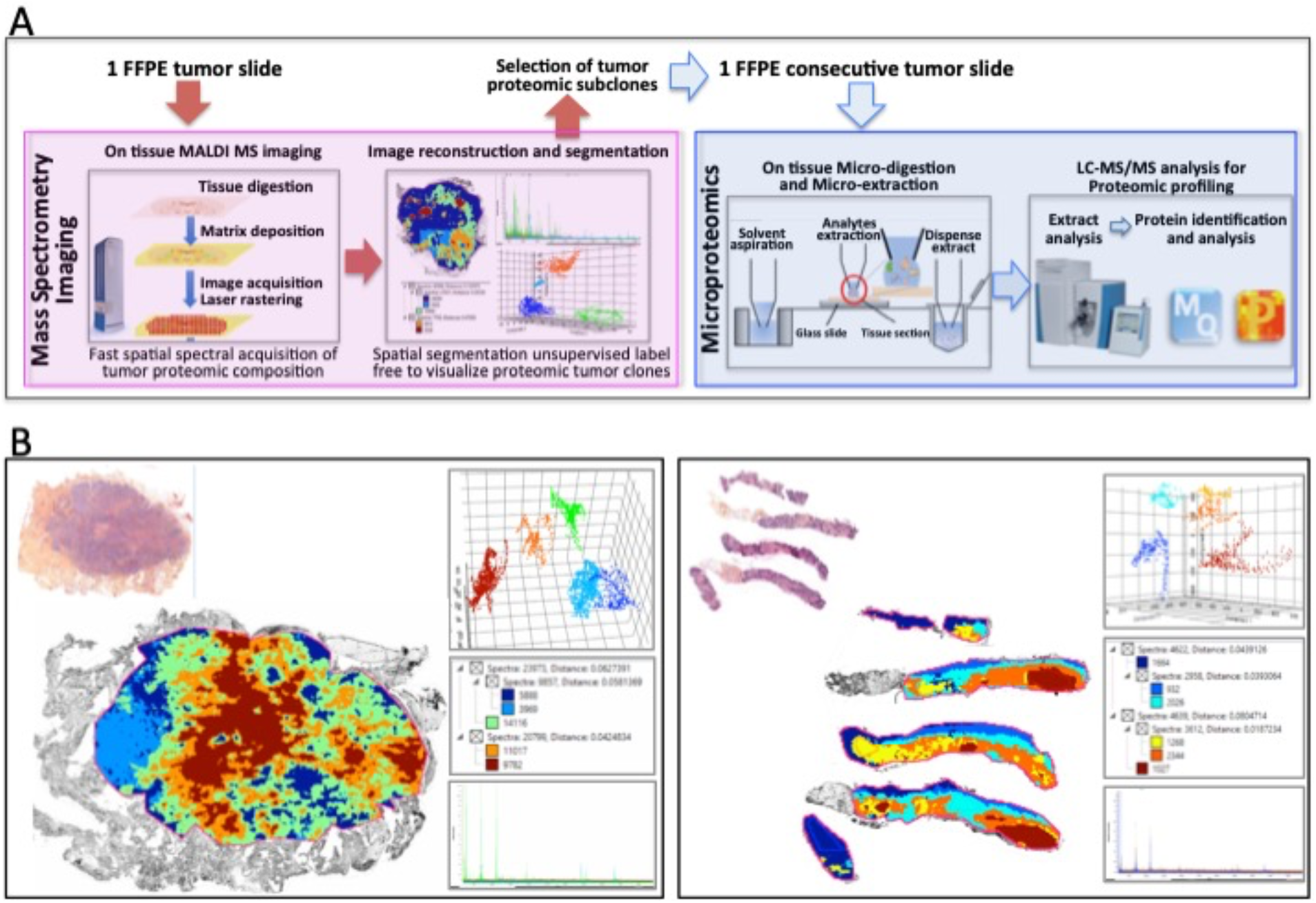
Clonal proteome analysis in breast cancer. (A) Workflow for on tissue analysis of tumor proteomic heterogeneity using spatially resolved microproteomics guided by MALDI MSI (B) The presence of tumor proteomic clones revealed by MSI was illustrated in a primary tumor (left) from a surgical resection (case 42) and a metastatic sample (right) collected with a fine needle biopsy (case 22). In each sample vignette, the MALDI MS imaging is displayed with the histological HPS picture (upper left), the principal component analysis of the proteomic clones (upper right), the segmentation tree (middle right), and the spectra of the clones (bottom right).

**Figure 2:**
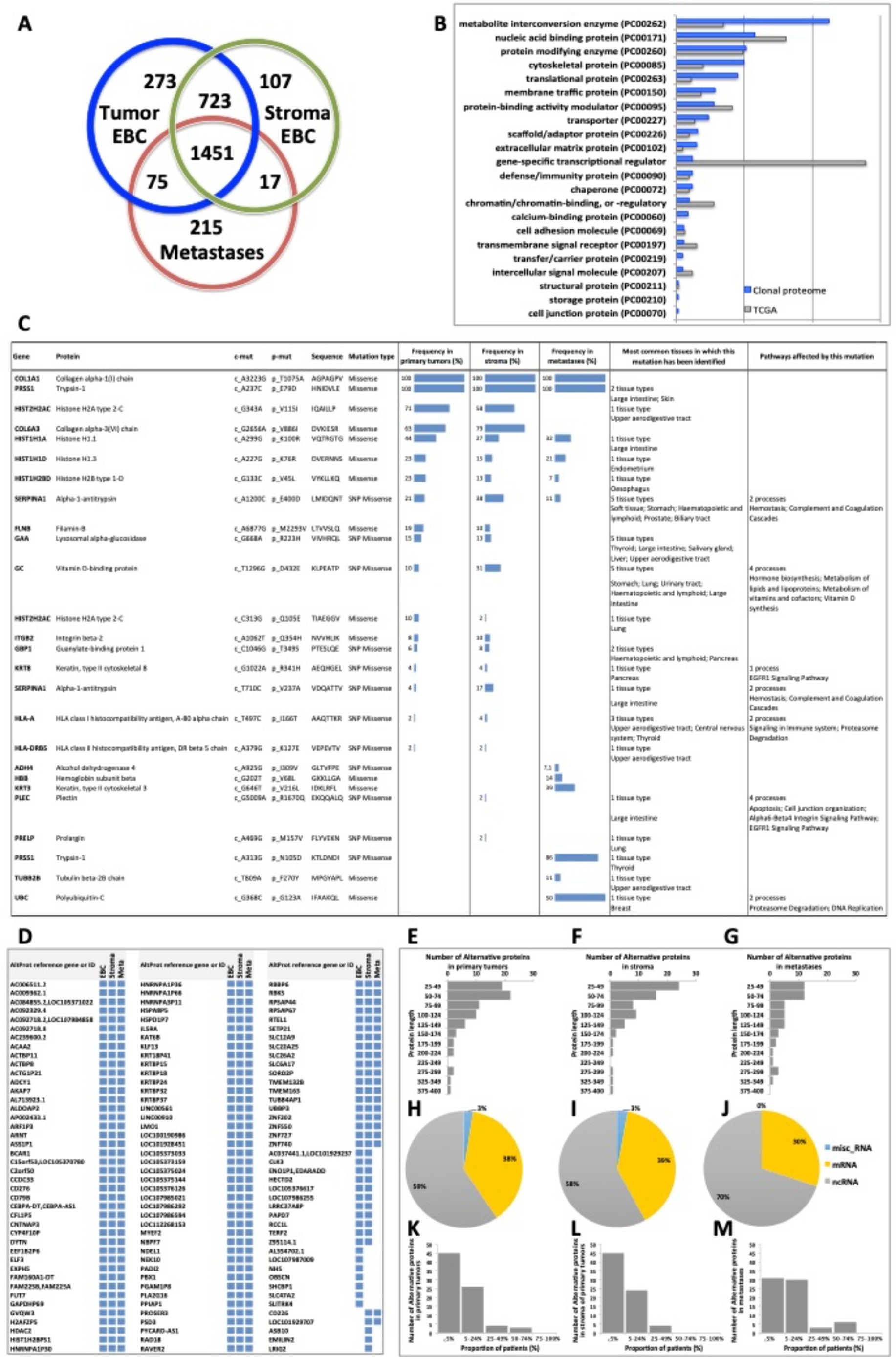
(A) The Venn diagram showing the number of proteins specific or shared among primary tumors (blue), stroma (green) and metastases (red). (B) Distribution of protein classes (in %) in the clonal proteome dataset (in blue) compared with TCGA dataset (in grey). (C) Mutated proteins identified using mass spectrometry, with their frequency in primary tumors, stroma and metastases, and the tissues in which they have been reported, along with the processes they affected. (D) 126 Alternative proteins identified by mass spectrometry; their length is indicated in (E) primary tumor samples, (F) stroma samples and (G) metastases. Their coding RNA distribution is shown in (H) (I) (J), respectively. The frequency of alternative proteins among patients is shown in (K) primary tumors, (L) stroma and (M) metastases. AltProt: alternative proteins; EBC: early breast cancer; ID: identification; Meta: metastases; SNP: single nucleotide polymorphism

### Luminal tumors clonal proteome landscape among classic and modified proteins

To fully understand the molecular information brought by our approach and its relevance to drug target discovery, the clonal proteomic dataset was further compared to the TCGA breast cancer database and two transcriptomic panels, BC360 (nanoString®) and CDx (Foundation One®). The Venn diagram in **Figure 3A** showed that only few proteins of the clonal proteomic dataset (identified by their reference gene) were shared with TCGA, BC360 or CDx panels, both in primary tumors and metastases. 2264 and 1562 proteins were exclusively found in the clonal proteome dataset of primary tumors and metastases respectively (**Supplementary material 8**). Enrichment analysis using Panther software identified 139 pathways in the clonal proteome dataset. Only 55% of these pathways were also present in the TCGA dataset, 68% in BC360 and 50% in CDx. The pathways and processes identified were differentially distributed across the datasets as depicted in the heatmap in **Figure 3B**. Pathways over-represented in the clonal proteome dataset were integrin or inflammation mediated by chemokine and cytokine signaling pathways, cytoskeletal regulation by Rho GTPase, the ubiquitin proteasome pathway, glycolysis,de novo purine biosynthesis or DNA replication. Under-represented processes were mainly signaling pathways or the oxidative stress response. 41 pathways were exclusive to the clonal proteome (**Table 2**), mainly metabolic pathways involved in amino acid, lipid or nucleic acid synthesis. Seven of these pathways have been suggested as candidate for drug targeting, 22 have been associated with breast cancer in experimental or clinical reports, the remaining 19 pathways have been understudied in breast cancer (**Table 2**). The mutated proteins identified have been reported in a variety of human tissues; only mutated polyubiquitin-C has been reported in breast tissue with an impact on proteasome degradation and DNA replication. The other mutated proteins may affect hemostasis, complement and coagulation cascades, hormone biosynthesis, metabolism, EGFR1 signaling, signaling in the immune system, apoptosis, cell junction organization, or integrin signaling (**Figure 2C**). Enrichment analyses performed on the identified mutated proteins using their reference genes showed three main biological processes: expression of interferon gamma genes, apoptosis, and senescence (**Figures 3C and 3D**). Only 4 of the reference genes (COL6A3, COL1A1, HBB, HLA-A) were also found in TCGA or BC360 datasets (none in CDx). The functions of the alternative proteins identified are not known yet. Among their known reference genes, only 8 (ARNT, CD79B, KAT6B, LMO1, PBX1, CD276, ELF3, HDAC2) were also present in TCGA, BC360 or CDx panels.

**Table 2:**
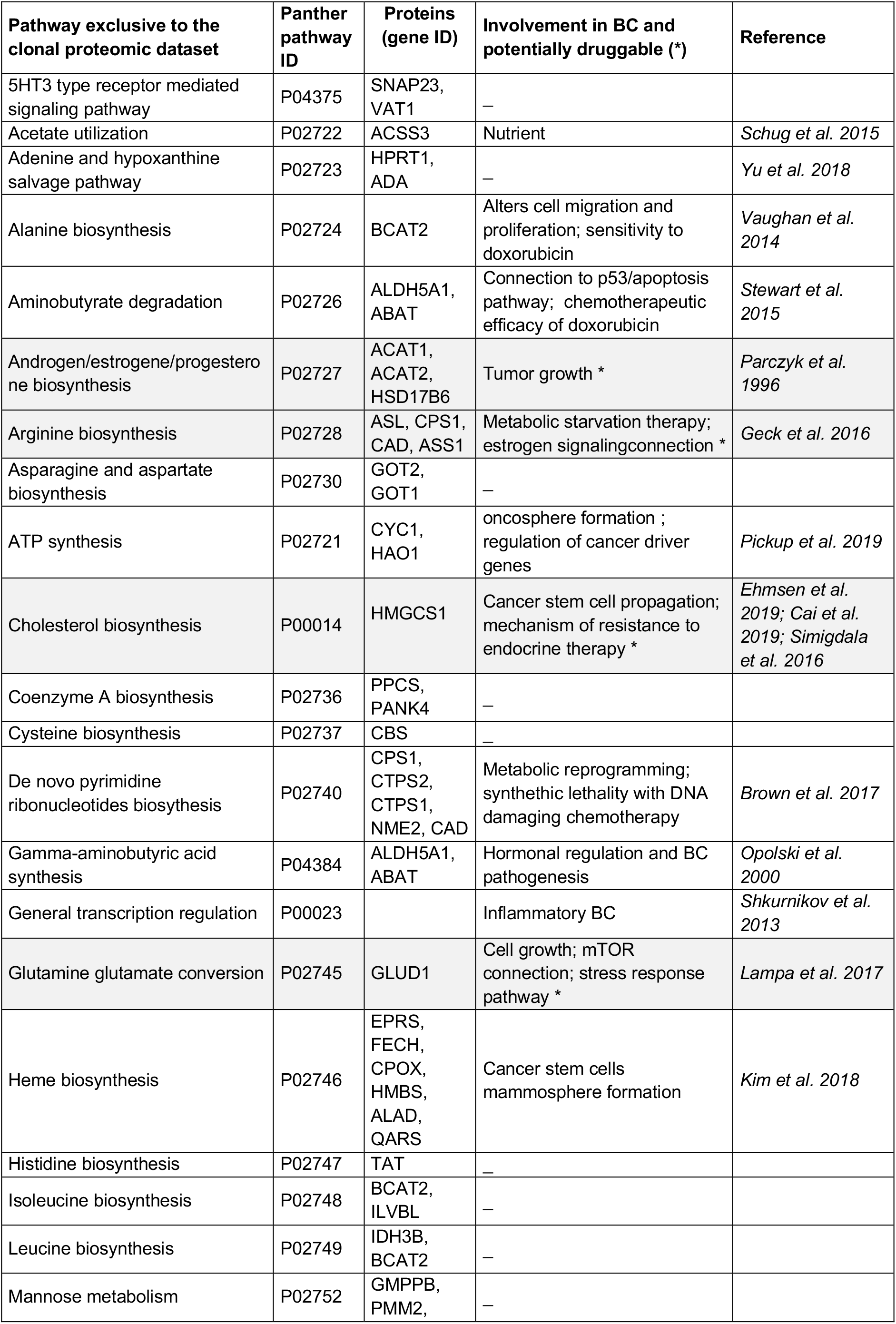
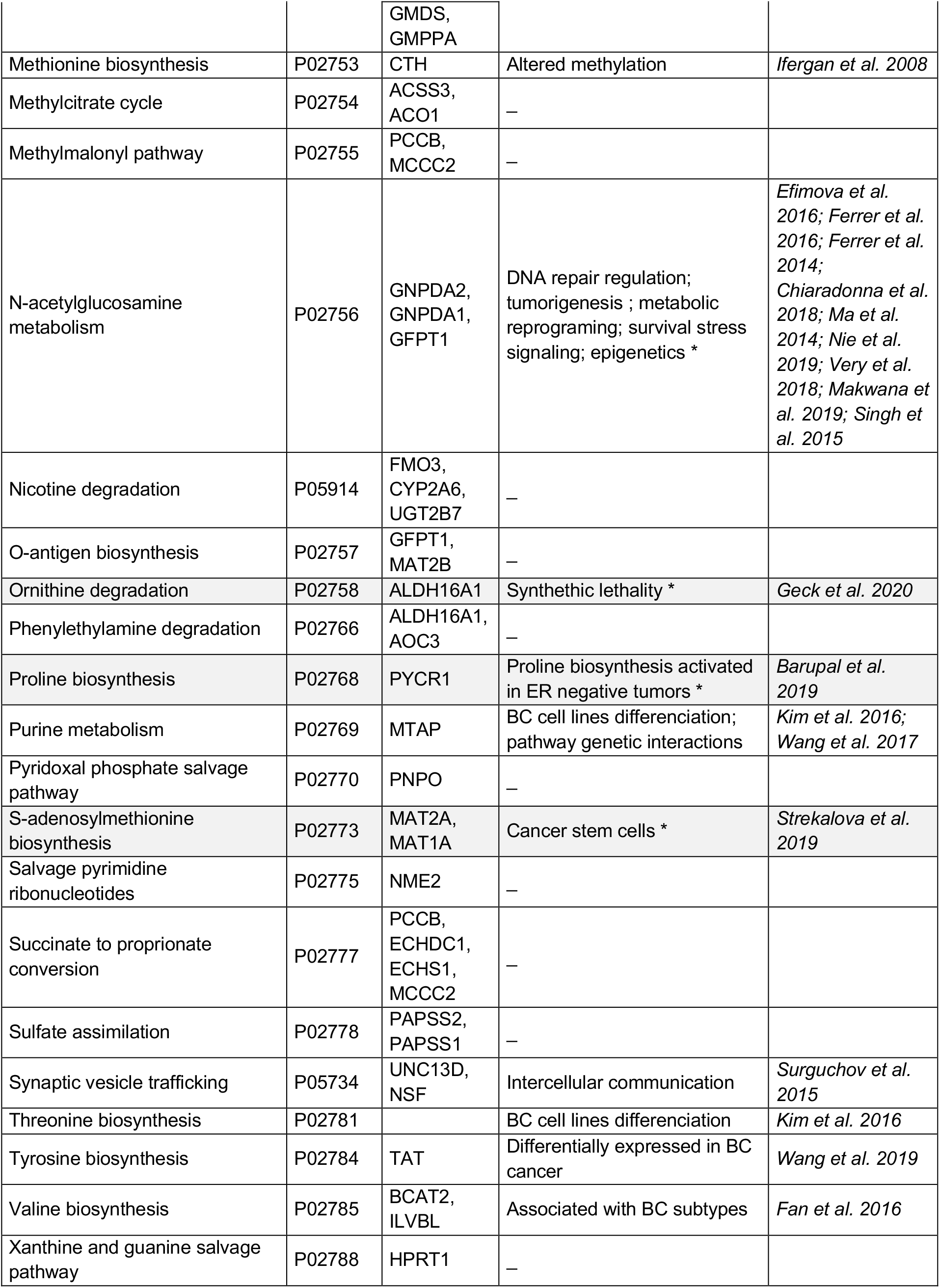
Pathways exclusive to the clonal proteomic dataset

**Figure 3:**
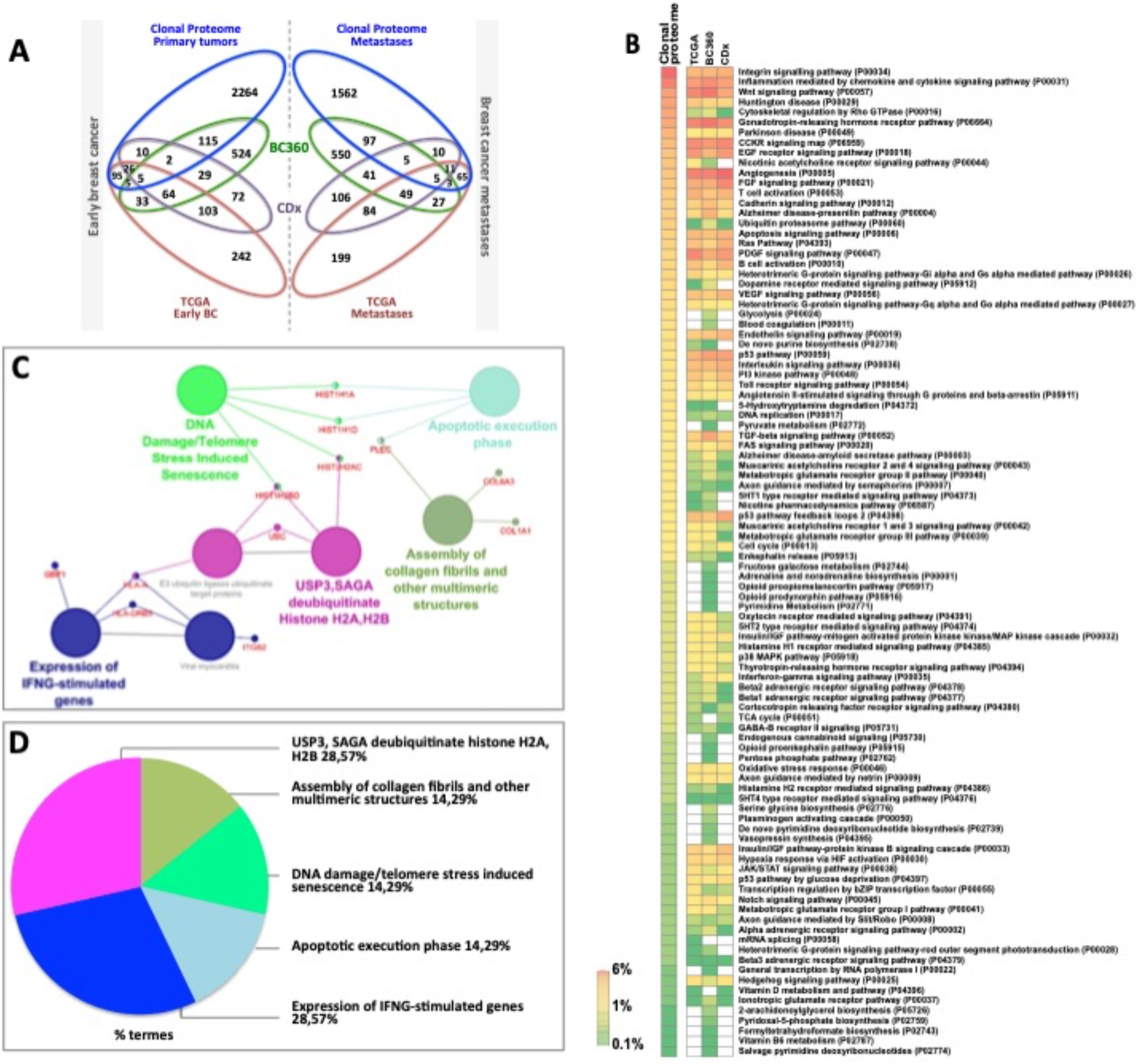
(A) Venn diagram comparing the clonal proteome dataset with TCGA, BC360 and CDx panels, in primary breast cancer (left) or metastases (right). (B) Panther pathways heatmap showing gene distribution between the datasets. Pathways over-represented are colored in orange and those under represented in green. (C) Mutated protein networks and (D) biological processes distribution analyzed using Cytoscape and Cluego.

### Clonal proteome druggability and interactions with approved drugs

The clonal proteome dataset was reviewed against DrugCentral database to determine the number of proteins targetable, their level of druggability and their interaction with approved drugs. Among the proteins identified in the clonal proteome dataset, 1495 proteins were targetable with a level of druggability high for 52% of them (known mechanism of action and drug interaction, Tclin), while 39% had a lower level of knowledge (Tchem), and 9% had no known drug or small molecule interaction (Tbio) (**Figure 4A**). The highest number of druggable targets was observed in the clonal proteome compared to the genomic and transcriptomic datasets. The proportion of less known targets was also greater in the clonal proteome dataset (47%) compared to TCGA (18%), BC360 (23%) or CDx (25%) (**Figure 4A**). The main target classes in the clonal proteome dataset were enzymes (60%), kinases (23%) and transporters (7%), whereas kinases were dominant in the other datasets (46% to 77%) (**Figure 4B**). The number of protein and drug interactions with antineoplastic and immunomodulating agents were up to 309 in the clonal proteome dataset, 485 in TCGA, 506 in BC360, and 647 in CDx (**Figure 4C and Supplementary material 9**). Among the anticancer drugs, 35 drugs matched uniquely with the clonal proteome dataset, with only 7 of them already approved in breast cancer. The number of target and drug interactions with non-anticancer drugs (such as agents targeting the cardiovascular system, metabolism, the musculoskeletal or the nervous systems) was higher in the clonal proteome dataset (540 interactions) compared to TCGA (83 interactions), BC360 (419 interactions), or CDx (172 interactions) (**Figure 4C**).

**Figure 4:**
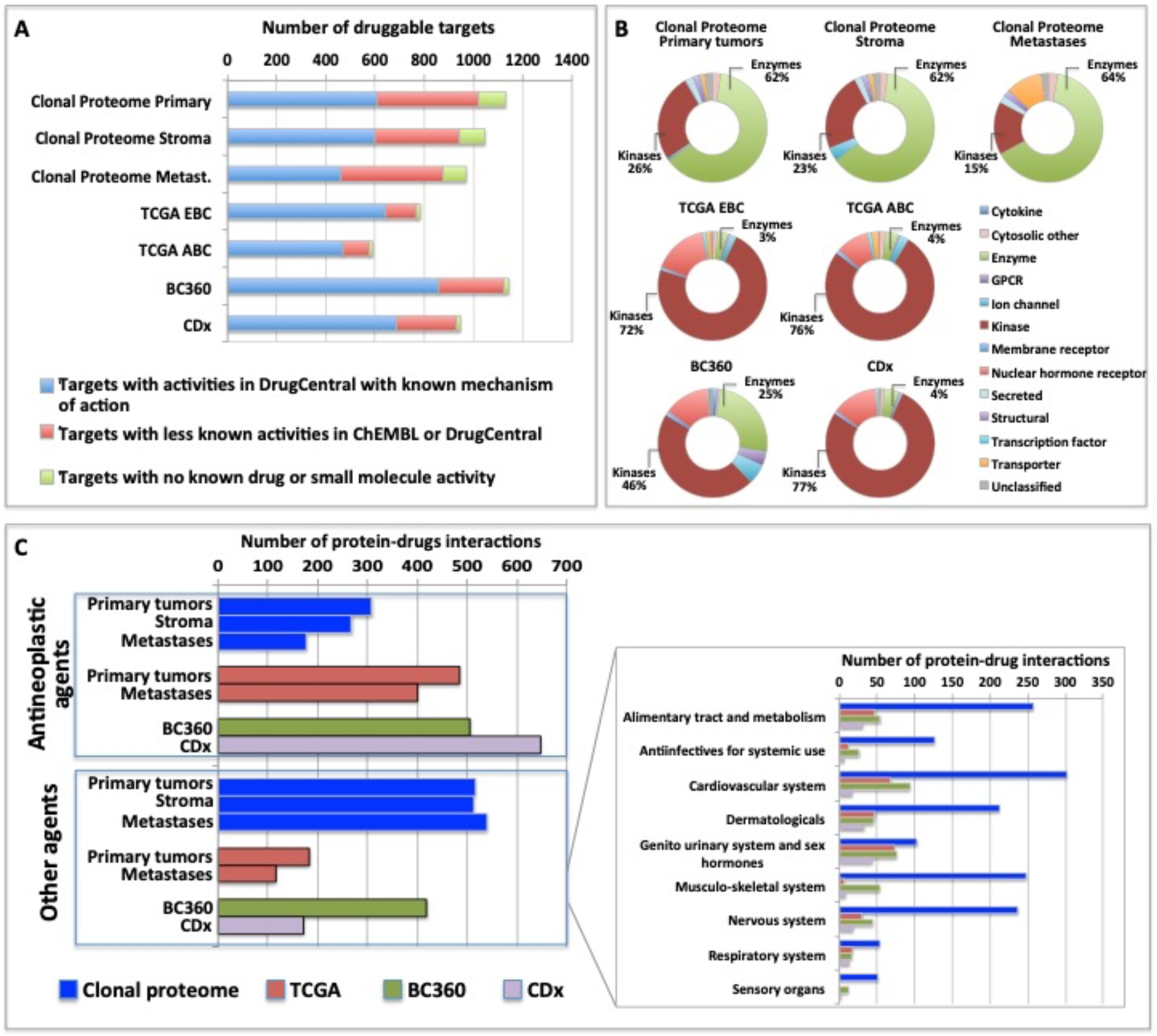
Druggable targets identified (A) in the clonal proteome, TCGA, CDx, and BC360 datasets using DrugCentral, and the druggability level as defined by IDG-KMC (https://druggablegenome.net/ProteinFam). Known targets (Tclin) are in blue, less known targets are in orange (Tchem) and targets with no known drug are in red (Tbio). (B) Target class distribution among the datasets, and (C) matching drugs, both approved antineoplastic drugs or other drugs (non-anticancer drugs) described using the ATC classification. ABC: advanced breast cancer; EBC: early breast cancer; Metas: metastases

### Proteins and processes of interest in the clonal proteome dataset for target discovery

In the clonal proteome dataset, proteins shared among samples or specific to primary tumors, stroma or metastases, or differentially expressed may associate with biological processes intrinsic to breast cancer stage, tumor microenvironment, or progression. These proteins may therefore be of interest, especially if they also associate with breast cancer survival. Protein distribution among patients showed that 200 proteins were found in all primary tumor samples (**Figure 5A**), 65 proteins were shared among all the stromal samples (**Figure 5B**), 98 proteins were found in all the metastases samples **(Figure 5C)**, and 37 proteins were present in all the 124 clonal samples (**Figure 5D**). Enrichment analyses for specific pathways showed as main biological processes in primary tumors AUF1, DNA-PK, S193-KSRP, or CDK5 related activities (**Figure 5E**), in stroma BGN activity, keratin sulfate cleavage, AUF1 ubiquitinylation, and C5 pathway activity (**Figure 5F**), and in metastases DCN, HSP90, and MAP2Ks related activities (**Figure 5G**). In addition, ficolin-rich granule exocytosis and cellular response to heat stress were among the main processes found in all samples (**Figure 5H**). Enrichment analyses of proteins specific to primary tumors (n=273), stroma (n=107) or metastases (n=215) showed biological processes related to membrane components of the endoplasmic reticulum, RAS or MAPK signaling in primary tumors (**Figure 6A**), AP-2, clathrin, or PP2A activities or ketone body metabolism in stroma (**Figure 6B**), oxidation, contractile fibers processes, and drug metabolism in metastases (**Figure 6C**).

**Figure 5:**
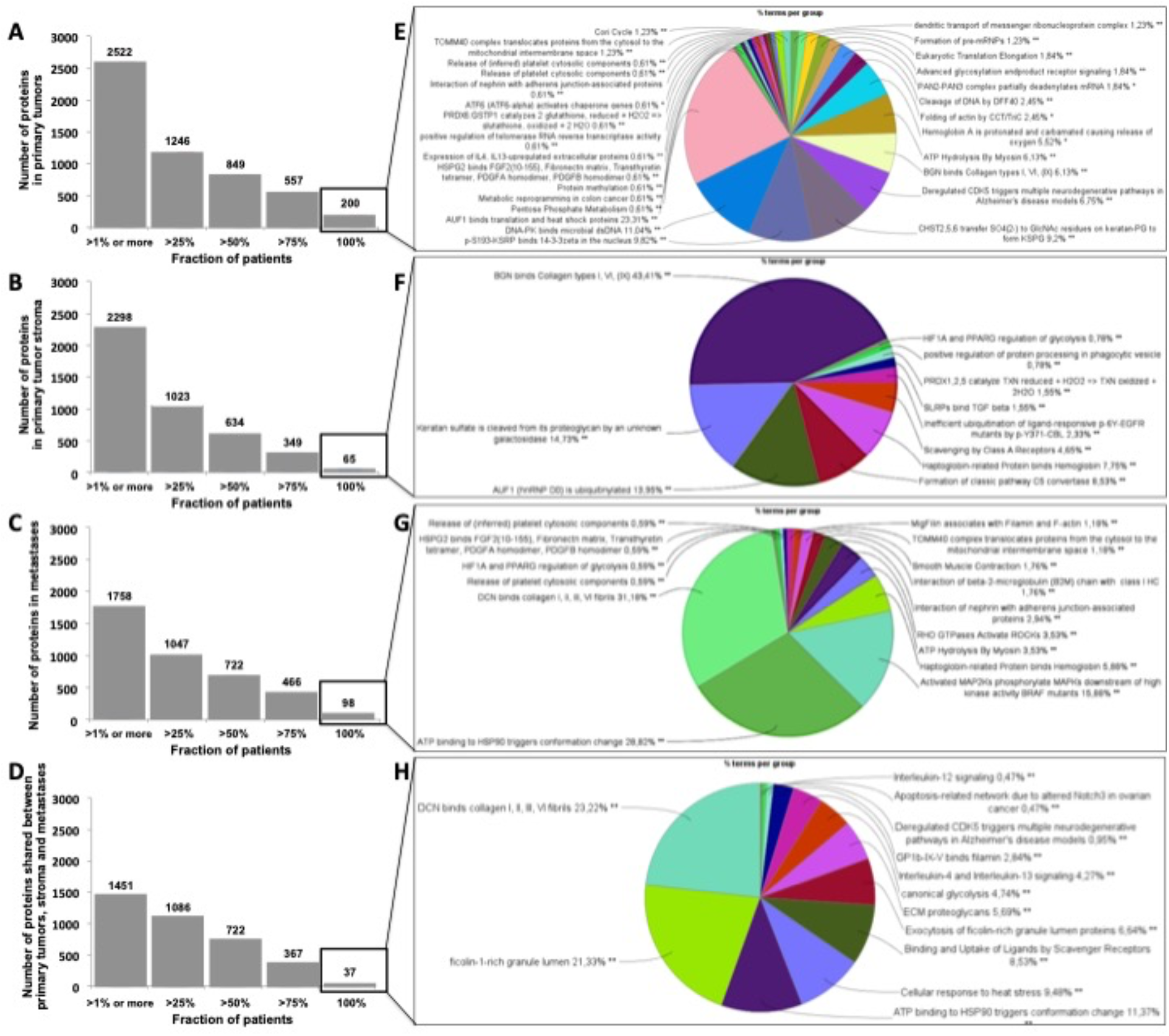
Distribution of proteins among patients in (A) primary tumors, (B) in stroma, (C) in metastases, and (D) shared in all samples. Biological processes enriched (in %) from proteins shared by all patients in (A) (B) (C) (D) are represented as pie charts in (E) (F) (G) (H), respectively. Analyses were performed with Cytoscape and ClueGo.

**Figure 6:**
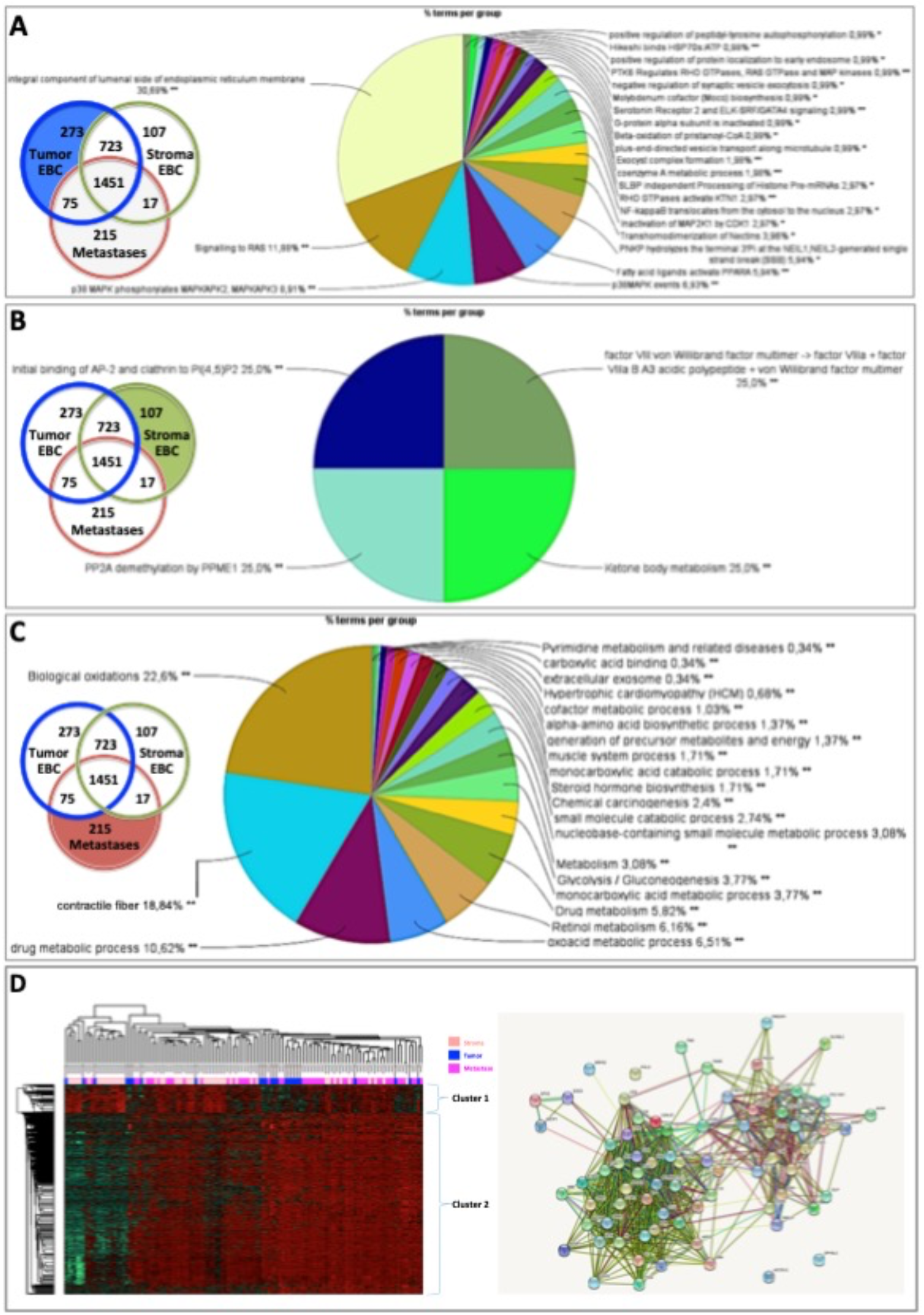
Biological processes enriched from proteins specifically found in (A) primary tumors, (B) stroma, or (C) metastases. Proteins differentially expressed in (D) primary tumors, stroma and metastases were analyzed using a multiple sample test ANOVA with a p<0.01 and represented in a heat map (on the left) identifying 2 clusters separating the stroma (Cluster 1) from the primary tumor and the metastases (Cluster 2). The String networks of the clusters are shown on the bottom right Analyses were performed with Cytoscape, ClueGo and String; proportions of processes in %.

Proteins differentially expressed among primary tumors, stroma and metastases were identified using a multiple sample test ANOVA with a p<0.01. A total of 662 proteins showed a significant difference in expression among the 3 groups. Two clusters have been identified separating the stroma (Cluster 1) from the primary tumor and the metastases (Cluster 2) (**Figure 6D**). String analysis of Cluster 1 revealed two separated networks linked by VCAN *i*.*e*. one centered on immune response inhibition and one on collagen proteins and protein in interaction with the extracellular matrix (**Supplementary material 10**). In Cluster 2, gene ontology reflects the presence of paraspeckles, VCP-NSFL1C complex, cytosolic small ribosomal subunit, cytosolic and polysomal ribosome, SNP and RNP complexes networks. KEGG Pathways identify as major networks the ones related to metabolism (lipids, glycosgelysis, pyruvate, proteins, carbon, butanoate, amino acid residues), antigen processing and presentation (**Supplementary material 10**). The relationship between the proteins of interest in the clonal proteome (shared, specific or differentially expressed) with TCGA, BC360 or CDx datasets on the one hand, and with survival on the other hand was detailed in **Figure 7**. Less than 5% of these proteins of interest were shared with the genomic/transcriptomic datasets, and 25% were associated with distant metastases free survival (DMFS) (n=222) or overall survival (OS) (n=227). Enrichment analyses of genes associated with both breast cancer DMFS and OS showed their involvement in natural killer cell mediated cytotoxicity, drug metabolism, muscle filament, ERBB2 and leptin signaling, aminoacid synthesis and B cell receptor signaling (**Figure 8A**). Among the proteins of interest, 48 (5%) had interactions with known drugs, mostly non-anticancer agents such as colchicine, acemetacin, aceclofenac (musculo-skeletal system), astemizole (respiratory system), eptifibatide (blood system), or acetyldigitoxin (cardiovascular system), which have shown anti-tumor activity experimentally in breast cancer (**Figure 8B**). Among the mutated proteins, 10 reference genes were associated with breast cancer DMFS (**Figure 9A**) or OS (**Figure 9B**) and were involved in the response to interferon gamma.

**Figure 7:**
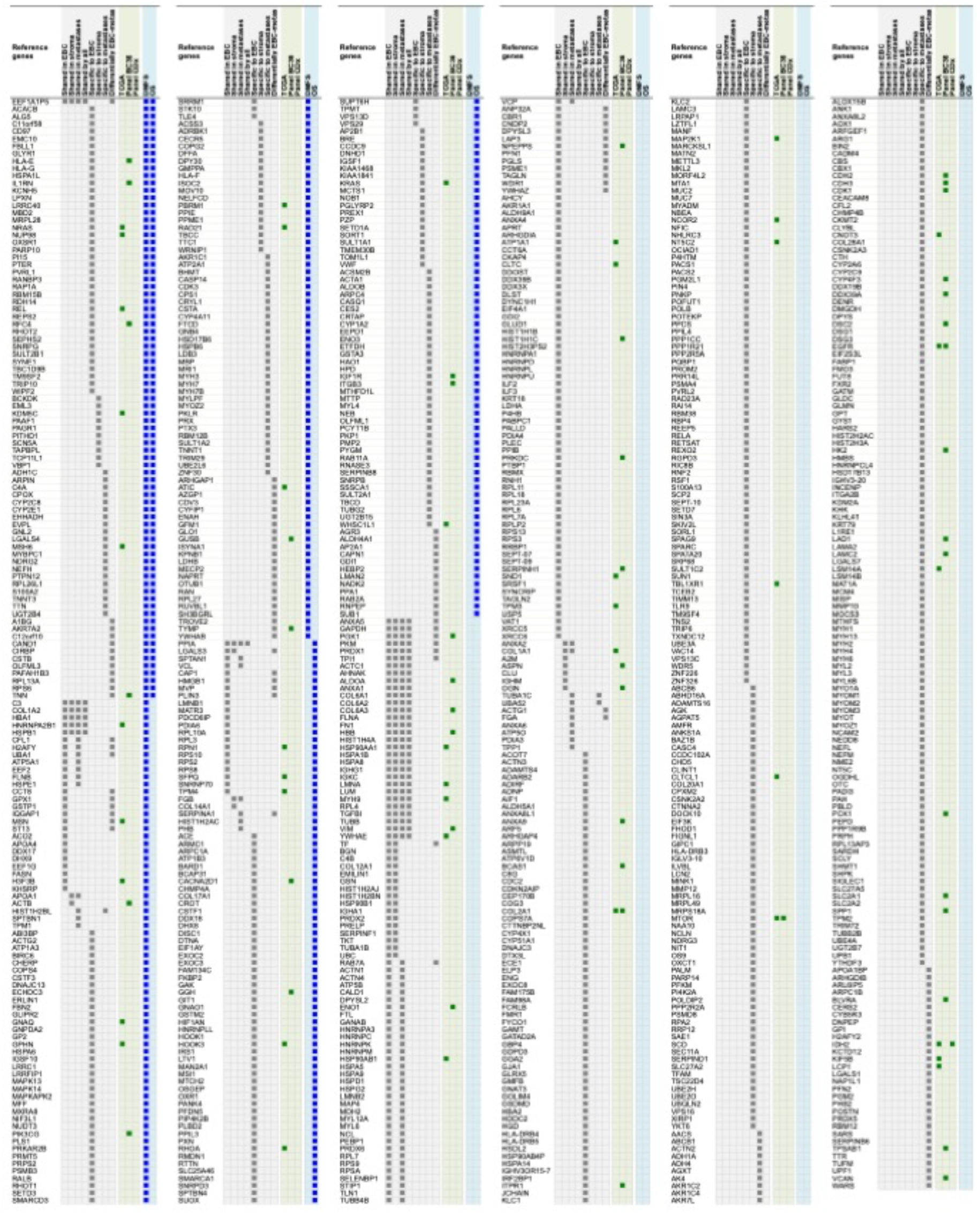
Proteins shared, specific or differentially expressed in the clonal proteome dataset (in grey). 892 proteins are indicated with their reference gene. Proteins also found in TCGA, BC360 or CDx datasets are indicated in green. Genes associated with DMFS or OS using publically available databases (Gyorffy et al. 2010) are shown in blue. EBC: early breast cancer; DMFS: distant metastases free survival; OS: overall survival.

**Figure 8:**
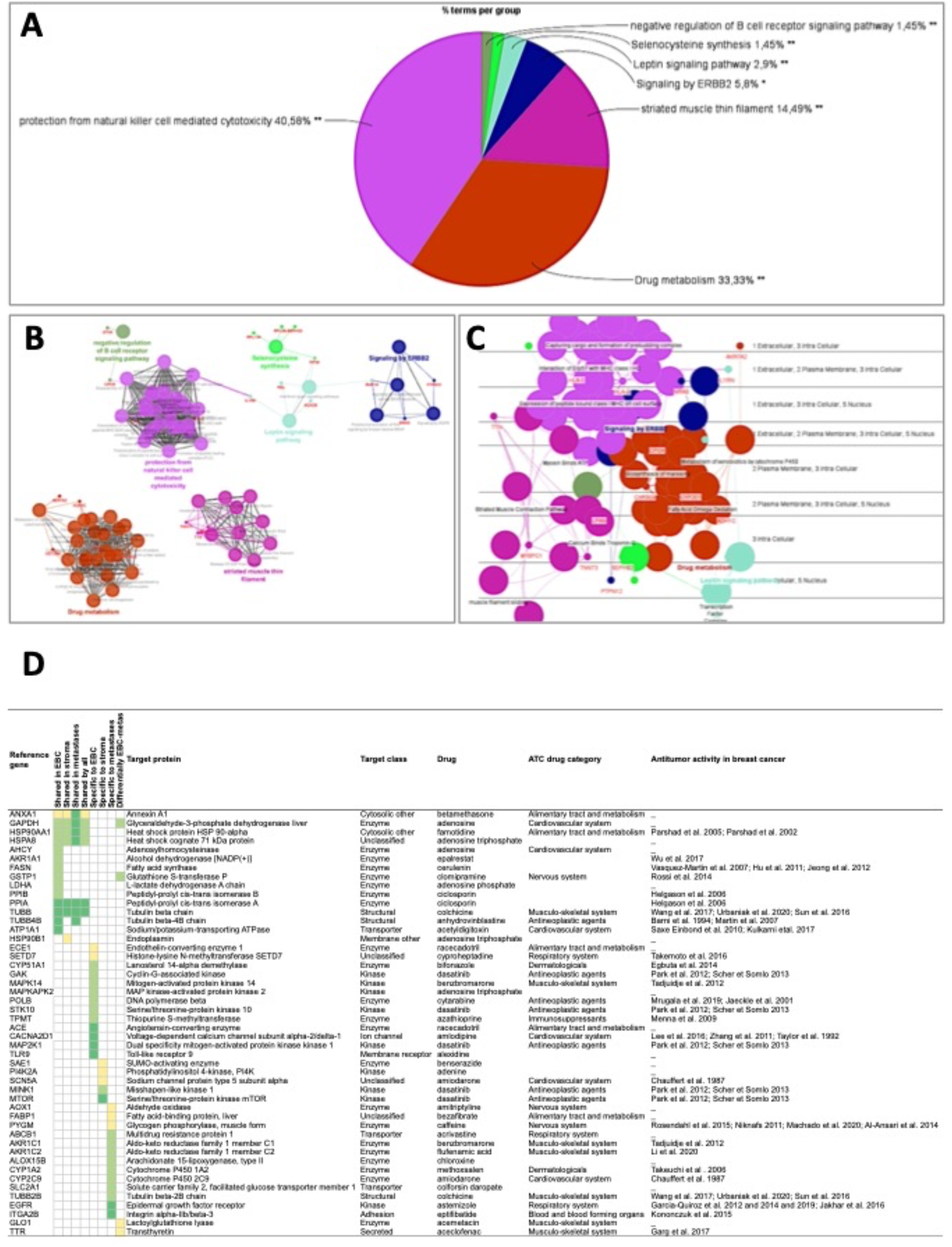
Analyses of the proteins from the clonal proteome associated both with DMFS and OS, showing (A) enrichment, (B) networks analysis, (C) cerebral layout of cellular distribution, and (D) their druggability. In the table, targets with a known drug-target interaction are in dark green, those with a less known interaction are in green, and those with limited data are in yellow. The target class, matching drug name, ATC classification and reported antitumor activity in breast cancer are indicated.

**Figure 9:**
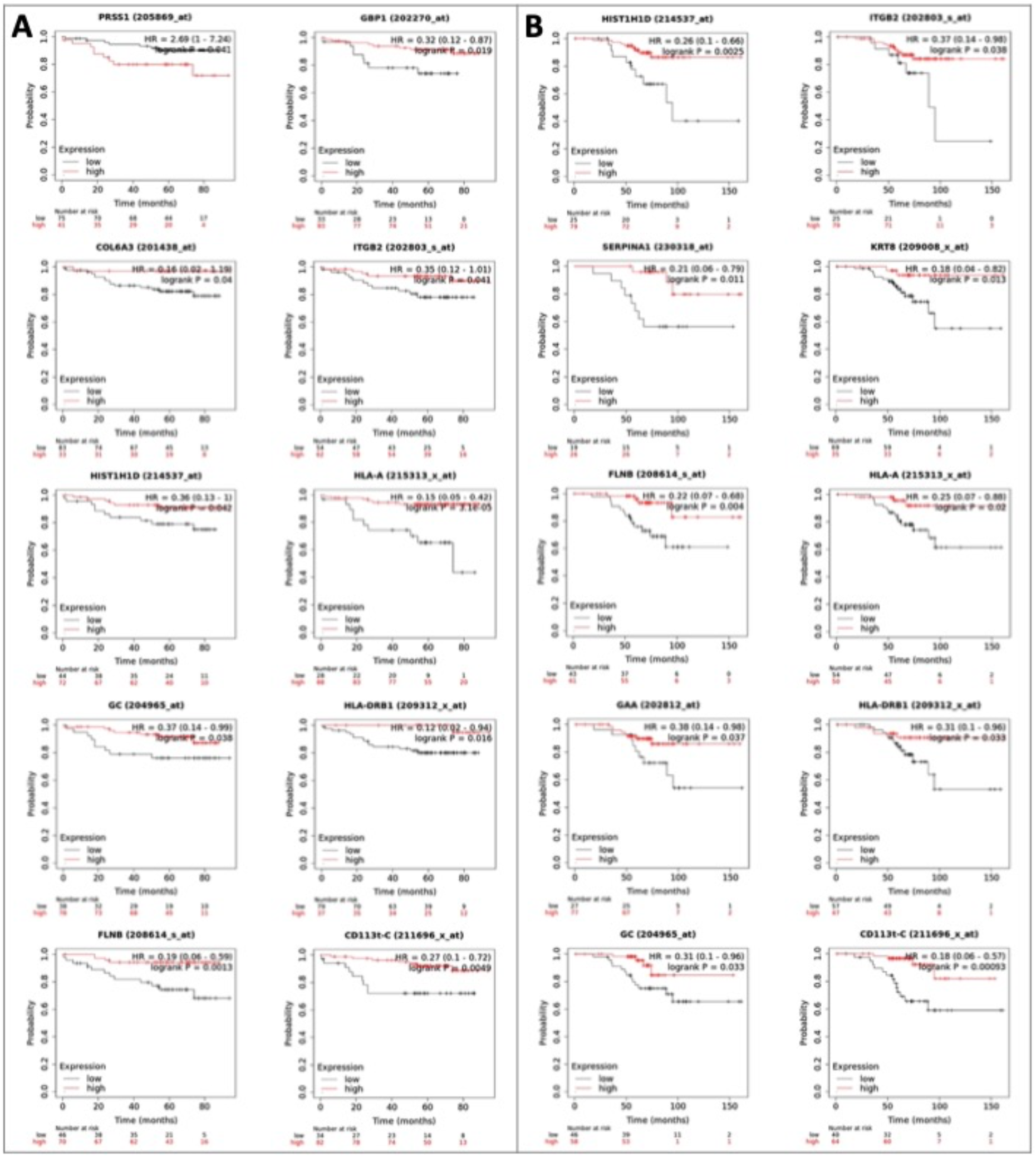
Reference genes of the mutated proteins identified in the clonal proteome dataset associated with breast cancer (A) DMFS or (B) OS. The breast cancer Kaplan-Meier plotter tool was used to run multiple reference gene testing in publically available estrogen receptor positive and HER2 negative cohorts. A logrank p<0.05 was considered significant. DMFS: distant metastases free survival; HR: hazard ratio; OS: overall survival.

## Discussion

The present study showed that MALDI MSI combined with microproteomics successfully mapped and profiled specific tumor subpopulations in luminal breast cancers based on their intra-tumor proteomic heterogeneity. This unsupervised and label-free technology characterized the tumors conventional proteome along with the mutated and alternative proteomes, at a clonal level, to identify candidate druggable targets. Our MS imaging and microproteomic technology offers the advantage of identifying proteomic clones in situ on intact tumors. In MALDI MSI, the signal intensities are recorded for analytes at specific x,y coordinates of the tissue section in their native states. MSI produces images of the scanned area where each pixel contains the MS spectrum at this location. In the present study, 204 spectra were generated by square millimeter. Spectra are high-dimensional vectors (typically in the order of 10^5^ dimensions), making MSI data similar to hyperspectral images. The spectrum produced by MSI at a given location represents a signature of the molecules present at this location. This proteomic signature was used to provide a label free unbiased method to map and visualize tumor functional heterogeneity and perform directed proteomic profiling on selected tumor subpopulations. This label free method is a strength compared to multiplex technics that require few selected markers (Giesen et al. 2014). This makes the MSI-microproteomics technology particularly suited to discovery. Relevant candidate tumor targets for drug development are sought mainly among tumor genomic alterations because of their putative role as oncogenic drivers and in drug resistance. Implementation of this concept in the clinic has yielded mitigated benefit for patients (Marquart et al. 2018). Moreover, the majority of oncogenic mutations are not druggable (Beltran et al. 2015). Target inference from bioinformatic analyses based on genomic data may not provide knowledge precise enough about the functional state of the tumor and its diversity to identify relevant targets. Searching candidate targets among proteins circumvents these limitations. The performance and usefulness of the MSI-microproteomic technology was previously reported in solid tumors such as ovarian cancer or gliomas to help finding novel biomarkers or refining diagnosis classification (Delcourt et al. 2017; Le Rhun et al. 2017). Additionally, MSI-microproteomics tumor subpopulation scale allowed a successful identification of specific tumor stroma proteins. This is a significant advantage given the involvement of the tumor microenvironment in drug response (Nakasone et al. 2012), contrary to single cell methods, which cannot analyze intercellular communications in their intact microenvironment.

The clonal proteome showed a rich landscape of proteins and biological processes compared to genomic or transcriptomic datasets. The overlap with TCGA data and transcriptomic panels was limited and a distinct distribution of biological processes was observed in enrichment analyses. The clonal proteomic dataset provided more information on enzymatic and metabolic processes. A study by Patel et al. reporting on a computational assessment of drug targets showed that enzymes were the most frequent protein class and the most druggable (Patel et al., 2013). The clonal proteome revealed 41 exclusive metabolic pathways, most of them understudied in relation to breast cancer. This was of particular interest because tumor metabolic phenotype has been recognized as a hallmark of cancer and is involved in drug resistance (Wang et al. 2017). Transcriptomic panels were enriched with kinases or immune processes as expected. The discrepancy with TCGA data may be related to alternative splicing and post-translational modifications revealed by mass spectrometry analyses that cannot be predicted by genome databases. Although the bulk of targetable proteins identified might not be involved in driver oncogenic pathways to which cancer cells are addicted, focusing on common and cell type- or stage-specific proteins and processes might increase their relevance. The present work showed that a large number of proteins in the clonal proteome dataset were associated with breast cancer outcome and highlighted shared or specific biological processes.

An additional strength of the present mass spectrometry technology relies on the detection of altered proteins, such as those originating from missense mutations or single nucleotide polymorphism missense mutations, and a newly recognized type of proteins named alternative proteins (or ghost proteins) because of their translation from alternative open reading frames. Although their functions cannot be predicted from their reference gene, their presence may reveal altered biological processes. Alternative proteins represent a vast class of proteins with still largely unknown biological functions, thus expanding the proteome complexity (Cardon et al. 2020). This field of research offers exciting perspectives about the functions of these modified proteins related to cancer and their potential impact on drug target interactions.

Our study showed that a clonal proteomic analysis brought additional non-redundant molecular information. The proteins and pathways uniquely identified in the clonal proteomic dataset may offer opportunities to identify novel drug targets. Drug development struggles with the molecular heterogeneity of tumor subpopulations, potentially leading to a differential target expression among cancer cells, which contributes to drug resistance. This has stimulated the development of multi-targeted therapeutic strategies (Sicklick et al. 2019; Al-Lazikani et al. 2012), facilitated by the fast expansion of the drug pipeline. Interestingly, a high proportion of the proteins in the clonal proteome dataset were druggable, with interactions with a variety of drug classes, either antineoplastic agents or non-anticancer drugs. Therefore, the MSI-microproteomic technology may offer opportunities in strategies of systematic high-throughput unbiased drug target screening for drug combination or repositioning (Wurth et al. 2016). A significant number of proteins had partially or not yet known drug interactions, showing also the potential of our approach for discovery.

To tackle tumor complexity, system biology approaches are developing to reveal therapeutic opportunities associated with the multiple dimensions of cancer through integration of tumor genome, phenome and other omics data (Griffiths et al. 2019; Kristensen et al. 2014). Accessing sufficient quantities of tumor tissue to perform all the omics analyses represents a technical challenge. Our technology uses only a limited amount of tumor tissue while maintaining the whole tissue section integrity allowing it to be re-used for additional experiments. For this reason and the large amount of data generated, the MSI-microproteomic technology is suited to multiomic strategies.

The present study has some limitations. The bisecting kmeans method visualizes and controls the number of tumors’ subclones manually. No data is yet available to determine the optimal group-split for breast cancer subclones identification with respect to clinical relevance. A study by Balluff et al. analyzed the clinical significance of the tumor subpopulations identified from tumor segmentation to up to 10 group-split in gastric tumors. The study showed that a significant prognostic effect became visible after 3 to 6 splits because it revealed smaller tumor subpopulations. Three to four segmentation splits were sufficient to categorize gastric subclones into poor, medium or good survival groups (Balluff et al. 2012). More data are needed to determine whether breast cancer subclones identified by MSI follow a similar pattern. In the present study, the number of split was 3 to 4 to identify tumor subpopulations with main differences among their proteomic signatures. Our two-step workflow requires a semi-automated microproteomic part because the current system does not allow protein identification directly from the MSI spectra. Further instrumental and bioinformatics developments are necessary to achieve a full automation and further increase the analysis speed of the platform and improve the spatial resolution to reach cellular resolution. The clonal proteome dataset had an underrepresentation in nuclear proteins as a consequence of owner in situ microdigestion and microextraction methods calibrated to capture proteins globally for discovery. Developments to enrich the clonal extract in nuclear proteins, phosphorylated or membrane proteins is foreseeable for specific protein classes analyses. The MALDI MSI combined with microproteomic technology provided a large amount of molecular information about classic proteins, mutated proteins and also alternative proteins in breast cancer subpopulations. However, assessment of their relevance as target candidates requires a validation step, which was beyond the scope of the present study. Drug target validation is a laborious multi-step process requiring sufficient pre-clinical and clinical validation to finally vet a candidate target (Settleman et al. 2018).

In conclusion, spatially resolved MSI combined with microproteomics represents a unique tool to perform a label-free multidimensional proteomic characterization of intratumor heterogeneity. By improving knowledge about tumor functional clones, this technology has the potential to expand the libraries used for drug discovery with clone-specific biomarkers and targets and offers opportunities for drug repurposing and combinatorial strategies in breast cancer. Its applicability to small samples, to FFPE samples, and its scalability thanks to the speed of analysis of current and next generation mass spectrometry instruments, make MSI-microproteomics integration to precision oncology tools foreseeable in a near future to implement clone-tailored therapies.

## Data Availability

All raw data are available as supplementary material files and on ProteomeXchange with identifier PXD024134.

## Abbreviations

ABC: advanced breast cancer
ACN: acetonitril
AGC: automatic gain control
AltProt: alternative proteins
ANOVA: analysis of variance
ATC: anatomical therapeutic chemical
BC: breast cancer
Da: dalton
DMFS: distant metastases free survival
EBC: early breast cancer
EMA: european medicines agency
FDA: food and drug administration
FDR: false discovery rate
FFPE: formalin-fixed paraffin-embedded
HCCA: hydroxycinnamic acid
HER2: human epidermal growth factor receptor-2
HR: hazard ratio
HUGO: human genome organization
ID: identification
IDG: illuminating the druggable genome
IDG-KMC: illuminating the druggable genome knowledge management center
ITO: indium tin oxide
LC: liquid chromatography
LESA: liquid extraction surface analysis
LFQ: label-free quantification
MALDI: matrix assisted laser desorption ionization
MeOH: methanol
Meta: metastases
MS: mass spectrometry
MSI: mass spectrometry imaging
OS: overall survival
PMDA: pharmaceuticals and medical devices agency
PSM: peptide spectrum matches
SNP: single nucleotide polymorphism
TDL: target development level
TFA: trifluoroacetic acid
TIC: total ion count

## DECLARATIONS

Ethics approval and consent to participate: All the analyses of this retrospective study were approved by the local Research and Ethics Committee (Centre Oscar Lambret) in accordance with the French and European legislation. Prior to the analyses, patients signed an informed consent and authorization form for the use of their data and biological samples. No personal information was used in these experiments, and a random number was assigned to each sample.

### Consent for publication

All patient consent for publication.

### Competing interests

**All** the authors declare they have no competing interests.

### Funding

This work was funded by Inserm and Centre Oscar Lambret.

### Author contributions

N.H. wrote the manuscript original draft; N.H. designed the study; S. A. and N.H. performed the analyses; N.H. and D.B. selected and collected the breast cancer samples; Y-M.R. and D.B. performed histology and validated diagnostics; N.H., S.A., T.C., I.F. and M.S. analyzed the data; I.F., M.S., T.C. and S.A. corrected the manuscript; N.H, I.F. and M.S. supervised the project and M.S., I.F. and N.H. provided the funding.

## Figure Legends

**Supplementary Table 1:** Information concerning patients and type of tumors identified by the pathologist

| Sample ID | Tissue type | Stage | Patient ID* | Method of sample collection and concomitant treatment |
| --- | --- | --- | --- | --- |
| 1 | breast | EBC | 1 | surgery |
| 2 | breast | EBC | 2 | surgery |
| 3 | breast | EBC | 3 | surgery |
| 4 | breast | EBC | 4 | surgery ; chemotherapy |
| 5 | breast | EBC | 5 | surgery |
| 6 | breast | EBC | 6 | surgery |
| 7 | breast | EBC | 7 | surgery |
| 8 | breast | EBC | 8 | surgery |
| 9 | breast | EBC | 9 | surgery |
| 10 | breast | EBC | 10 | surgery |
| 11 | breast | EBC | 11 | surgery |
| 12 | breast | EBC | 12 | surgery |
| 13 | breast | EBC | 13 | surgery |
| 14 | breast | EBC | 14 | surgery |
| 15 | breast | EBC | 15 | surgery |
| 16 | breast | EBC | 16 | surgery |
| 17 | breast | EBC | 17 | surgery |
| 18 | breast | EBC | 18 | surgery |
| 19 | breast | EBC | 19 | surgery |
| 20 | breast | EBC | 20 | surgery |
| 21 | breast | EBC | 21 | surgery |
| 22 | breast | EBC | 22 | surgery |
| 23 | breast | EBC | 23 | surgery |
| 24 | breast | EBC | 24 | surgery |
| 25 | breast | EBC | 4 | surgery ; chemotherapy |
| 26 | breast | EBC | 25 | surgery |
| 27 | breast | EBC | 26 | surgery |
| 28 | breast | EBC | 27 | surgery |
| 29 | breast | EBC | 28 | surgery |
| 30 | breast | EBC | 29 | surgery |
| 31 | breast | EBC | 30 | surgery |
| 32 | breast | EBC | 31 | surgery |
| 33 | breast | EBC | 32 | surgery |
| 34 | breast | EBC | 33 | surgery |
| 35 | breast | EBC | 34 | surgery |
| 36 | breast | EBC | 35 | surgery |
| 37 | breast | EBC | 36 | surgery |
| 38 | breast | EBC | 37 | surgery |
| 39 | breast | EBC | 38 | surgery |
| 40 | breast | EBC | 39 | surgery |
| 41 | breast | EBC | 40 | surgery |
| 42 | breast | EBC | 41 | surgery |
| 43 | breast | EBC | 42 | surgery |
| 44 | breast | EBC | 43 | surgery |
| 45 | breast | EBC | 44 | surgery |
| 46 | breast | EBC | 45 | surgery |
| 47 | breast | EBC | 46 | surgery |
| 48 | breast | EBC | 47 | surgery |
| 49 | breast | EBC | 48 | surgery |
| 50 | breast | EBC | 49 | surgery |
| 51 | breast | EBC | 50 | surgery |
| 52 | breast | EBC | 51 | surgery |
| 53 | skin | ABC | 52 | surgical biopsy ; endocrine therapy |
| 54 | skin | ABC | 53 | surgical biopsy ; endocrine therapy |
| 55 | node | ABC | 53 | surgical biopsy ; endocrine therapy |
| 56 | skin | ABC | 54 | surgical biopsy ; endocrine therapy |
| 57 | node | ABC | 53 | surgical biopsy ; endocrine therapy |
| 58 | skin | ABC | 54 | surgical biopsy ; endocrine therapy |
| 59 | bone | ABC | 52 | FNB ; chemotherapy |
| 60 | liver | ABC | 39 | FNB |
| 61 | node | ABC | 55 | surgical biopsy ; chemotherapy |
| 62 | liver | ABC | 56 | FNB ; chemotherapy |
| 63 | skin | ABC | 57 | surgical biopsy ; endocrine therapy |
| 64 | liver | ABC | 39 | FNB ; chemotherapy |
| 65 | node | ABC | 56 | FNB ; chemotherapy |
| 66 | liver | ABC | 58 | FNB ; endocrine therapy |
| 67 | skin | ABC | 57 | surgical biopsy |
| 68 | skin | ABC | 4 | FNB ; chemotherapy |
| 69 | bone | ABC | 58 | FNB ; endocrine therapy |
| 70 | skin | ABC | 57 | FNB |
| 71 | liver | ABC | 59 | FNB ; endocrine therapy |
| 72 | node | ABC | 60 | FNB |
| 73 | skin | ABC | 57 | FNB |
| 74 | bone | ABC | 60 | FNB ; endocrine therapy |
| 75 | node | ABC | 17 | FNB ; chemotherapy |
| 76 | skin | ABC | 54 | FNB ; endocrine therapy |

\* Some patients had multiple sample collection through the course of the disease; ABC: advanced breast cancer; EBC: early breast cancer; FNB: fine needle biopsy

**Supplementary material 1:** TCGA database of mutations and CNV alterations in early and advanced breast cancers

**Supplementary material 2:** BC360 panel gene list

**Supplementary material 3:** CDx panel gene list

**Supplementary material 4:** MALDI MSI of 52 cases of primary tumors showing the spatial proteomic heterogeneity of the tumors. In each sample vignette, the MALDI MS imaging is displayed with the histological HPS picture (upper left), the principal component analysis of the proteomic clones (upper right), the segmentation tree (middle right), and the spectra of the clones (bottom right).

**Supplementary material 5:** MALDI MSI of 24 cases of metastases showing the spatial proteomic heterogeneity of the tumors. In each sample vignette, the MALDI MS imaging is displayed with the histological HPS picture (upper left), the principal component analysis of the proteomic clones (upper right), the segmentation tree (middle right), and the spectra of the clones (bottom right).

**Supplementary material 6:** Number of total proteins identified according to (A) tissue types (bone n=3 ; liver n=5 ; nodes n=6 ; skin n=10) or (B) to the sampling method (B), i.e. surgery (n=52), surgical biopsy (n=9) or fine needle core biopsy (n=15). Line in the middle of the box is the median, the upper and lower sides of the box represent the 75th and 25th percentiles respectively, and the outside lines are the extremes.

**Supplementary material 7:** Panther analysis of the clonal proteome landscape showing (A) the protein class distribution (in %), and (B) a comparison of the distribution between primary tumors and stroma (relative difference in % in blue) and between primary tumors and metastases (relative difference in % in red).

**Supplementary material 8:** Reference gene distribution across the four datasets (clonal proteome, TCGA, BC360, CDx)

**Supplementary material 9:** Targets interacting with anticancer drugs across the four datasets (clonal proteome, TCGA, BC360, CDx)

**Supplementary material 10:** String networks in stroma (cluster 1) and in primary tumors and metastases (cluster 2)

## Notes

### Competing Interest Statement

The authors have declared no competing interest.

### Clinical Trial

N/A

