## supplementary material (1-10) for "Path to drugging functional clones of luminal breast cancers using in-depth proteomics with spatially resolved mass spectrometry guided by MALDI imaging"

Supplemental material 1

| Genes TCGA early BC | Name | Genes TCGA advanced BC | Name |
|---|---|---|---|
| **TP53** | tumor protein p53 | **PIK3CA** | phosphatidylinositol-4,5-bisphosphate 3-kinase, catalytic subunit alpha |
| **PIK3CA** | phosphatidylinositol-4,5-bisphosphate 3-kinase, catalytic subunit alpha | **TP53** | tumor protein p53 |
| **CDH1** | cadherin 1, type 1, E-cadherin (epithelial) | **CLIP1** | CAP-GLY domain containing linker protein 1 |
| **GATA3** | GATA binding protein 3 | **MUC1** | mucin 1, cell surface associated |
| **KMT2C** | lysine (K)-specific methyltransferase 2C | **TSHR** | thyroid stimulating hormone receptor |
| **MAP3K1** | mitogen-activated protein kinase kinase kinase 1, E3 ubiquitin protein ligase | **FGFR2** | fibroblast growth factor receptor 2 |
| **PTEN** | phosphatase and tensin homolog | **CBLB** | Cbl proto-oncogene B, E3 ubiquitin protein ligase |
| **NCOR1** | nuclear receptor corepressor 1 | **IL21R** | interleukin 21 receptor |
| **NF1** | neurofibromin 1 | **AMER1** | APC membrane recruitment protein 1 |
| **RUNX1** | runt-related transcription factor 1 | **NSD1** | nuclear receptor binding SET domain protein 1 |
| **ARID1A** | AT rich interactive domain 1A (SWI-like) | **ETNK1** | ethanolamine kinase 1 |
| **PIK3R1** | phosphoinositide-3-kinase, regulatory subunit 1 (alpha) | **AKAP9** | A kinase (PRKA) anchor protein 9 |
| **MAP2K4** | mitogen-activated protein kinase kinase 4 | **PTPRB** | protein tyrosine phosphatase, receptor type, B |
| **RNF213** | ring finger protein 213 | **ERBB2** | erb-b2 receptor tyrosine kinase 2 |
| **KMT2D** | lysine (K)-specific methyltransferase 2D | **NOTCH1** | notch 1 |
| **SPEN** | spen family transcriptional repressor | **TPR** | translocated promoter region, nuclear basket protein |
| **AKAP9** | A kinase (PRKA) anchor protein 9 | **FLI1** | Fli-1 proto-oncogene, ETS transcription factor |
| **FAT1** | FAT atypical cadherin 1 | **CDH1** | cadherin 1, type 1, E-cadherin (epithelial) |
| **MYH9** | myosin, heavy chain 9, non-muscle | **MEN1** | multiple endocrine neoplasia I |
| **TBX3** | T-box 3 | **MLLT10** | myeloid/lymphoid or mixed-lineage leukemia (trithorax homolog, Drosophila); translocated to, 10 |
| **MED12** | mediator complex subunit 12 | **CAMTA1** | calmodulin binding transcription activator 1 |
| **KMT2A** | lysine (K)-specific methyltransferase 2A | **AKT1** | v-akt murine thymoma viral oncogene homolog 1 |
| **MLLT4** | myeloid/lymphoid or mixed-lineage leukemia (trithorax homolog, Drosophila); translocated to, 4 | **NFATC2** | nuclear factor of activated T-cells, cytoplasmic, calcineurin-dependent 2 |
| **ATRX** | alpha thalassemia/mental retardation syndrome X-linked | **NOTCH2** | notch 2 |
| **FOXA1** | forkhead box A1 | **CHD4** | chromodomain helicase DNA binding protein 4 |
| **PTPRB** | protein tyrosine phosphatase, receptor type, B | **FLCN** | folliculin |
| **ERBB2** | erb-b2 receptor tyrosine kinase 2 | **MSI2** | musashi RNA-binding protein 2 |
| **ATM** | ATM serine/threonine kinase | **CREB1** | cAMP responsive element binding protein 1 |
| **TPR** | translocated promoter region, nuclear basket protein | **RALGDS** | ral guanine nucleotide dissociation stimulator |
| **AKT1** | v-akt murine thymoma viral oncogene homolog 1 | **ABL2** | ABL proto-oncogene 2, non-receptor tyrosine kinase |
| **UBR5** | ubiquitin protein ligase E3 component n-recognin 5 | **NDRG1** | N-myc downstream regulated 1 |
| **CHD4** | chromodomain helicase DNA binding protein 4 | **KCNJ5** | potassium channel, inwardly rectifying subfamily J, member 5 |
| **BRCA1** | breast cancer 1, early onset | **ERBB3** | erb-b2 receptor tyrosine kinase 3 |
| **ARID1B** | AT rich interactive domain 1B (SWI1-like) | **CHEK2** | checkpoint kinase 2 |
| **MTOR** | mechanistic target of rapamycin (serine/threonine kinase) | **MYCL** | v-myc avian myelocytomatosis viral oncogene lung carcinoma derived homolog |
| **BRCA2** | breast cancer 2, early onset | **BCL11B** | B-cell CLL/lymphoma 11B (zinc finger protein) |
| **NOTCH2** | notch 2 | **PTPN11** | protein tyrosine phosphatase, non-receptor type 11 |
| **SETD2** | SET domain containing 2 | **PCSK7** | proprotein convertase subtilisin/kexin type 7 |
| **CBFB** | core-binding factor, beta subunit | **GNAS** | GNAS complex locus |
| **RB1** | retinoblastoma 1 | **SOCS1** | suppressor of cytokine signaling 1 |
| **GRIN2A** | glutamate receptor, ionotropic, N-methyl D-aspartate 2A | **GNA11** | guanine nucleotide binding protein (G protein), alpha 11 (Gq class) |
| **FAT4** | FAT atypical cadherin 4 | **RAD21** | RAD21 homolog (S. pombe) |

| | | | |
|---|---|---|---|
| **RUNX1T1** | runt-related transcription factor 1; translocated to, 1 (cyclin D-related) | **COX6C** | cytochrome c oxidase subunit VIc |
| **ERBB3** | erb-b2 receptor tyrosine kinase 3 | **LIFR** | leukemia inhibitory factor receptor alpha |
| **TET1** | tet methylcytosine dioxygenase 1 | **PMS2** | PMS2 postmeiotic segregation increased 2 (S. cerevisiae) |
| **ATR** | ATR serine/threonine kinase | **ACKR3** | atypical chemokine receptor 3 |
| **SF3B1** | splicing factor 3b, subunit 1, 155kDa | **MAP2K2** | mitogen-activated protein kinase kinase 2 |
| **PDE4DIP** | phosphodiesterase 4D interacting protein | **EZR** | ezrin |
| **AMER1** | APC membrane recruitment protein 1 | **FGFR1OP** | FGFR1 oncogene partner |
| **KDM6A** | lysine (K)-specific demethylase 6A | **CARS** | cysteinyl-tRNA synthetase |
| **CREBBP** | CREB binding protein | **MTOR** | mechanistic target of rapamycin (serine/threonine kinase) |
| **TRRAP** | transformation/transcription domain-associated protein | **PPP6C** | protein phosphatase 6, catalytic subunit |
| **STAG2** | stromal antigen 2 | **FANCF** | Fanconi anemia, complementation group F |
| **MYH11** | myosin, heavy chain 11, smooth muscle | **ATM** | ATM serine/threonine kinase |
| **ZFHX3** | zinc finger homeobox 3 | **TET2** | tet methylcytosine dioxygenase 2 |
| **KAT6B** | K(lysine) acetyltransferase 6B | **FH** | fumarate hydratase |
| **CTCF** | CCCTC-binding factor (zinc finger protein) | **PRF1** | perforin 1 (pore forming protein) |
| **CASP8** | caspase 8, apoptosis-related cysteine peptidase | **IKZF1** | IKAROS family zinc finger 1 (Ikaros) |
| **ATP2B3** | ATPase, Ca++ transporting, plasma membrane 3 | **FOXA1** | forkhead box A1 |
| **EP300** | E1A binding protein p300 | **NUP214** | nucleoporin 214kDa |
| **TET2** | tet methylcytosine dioxygenase 2 | **BRAF** | B-Raf proto-oncogene, serine/threonine kinase |
| **ROS1** | ROS proto-oncogene 1 , receptor tyrosine kinase | **SMAD3** | SMAD family member 3 |
| **SMARCA4** | SWI/SNF related, matrix associated, actin dependent regulator of chromatin, subfamily a, member 4 | **USP8** | ubiquitin specific peptidase 8 |
| **EGFR** | epidermal growth factor receptor | **VTI1A** | vesicle transport through interaction with t-SNAREs 1A |
| **AFF3** | AF4/FMR2 family, member 3 | **SF3B1** | splicing factor 3b, subunit 1, 155kDa |
| **ARID2** | AT rich interactive domain 2 (ARID, RFX-like) | **RBM15** | RNA binding motif protein 15 |
| **ERBB4** | erb-b2 receptor tyrosine kinase 4 | **FLT3** | fms-related tyrosine kinase 3 |
| **NIN** | ninein (GSK3B interacting protein) | **NKX2-1** | NK2 homeobox 1 |
| **RANBP2** | RAN binding protein 2 | **TAL1** | T-cell acute lymphocytic leukemia 1 |
| **CNTRL** | centriolin | **OLIG2** | oligodendrocyte lineage transcription factor 2 |
| **FBXW7** | F-box and WD repeat domain containing 7, E3 ubiquitin protein ligase | **sept-05** | septin 5 |
| **FANCD2** | Fanconi anemia, complementation group D2 | **CDX2** | caudal type homeobox 2 |
| **AFF1** | AF4/FMR2 family, member 1 | **HIF1A** | hypoxia inducible factor 1, alpha subunit (basic helix-loop-helix transcription factor) |
| **CACNA1D** | calcium channel, voltage-dependent, L type, alpha 1D subunit | **MYOD1** | myogenic differentiation 1 |
| **KDM5A** | lysine (K)-specific demethylase 5A | **LZTR1** | leucine-zipper-like transcription regulator 1 |
| **BCR** | breakpoint cluster region | **MDM2** | MDM2 proto-oncogene, E3 ubiquitin protein ligase |
| **BLM** | Bloom syndrome, RecQ helicase-like | **FAS** | Fas cell surface death receptor |
| **KAT6A** | K(lysine) acetyltransferase 6A | **HOXC13** | homeobox C13 |
| **PTCH1** | patched 1 | **ELL** | elongation factor RNA polymerase II |
| **BCL9** | B-cell CLL/lymphoma 9 | **MAX** | MYC associated factor X |
| **APC** | adenomatous polyposis coli | **PSIP1** | PC4 and SFRS1 interacting protein 1 |
| **PTPRC** | protein tyrosine phosphatase, receptor type, C | **FOXL2** | forkhead box L2 |
| **TRIP11** | thyroid hormone receptor interactor 11 | **FBXW7** | F-box and WD repeat domain containing 7, E3 ubiquitin protein ligase |
| **ASXL1** | additional sex combs like transcriptional regulator 1 | **HNF1A** | HNF1 homeobox A |
| **MYO5A** | myosin VA (heavy chain 12, myoxin) | **HSP90AB1** | heat shock protein 90kDa alpha (cytosolic), class B member 1 |
| **NSD1** | nuclear receptor binding SET domain protein 1 | **PTPRC** | protein tyrosine phosphatase, receptor type, C |
| **NUP98** | nucleoporin 98kDa | **DCTN1** | dynactin 1 |
| **NUMA1** | nuclear mitotic apparatus protein 1 | **SRSF2** | serine/arginine-rich splicing factor 2 |

| | | | |
|---|---|---|---|
| **MECOM** | MDS1 and EVI1 complex locus | **NUMA1** | nuclear mitotic apparatus protein 1 |
| **CLTC** | clathrin, heavy chain (Hc) | **PML** | promyelocytic leukemia |
| **MAP3K13** | mitogen-activated protein kinase kinase kinase 13 | **MYH9** | myosin, heavy chain 9, non-muscle |
| **XPO1** | exportin 1 | **MECOM** | MDS1 and EVI1 complex locus |
| **ESR1** | estrogen receptor 1 | **ECT2L** | epithelial cell transforming 2 like |
| **CDK12** | cyclin-dependent kinase 12 | **BRCA1** | breast cancer 1, early onset |
| **BRIP1** | BRCA1 interacting protein C-terminal helicase 1 | **FUS** | FUS RNA binding protein |
| **WHSC1L1** | Wolf-Hirschhorn syndrome candidate 1-like 1 | **PAFAH1B2** | platelet-activating factor acetylhydrolase 1b, catalytic subunit 2 (30kDa) |
| **JAK3** | Janus kinase 3 | **SDHD** | succinate dehydrogenase complex, subunit D, integral membrane protein |
| **EIF4A2** | eukaryotic translation initiation factor 4A2 | **PIM1** | Pim-1 proto-oncogene, serine/threonine kinase |
| **BCOR** | BCL6 corepressor | **RAC1** | ras-related C3 botulinum toxin substrate 1 (rho family, small GTP binding protein Rac1) |
| **CLIP1** | CAP-GLY domain containing linker protein 1 | **MYCN** | v-myc avian myelocytomatosis viral oncogene neuroblastoma derived homolog |
| **ZNF521** | zinc finger protein 521 | **CD274** | CD274 molecule |
| **ZMYM2** | zinc finger, MYM-type 2 | **ZBTB16** | zinc finger and BTB domain containing 16 |
| **KTN1** | kinectin 1 (kinesin receptor) | **EML4** | echinoderm microtubule associated protein like 4 |
| **DDX5** | DEAD (Asp-Glu-Ala-Asp) box helicase 5 | **TFRC** | transferrin receptor |
| **CARD11** | caspase recruitment domain family, member 11 | **FHIT** | fragile histidine triad |
| **POLE** | polymerase (DNA directed), epsilon, catalytic subunit | **PTEN** | phosphatase and tensin homolog |
| **PLCG1** | phospholipase C, gamma 1 | **FANCA** | Fanconi anemia, complementation group A |
| **CUX1** | cut-like homeobox 1 | **CBL** | Cbl proto-oncogene, E3 ubiquitin protein ligase |
| **COL1A1** | collagen, type I, alpha 1 | **MAFB** | v-maf avian musculoaponeurotic fibrosarcoma oncogene homolog B |
| **GNAS** | GNAS complex locus | **TNFRSF17** | tumor necrosis factor receptor superfamily, member 17 |
| **CDKN1B** | cyclin-dependent kinase inhibitor 1B (p27, Kip1) | **CLTC** | clathrin, heavy chain (Hc) |
| **FGFR2** | fibroblast growth factor receptor 2 | **JAK2** | Janus kinase 2 |
| **CBLB** | Cbl proto-oncogene B, E3 ubiquitin protein ligase | **ERCC5** | excision repair cross-complementation group 5 |
| **JAK1** | Janus kinase 1 | **TERT** | telomerase reverse transcriptase |
| **MAML2** | mastermind-like 2 (Drosophila) | **TMPRSS2** | transmembrane protease, serine 2 |
| **TSC2** | tuberous sclerosis 2 | **SUZ12** | SUZ12 polycomb repressive complex 2 subunit |
| **CAMTA1** | calmodulin binding transcription activator 1 | **IL7R** | interleukin 7 receptor |
| **PTPN13** | protein tyrosine phosphatase, non-receptor type 13 (APO-1/CD95 (Fas)-associated phosphatase) | **MAP3K13** | mitogen-activated protein kinase kinase kinase 13 |
| **ALK** | anaplastic lymphoma receptor tyrosine kinase | **FUBP1** | far upstream element (FUSE) binding protein 1 |
| **DICER1** | dicer 1, ribonuclease type III | **CBFA2T3** | core-binding factor, runt domain, alpha subunit 2; translocated to, 3 |
| **IKBKB** | inhibitor of kappa light polypeptide gene enhancer in B-cells, kinase beta | **ARID1A** | AT rich interactive domain 1A (SWI-like) |
| **PBRM1** | polybromo 1 | **ASPSCR1** | alveolar soft part sarcoma chromosome region, candidate 1 |
| **CYLD** | cylindromatosis (turban tumor syndrome) | **NF2** | neurofibromin 2 (merlin) |
| **DCTN1** | dynactin 1 | **CIITA** | class II, major histocompatibility complex, transactivator |
| **AFF4** | AF4/FMR2 family, member 4 | **CCDC6** | coiled-coil domain containing 6 |
| **FGFR1** | fibroblast growth factor receptor 1 | **TNFRSF14** | tumor necrosis factor receptor superfamily, member 14 |
| **PDGFRA** | platelet-derived growth factor receptor, alpha polypeptide | **KLF6** | Kruppel-like factor 6 |
| **TNFAIP3** | tumor necrosis factor, alpha-induced protein 3 | **EWSR1** | EWS RNA-binding protein 1 |
| **ATP1A1** | ATPase, Na+/K+ transporting, alpha 1 polypeptide | **FGFR3** | fibroblast growth factor receptor 3 |
| **SMAD4** | SMAD family member 4 | **LCP1** | lymphocyte cytosolic protein 1 (L-plastin) |
| **STAT6** | signal transducer and activator of transcription 6, interleukin-4 induced | **HOXA9** | homeobox A9 |
| **RANBP17** | RAN binding protein 17 | **DAXX** | death-domain associated protein |

| | | | |
|---|---|---|---|
| **LIFR** | leukemia inhibitory factor receptor alpha | **PRDM1** | PR domain containing 1, with ZNF domain |
| **HSP90AB1** | heat shock protein 90kDa alpha (cytosolic), class B member 1 | **TCF3** | transcription factor 3 |
| **MSH6** | mutS homolog 6 | **GNAQ** | guanine nucleotide binding protein (G protein), q polypeptide |
| **TBL1XR1** | transducin (beta)-like 1 X-linked receptor 1 | **PAX8** | paired box 8 |
| **BAP1** | BRCA1 associated protein-1 (ubiquitin carboxy-terminal hydrolase) | **PIK3R1** | phosphoinositide-3-kinase, regulatory subunit 1 (alpha) |
| **BCL11A** | B-cell CLL/lymphoma 11A (zinc finger protein) | **BRCA2** | breast cancer 2, early onset |
| **CASC5** | cancer susceptibility candidate 5 | **FGFR1** | fibroblast growth factor receptor 1 |
| **MET** | MET proto-oncogene, receptor tyrosine kinase | **SDC4** | syndecan 4 |
| **CEP89** | centrosomal protein 89kDa | **TBL1XR1** | transducin (beta)-like 1 X-linked receptor 1 |
| **TRIM33** | tripartite motif containing 33 | **FOXO3** | forkhead box O3 |
| **NOTCH1** | notch 1 | **CCND3** | cyclin D3 |
| **KIAA1549** | KIAA1549 | **FGFR4** | fibroblast growth factor receptor 4 |
| **DNM2** | dynamin 2 | **BCL7A** | B-cell CLL/lymphoma 7A |
| **CHN1** | chimerin 1 | **ZMYM2** | zinc finger, MYM-type 2 |
| **SETBP1** | SET binding protein 1 | **CCND1** | cyclin D1 |
| **WRN** | Werner syndrome, RecQ helicase-like | **JAK1** | Janus kinase 1 |
| **NR4A3** | nuclear receptor subfamily 4, group A, member 3 | **BAP1** | BRCA1 associated protein-1 (ubiquitin carboxy-terminal hydrolase) |
| **MLLT10** | myeloid/lymphoid or mixed-lineage leukemia (trithorax homolog, Drosophila); translocated to, 10 | **NAB2** | NGFI-A binding protein 2 (EGR1 binding protein 2) |
| **KDM5C** | lysine (K)-specific demethylase 5C | **LCK** | LCK proto-oncogene, Src family tyrosine kinase |
| **ETV6** | ets variant 6 | **CSF3R** | colony stimulating factor 3 receptor (granulocyte) |
| **ABL2** | ABL proto-oncogene 2, non-receptor tyrosine kinase | **HOXA11** | homeobox A11 |
| **RAD21** | RAD21 homolog (S. pombe) | **ZNF384** | zinc finger protein 384 |
| **PMS2** | PMS2 postmeiotic segregation increased 2 (S. cerevisiae) | **PPP2R1A** | protein phosphatase 2, regulatory subunit A, alpha |
| **XPC** | xeroderma pigmentosum, complementation group C | **SDHB** | succinate dehydrogenase complex, subunit B, iron sulfur (Ip) |
| **ERCC5** | excision repair cross-complementation group 5 | **BCR** | breakpoint cluster region |
| **PRDM1** | PR domain containing 1, with ZNF domain | **ERBB4** | erb-b2 receptor tyrosine kinase 4 |
| **FOXO4** | forkhead box O4 | **SFPQ** | splicing factor proline/glutamine-rich |
| **SPOP** | speckle-type POZ protein | **NTRK3** | neurotrophic tyrosine kinase, receptor, type 3 |
| **STAT3** | signal transducer and activator of transcription 3 (acute-phase response factor) | **SMARCB1** | SWI/SNF related, matrix associated, actin dependent regulator of chromatin, subfamily b, member 1 |
| **TFE3** | transcription factor binding to IGHM enhancer 3 | **TCEA1** | transcription elongation factor A (SII), 1 |
| **ITK** | IL2-inducible T-cell kinase | **MAP2K4** | mitogen-activated protein kinase kinase 4 |
| **NCOA2** | nuclear receptor coactivator 2 | **HOXC11** | homeobox C11 |
| **RBM10** | RNA binding motif protein 10 | **NCOR2** | nuclear receptor corepressor 2 |
| **ERCC3** | excision repair cross-complementation group 3 | **HSP90AA1** | heat shock protein 90kDa alpha (cytosolic), class A member 1 |
| **EPS15** | epidermal growth factor receptor pathway substrate 15 | **NUTM2A** | NUT family member 2A |
| **ZCCHC8** | zinc finger, CCHC domain containing 8 | **NRG1** | neuregulin 1 |
| **TSC1** | tuberous sclerosis 1 | **KMT2A** | lysine (K)-specific methyltransferase 2A |
| **ERC1** | ELKS/RAB6-interacting/CAST family member 1 | **NUTM1** | NUT midline carcinoma, family member 1 |
| **NFATC2** | nuclear factor of activated T-cells, cytoplasmic, calcineurin-dependent 2 | **EIF3E** | eukaryotic translation initiation factor 3, subunit E |
| **KDR** | kinase insert domain receptor | **HOXD13** | homeobox D13 |
| **DNMT3A** | DNA (cytosine-5-)-methyltransferase 3 alpha | **NFE2L2** | nuclear factor, erythroid 2-like 2 |
| **TRIM24** | tripartite motif containing 24 | **NRAS** | neuroblastoma RAS viral (v-ras) oncogene homolog |
| **PDGFRB** | platelet-derived growth factor receptor, beta polypeptide | **PDGFRA** | platelet-derived growth factor receptor, alpha polypeptide |
| **TPM3** | tropomyosin 3 | **DDX10** | DEAD (Asp-Glu-Ala-Asp) box polypeptide 10 |

| | | | |
|---|---|---|---|
| **FOXP1** | forkhead box P1 | **RSPO2** | R-spondin 2 |
| **TCF12** | transcription factor 12 | **RNF213** | ring finger protein 213 |
| **KIT** | v-kit Hardy-Zuckerman 4 feline sarcoma viral oncogene homolog | **ELN** | elastin |
| **CNOT3** | CCR4-NOT transcription complex, subunit 3 | **CXCR4** | chemokine (C-X-C motif) receptor 4 |
| **EXT2** | exostosin glycosyltransferase 2 | **CLTCL1** | clathrin, heavy chain-like 1 |
| **PCSK7** | proprotein convertase subtilisin/kexin type 7 | **BMPR1A** | bone morphogenetic protein receptor, type IA |
| **SS18** | synovial sarcoma translocation, chromosome 18 | **TET1** | tet methylcytosine dioxygenase 1 |
| **CARS** | cysteinyl-tRNA synthetase | **C2orf44** | chromosome 2 open reading frame 44 |
| **NUP214** | nucleoporin 214kDa | **RB1** | retinoblastoma 1 |
| **TSHR** | thyroid stimulating hormone receptor | **ASXL1** | additional sex combs like transcriptional regulator 1 |
| **BRD4** | bromodomain containing 4 | **MAML2** | mastermind-like 2 (Drosophila) |
| **PML** | promyelocytic leukemia | **NTRK1** | neurotrophic tyrosine kinase, receptor, type 1 |
| **JAK2** | Janus kinase 2 | **GMPS** | guanine monphosphate synthase |
| **FOXO3** | forkhead box O3 | **TNFAIP3** | tumor necrosis factor, alpha-induced protein 3 |
| **NTRK3** | neurotrophic tyrosine kinase, receptor, type 3 | **MYO5A** | myosin VA (heavy chain 12, myoxin) |
| **NCOR2** | nuclear receptor corepressor 2 | **SRSF3** | serine/arginine-rich splicing factor 3 |
| **RPL10** | ribosomal protein L10 | **KIAA1598** | KIAA1598 |
| **ARHGEF12** | Rho guanine nucleotide exchange factor (GEF) 12 | **NIN** | ninein (GSK3B interacting protein) |
| **ERCC2** | excision repair cross-complementation group 2 | **MYH11** | myosin, heavy chain 11, smooth muscle |
| **CCND2** | cyclin D2 | **SDHAF2** | succinate dehydrogenase complex assembly factor 2 |
| **USP6** | ubiquitin specific peptidase 6 | **RPL22** | ribosomal protein L22 |
| **THRAP3** | thyroid hormone receptor associated protein 3 | **TBX3** | T-box 3 |
| **MSN** | moesin | **CASC5** | cancer susceptibility candidate 5 |
| **AR** | androgen receptor | **HOXA13** | homeobox A13 |
| **MLLT3** | myeloid/lymphoid or mixed-lineage leukemia (trithorax homolog, Drosophila); translocated to, 3 | **KTN1** | kinectin 1 (kinesin receptor) |
| **PALB2** | partner and localizer of BRCA2 | **UBR5** | ubiquitin protein ligase E3 component n-recognin 5 |
| **EBF1** | early B-cell factor 1 | **MLF1** | myeloid leukemia factor 1 |
| **PER1** | period circadian clock 1 | **ELK4** | ELK4, ETS-domain protein (SRF accessory protein 1) |
| **HIP1** | huntingtin interacting protein 1 | **IL6ST** | interleukin 6 signal transducer |
| **PCM1** | pericentriolar material 1 | **MLLT1** | myeloid/lymphoid or mixed-lineage leukemia (trithorax homolog, Drosophila); translocated to, 1 |
| **RNF43** | ring finger protein 43 | **TLX3** | T-cell leukemia homeobox 3 |
| **RABEP1** | rabaptin, RAB GTPase binding effector protein 1 | **HLA-A** | major histocompatibility complex, class I, A |
| **RET** | ret proto-oncogene | **LMO1** | LIM domain only 1 (rhombotin 1) |
| **PMS1** | PMS1 postmeiotic segregation increased 1 (S. cerevisiae) | **STRN** | striatin, calmodulin binding protein |
| **sept-06** | septin 6 | **ATP1A1** | ATPase, Na+/K+ transporting, alpha 1 polypeptide |
| **PAX3** | paired box 3 | **SPOP** | speckle-type POZ protein |
| **MYB** | v-myb avian myeloblastosis viral oncogene homolog | **H3F3B** | H3 histone, family 3B (H3.3B) |
| **CHEK2** | checkpoint kinase 2 | **PHF6** | PHD finger protein 6 |
| **BCL11B** | B-cell CLL/lymphoma 11B (zinc finger protein) | **ESR1** | estrogen receptor 1 |
| **RAF1** | Raf-1 proto-oncogene, serine/threonine kinase | **STAT3** | signal transducer and activator of transcription 3 (acute-phase response factor) |
| **PSIP1** | PC4 and SFRS1 interacting protein 1 | **WWTR1** | WW domain containing transcription regulator 1 |
| **FANCA** | Fanconi anemia, complementation group A | **CCNB1IP1** | cyclin B1 interacting protein 1, E3 ubiquitin protein ligase |
| **WHSC1** | Wolf-Hirschhorn syndrome candidate 1 | **EXT1** | exostosin glycosyltransferase 1 |
| **IL7R** | interleukin 7 receptor | **SMARCA4** | SWI/SNF related, matrix associated, actin dependent regulator of chromatin, subfamily a, member 4 |
| **EWSR1** | EWS RNA-binding protein 1 | **MLLT11** | myeloid/lymphoid or mixed-lineage leukemia (trithorax homolog, Drosophila); translocated to, 11 |
| **HSP90AA1** | heat shock protein 90kDa alpha (cytosolic), class A member 1 | **ACVR1** | activin A receptor, type I |

| | | | |
|---|---|---|---|
| **IL6ST** | interleukin 6 signal transducer | **KDM6A** | lysine (K)-specific demethylase 6A |
| **RPL5** | ribosomal protein L5 | **DDX5** | DEAD (Asp-Glu-Ala-Asp) box helicase 5 |
| **PPFIBP1** | PTPRF interacting protein, binding protein 1 (liprin beta 1) | **RPL10** | ribosomal protein L10 |
| **PRRX1** | paired related homeobox 1 | **HOXD11** | homeobox D11 |
| **SMAD2** | SMAD family member 2 | **SLC45A3** | solute carrier family 45, member 3 |
| **MEN1** | multiple endocrine neoplasia I | **STK11** | serine/threonine kinase 11 |
| **KIF5B** | kinesin family member 5B | **GRIN2A** | glutamate receptor, ionotropic, N-methyl D-aspartate 2A |
| **COL2A1** | collagen, type II, alpha 1 | **AXIN1** | axin 1 |
| **MLH1** | mutL homolog 1 | **NCOR1** | nuclear receptor corepressor 1 |
| **TGFBR2** | transforming growth factor, beta receptor II (70/80kDa) | **RHOH** | ras homolog family member H |
| **WAS** | Wiskott-Aldrich syndrome | **ZNF331** | zinc finger protein 331 |
| **SPECC1** | sperm antigen with calponin homology and coiled-coil domains 1 | **NPM1** | nucleophosmin (nucleolar phosphoprotein B23, numatrin) |
| **MKL1** | megakaryoblastic leukemia (translocation) 1 | **JAZF1** | JAZF zinc finger 1 |
| **LPP** | LIM domain containing preferred translocation partner in lipoma | **sept-09** | septin 9 |
| **ABL1** | ABL proto-oncogene 1, non-receptor tyrosine kinase | **GPC3** | glypican 3 |
| **RALGDS** | ral guanine nucleotide dissociation stimulator | **ARID1B** | AT rich interactive domain 1B (SWI1-like) |
| **FGFR1OP** | FGFR1 oncogene partner | **PICALM** | phosphatidylinositol binding clathrin assembly protein |
| **PRF1** | perforin 1 (pore forming protein) | **TRIM33** | tripartite motif containing 33 |
| **NACA** | nascent polypeptide-associated complex alpha subunit | **HMGA1** | high mobility group AT-hook 1 |
| **HIF1A** | hypoxia inducible factor 1, alpha subunit (basic helix-loop-helix transcription factor) | **CANT1** | calcium activated nucleotidase 1 |
| **ELL** | elongation factor RNA polymerase II | **ARHGEF12** | Rho guanine nucleotide exchange factor (GEF) 12 |
| **FUS** | FUS RNA binding protein | **RECQL4** | RecQ protein-like 4 |
| **EML4** | echinoderm microtubule associated protein like 4 | **AFF1** | AF4/FMR2 family, member 1 |
| **CBL** | Cbl proto-oncogene, E3 ubiquitin protein ligase | **ZFHX3** | zinc finger homeobox 3 |
| **CIITA** | class II, major histocompatibility complex, transactivator | **TFPT** | TCF3 (E2A) fusion partner (in childhood Leukemia) |
| **WT1** | Wilms tumor 1 | **KMT2C** | lysine (K)-specific methyltransferase 2C |
| **PPP2R1A** | protein phosphatase 2, regulatory subunit A, alpha | **RPL5** | ribosomal protein L5 |
| **NRG1** | neuregulin 1 | **RAD51B** | RAD51 paralog B |
| **NFE2L2** | nuclear factor, erythroid 2-like 2 | **GATA3** | GATA binding protein 3 |
| **ARHGAP26** | Rho GTPase activating protein 26 | **NCOA1** | nuclear receptor coactivator 1 |
| **KRAS** | Kirsten rat sarcoma viral oncogene homolog | **OMD** | osteomodulin |
| **EXT1** | exostosin glycosyltransferase 1 | **KLK2** | kallikrein-related peptidase 2 |
| **RECQL4** | RecQ protein-like 4 | **CARD11** | caspase recruitment domain family, member 11 |
| **DDB2** | damage-specific DNA binding protein 2, 48kDa | **ERCC2** | excision repair cross-complementation group 2 |
| **FLI1** | Fli-1 proto-oncogene, ETS transcription factor | **NBN** | nibrin |
| **PRKAR1A** | protein kinase, cAMP-dependent, regulatory, type I, alpha | **CREBBP** | CREB binding protein |
| **ETV1** | ets variant 1 | **TRRAP** | transformation/transcription domain-associated protein |
| **ETV5** | ets variant 5 | **SH3GL1** | SH3-domain GRB2-like 1 |
| **CDH11** | cadherin 11, type 2, OB-cadherin (osteoblast) | **CCND2** | cyclin D2 |
| **SMO** | smoothened, frizzled class receptor | **SPEN** | spen family transcriptional repressor |
| **TCF7L2** | transcription factor 7-like 2 (T-cell specific, HMG-box) | **MAP3K1** | mitogen-activated protein kinase kinase kinase 1, E3 ubiquitin protein ligase |
| **LRIG3** | leucine-rich repeats and immunoglobulin-like domains 3 | **CDK12** | cyclin-dependent kinase 12 |
| **MSH2** | mutS homolog 2 | **KIAA1549** | KIAA1549 |
| **SRGAP3** | SLIT-ROBO Rho GTPase activating protein 3 | **DDX6** | DEAD (Asp-Glu-Ala-Asp) box helicase 6 |
| **ELF4** | E74-like factor 4 (ets domain transcription factor) | **SMAD4** | SMAD family member 4 |

| | | | |
|---|---|---|---|
| **HEY1** | hes-related family bHLH transcription factor with YRPW motif 1 | **CHIC2** | cysteine-rich hydrophobic domain 2 |
| **HOOK3** | hook microtubule-tethering protein 3 | **MYC** | v-myc avian myelocytomatosis viral oncogene homolog |
| **ERG** | v-ets avian erythroblastosis virus E26 oncogene homolog | **SND1** | staphylococcal nuclease and tudor domain containing 1 |
| **EZH2** | enhancer of zeste 2 polycomb repressive complex 2 subunit | **BIRC3** | baculoviral IAP repeat containing 3 |
| **PTPRK** | protein tyrosine phosphatase, receptor type, K | **USP6** | ubiquitin specific peptidase 6 |
| **AXIN2** | axin 2 | **NUP98** | nucleoporin 98kDa |
| **NFIB** | nuclear factor I/B | **RMI2** | RecQ mediated genome instability 2 |
| **ERCC4** | excision repair cross-complementation group 4 | **PDCD1LG2** | programmed cell death 1 ligand 2 |
| **PPARG** | peroxisome proliferator-activated receptor gamma | **THRAP3** | thyroid hormone receptor associated protein 3 |
| **CDC73** | cell division cycle 73 | **KDSR** | 3-ketodihydrosphingosine reductase |
| **BRAF** | B-Raf proto-oncogene, serine/threonine kinase | **PRRX1** | paired related homeobox 1 |
| **USP8** | ubiquitin specific peptidase 8 | **CASP8** | caspase 8, apoptosis-related cysteine peptidase |
| **FLT3** | fms-related tyrosine kinase 3 | **PRKAR1A** | protein kinase, cAMP-dependent, regulatory, type I, alpha |
| **TERT** | telomerase reverse transcriptase | **STAT6** | signal transducer and activator of transcription 6, interleukin-4 induced |
| **FUBP1** | far upstream element (FUSE) binding protein 1 | **PRDM16** | PR domain containing 16 |
| **NF2** | neurofibromin 2 (merlin) | **CHN1** | chimerin 1 |
| **CSF3R** | colony stimulating factor 3 receptor (granulocyte) | **BRIP1** | BRCA1 interacting protein C-terminal helicase 1 |
| **SLC34A2** | solute carrier family 34 (type II sodium/phosphate cotransporter), member 2 | **SET** | SET nuclear proto-oncogene |
| **SFPQ** | splicing factor proline/glutamine-rich | **IRF4** | interferon regulatory factor 4 |
| **NUTM1** | NUT midline carcinoma, family member 1 | **ARNT** | aryl hydrocarbon receptor nuclear translocator |
| **EIF3E** | eukaryotic translation initiation factor 3, subunit E | **ETV1** | ets variant 1 |
| **ELN** | elastin | **ATP2B3** | ATPase, Ca++ transporting, plasma membrane 3 |
| **CLTCL1** | clathrin, heavy chain-like 1 | **U2AF1** | U2 small nuclear RNA auxiliary factor 1 |
| **NTRK1** | neurotrophic tyrosine kinase, receptor, type 1 | **ETV5** | ets variant 5 |
| **ACSL6** | acyl-CoA synthetase long-chain family member 6 | **LMNA** | lamin A/C |
| **PAX5** | paired box 5 | **RANBP17** | RAN binding protein 17 |
| **SLC45A3** | solute carrier family 45, member 3 | **PAX7** | paired box 7 |
| **AXIN1** | axin 1 | **DEK** | DEK proto-oncogene |
| **PICALM** | phosphatidylinositol binding clathrin assembly protein | **NCOA2** | nuclear receptor coactivator 2 |
| **CANT1** | calcium activated nucleotidase 1 | **KAT6B** | K(lysine) acetyltransferase 6B |
| **NCOA1** | nuclear receptor coactivator 1 | **NFKB2** | nuclear factor of kappa light polypeptide gene enhancer in B-cells 2 (p49/p100) |
| **NBN** | nibrin | **GAS7** | growth arrest-specific 7 |
| **CTNNB1** | catenin (cadherin-associated protein), beta 1, 88kDa | **FSTL3** | follistatin-like 3 (secreted glycoprotein) |
| **NFKB2** | nuclear factor of kappa light polypeptide gene enhancer in B-cells 2 (p49/p100) | **CDKN2A** | cyclin-dependent kinase inhibitor 2A |
| **HLF** | hepatic leukemia factor | **MDS2** | myelodysplastic syndrome 2 translocation associated |
| **STAT5B** | signal transducer and activator of transcription 5B | **ATIC** | 5-aminoimidazole-4-carboxamide ribonucleotide formyltransferase/IMP cyclohydrolase |
| **STIL** | SCL/TAL1 interrupting locus | **CACNA1D** | calcium channel, voltage-dependent, L type, alpha 1D subunit |
| **HMGA2** | high mobility group AT-hook 2 | **CREB3L2** | cAMP responsive element binding protein 3-like 2 |
| **PRCC** | papillary renal cell carcinoma (translocation-associated) | **ABI1** | abl-interactor 1 |
| **FIP1L1** | factor interacting with PAPOLA and CPSF1 | **NCKIPSD** | NCK interacting protein with SH3 domain |
| **TCL6** | T-cell leukemia/lymphoma 6 (non-protein coding) | **ERCC3** | excision repair cross-complementation group 3 |
| **IKZF1** | IKAROS family zinc finger 1 (Ikaros) | **MLLT3** | myeloid/lymphoid or mixed-lineage leukemia (trithorax homolog, Drosophila); translocated to, 3 |
| **MALT1** | MALT1 paracaspase | **MNX1** | motor neuron and pancreas homeobox 1 |

| | | | |
|---|---|---|---|
| **SMAD3** | SMAD family member 3 | **MN1** | meningioma (disrupted in balanced translocation) 1 |
| **RBM15** | RNA binding motif protein 15 | **WHSC1L1** | Wolf-Hirschhorn syndrome candidate 1-like 1 |
| **sept-05** | septin 5 | **BCL2** | B-cell CLL/lymphoma 2 |
| **LZTR1** | leucine-zipper-like transcription regulator 1 | **SS18L1** | synovial sarcoma translocation gene on chromosome 18-like 1 |
| **ZBTB16** | zinc finger and BTB domain containing 16 | **ZCCHC8** | zinc finger, CCHC domain containing 8 |
| **TFRC** | transferrin receptor | **WRN** | Werner syndrome, RecQ helicase-like |
| **SUZ12** | SUZ12 polycomb repressive complex 2 subunit | **TSC1** | tuberous sclerosis 1 |
| **LSM14A** | LSM14A, SCD6 homolog A (S. cerevisiae) | **PALB2** | partner and localizer of BRCA2 |
| **TNFRSF14** | tumor necrosis factor receptor superfamily, member 14 | **POLE** | polymerase (DNA directed), epsilon, catalytic subunit |
| **DAXX** | death-domain associated protein | **PHOX2B** | paired-like homeobox 2b |
| **FGFR4** | fibroblast growth factor receptor 4 | **PDE4DIP** | phosphodiesterase 4D interacting protein |
| **LCK** | LCK proto-oncogene, Src family tyrosine kinase | **KIF5B** | kinesin family member 5B |
| **HOXA11** | homeobox A11 | **SDHA** | succinate dehydrogenase complex, subunit A, flavoprotein (Fp) |
| **ZNF384** | zinc finger protein 384 | **GPHN** | gephyrin |
| **AKT2** | v-akt murine thymoma viral oncogene homolog 2 | **AP002884.2** | AP002884.2 |
| **FBXO11** | F-box protein 11 | **FLT4** | fms-related tyrosine kinase 4 |
| **RPN1** | ribophorin I | **SH2B3** | SH2B adaptor protein 3 |
| **ETNK1** | ethanolamine kinase 1 | **TCF7L2** | transcription factor 7-like 2 (T-cell specific, HMG-box) |
| **GPC3** | glypican 3 | **FNBP1** | formin binding protein 1 |
| **RAD51B** | RAD51 paralog B | **CRTC1** | CREB regulated transcription coactivator 1 |
| **DDX6** | DEAD (Asp-Glu-Ala-Asp) box helicase 6 | **PER1** | period circadian clock 1 |
| **SND1** | staphylococcal nuclease and tudor domain containing 1 | **SMARCE1** | SWI/SNF related, matrix associated, actin dependent regulator of chromatin, subfamily e, member 1 |
| **ARNT** | aryl hydrocarbon receptor nuclear translocator | **HIP1** | huntingtin interacting protein 1 |
| **PHOX2B** | paired-like homeobox 2b | **PLAG1** | pleiomorphic adenoma gene 1 |
| **FLT4** | fms-related tyrosine kinase 4 | **BLM** | Bloom syndrome, RecQ helicase-like |
| **GATA2** | GATA binding protein 2 | **IDH2** | isocitrate dehydrogenase 2 (NADP+), mitochondrial |
| **CIC** | capicua transcriptional repressor | **NF1** | neurofibromin 1 |
| **MITF** | microphthalmia-associated transcription factor | **MLLT4** | myeloid/lymphoid or mixed-lineage leukemia (trithorax homolog, Drosophila); translocated to, 4 |
| **MAP2K1** | mitogen-activated protein kinase kinase 1 | **JAK3** | Janus kinase 3 |
| **HNRNPA2B1** | heterogeneous nuclear ribonucleoprotein A2/B1 | **CLP1** | cleavage and polyadenylation factor I subunit 1 |
| **BRD3** | bromodomain containing 3 | **STAT5B** | signal transducer and activator of transcription 5B |
| **PRKACA** | protein kinase, cAMP-dependent, catalytic, alpha | **CIC** | capicua transcriptional repressor |
| **RHOA** | ras homolog family member A | **FOXO1** | forkhead box O1 |
| **IDH1** | isocitrate dehydrogenase 1 (NADP+), soluble | **KAT6A** | K(lysine) acetyltransferase 6A |
| **RARA** | retinoic acid receptor, alpha | **PCM1** | pericentriolar material 1 |
| **TRAF7** | TNF receptor-associated factor 7, E3 ubiquitin protein ligase | **NFKBIE** | nuclear factor of kappa light polypeptide gene enhancer in B-cells inhibitor, epsilon |
| **TOP1** | topoisomerase (DNA) I | **MUTYH** | mutY homolog |
| **NDRG1** | N-myc downstream regulated 1 | **RANBP2** | RAN binding protein 2 |
| **MYCL** | v-myc avian myelocytomatosis viral oncogene lung carcinoma derived homolog | **MITF** | microphthalmia-associated transcription factor |
| **CCNE1** | cyclin E1 | **FEV** | FEV (ETS oncogene family) |
| **GNA11** | guanine nucleotide binding protein (G protein), alpha 11 (Gq class) | **ERC1** | ELKS/RAB6-interacting/CAST family member 1 |
| **SSX1** | synovial sarcoma, X breakpoint 1 | **CD79B** | CD79b molecule, immunoglobulin-associated beta |
| **EZR** | ezrin | **RUNX1** | runt-related transcription factor 1 |
| **MUC1** | mucin 1, cell surface associated | **BCL6** | B-cell CLL/lymphoma 6 |
| **HNF1A** | HNF1 homeobox A | **RNF43** | ring finger protein 43 |
| **SRSF2** | serine/arginine-rich splicing factor 2 | **SUFU** | suppressor of fused homolog (Drosophila) |

| | | | |
|---|---|---|---|
| **ECT2L** | epithelial cell transforming 2 like | **ALDH2** | aldehyde dehydrogenase 2 family (mitochondrial) |
| **PAX8** | paired box 8 | **EIF4A2** | eukaryotic translation initiation factor 4A2 |
| **DDX10** | DEAD (Asp-Glu-Ala-Asp) box polypeptide 10 | **RABEP1** | rabaptin, RAB GTPase binding effector protein 1 |
| **KIAA1598** | KIAA1598 | **CRTC3** | CREB regulated transcription coactivator 3 |
| **FAM46C** | family with sequence similarity 46, member C | **MAP2K1** | mitogen-activated protein kinase kinase 1 |
| **KLK2** | kallikrein-related peptidase 2 | **FCGR2B** | Fc fragment of IgG, low affinity IIb, receptor (CD32) |
| **PRDM16** | PR domain containing 16 | **STIL** | SCL/TAL1 interrupting locus |
| **CREB3L1** | cAMP responsive element binding protein 3-like 1 | **JUN** | jun proto-oncogene |
| **KLF4** | Kruppel-like factor 4 (gut) | **COL2A1** | collagen, type II, alpha 1 |
| **GAS7** | growth arrest-specific 7 | **PLCG1** | phospholipase C, gamma 1 |
| **GOPC** | golgi-associated PDZ and coiled-coil motif containing | **TFEB** | transcription factor EB |
| **TPM4** | tropomyosin 4 | **PTPN13** | protein tyrosine phosphatase, non-receptor type 13 (APO-1/CD95 (Fas)-associated phosphatase) |
| **PLAG1** | pleiomorphic adenoma gene 1 | **PRCC** | papillary renal cell carcinoma (translocation-associated) |
| **CD79B** | CD79b molecule, immunoglobulin-associated beta | **HNRNPA2B1** | heterogeneous nuclear ribonucleoprotein A2/B1 |
| **FCGR2B** | Fc fragment of IgG, low affinity IIb, receptor (CD32) | **RET** | ret proto-oncogene |
| **FANCC** | Fanconi anemia, complementation group C | **HEY1** | hes-related family bHLH transcription factor with YRPW motif 1 |
| **FAM131B** | family with sequence similarity 131, member B | **ATF1** | activating transcription factor 1 |
| **HRAS** | Harvey rat sarcoma viral oncogene homolog | **PMS1** | PMS1 postmeiotic segregation increased 1 (S. cerevisiae) |
| **FCRL4** | Fc receptor-like 4 | **BRD3** | bromodomain containing 3 |
| **SMARCD1** | SWI/SNF related, matrix associated, actin dependent regulator of chromatin, subfamily d, member 1 | **KDM5A** | lysine (K)-specific demethylase 5A |
| **POU5F1** | POU class 5 homeobox 1 | **HOOK3** | hook microtubule-tethering protein 3 |
| **REL** | v-rel avian reticuloendotheliosis viral oncogene homolog | **ERG** | v-ets avian erythroblastosis virus E26 oncogene homolog |
| **PTPN11** | protein tyrosine phosphatase, non-receptor type 11 | **KDR** | kinase insert domain receptor |
| **ACKR3** | atypical chemokine receptor 3 | **MDM4** | MDM4, p53 regulator |
| **FH** | fumarate hydratase | **LHFP** | lipoma HMGIC fusion partner |
| **HOXC13** | homeobox C13 | **ALK** | anaplastic lymphoma receptor tyrosine kinase |
| **MAX** | MYC associated factor X | **FIP1L1** | factor interacting with PAPOLA and CPSF1 |
| **PWWP2A** | PWWP domain containing 2A | **DNMT3A** | DNA (cytosine-5-)-methyltransferase 3 alpha |
| **PIM1** | Pim-1 proto-oncogene, serine/threonine kinase | **LASP1** | LIM and SH3 protein 1 |
| **MAFB** | v-maf avian musculoaponeurotic fibrosarcoma oncogene homolog B | **BCL9** | B-cell CLL/lymphoma 9 |
| **ASPSCR1** | alveolar soft part sarcoma chromosome region, candidate 1 | **FAM131B** | family with sequence similarity 131, member B |
| **LCP1** | lymphocyte cytosolic protein 1 (L-plastin) | **PATZ1** | POZ (BTB) and AT hook containing zinc finger 1 |
| **TCF3** | transcription factor 3 | **TRIM24** | tripartite motif containing 24 |
| **CCND3** | cyclin D3 | **HRAS** | Harvey rat sarcoma viral oncogene homolog |
| **NAB2** | NGFI-A binding protein 2 (EGR1 binding protein 2) | **EZH2** | enhancer of zeste 2 polycomb repressive complex 2 subunit |
| **TCEA1** | transcription elongation factor A (SII), 1 | **RHOA** | ras homolog family member A |
| **CXCR4** | chemokine (C-X-C motif) receptor 4 | **SPECC1** | sperm antigen with calponin homology and coiled-coil domains 1 |
| **STRN** | striatin, calmodulin binding protein | **SYK** | spleen tyrosine kinase |
| **CCNB1IP1** | cyclin B1 interacting protein 1, E3 ubiquitin protein ligase | **RUNX1T1** | runt-related transcription factor 1; translocated to, 1 (cyclin D-related) |
| **NONO** | non-POU domain containing, octamer-binding | **PAX3** | paired box 3 |
| **ZNF331** | zinc finger protein 331 | **MKL1** | megakaryoblastic leukemia (translocation) 1 |
| **SEPT09** | septin 9 | **SOX2** | SRY (sex determining region Y)-box 2 |
| **FANCG** | Fanconi anemia, complementation group G | **MTCP1** | mature T-cell proliferation 1 |
| **PAX7** | paired box 7 | **CUX1** | cut-like homeobox 1 |

| | | | |
|---|---|---|---|
| **GOLGA5** | golgin A5 | **TPM3** | tropomyosin 3 |
| **CREB3L2** | cAMP responsive element binding protein 3-like 2 | **FCRL4** | Fc receptor-like 4 |
| **ABI1** | abl-interactor 1 | **AXIN2** | axin 2 |
| **NCKIPSD** | NCK interacting protein with SH3 domain | **RAP1GDS1** | RAP1, GTP-GDP dissociation stimulator 1 |
| **GPHN** | gephyrin | **FOXP1** | forkhead box P1 |
| **FNBP1** | formin binding protein 1 | **TRIM27** | tripartite motif containing 27 |
| **BCL6** | B-cell CLL/lymphoma 6 | **PBX1** | pre-B-cell leukemia homeobox 1 |
| **JUN** | jun proto-oncogene | **BCL10** | B-cell CLL/lymphoma 10 |
| **LASP1** | LIM and SH3 protein 1 | **FANCE** | Fanconi anemia, complementation group E |
| **SYK** | spleen tyrosine kinase | **TCF12** | transcription factor 12 |
| **TFG** | TRK-fused gene | **LEF1** | lymphoid enhancer-binding factor 1 |
| **RAP1GDS1** | RAP1, GTP-GDP dissociation stimulator 1 | **SDHC** | succinate dehydrogenase complex, subunit C, integral membrane protein, 15kDa |
| **BCL10** | B-cell CLL/lymphoma 10 | **POU2AF1** | POU class 2 associating factor 1 |
| **TTL** | tubulin tyrosine ligase | **CDK6** | cyclin-dependent kinase 6 |
| **BUB1B** | BUB1 mitotic checkpoint serine/threonine kinase B | **C15orf65** | chromosome 15 open reading frame 65 |
| **MSI2** | musashi RNA-binding protein 2 | **AFF3** | AF4/FMR2 family, member 3 |
| **CREB1** | cAMP responsive element binding protein 1 | **NFIB** | nuclear factor I/B |
| **MAP2K2** | mitogen-activated protein kinase kinase 2 | **KIT** | v-kit Hardy-Zuckerman 4 feline sarcoma viral oncogene homolog |
| **PPP6C** | protein phosphatase 6, catalytic subunit | **IKBKB** | inhibitor of kappa light polypeptide gene enhancer in B-cells, kinase beta |
| **FANCF** | Fanconi anemia, complementation group F | **LPP** | LIM domain containing preferred translocation partner in lipoma |
| **MDM2** | MDM2 proto-oncogene, E3 ubiquitin protein ligase | **ABL1** | ABL proto-oncogene 1, non-receptor tyrosine kinase |
| **PDGFB** | platelet-derived growth factor beta polypeptide | **COL1A1** | collagen, type I, alpha 1 |
| **CD274** | CD274 molecule | **CEBPA** | CCAAT/enhancer binding protein (C/EBP), alpha |
| **MYD88** | myeloid differentiation primary response 88 | **IDH1** | isocitrate dehydrogenase 1 (NADP+), soluble |
| **CCDC6** | coiled-coil domain containing 6 | **SMARCD1** | SWI/SNF related, matrix associated, actin dependent regulator of chromatin, subfamily d, member 1 |
| **BCL7A** | B-cell CLL/lymphoma 7A | **RARA** | retinoic acid receptor, alpha |
| **CD79A** | CD79a molecule, immunoglobulin-associated alpha | **KMT2D** | lysine (K)-specific methyltransferase 2D |
| **POT1** | protection of telomeres 1 | **POU5F1** | POU class 5 homeobox 1 |
| **NUTM2A** | NUT family member 2A | **H3F3A** | H3 histone, family 3A |
| **RSPO2** | R-spondin 2 | **PBRM1** | polybromo 1 |
| **GMPS** | guanine monphosphate synthase | **SETD2** | SET domain containing 2 |
| **SRSF3** | serine/arginine-rich splicing factor 3 | **CHCHD7** | coiled-coil-helix-coiled-coil-helix domain containing 7 |
| **HOXA13** | homeobox A13 | **ACSL3** | acyl-CoA synthetase long-chain family member 3 |
| **MLF1** | myeloid leukemia factor 1 | **ETV4** | ets variant 4 |
| **H3F3B** | H3 histone, family 3B (H3.3B) | **MYB** | v-myb avian myeloblastosis viral oncogene homolog |
| **BCL3** | B-cell CLL/lymphoma 3 | **TTL** | tubulin tyrosine ligase |
| **WWTR1** | WW domain containing transcription regulator 1 | **YWHAE** | tyrosine 3-monooxygenase/tryptophan 5-monooxygenase activation protein, epsilon |
| **HERPUD1** | homocysteine-inducible, endoplasmic reticulum stress-inducible, ubiquitin-like domain member 1 | **MPL** | MPL proto-oncogene, thrombopoietin receptor |
| **RHOH** | ras homolog family member H | **MAF** | v-maf avian musculoaponeurotic fibrosarcoma oncogene homolog |
| **TFPT** | TCF3 (E2A) fusion partner (in childhood Leukemia) | **ERCC4** | excision repair cross-complementation group 4 |
| **OMD** | osteomodulin | **BUB1B** | BUB1 mitotic checkpoint serine/threonine kinase B |
| **MYC** | v-myc avian myelocytomatosis viral oncogene homolog | **EP300** | E1A binding protein p300 |
| **SET** | SET nuclear proto-oncogene | **PTK6** | protein tyrosine kinase 6 |
| **LMNA** | lamin A/C | **CNOT3** | CCR4-NOT transcription complex, subunit 3 |

| | | | |
|---|---|---|---|
| **DEK** | DEK proto-oncogene | **TOP1** | topoisomerase (DNA) I |
| **ATIC** | 5-aminoimidazole-4-carboxamide ribonucleotide formyltransferase/IMP cyclohydrolase | | |
| **MN1** | meningioma (disrupted in balanced translocation) 1 | | |
| **BCL2** | B-cell CLL/lymphoma 2 | | |
| **WIF1** | WNT inhibitory factor 1 | | |
| **SDHA** | succinate dehydrogenase complex, subunit A, flavoprotein (Fp) | | |
| **SH2B3** | SH2B adaptor protein 3 | | |
| **SMARCE1** | SWI/SNF related, matrix associated, actin dependent regulator of chromatin, subfamily e, member 1 | | |
| **SUFU** | suppressor of fused homolog (Drosophila) | | |
| **ATF1** | activating transcription factor 1 | | |
| **MDM4** | MDM4, p53 regulator | | |
| **SOX2** | SRY (sex determining region Y)-box 2 | | |
| **SDHC** | succinate dehydrogenase complex, subunit C, integral membrane protein, 15kDa | | |
| **H3F3A** | H3 histone, family 3A | | |
| **ACSL3** | acyl-CoA synthetase long-chain family member 3 | | |
| **MAF** | v-maf avian musculoaponeurotic fibrosarcoma oncogene homolog | | |
| **FLCN** | folliculin | | |
| **KCNJ5** | potassium channel, inwardly rectifying subfamily J, member 5 | | |
| **SBDS** | Shwachman-Bodian-Diamond syndrome | | |
| **TAL1** | T-cell acute lymphocytic leukemia 1 | | |
| **MYCN** | v-myc avian myelocytomatosis viral oncogene neuroblastoma derived homolog | | |
| **DDIT3** | DNA-damage-inducible transcript 3 | | |
| **TNFRSF17** | tumor necrosis factor receptor superfamily, member 17 | | |
| **SDC4** | syndecan 4 | | |
| **SDHB** | succinate dehydrogenase complex, subunit B, iron sulfur (Ip) | | |
| **SMARCB1** | SWI/SNF related, matrix associated, actin dependent regulator of chromatin, subfamily b, member 1 | | |
| **HOXD13** | homeobox D13 | | |
| **BMPR1A** | bone morphogenetic protein receptor, type IA | | |
| **P2RY8** | purinergic receptor P2Y, G-protein coupled, 8 | | |
| **RPL22** | ribosomal protein L22 | | |
| **PHF6** | PHD finger protein 6 | | |
| **STK11** | serine/threonine kinase 11 | | |
| **DNAJB1** | DnaJ (Hsp40) homolog, subfamily B, member 1 | | |
| **CHIC2** | cysteine-rich hydrophobic domain 2 | | |
| **BIRC3** | baculoviral IAP repeat containing 3 | | |
| **GATA1** | GATA binding protein 1 (globin transcription factor 1) | | |
| **IRF4** | interferon regulatory factor 4 | | |
| **SS18L1** | synovial sarcoma translocation gene on chromosome 18-like 1 | | |
| **MUTYH** | mutY homolog | | |
| **CRTC3** | CREB regulated transcription coactivator 3 | | |
| **TFEB** | transcription factor EB | | |
| **LHFP** | lipoma HMGIC fusion partner | | |
| **PATZ1** | POZ (BTB) and AT hook containing zinc finger 1 | | |

| | |
|---|---|
| **PBX1** | pre-B-cell leukemia homeobox 1 |
| **FANCE** | Fanconi anemia, complementation group E |
| **LEF1** | lymphoid enhancer-binding factor 1 |
| **CDK6** | cyclin-dependent kinase 6 |
| **C15orf65** | chromosome 15 open reading frame 65 |
| **MPL** | MPL proto-oncogene, thrombopoietin receptor |
| **PTK6** | protein tyrosine kinase 6 |
| **TAL2** | T-cell acute lymphocytic leukemia 2 |
| **LYL1** | lymphoblastic leukemia associated hematopoiesis regulator 1 |
| **CDX2** | caudal type homeobox 2 |
| **FAS** | Fas cell surface death receptor |
| **RAC1** | ras-related C3 botulinum toxin substrate 1 (rho family, small GTP binding protein Rac1) |
| **FHIT** | fragile histidine triad |
| **KLF6** | Kruppel-like factor 6 |
| **FGFR3** | fibroblast growth factor receptor 3 |
| **HOXA9** | homeobox A9 |
| **GNAQ** | guanine nucleotide binding protein (G protein), q polypeptide |
| **CCND1** | cyclin D1 |
| **CALR** | calreticulin |
| **HOXC11** | homeobox C11 |
| **NRAS** | neuroblastoma RAS viral (v-ras) oncogene homolog |
| **IL21R** | interleukin 21 receptor |
| **C2orf44** | chromosome 2 open reading frame 44 |
| **SDHAF2** | succinate dehydrogenase complex assembly factor 2 |
| **ELK4** | ELK4, ETS-domain protein (SRF accessory protein 1) |
| **LMO2** | LIM domain only 2 (rhombotin-like 1) |
| **MLLT1** | myeloid/lymphoid or mixed-lineage leukemia (trithorax homolog, Drosophila); translocated to, 1 |
| **HLA-A** | major histocompatibility complex, class I, A |
| **LMO1** | LIM domain only 1 (rhombotin 1) |
| **ACVR1** | activin A receptor, type I |
| **NPM1** | nucleophosmin (nucleolar phosphoprotein B23, numatrin) |
| **HMGA1** | high mobility group AT-hook 1 |
| **SH3GL1** | SH3-domain GRB2-like 1 |
| **MDS2** | myelodysplastic syndrome 2 translocation associated |
| **MNX1** | motor neuron and pancreas homeobox 1 |
| **FOXO1** | forkhead box O1 |
| **NFKBIE** | nuclear factor of kappa light polypeptide gene enhancer in B-cells inhibitor, epsilon |
| **FEV** | FEV (ETS oncogene family) |
| **IL2** | interleukin 2 |
| **RSPO3** | R-spondin 3 |
| **MTCP1** | mature T-cell proliferation 1 |
| **TRIM27** | tripartite motif containing 27 |
| **POU2AF1** | POU class 2 associating factor 1 |
| **CBLC** | Cbl proto-oncogene C, E3 ubiquitin protein ligase |
| **CHCHD7** | coiled-coil-helix-coiled-coil-helix domain containing 7 |

| | |
|---|---|
| **HMGN2P46** | high mobility group nucleosomal binding domain 2 pseudogene 46 |
| **SOCS1** | suppressor of cytokine signaling 1 |
| **COX6C** | cytochrome c oxidase subunit VIc |
| **VTI1A** | vesicle transport through interaction with t-SNAREs 1A |
| **NKX2-1** | NK2 homeobox 1 |
| **FOXL2** | forkhead box L2 |
| **PAFAH1B2** | platelet-activating factor acetylhydrolase 1b, catalytic subunit 2 (30kDa) |
| **SDHD** | succinate dehydrogenase complex, subunit D, integral membrane protein |
| **TMPRSS2** | transmembrane protease, serine 2 |
| **CBFA2T3** | core-binding factor, runt domain, alpha subunit 2; translocated to, 3 |
| **CD74** | CD74 molecule, major histocompatibility complex, class II invariant chain |
| **CDK4** | cyclin-dependent kinase 4 |
| **CNBP** | CCHC-type zinc finger, nucleic acid binding protein |
| **TLX3** | T-cell leukemia homeobox 3 |
| **HOXD11** | homeobox D11 |
| **JAZF1** | JAZF zinc finger 1 |
| **CDKN2C** | cyclin-dependent kinase inhibitor 2C (p18, inhibits CDK4) |
| **BTG1** | B-cell translocation gene 1, anti-proliferative |
| **PDCD1LG2** | programmed cell death 1 ligand 2 |
| **KDSR** | 3-ketodihydrosphingosine reductase |
| **SSX2** | synovial sarcoma, X breakpoint 2 |
| **U2AF1** | U2 small nuclear RNA auxiliary factor 1 |
| **NT5C2** | 5'-nucleotidase, cytosolic II |
| **ZRSR2** | zinc finger (CCCH type), RNA-binding motif and serine/arginine rich 2 |
| **CRTC1** | CREB regulated transcription coactivator 1 |
| **IDH2** | isocitrate dehydrogenase 2 (NADP+), mitochondrial |
| **CLP1** | cleavage and polyadenylation factor I subunit 1 |
| **ALDH2** | aldehyde dehydrogenase 2 family (mitochondrial) |
| **NUTM2B** | NUT family member 2B |
| **CEBPA** | CCAAT/enhancer binding protein (C/EBP), alpha |
| **YWHAE** | tyrosine 3-monooxygenase/tryptophan 5-monooxygenase activation protein, epsilon |
| **VHL** | von Hippel-Lindau tumor suppressor, E3 ubiquitin protein ligase |
| **OLIG2** | oligodendrocyte lineage transcription factor 2 |
| **MYOD1** | myogenic differentiation 1 |
| **MLLT11** | myeloid/lymphoid or mixed-lineage leukemia (trithorax homolog, Drosophila); translocated to, 11 |
| **RMI2** | RecQ mediated genome instability 2 |
| **FSTL3** | follistatin-like 3 (secreted glycoprotein) |
| **CDKN2A** | cyclin-dependent kinase inhibitor 2A |
| **XPA** | xeroderma pigmentosum, complementation group A |
| **AP002884.2** | AP002884.2 |
| **TLX1** | T-cell leukemia homeobox 1 |
| **TCL1A** | T-cell leukemia/lymphoma 1A |
| **CRLF2** | cytokine receptor-like factor 2 |
| **ETV4** | ets variant 4 |

Supplemental material 2

| Gene Symbol | Panel BC 360 -Official Full Name |
|---|---|
| **ABCA8** | ATP binding cassette subfamily A member 8 |
| **ACTR3B** | ARP3 actin related protein 3 homolog B |
| **ACVR1B** | activin A receptor type 1B |
| **ACVR1C** | activin A receptor type 1C |
| **ACVRL1** | activin A receptor like type 1 |
| **ADAM12** | ADAM metallopeptidase domain 12 |
| **ADCY9** | adenylate cyclase 9 |
| **ADD1** | adducin 1 |
| **ADM** | adrenomedullin |
| **AGR2** | anterior gradient 2, protein disulphide isomerase family member |
| **AGT** | angiotensinogen |
| **AGTR1** | angiotensin II receptor type 1 |
| **AKT3** | AKT serine/threonine kinase 3 |
| **ALDH1A1** | aldehyde dehydrogenase 1 family member A1 |
| **ALDOA** | aldolase, fructose-bisphosphate A |
| **ANGPT1** | angiopoietin 1 |
| **ANLN** | anillin actin binding protein |
| **ANXA9** | annexin A9 |
| **APH1B** | aph-1 homolog B, gamma-secretase subunit |
| **APOD** | apolipoprotein D |
| **APOE** | apolipoprotein E |
| **AR** | androgen receptor |
| **AREG** | amphiregulin |
| **ARID1A** | AT-rich interaction domain 1A |
| **ARNT2** | aryl hydrocarbon receptor nuclear translocator 2 |
| **ASPM** | abnormal spindle microtubule assembly |
| **ASPN** | asporin |
| **ATAD2** | ATPase family, AAA domain containing 2 |
| **ATM** | ATM serine/threonine kinase |
| **ATP10B** | ATPase phospholipid transporting 10B (putative) |
| **AURKA** | aurora kinase A |
| **AURKB** | aurora kinase B |
| **AXIN1** | axin 1 |
| **AXIN2** | axin 2 |
| **B3GNT3** | UDP-GlcNAc:betaGal beta-1,3-N-acetylglucosaminyltransferase 3 |
| **BAD** | BCL2 associated agonist of cell death |
| **BAG1** | BCL2 associated athanogene 1 |
| **BAIAP2L1** | BAI1 associated protein 2 like 1 |
| **BAIAP3** | BAI1 associated protein 3 |
| **BAMBI** | BMP and activin membrane bound inhibitor |
| **BAX** | BCL2 associated X, apoptosis regulator |
| **BBC3** | BCL2 binding component 3 |
| **BBOX1** | gamma-butyrobetaine hydroxylase 1 |
| **BCAS1** | breast carcinoma amplified sequence 1 |
| **BCL11A** | B-cell CLL/lymphoma 11A |
| **BCL2** | BCL2, apoptosis regulator |
| **BCL2A1** | BCL2 related protein A1 |
| **BCL2L1** | BCL2 like 1 |
| **BCL6B** | B-cell CLL/lymphoma 6B |
| **BDNF** | brain derived neurotrophic factor |
| **BIRC5** | baculoviral IAP repeat containing 5 |
| **BLM** | Bloom syndrome RecQ like helicase |
| **BLVRA** | biliverdin reductase A |
| **BMP2** | bone morphogenetic protein 2 |
| **BMP4** | bone morphogenetic protein 4 |
| **BMP5** | bone morphogenetic protein 5 |
| **BMP6** | bone morphogenetic protein 6 |
| **BMP7** | bone morphogenetic protein 7 |
| **BMP8A** | bone morphogenetic protein 8a |
| **BMPR1A** | bone morphogenetic protein receptor type 1A |
| **BMPR1B** | bone morphogenetic protein receptor type 1B |
| **BMPR2** | bone morphogenetic protein receptor type 2 |
| **BNC2** | basonuclin 2 |
| **BNIP3** | BCL2 interacting protein 3 |
| **BORCS7** | BLOC-1 related complex subunit 7 |
| **BRCA1** | BRCA1, DNA repair associated |
| **BRCA2** | BRCA2, DNA repair associated |
| **BTG2** | BTG anti-proliferation factor 2 |
| **C5orf38** | chromosome 5 open reading frame 38 |
| **CA12** | carbonic anhydrase 12 |
| **CACNA1D** | calcium voltage-gated channel subunit alpha1 D |
| **CACNA1H** | calcium voltage-gated channel subunit alpha1 H |
| **CACNA2D1** | calcium voltage-gated channel auxiliary subunit alpha2delta 1 |
| **CACNA2D3** | calcium voltage-gated channel auxiliary subunit alpha2delta 3 |

| | |
|---|---|
| **CACNG1** | calcium voltage-gated channel auxiliary subunit gamma 1 |
| **CACNG4** | calcium voltage-gated channel auxiliary subunit gamma 4 |
| **CACNG6** | calcium voltage-gated channel auxiliary subunit gamma 6 |
| **CALML5** | calmodulin like 5 |
| **CAMK2B** | calcium/calmodulin dependent protein kinase II beta |
| **CAV1** | caveolin 1 |
| **CBLC** | Cbl proto-oncogene C |
| **CCL2** | C-C motif chemokine ligand 2 |
| **CCL21** | C-C motif chemokine ligand 21 |
| **CCL3L1** | C-C motif chemokine ligand 3 like 1 |
| **CCL4** | C-C motif chemokine ligand 4 |
| **CCL5** | C-C motif chemokine ligand 5 |
| **CCL7** | C-C motif chemokine ligand 7 |
| **CCL8** | C-C motif chemokine ligand 8 |
| **CCNA1** | cyclin A1 |
| **CCNA2** | cyclin A2 |
| **CCNB1** | cyclin B1 |
| **CCND1** | cyclin D1 |
| **CCND2** | cyclin D2 |
| **CCNE1** | cyclin E1 |
| **CCNE2** | cyclin E2 |
| **CCR1** | C-C motif chemokine receptor 1 |
| **CCR2** | C-C motif chemokine receptor 2 |
| **CCR5** | C-C motif chemokine receptor 5 (gene/pseudogene) |
| **CD163** | CD163 molecule |
| **CD19** | CD19 molecule |
| **CD1E** | CD1e molecule |
| **CD24** | CD24 molecule |
| **CD27** | CD27 molecule |
| **CD274** | CD274 molecule |
| **CD276** | CD276 molecule |
| **CD34** | CD34 molecule |
| **CD36** | CD36 molecule |
| **CD44** | CD44 molecule (Indian blood group) |
| **CD68** | CD68 molecule |
| **CD84** | CD84 molecule |
| **CD8A** | CD8a molecule |
| **CD8B** | CD8b molecule |
| **CDC14A** | cell division cycle 14A |
| **CDC14B** | cell division cycle 14B |
| **CDC20** | cell division cycle 20 |
| **CDC25A** | cell division cycle 25A |
| **CDC25B** | cell division cycle 25B |
| **CDC25C** | cell division cycle 25C |
| **CDC6** | cell division cycle 6 |
| **CDC7** | cell division cycle 7 |
| **CDCA5** | cell division cycle associated 5 |
| **CDCA7L** | cell division cycle associated 7 like |
| **CDCA8** | cell division cycle associated 8 |
| **CDH1** | cadherin 1 |
| **CDH2** | cadherin 2 |
| **CDH3** | cadherin 3 |
| **CDH5** | cadherin 5 |
| **CDK1** | cyclin dependent kinase 1 |
| **CDK4** | cyclin dependent kinase 4 |
| **CDK6** | cyclin dependent kinase 6 |
| **CDKN1A** | cyclin dependent kinase inhibitor 1A |
| **CDKN1B** | cyclin dependent kinase inhibitor 1B |
| **CDKN1C** | cyclin dependent kinase inhibitor 1C |
| **CDKN2A** | cyclin dependent kinase inhibitor 2A |
| **CDKN2B** | cyclin dependent kinase inhibitor 2B |
| **CDKN2C** | cyclin dependent kinase inhibitor 2C |
| **CDKN2D** | cyclin dependent kinase inhibitor 2D |
| **CDKN3** | cyclin dependent kinase inhibitor 3 |
| **CEACAM5** | carcinoembryonic antigen related cell adhesion molecule 5 |
| **CEACAM6** | carcinoembryonic antigen related cell adhesion molecule 6 |
| **CENPF** | centromere protein F |
| **CEP55** | centrosomal protein 55 |
| **CETN2** | centrin 2 |
| **CFD** | complement factor D |
| **CHAD** | chondroadherin |
| **CHEK2** | checkpoint kinase 2 |
| **CHI3L1** | chitinase 3 like 1 |
| **CHIT1** | chitinase 1 |
| **CHRNA5** | cholinergic receptor nicotinic alpha 5 subunit |
| **CKB** | creatine kinase B |
| **CKMT1A** | creatine kinase, mitochondrial 1A |

| | |
|---|---|
| **CKS1B** | CDC28 protein kinase regulatory subunit 1B |
| **CLDN1** | claudin 1 |
| **CLDN3** | claudin 3 |
| **CLDN4** | claudin 4 |
| **CLDN7** | claudin 7 |
| **CLEC14A** | C-type lectin domain containing 14A |
| **CLEC5A** | C-type lectin domain containing 5A |
| **CMKLR1** | chemerin chemokine-like receptor 1 |
| **CNTFR** | ciliary neurotrophic factor receptor |
| **COL11A1** | collagen type XI alpha 1 chain |
| **COL27A1** | collagen type XXVII alpha 1 chain |
| **COL2A1** | collagen type II alpha 1 chain |
| **COL4A6** | collagen type IV alpha 6 chain |
| **COL6A3** | collagen type VI alpha 3 chain |
| **COL7A1** | collagen type VII alpha 1 chain |
| **COL9A3** | collagen type IX alpha 3 chain |
| **COLEC12** | collectin subfamily member 12 |
| **COMP** | cartilage oligomeric matrix protein |
| **CPA3** | carboxypeptidase A3 |
| **CREBBP** | CREB binding protein |
| **CRYAB** | crystallin alpha B |
| **CSF3R** | colony stimulating factor 3 receptor |
| **CTSW** | cathepsin W |
| **CXADR** | CXADR, Ig-like cell adhesion molecule |
| **CXCL10** | C-X-C motif chemokine ligand 10 |
| **CXCL12** | C-X-C motif chemokine ligand 12 |
| **CXCL13** | C-X-C motif chemokine ligand 13 |
| **CXCL5** | C-X-C motif chemokine ligand 5 |
| **CXCL8** | C-X-C motif chemokine ligand 8 |
| **CXCL9** | C-X-C motif chemokine ligand 9 |
| **CXCR6** | C-X-C motif chemokine receptor 6 |
| **CXorf36** | chromosome X open reading frame 36 |
| **CXXC5** | CXXC finger protein 5 |
| **CYBB** | cytochrome b-245 beta chain |
| **CYP4F3** | cytochrome P450 family 4 subfamily F member 3 |
| **DCN** | decorin |
| **DDB2** | damage specific DNA binding protein 2 |
| **DDR2** | discoidin domain receptor tyrosine kinase 2 |
| **DDX39A** | DExD-box helicase 39A |
| **DEPDC1** | DEP domain containing 1 |
| **DHRS2** | dehydrogenase/reductase 2 |
| **DKK1** | dickkopf WNT signaling pathway inhibitor 1 |
| **DKK2** | dickkopf WNT signaling pathway inhibitor 2 |
| **DLGAP5** | DLG associated protein 5 |
| **DLL1** | delta like canonical Notch ligand 1 |
| **DLL3** | delta like canonical Notch ligand 3 |
| **DLL4** | delta like canonical Notch ligand 4 |
| **DNAJC12** | DnaJ heat shock protein family (Hsp40) member C12 |
| **DPT** | dermatopontin |
| **DSC2** | desmocollin 2 |
| **DTX1** | deltex E3 ubiquitin ligase 1 |
| **DTX3** | deltex E3 ubiquitin ligase 3 |
| **DTX4** | deltex E3 ubiquitin ligase 4 |
| **DUSP4** | dual specificity phosphatase 4 |
| **DUSP6** | dual specificity phosphatase 6 |
| **E2F1** | E2F transcription factor 1 |
| **E2F5** | E2F transcription factor 5 |
| **ECM2** | extracellular matrix protein 2 |
| **EDN1** | endothelin 1 |
| **EDNRB** | endothelin receptor type B |
| **EFNA3** | ephrin A3 |
| **EFNA5** | ephrin A5 |
| **EGF** | epidermal growth factor |
| **EGFR** | epidermal growth factor receptor |
| **EGLN2** | egl-9 family hypoxia inducible factor 2 |
| **EGLN3** | egl-9 family hypoxia inducible factor 3 |
| **EIF2AK3** | eukaryotic translation initiation factor 2 alpha kinase 3 |
| **EIF3B** | eukaryotic translation initiation factor 3 subunit B |
| **EIF4E2** | eukaryotic translation initiation factor 4E family member 2 |
| **EIF4EBP1** | eukaryotic translation initiation factor 4E binding protein 1 |
| **ELF3** | E74 like ETS transcription factor 3 |
| **ELK3** | ELK3, ETS transcription factor |
| **ELOVL2** | ELOVL fatty acid elongase 2 |
| **EMCN** | endomucin |
| **ENO1** | enolase 1 |
| **ENPP2** | ectonucleotide pyrophosphatase/phosphodiesterase 2 |
| **EP300** | E1A binding protein p300 |

| | |
|---|---|
| **EPAS1** | endothelial PAS domain protein 1 |
| **ERBB2** | erb-b2 receptor tyrosine kinase 2 |
| **ERBB4** | erb-b2 receptor tyrosine kinase 4 |
| **ERCC1** | ERCC excision repair 1, endonuclease non-catalytic subunit |
| **EREG** | epiregulin |
| **ESPL1** | extra spindle pole bodies like 1, separase |
| **ESR1** | estrogen receptor 1 |
| **ETV4** | ETS variant 4 |
| **ETV7** | ETS variant 7 |
| **EXO1** | exonuclease 1 |
| **EYA1** | EYA transcriptional coactivator and phosphatase 1 |
| **EYA2** | EYA transcriptional coactivator and phosphatase 2 |
| **EYA4** | EYA transcriptional coactivator and phosphatase 4 |
| **F3** | coagulation factor III, tissue factor |
| **FAM124B** | family with sequence similarity 124 member B |
| **FAM198B** | family with sequence similarity 198 member B |
| **FAM214A** | family with sequence similarity 214 member A |
| **FAM83D** | family with sequence similarity 83 member D |
| **FANCF** | Fanconi anemia complementation group F |
| **FAP** | fibroblast activation protein alpha |
| **FBN1** | fibrillin 1 |
| **FGF1** | fibroblast growth factor 1 |
| **FGF10** | fibroblast growth factor 10 |
| **FGF12** | fibroblast growth factor 12 |
| **FGF13** | fibroblast growth factor 13 |
| **FGF18** | fibroblast growth factor 18 |
| **FGF2** | fibroblast growth factor 2 |
| **FGF7** | fibroblast growth factor 7 |
| **FGF9** | fibroblast growth factor 9 |
| **FGFR2** | fibroblast growth factor receptor 2 |
| **FGFR3** | fibroblast growth factor receptor 3 |
| **FGFR4** | fibroblast growth factor receptor 4 |
| **FGL2** | fibrinogen like 2 |
| **FHL1** | four and a half LIM domains 1 |
| **FLI1** | Fli-1 proto-oncogene, ETS transcription factor |
| **FLNC** | filamin C |
| **FLRT3** | fibronectin leucine rich transmembrane protein 3 |
| **FLT3** | fms related tyrosine kinase 3 |
| **FNBP1** | formin binding protein 1 |
| **FOS** | Fos proto-oncogene, AP-1 transcription factor subunit |
| **FOSL1** | FOS like 1, AP-1 transcription factor subunit |
| **FOXA1** | forkhead box A1 |
| **FOXC1** | forkhead box C1 |
| **FOXC2** | forkhead box C2 |
| **FOXM1** | forkhead box M1 |
| **FOXP3** | forkhead box P3 |
| **FREM2** | FRAS1 related extracellular matrix protein 2 |
| **FST** | follistatin |
| **FSTL1** | follistatin like 1 |
| **FSTL3** | follistatin like 3 |
| **FUT3** | fucosyltransferase 3 (Lewis blood group) |
| **FXYD3** | FXYD domain containing ion transport regulator 3 |
| **FZD10** | frizzled class receptor 10 |
| **FZD7** | frizzled class receptor 7 |
| **FZD8** | frizzled class receptor 8 |
| **FZD9** | frizzled class receptor 9 |
| **GABRP** | gamma-aminobutyric acid type A receptor pi subunit |
| **GADD45A** | growth arrest and DNA damage inducible alpha |
| **GADD45B** | growth arrest and DNA damage inducible beta |
| **GADD45G** | growth arrest and DNA damage inducible gamma |
| **GAS1** | growth arrest specific 1 |
| **GATA3** | GATA binding protein 3 |
| **GATA4** | GATA binding protein 4 |
| **GDF15** | growth differentiation factor 15 |
| **GDF5** | growth differentiation factor 5 |
| **GGH** | gamma-glutamyl hydrolase |
| **GHR** | growth hormone receptor |
| **GJB2** | gap junction protein beta 2 |
| **GLI3** | GLI family zinc finger 3 |
| **GNG4** | G protein subunit gamma 4 |
| **GNLY** | granulysin |
| **GPC4** | glypican 4 |
| **GPR160** | G protein-coupled receptor 160 |
| **GPX3** | glutathione peroxidase 3 |
| **GRB2** | growth factor receptor bound protein 2 |
| **GRB7** | growth factor receptor bound protein 7 |
| **GREM1** | gremlin 1, DAN family BMP antagonist |

| | |
|---|---|
| **GRIA3** | glutamate ionotropic receptor AMPA type subunit 3 |
| **GRIN1** | glutamate ionotropic receptor NMDA type subunit 1 |
| **GRIN2A** | glutamate ionotropic receptor NMDA type subunit 2A |
| **GSK3B** | glycogen synthase kinase 3 beta |
| **GTF2H2** | general transcription factor IIH subunit 2 |
| **GZMA** | granzyme A |
| **GZMB** | granzyme B |
| **GZMH** | granzyme H |
| **GZMM** | granzyme M |
| **HAPLN1** | hyaluronan and proteoglycan link protein 1 |
| **HAS1** | hyaluronan synthase 1 |
| **HBB** | hemoglobin subunit beta |
| **HDAC1** | histone deacetylase 1 |
| **HDAC10** | histone deacetylase 10 |
| **HDAC11** | histone deacetylase 11 |
| **HDAC2** | histone deacetylase 2 |
| **HDAC5** | histone deacetylase 5 |
| **HDAC6** | histone deacetylase 6 |
| **HDC** | histidine decarboxylase |
| **HEG1** | heart development protein with EGF like domains 1 |
| **HELLS** | helicase, lymphoid specific |
| **HEMK1** | HemK methyltransferase family member 1 |
| **HES1** | hes family bHLH transcription factor 1 |
| **HGF** | hepatocyte growth factor |
| **HIF1A** | hypoxia inducible factor 1 alpha subunit |
| **HIST1H1C** | histone cluster 1 H1 family member c |
| **HIST1H2BH** | histone cluster 1 H2B family member h |
| **HIST1H3H** | histone cluster 1 H3 family member h |
| **HIST3H2BB** | histone cluster 3 H2B family member b |
| **HK2** | hexokinase 2 |
| **HLA-A** | H1 histone family member 0 |
| **HLA-B** | major histocompatibility complex, class I, B |
| **HLA-C** | major histocompatibility complex, class I, C |
| **HLA-DMA** | major histocompatibility complex, class II, DM alpha |
| **HLA-DMB** | major histocompatibility complex, class II, DM beta |
| **HLA-DOB** | major histocompatibility complex, class II, DO beta |
| **HLA-DPA1** | major histocompatibility complex, class II, DP alpha 1 |
| **HLA-DPB1** | major histocompatibility complex, class II, DP beta 1 |
| **HLA-DQA1** | major histocompatibility complex, class II, DQ alpha 1 |
| **HLA-DQB1** | major histocompatibility complex, class II, DQ beta 1 |
| **HLA-DRA** | major histocompatibility complex, class II, DR alpha |
| **HLA-DRB1** | major histocompatibility complex, class II, DR beta 1 |
| **HLA-E** | major histocompatibility complex, class I, E |
| **HMGA1** | high mobility group AT-hook 1 |
| **HNF1A** | HNF1 homeobox A |
| **HOXA5** | homeobox A5 |
| **HOXA7** | homeobox A7 |
| **HOXA9** | homeobox A9 |
| **HOXB13** | homeobox B13 |
| **HOXB3** | homeobox B3 |
| **HSPA2** | heat shock protein family A (Hsp70) member 2 |
| **IBSP** | integrin binding sialoprotein |
| **ICAM1** | intercellular adhesion molecule 1 |
| **ID1** | inhibitor of DNA binding 1, HLH protein |
| **ID2** | inhibitor of DNA binding 2 |
| **ID4** | inhibitor of DNA binding 4, HLH protein |
| **IDO1** | indoleamine 2,3-dioxygenase 1 |
| **IFT140** | intraflagellar transport 140 |
| **IGF1** | insulin like growth factor 1 |
| **IGF1R** | insulin like growth factor 1 receptor |
| **IKZF3** | IKAROS family zinc finger 3 |
| **IL10RA** | interleukin 10 receptor subunit alpha |
| **IL11RA** | interleukin 11 receptor subunit alpha |
| **IL12RB2** | interleukin 12 receptor subunit beta 2 |
| **IL13RA1** | interleukin 13 receptor subunit alpha 1 |
| **IL1B** | interleukin 1 beta |
| **IL1R2** | interleukin 1 receptor type 2 |
| **IL1RN** | interleukin 1 receptor antagonist |
| **IL20RA** | interleukin 20 receptor subunit alpha |
| **IL20RB** | interleukin 20 receptor subunit beta |
| **IL22RA2** | interleukin 22 receptor subunit alpha 2 |
| **IL24** | interleukin 24 |
| **IL2RA** | interleukin 2 receptor subunit alpha |
| **IL2RB** | interleukin 2 receptor subunit beta |
| **IL3RA** | interleukin 3 receptor subunit alpha |
| **IL4R** | interleukin 4 receptor |
| **IL6** | interleukin 6 |

| | |
|---|---|
| **IL6R** | interleukin 6 receptor |
| **IL7R** | interleukin 7 receptor |
| **INHBA** | inhibin beta A subunit |
| **INHBB** | inhibin beta B subunit |
| **IRF6** | interferon regulatory factor 6 |
| **IRX1** | iroquois homeobox 1 |
| **ISG15** | ISG15 ubiquitin-like modifier |
| **ISM1** | isthmin 1 |
| **ITGA6** | integrin subunit alpha 6 |
| **ITGAV** | integrin subunit alpha V |
| **ITGB1** | integrin subunit beta 1 |
| **ITGB3** | integrin subunit beta 3 |
| **ITGB6** | integrin subunit beta 6 |
| **ITPR1** | inositol 1,4,5-trisphosphate receptor type 1 |
| **JAG1** | jagged 1 |
| **JAG2** | jagged 2 |
| **JAK1** | Janus kinase 1 |
| **JAK2** | Janus kinase 2 |
| **JAK3** | Janus kinase 3 |
| **JAM2** | junctional adhesion molecule 2 |
| **JCAD** | junctional cadherin 5 associated |
| **JUN** | Jun proto-oncogene, AP-1 transcription factor subunit |
| **KAT2B** | lysine acetyltransferase 2B |
| **KCNB1** | potassium voltage-gated channel subfamily B member 1 |
| **KDR** | kinase insert domain receptor |
| **KIAA0040** | KIAA0040 |
| **KIF11** | kinesin family member 11 |
| **KIF14** | kinesin family member 14 |
| **KIF23** | kinesin family member 23 |
| **KIF2C** | kinesin family member 2C |
| **KIFC1** | kinesin family member C1 |
| **KIT** | KIT proto-oncogene receptor tyrosine kinase |
| **KLRK1** | killer cell lectin like receptor K1 |
| **KRT14** | keratin 14 |
| **KRT17** | keratin 17 |
| **KRT5** | keratin 5 |
| **KRT6B** | keratin 6B |
| **KRT7** | keratin 7 |
| **LAD1** | ladinin 1 |
| **LAG3** | lymphocyte activating 3 |
| **LAMA3** | laminin subunit alpha 3 |
| **LAMB3** | laminin subunit beta 3 |
| **LAMC2** | laminin subunit gamma 2 |
| **LEF1** | lymphoid enhancer binding factor 1 |
| **LEFTY2** | left-right determination factor 2 |
| **LEMD1** | LEM domain containing 1 |
| **LEP** | leptin |
| **LEPR** | leptin receptor |
| **LFNG** | LFNG O-fucosylpeptide 3-beta-N-acetylglucosaminyltransferase |
| **LIF** | LIF, interleukin 6 family cytokine |
| **LIFR** | LIF receptor alpha |
| **LINC02381** | long intergenic non-protein coding RNA 2381 |
| **LPL** | lipoprotein lipase |
| **LRP2** | LDL receptor related protein 2 |
| **LRRC32** | leucine rich repeat containing 32 |
| **LTB** | lymphotoxin beta |
| **LTBP1** | latent transforming growth factor beta binding protein 1 |
| **MAD2L1** | mitotic arrest deficient 2 like 1 |
| **MAF** | MAF bZIP transcription factor |
| **MAML2** | mastermind like transcriptional coactivator 2 |
| **MAP2K4** | mitogen-activated protein kinase kinase 4 |
| **MAP3K12** | mitogen-activated protein kinase kinase kinase 12 |
| **MAPK1** | mitogen-activated protein kinase 1 |
| **MAPK10** | mitogen-activated protein kinase 10 |
| **MAPK3** | mitogen-activated protein kinase 3 |
| **MAPK8IP2** | mitogen-activated protein kinase 8 interacting protein 2 |
| **MAPT** | microtubule associated protein tau |
| **MARCO** | macrophage receptor with collagenous structure |
| **MCM2** | minichromosome maintenance complex component 2 |
| **MCM3** | minichromosome maintenance complex component 3 |
| **MDM2** | MDM2 proto-oncogene |
| **MED1** | mediator complex subunit 1 |
| **MELK** | maternal embryonic leucine zipper kinase |
| **MEOX2** | mesenchyme homeobox 2 |
| **MET** | MET proto-oncogene, receptor tyrosine kinase |
| **MFNG** | MFNG O-fucosylpeptide 3-beta-N-acetylglucosaminyltransferase |
| **MIA** | melanoma inhibitory activity |

| | |
|---|---|
| **MIS18A** | MIS18 kinetochore protein A |
| **MKI67** | marker of proliferation Ki-67 |
| **MLH1** | mutL homolog 1 |
| **MLLT3** | MLLT3, super elongation complex subunit |
| **MLPH** | melanophilin |
| **MME** | membrane metalloendopeptidase |
| **MMP11** | matrix metallopeptidase 11 |
| **MMP14** | matrix metallopeptidase 14 |
| **MMP3** | matrix metallopeptidase 3 |
| **MMP7** | matrix metallopeptidase 7 |
| **MMP9** | matrix metallopeptidase 9 |
| **MMRN2** | multimerin 2 |
| **MRE11** | MRE11 homolog, double strand break repair nuclease |
| **MS4A2** | membrane spanning 4-domains A2 |
| **MSR1** | macrophage scavenger receptor 1 |
| **MT1G** | metallothionein 1G |
| **MTOR** | mechanistic target of rapamycin kinase |
| **MUC1** | mucin 1, cell surface associated |
| **MUS81** | MUS81 structure-specific endonuclease subunit |
| **MYBL2** | MYB proto-oncogene like 2 |
| **MYC** | MYC proto-oncogene, bHLH transcription factor |
| **MYCN** | MYCN proto-oncogene, bHLH transcription factor |
| **MYCT1** | MYC target 1 |
| **NASP** | nuclear autoantigenic sperm protein |
| **NAT1** | N-acetyltransferase 1 |
| **NCAM1** | neural cell adhesion molecule 1 |
| **NCAPH2** | non-SMC condensin II complex subunit H2 |
| **NDC80** | NDC80, kinetochore complex component |
| **NDP** | NDP, norrin cystine knot growth factor |
| **NEIL1** | nei like DNA glycosylase 1 |
| **NEIL2** | nei like DNA glycosylase 2 |
| **NEIL3** | nei like DNA glycosylase 3 |
| **NEO1** | neogenin 1 |
| **NETO2** | neuropilin and tolloid like 2 |
| **NFATC1** | nuclear factor of activated T-cells 1 |
| **NFKBIZ** | NFKB inhibitor zeta |
| **NGFR** | nerve growth factor receptor |
| **NKG7** | natural killer cell granule protein 7 |
| **NOD2** | nucleotide binding oligomerization domain containing 2 |
| **NOTCH1** | notch 1 |
| **NOTCH2** | notch 2 |
| **NOTCH3** | notch 3 |
| **NPEPPS** | aminopeptidase puromycin sensitive |
| **NPR1** | natriuretic peptide receptor 1 |
| **NR4A1** | nuclear receptor subfamily 4 group A member 1 |
| **NR4A3** | nuclear receptor subfamily 4 group A member 3 |
| **NRCAM** | neuronal cell adhesion molecule |
| **NRXN1** | neurexin 1 |
| **NRXN3** | neurexin 3 |
| **NSD1** | nuclear receptor binding SET domain protein 1 |
| **NSD3** | nuclear receptor binding SET domain protein 3 |
| **NTRK2** | neurotrophic receptor tyrosine kinase 2 |
| **NUDT1** | nudix hydrolase 1 |
| **NUF2** | NUF2, NDC80 kinetochore complex component |
| **NUMBL** | NUMB like, endocytic adaptor protein |
| **NUPR1** | nuclear protein 1, transcriptional regulator |
| **OAS3** | 2'-5'-oligoadenylate synthetase 3 |
| **OCLN** | occludin |
| **OGN** | osteoglycin |
| **OLFML2B** | olfactomedin like 2B |
| **ORC6** | origin recognition complex subunit 6 |
| **PALB2** | partner and localizer of BRCA2 |
| **PALMD** | palmdelphin |
| **PARP1** | poly(ADP-ribose) polymerase 1 |
| **PARP2** | poly(ADP-ribose) polymerase 2 |
| **PARP4** | poly(ADP-ribose) polymerase family member 4 |
| **PAX5** | paired box 5 |
| **PAX8** | paired box 8 |
| **PBX3** | PBX homeobox 3 |
| **PCK1** | phosphoenolpyruvate carboxykinase 1 |
| **PCNA** | proliferating cell nuclear antigen |
| **PDCD1** | programmed cell death 1 |
| **PDCD1LG2** | programmed cell death 1 ligand 2 |
| **PDE9A** | phosphodiesterase 9A |
| **PDGFB** | platelet derived growth factor subunit B |
| **PDGFRA** | platelet derived growth factor receptor alpha |
| **PDGFRB** | platelet derived growth factor receptor beta |

| | |
|---|---|
| **PDK4** | pyruvate dehydrogenase kinase 4 |
| **PECAM1** | platelet and endothelial cell adhesion molecule 1 |
| **PFDN2** | prefoldin subunit 2 |
| **PGK1** | phosphoglycerate kinase 1 |
| **PGR** | progesterone receptor |
| **PHGDH** | phosphoglycerate dehydrogenase |
| **PIK3CA** | phosphatidylinositol-4,5-bisphosphate 3-kinase catalytic subunit alpha |
| **PIK3CD** | phosphatidylinositol-4,5-bisphosphate 3-kinase catalytic subunit delta |
| **PIK3CG** | phosphatidylinositol-4,5-bisphosphate 3-kinase catalytic subunit gamma |
| **PIK3R1** | phosphoinositide-3-kinase regulatory subunit 1 |
| **PIK3R2** | phosphoinositide-3-kinase regulatory subunit 2 |
| **PIK3R3** | phosphoinositide-3-kinase regulatory subunit 3 |
| **PIK3R5** | phosphoinositide-3-kinase regulatory subunit 5 |
| **PIM1** | Pim-1 proto-oncogene, serine/threonine kinase |
| **PIP** | prolactin induced protein |
| **PKMYT1** | protein kinase, membrane associated tyrosine/threonine 1 |
| **PLA1A** | phospholipase A1 member A |
| **PLA2G2A** | phospholipase A2 group IIA |
| **PLA2G3** | phospholipase A2 group III |
| **PLA2G4A** | phospholipase A2 group IVA |
| **PLA2G4F** | phospholipase A2 group IVF |
| **PLAT** | plasminogen activator, tissue type |
| **PLCB1** | phospholipase C beta 1 |
| **PLCB4** | phospholipase C beta 4 |
| **PLCE1** | phospholipase C epsilon 1 |
| **PLD1** | phospholipase D1 |
| **PMS2** | PMS1 homolog 2, mismatch repair system component |
| **POLD1** | DNA polymerase delta 1, catalytic subunit |
| **POLQ** | DNA polymerase theta |
| **POPDC3** | popeye domain containing 3 |
| **PPARG** | peroxisome proliferator activated receptor gamma |
| **PPARGC1A** | PPARG coactivator 1 alpha |
| **PPP2CB** | protein phosphatase 2 catalytic subunit beta |
| **PPP2R1A** | protein phosphatase 2 scaffold subunit Aalpha |
| **PPP2R2C** | protein phosphatase 2 regulatory subunit Bgamma |
| **PRC1** | protein regulator of cytokinesis 1 |
| **PREP** | prolyl endopeptidase |
| **PRF1** | perforin 1 |
| **PRKAA2** | protein kinase AMP-activated catalytic subunit alpha 2 |
| **PRKACA** | protein kinase cAMP-activated catalytic subunit alpha |
| **PRKACB** | protein kinase cAMP-activated catalytic subunit beta |
| **PRKCA** | protein kinase C alpha |
| **PRKCB** | protein kinase C beta |
| **PRKDC** | protein kinase, DNA-activated, catalytic polypeptide |
| **PRKX** | protein kinase, X-linked |
| **PRLR** | prolactin receptor |
| **PROM1** | prominin 1 |
| **PSAT1** | phosphoserine aminotransferase 1 |
| **PSMB10** | proteasome subunit beta 10 |
| **PSMB7** | proteasome subunit beta 7 |
| **PSMB9** | proteasome subunit beta 9 |
| **PTCH1** | patched 1 |
| **PTEN** | phosphatase and tensin homolog |
| **PTGDS** | prostaglandin D2 synthase |
| **PTGER3** | prostaglandin E receptor 3 |
| **PTGS2** | prostaglandin-endoperoxide synthase 2 |
| **PTTG1** | pituitary tumor-transforming 1 |
| **PYCARD** | PYD and CARD domain containing |
| **RAC2** | Rac family small GTPase 2 |
| **RAC3** | Rac family small GTPase 3 |
| **RAD51** | RAD51 recombinase |
| **RAD51C** | RAD51 paralog C |
| **RAD52** | RAD52 homolog, DNA repair protein |
| **RAD54L** | RAD54 like |
| **RARRES3** | retinoic acid receptor responder 3 |
| **RASAL1** | RAS protein activator like 1 |
| **RASGRF1** | Ras protein specific guanine nucleotide releasing factor 1 |
| **RASGRF2** | Ras protein specific guanine nucleotide releasing factor 2 |
| **RASGRP1** | RAS guanyl releasing protein 1 |
| **RB1** | RB transcriptional corepressor 1 |
| **RBL1** | RB transcriptional corepressor like 1 |
| **RBL2** | RB transcriptional corepressor like 2 |
| **RBX1** | ring-box 1 |
| **RELN** | reelin |
| **RFC4** | replication factor C subunit 4 |
| **RNASE2** | ribonuclease A family member 2 |
| **RNF103** | ring finger protein 103 |

| | |
|---|---|
| **ROBO4** | roundabout guidance receptor 4 |
| **ROCK1** | Rho associated coiled-coil containing protein kinase 1 |
| **ROCK2** | Rho associated coiled-coil containing protein kinase 2 |
| **RORA** | RAR related orphan receptor A |
| **RORB** | RAR related orphan receptor B |
| **RPS6KA5** | ribosomal protein S6 kinase A5 |
| **RPS6KB1** | ribosomal protein S6 kinase B1 |
| **RPS6KB2** | ribosomal protein S6 kinase B2 |
| **RRM2** | ribonucleotide reductase regulatory subunit M2 |
| **RUNX3** | runt related transcription factor 3 |
| **S100A14** | S100 calcium binding protein A14 |
| **S100A7** | S100 calcium binding protein A7 |
| **S1PR1** | sphingosine-1-phosphate receptor 1 |
| **SCARA5** | scavenger receptor class A member 5 |
| **SCUBE2** | signal peptide, CUB domain and EGF like domain containing 2 |
| **SELE** | selectin E |
| **SERBP1** | SERPINE1 mRNA binding protein 1 |
| **SERPINB5** | serpin family B member 5 |
| **SERPINH1** | serpin family H member 1 |
| **SFN** | stratifin |
| **SFRP1** | secreted frizzled related protein 1 |
| **SFRP2** | secreted frizzled related protein 2 |
| **SFRP4** | secreted frizzled related protein 4 |
| **SHC2** | SHC adaptor protein 2 |
| **SHC4** | SHC adaptor protein 4 |
| **SHE** | Src homology 2 domain containing E |
| **SHMT2** | serine hydroxymethyltransferase 2 |
| **SIDT1** | SID1 transmembrane family member 1 |
| **SIGIRR** | single Ig and TIR domain containing |
| **SIX1** | SIX homeobox 1 |
| **SKA3** | spindle and kinetochore associated complex subunit 3 |
| **SKP1** | S-phase kinase associated protein 1 |
| **SKP2** | S-phase kinase associated protein 2 |
| **SLC2A1** | solute carrier family 2 member 1 |
| **SLC39A6** | solute carrier family 39 member 6 |
| **SLC44A4** | solute carrier family 44 member 4 |
| **SLPI** | secretory leukocyte peptidase inhibitor |
| **SMAD1** | SMAD family member 1 |
| **SMAD3** | SMAD family member 3 |
| **SMAD4** | SMAD family member 4 |
| **SMAD5** | SMAD family member 5 |
| **SMC1B** | structural maintenance of chromosomes 1B |
| **SMO** | smoothened, frizzled class receptor |
| **SMURF2** | SMAD specific E3 ubiquitin protein ligase 2 |
| **SNAI1** | snail family transcriptional repressor 1 |
| **SNAI2** | snail family transcriptional repressor 2 |
| **SOCS1** | suppressor of cytokine signaling 1 |
| **SOCS2** | suppressor of cytokine signaling 2 |
| **SOCS3** | suppressor of cytokine signaling 3 |
| **SOX10** | SRY-box 10 |
| **SOX17** | SRY-box 17 |
| **SOX2** | SRY-box 2 |
| **SOX9** | SRY-box 9 |
| **SP1** | Sp1 transcription factor |
| **SPC25** | SPC25, NDC80 kinetochore complex component |
| **SPDEF** | SAM pointed domain containing ETS transcription factor |
| **SPN** | sialophorin |
| **SPP1** | secreted phosphoprotein 1 |
| **SPRY1** | sprouty RTK signaling antagonist 1 |
| **SPRY2** | sprouty RTK signaling antagonist 2 |
| **SPRY4** | sprouty RTK signaling antagonist 4 |
| **SRPX** | sushi repeat containing protein, X-linked |
| **ST6GALNAC2** | ST6 N-acetylgalactosaminide alpha-2,6-sialyltransferase 2 |
| **STAT1** | signal transducer and activator of transcription 1 |
| **STC1** | stanniocalcin 1 |
| **SUV39H2** | suppressor of variegation 3-9 homolog 2 |
| **SYTL4** | synaptotagmin like 4 |
| **TAP1** | transporter 1, ATP binding cassette subfamily B member |
| **TAP2** | transporter 2, ATP binding cassette subfamily B member |
| **TAPBP** | TAP binding protein |
| **TBC1D9** | TBC1 domain family member 9 |
| **TBX1** | T-box 1 |
| **TCEAL1** | transcription elongation factor A like 1 |
| **TCF4** | transcription factor 4 |
| **TCF7L1** | transcription factor 7 like 1 |
| **TEK** | TEK receptor tyrosine kinase |
| **TFDP1** | transcription factor Dp-1 |

| | |
|---|---|
| **TFF1** | trefoil factor 1 |
| **TFF3** | trefoil factor 3 |
| **TGFB1** | transforming growth factor beta 1 |
| **TGFB2** | transforming growth factor beta 2 |
| **TGFB3** | transforming growth factor beta 3 |
| **TGFBR2** | transforming growth factor beta receptor 2 |
| **THBS1** | thrombospondin 1 |
| **THBS2** | thrombospondin 2 |
| **THBS4** | thrombospondin 4 |
| **THY1** | Thy-1 cell surface antigen |
| **TIE1** | tyrosine kinase with immunoglobulin like and EGF like domains 1 |
| **TIGIT** | T-cell immunoreceptor with Ig and ITIM domains |
| **TIMP4** | TIMP metallopeptidase inhibitor 4 |
| **TLE3** | transducin like enhancer of split 3 |
| **TLR4** | toll like receptor 4 |
| **TLX1** | T-cell leukemia homeobox 1 |
| **TMEM45B** | transmembrane protein 45B |
| **TMPRSS2** | transmembrane protease, serine 2 |
| **TMPRSS4** | transmembrane protease, serine 4 |
| **TNF** | tumor necrosis factor |
| **TNFAIP6** | TNF alpha induced protein 6 |
| **TNFSF10** | TNF superfamily member 10 |
| **TNKS** | tankyrase |
| **TNKS2** | tankyrase 2 |
| **TNN** | tenascin N |
| **TOP2A** | DNA topoisomerase II alpha |
| **TP53** | tumor protein p53 |
| **TPSAB1** | tryptase alpha/beta 1 |
| **TRIP13** | thyroid hormone receptor interactor 13 |
| **TSPAN1** | tetraspanin 1 |
| **TSPAN7** | tetraspanin 7 |
| **TTK** | TTK protein kinase |
| **TTYH1** | tweety family member 1 |
| **TUBA4A** | tubulin alpha 4a |
| **TWIST1** | twist family bHLH transcription factor 1 |
| **TWIST2** | twist family bHLH transcription factor 2 |
| **TYK2** | tyrosine kinase 2 |
| **TYMP** | thymidine phosphorylase |
| **TYMS** | thymidylate synthetase |
| **UBE2C** | ubiquitin conjugating enzyme E2 C |
| **UBE2T** | ubiquitin conjugating enzyme E2 T |
| **VCAN** | versican |
| **VEGFA** | vascular endothelial growth factor A |
| **VEGFD** | vascular endothelial growth factor D |
| **VIM** | vimentin |
| **VIT** | vitrin |
| **WDR77** | WD repeat domain 77 |
| **WEE1** | WEE1 G2 checkpoint kinase |
| **WIF1** | WNT inhibitory factor 1 |
| **WNT10A** | Wnt family member 10A |
| **WNT11** | Wnt family member 11 |
| **WNT2** | Wnt family member 2 |
| **WNT4** | Wnt family member 4 |
| **WNT5A** | Wnt family member 5A |
| **WNT5B** | Wnt family member 5B |
| **WNT6** | Wnt family member 6 |
| **WNT7B** | Wnt family member 7B |
| **WRN** | Werner syndrome RecQ like helicase |
| **WT1** | Wilms tumor 1 |
| **XRCC2** | X-ray repair cross complementing 2 |
| **XRCC3** | X-ray repair cross complementing 3 |
| **ZBTB16** | zinc finger and BTB domain containing 16 |
| **ZEB1** | zinc finger E-box binding homeobox 1 |
| **ZEB2** | zinc finger E-box binding homeobox 2 |
| **ZFPM2** | zinc finger protein, FOG family member 2 |
| **ZFYVE9** | zinc finger FYVE-type containing 9 |
| **ZIC2** | Zic family member 2 |
| **ZNF205** | zinc finger protein 205 |
| **ABCF1** | ATP binding cassette subfamily F member 1 |
| **ACTB** | actin beta |
| **G6PD** | glucose-6-phosphate dehydrogenase |
| **GUSB** | glucuronidase beta |
| **MRPL19** | mitochondrial ribosomal protein L19 |
| **NRDE2** | NRDE-2, necessary for RNA interference, domain containing |
| **OAZ1** | ornithine decarboxylase antizyme 1 |
| **POLR2A** | RNA polymerase II subunit A |
| **PSMC4** | proteasome 26S subunit, ATPase 4 |

| | |
|---|---|
| **PUM1** | pumilio RNA binding family member 1 |
| **RPLP0** | ribosomal protein lateral stalk subunit P0 |
| **SDHA** | succinate dehydrogenase complex flavoprotein subunit A |
| **SF3A1** | splicing factor 3a subunit 1 |
| **STK11IP** | serine/threonine kinase 11 interacting protein |
| **TBC1D10B** | TBC1 domain family member 10B |
| **TBP** | TATA-box binding protein |
| **TFRC** | transferrin receptor |
| **UBB** | ubiquitin B |

Supplemental material 3

| |
|---|
| ABL1 |
| ACVR1B |
| AKT1 |
| AKT2 |
| AKT3 |
| ALK |
| ALOX12B |
| AMER1 |
| APC |
| AR |
| ARAF |
| ARFRP1 |
| ARID1A |
| ASXL1 |
| ATM |
| ATR |
| ATRX |
| AURKA |
| AURKB |
| AXIN1 |
| AXL |
| BAP1 |
| BARD1 |
| BCL2 |
| BCL2L1 |
| BCL2L2 |
| BCL6 |
| BCOR |
| BCORL1 |
| BRAF |
| BRCA1 |
| BRCA2 |
| BRD4 |
| BRIP1 |
| BTG1 |
| BTG2 |
| BTK |
| C11orf30 |
| CALR |
| CARD11 |
| CASP8 |
| CBFB |
| CBL |
| CCND1 |
| CCND2 |
| CCND3 |
| CCNE1 |
| CD22 |
| CD274 |
| CD70 |
| CD79A |
| CD79B |
| CDC73 |
| CDH1 |
| CDK12 |
| CDK4 |
| CDK6 |
| CDK8 |
| CDKN1A |
| CDKN1B |
| CDKN2A |
| CDKN2B |
| CDKN2C |
| CEBPA |
| CHEK1 |
| CHEK2 |

| |
|---|
| CIC |
| CREBBP |
| CRKL |
| CSF1R |
| CSF3R |
| CTCF |
| CTNNA1 |
| CTNNB1 |
| CUL3 |
| CUL4A |
| CXCR4 |
| CYP17A1 |
| DAXX |
| DDR1 |
| DDR2 |
| DIS3 |
| DNMT3A |
| DOT1L |
| EED |
| EGFR |
| EP300 |
| EPHA3 |
| EPHB1 |
| EPHB4 |
| ERBB2 |
| ERBB3 |
| ERBB4 |
| ERCC4 |
| ERG |
| ERRFI1 |
| ESR1 |
| EZH2 |
| FAM46C |
| FANCA |
| FANCC |
| FANCG |
| FANCL |
| FAS |
| FBXW7 |
| FGF10 |
| FGF12 |
| FGF14 |
| FGF19 |
| FGF23 |
| FGF3 |
| FGF4 |
| FGF6 |
| FGFR1 |
| FGFR2 |
| FGFR3 |
| FGFR4 |
| FH |
| FLCN |
| FLT1 |
| FLT3 |
| FOXL2 |
| FUBP1 |
| GABRA6 |
| GATA3 |
| GATA4 |
| GATA6 |
| GID4 |
| GNA11 |
| GNA13 |
| GNAQ |
| GNAS |
| GRM3 |
| GSK3B |

| |
|---|
| H3F3A |
| HDAC1 |
| HGF |
| HNF1A |
| HRAS |
| HSD3B1 |
| ID3 |
| IDH1 |
| IDH2 |
| IGF1R |
| IKBKE |
| IKZF1 |
| INPP4B |
| IRF2 |
| IRF4 |
| IRS2 |
| JAK1 |
| JAK2 |
| JAK3 |
| JUN |
| KDM5A |
| KDM5C |
| KDM6A |
| KDR |
| KEAP1 |
| KEL |
| KIT |
| KLHL6 |
| KMT2A |
| KMT2D |
| KRAS |
| LTK |
| LYN |
| MAF |
| MAP2K1 |
| MAP2K2 |
| MAP2K4 |
| MAP3K1 |
| MAP3K13 |
| MAPK1 |
| MCL1 |
| MDM2 |
| MDM4 |
| MED12 |
| MEF2B |
| MEN1 |
| MERTK |
| MET |
| MITF |
| MKNK1 |
| MLH1 |
| MPL |
| MRE11A |
| MSH2 |
| MSH3 |
| MSH6 |
| MST1R |
| MTAP |
| MTOR |
| MUTYH |
| MYC |
| MYCL |
| MYCN |
| MYD88 |
| NBN |
| NF1 |
| NF2 |
| NFE2L2 |

| |
|---|
| NFKBIA |
| NKX2-1 |
| NOTCH1 |
| NOTCH2 |
| NOTCH3 |
| NPM1 |
| NRAS |
| NT5C2 |
| NTRK1 |
| NTRK2 |
| NTRK3 |
| P2RY8 |
| Page |
| PALB2 |
| PARK2 |
| PARP1 |
| PARP2 |
| PARP3 |
| PAX5 |
| PBRM1 |
| PDCD1 |
| PDCD1LG2 |
| PDGFRA |
| PDGFRB |
| PDK1 |
| PIK3C2B |
| PIK3C2G |
| PIK3CA |
| PIK3CB |
| PIK3R1 |
| PIM1 |
| PMS2 |
| POLD1 |
| POLE |
| PPARG |
| PPP2R1A |
| PPP2R2A |
| PRDM1 |
| PRKAR1A |
| PRKCI |
| PTCH1 |
| PTEN |
| PTPN11 |
| PTPRO |
| QKI |
| RAC1 |
| RAD21 |
| RAD51 |
| RAD51B |
| RAD51C |
| RAD51D |
| RAD52 |
| RAD54L |
| RAF1 |
| RAL-0003-03 |
| RARA |
| RB1 |
| RBM10 |
| REL |
| RET |
| RICTOR |
| RNF43 |
| ROS1 |
| RPTOR |
| SDHA |
| SDHB |
| SDHC |
| SDHD |

| |
|---|
| SETD2 |
| SF3B1 |
| SGK1 |
| SMAD2 |
| SMAD4 |
| SMARCA4 |
| SMARCB1 |
| SMO |
| SNCAIP |
| SOCS1 |
| SOX2 |
| SOX9 |
| SPEN |
| SPOP |
| SRC |
| STAG2 |
| STAT3 |
| STK11 |
| SUFU |
| SYK |
| TBX3 |
| TEK |
| TET2 |
| TGFBR2 |
| TIPARP |
| TNFAIP3 |
| TNFRSF14 |
| TP53 |
| TSC1 |
| TSC2 |
| TYRO3 |
| U2AF1 |
| VEGFA |
| VHL |
| WHSC1 |
| WHSC1L1 |
| WT1 |
| XPO1 |
| XRCC2 |
| ZNF217 |
| ZNF703 |

Supplemental material 4

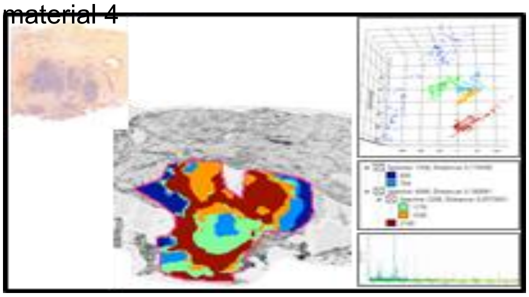

Case 1

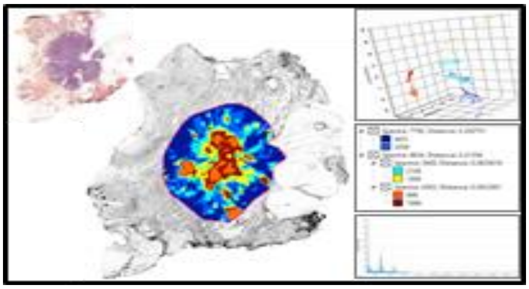

Case 2

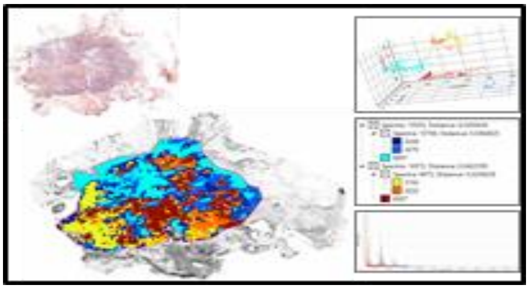

Case 3

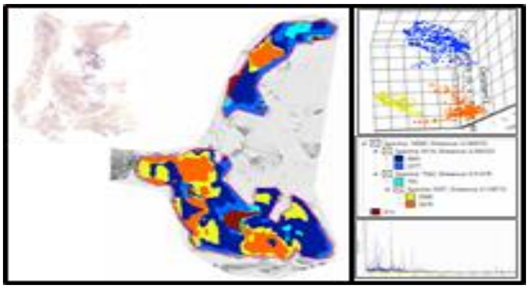

Case 4

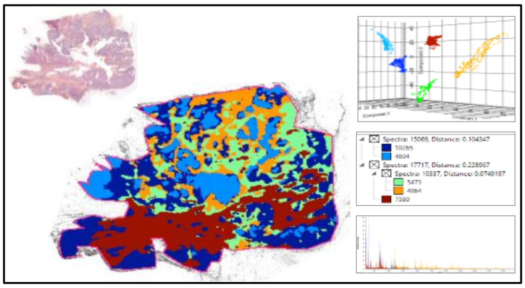

Case 5

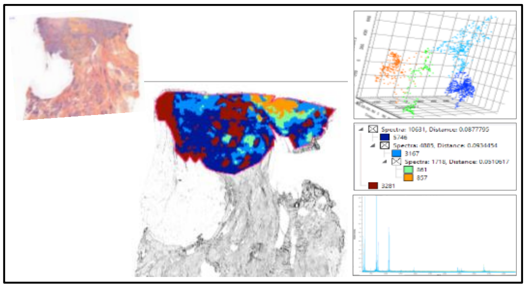

Case 6

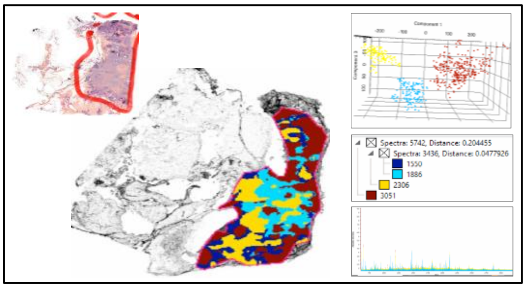

Case 7

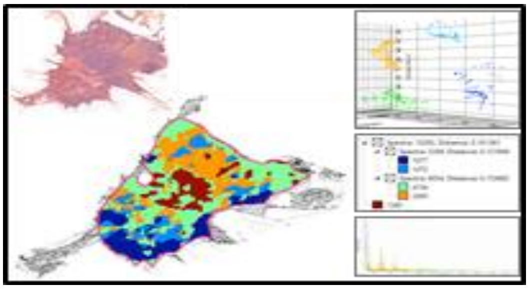

Case 8

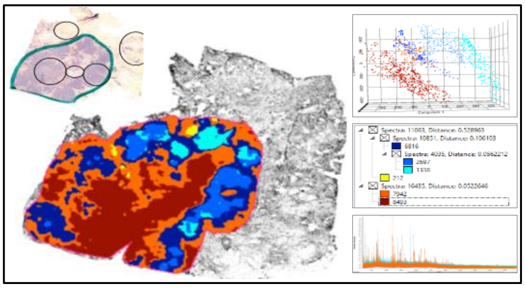

Case 9

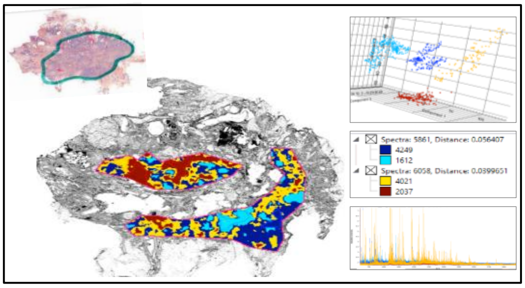

Case 10

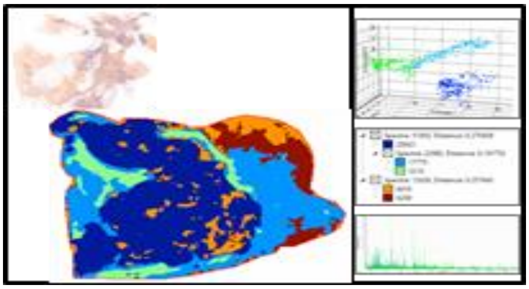

Case 11

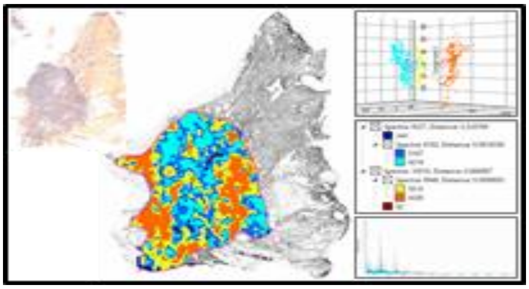

Case 12

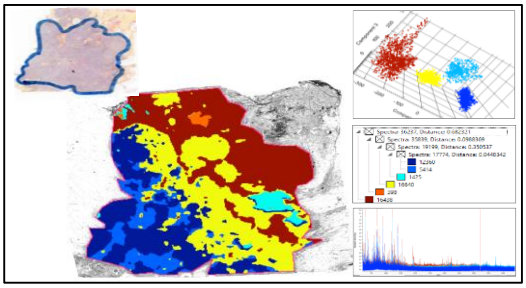

Case 13

Case 14

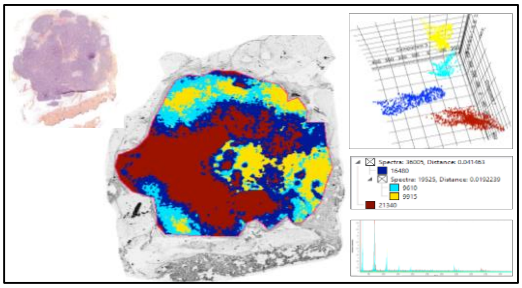

Case 15

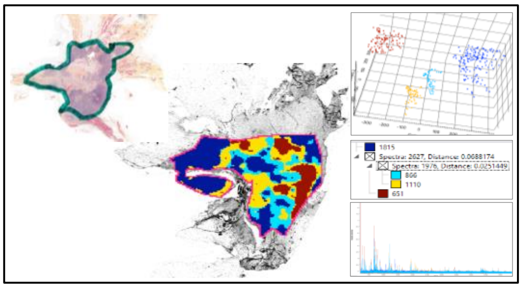

Case 16

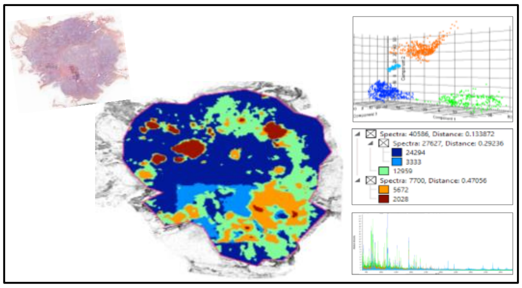

Case 17

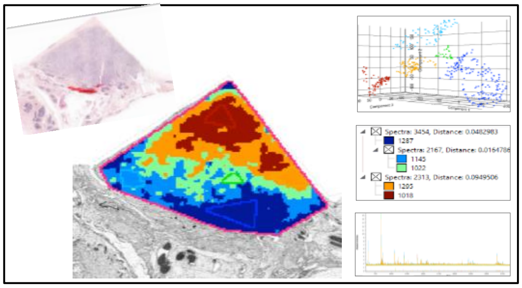

Case 18

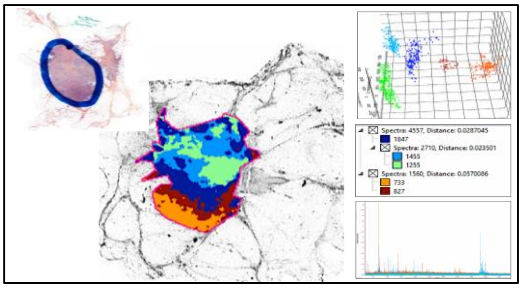

Case 19

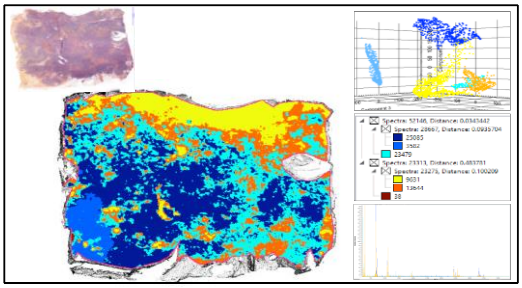

Case 20

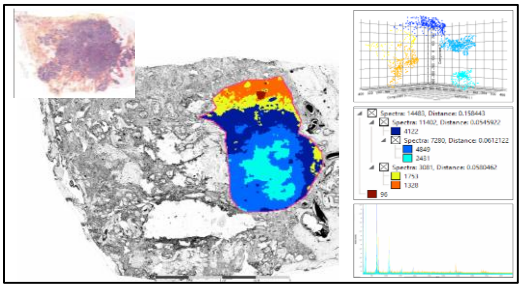

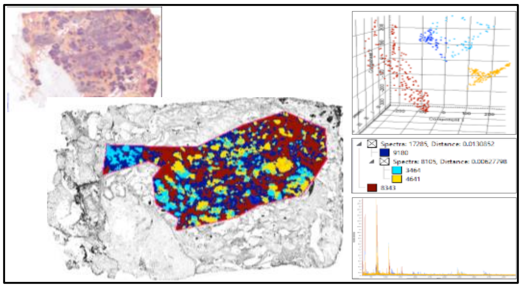

Case 21

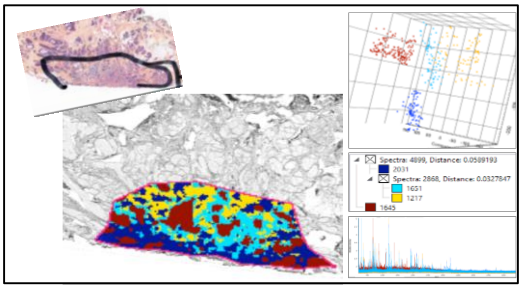

Case 22

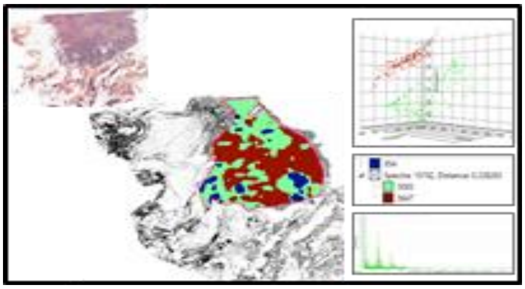

Case 23

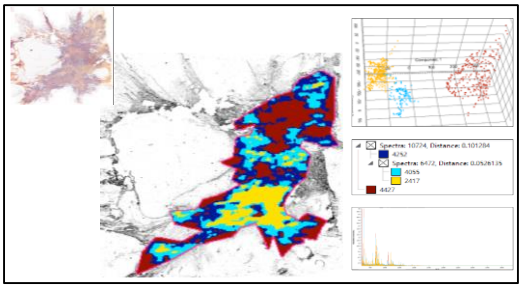

Case 24

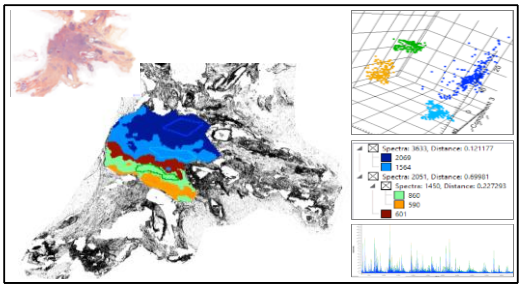

Case 25

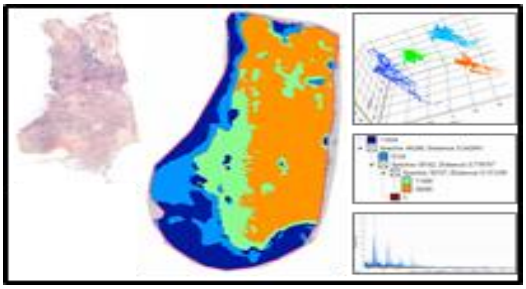

Case 26

Case 27

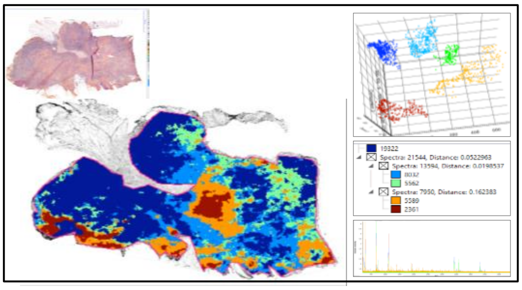

Case 28

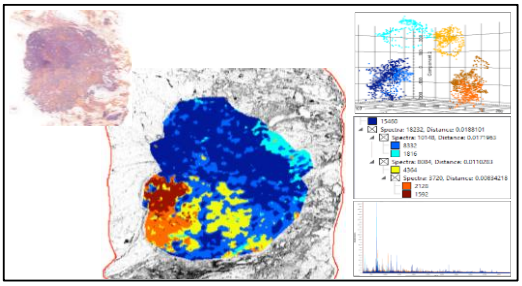

Case 29

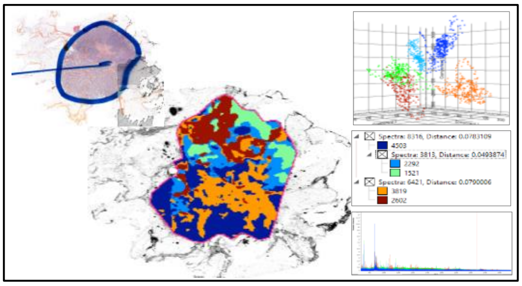

Case 30

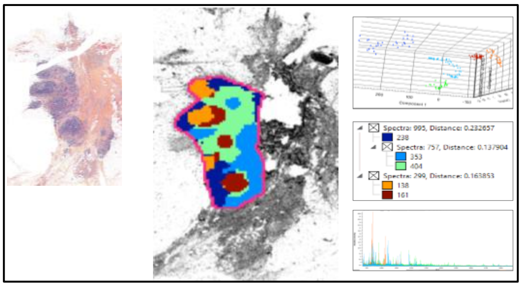

Case 31

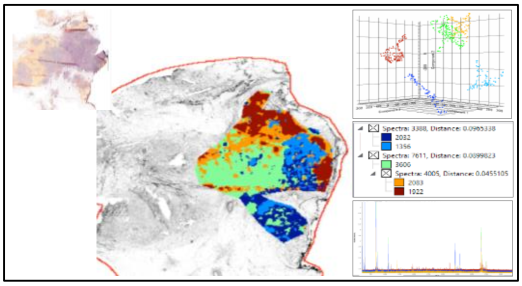

Case 32

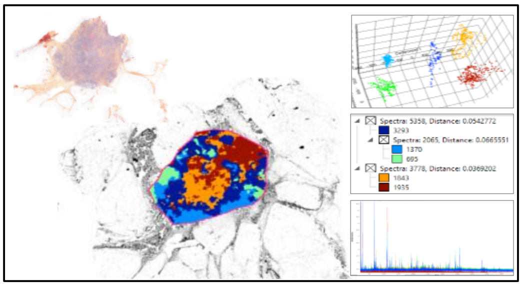

Case 33

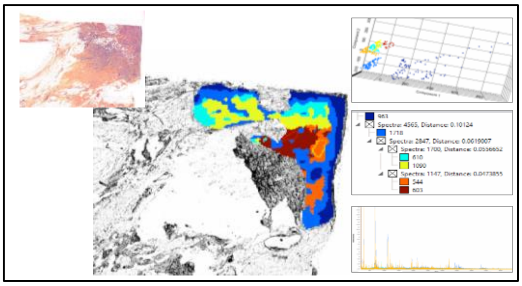

Case 34

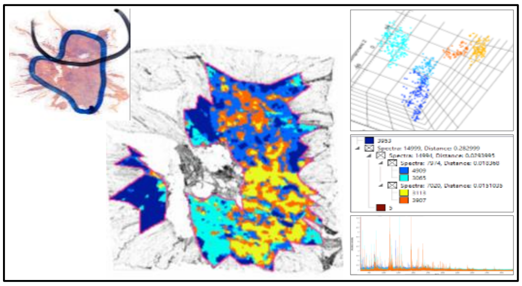

Case 35

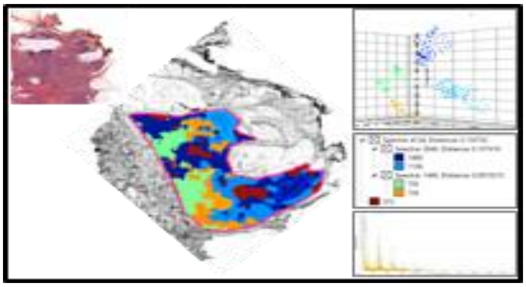

Case 36

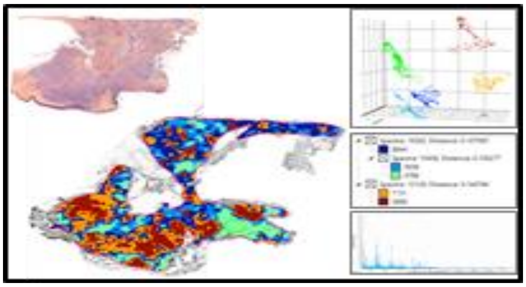

Case 37

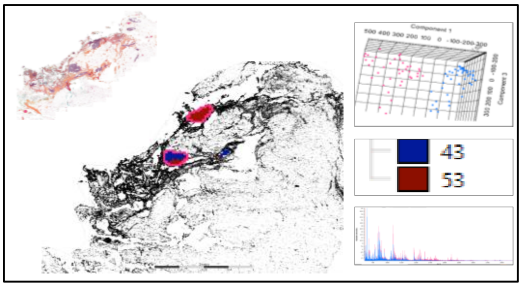

Case 38

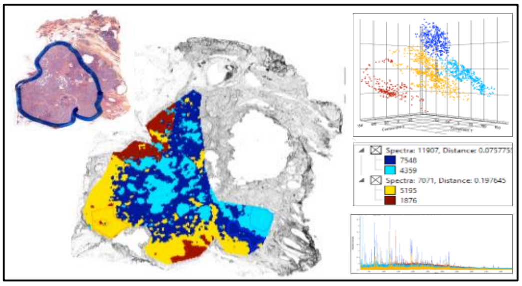

Case 39

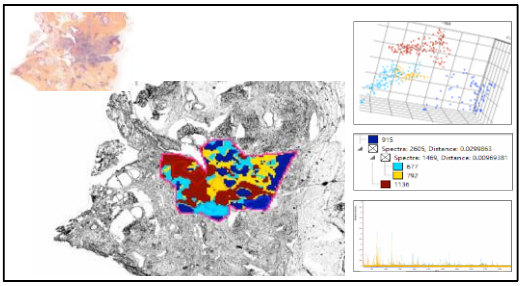

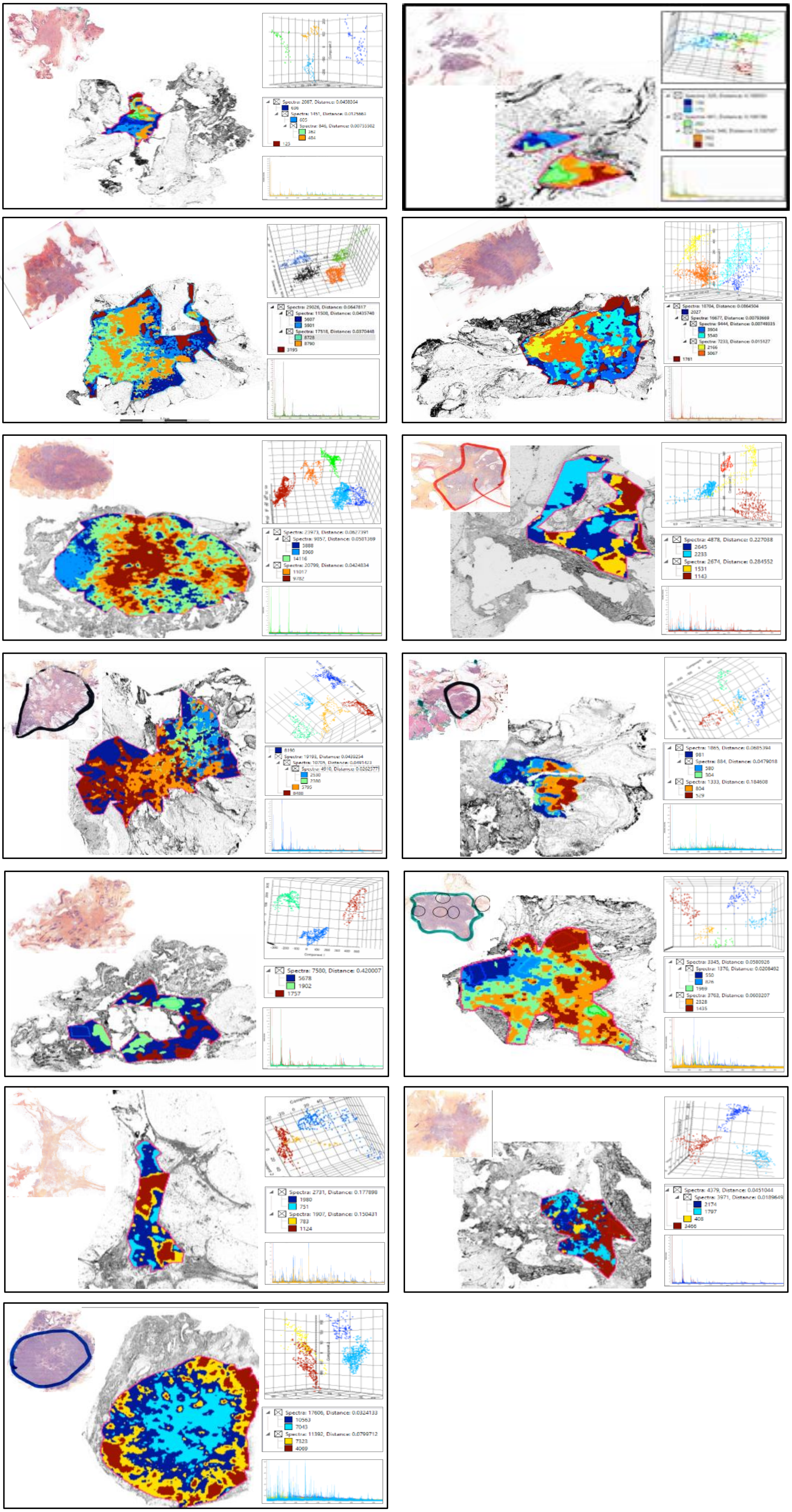

Case 40

Case 47

Case 41

Case 48

Case 42

Case 49

Case 43

Case 50

Case 44

Case 51

Case 45

Case 52

Case 46

Supplemental material 5

Case 1

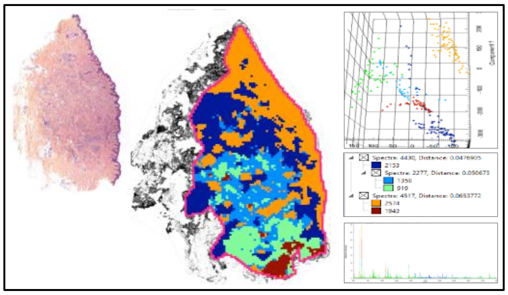

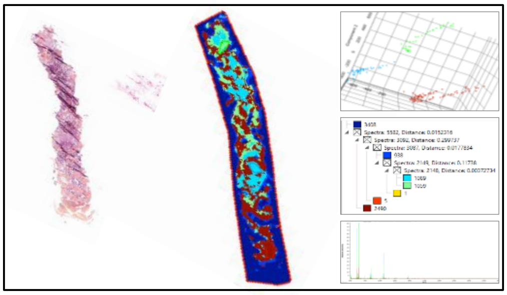

Case 7

Case 2

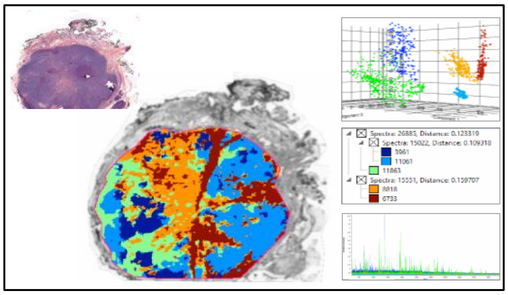

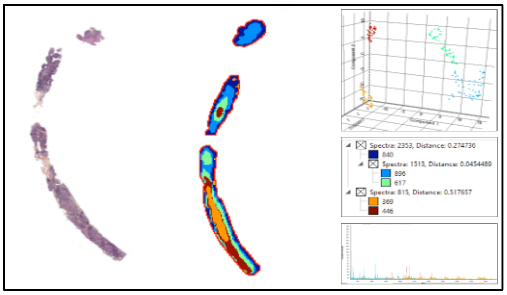

Case 8

Case 3

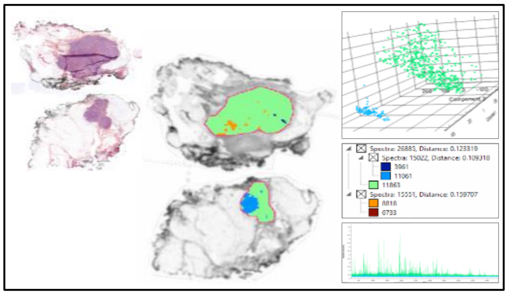

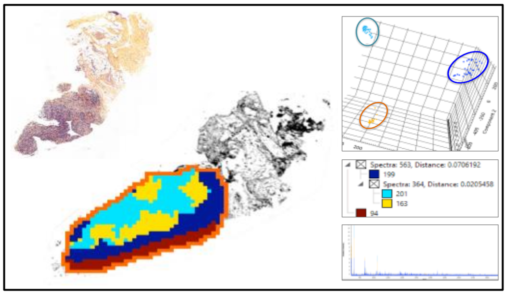

Case 9

Case 4

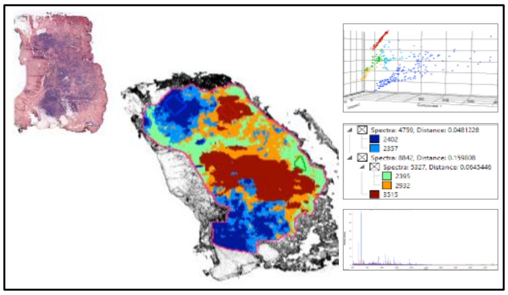

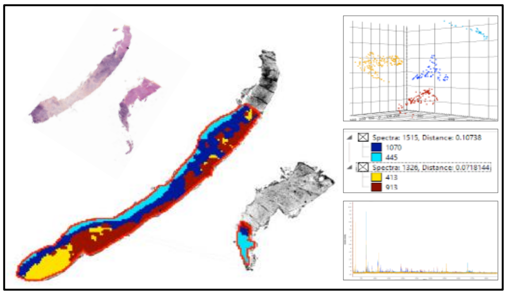

Case 10

Case 5

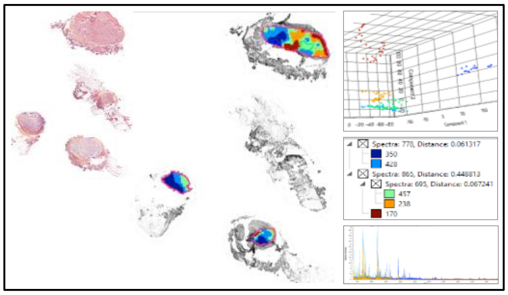

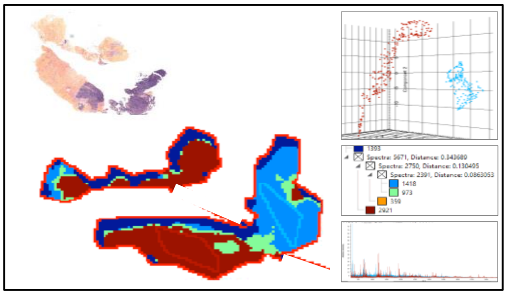

Case 11

Case 6

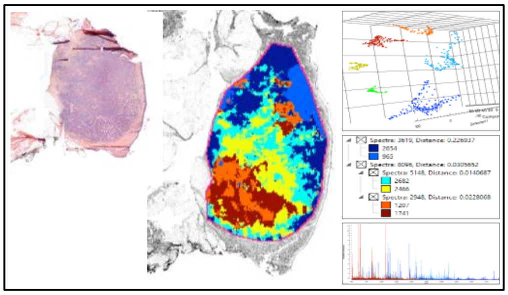

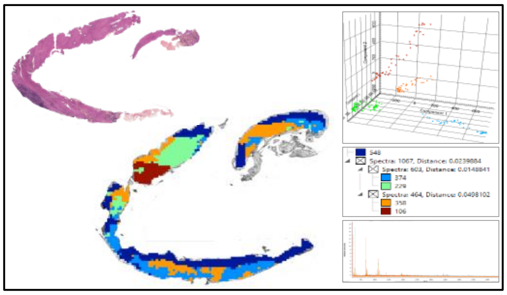

Case 12

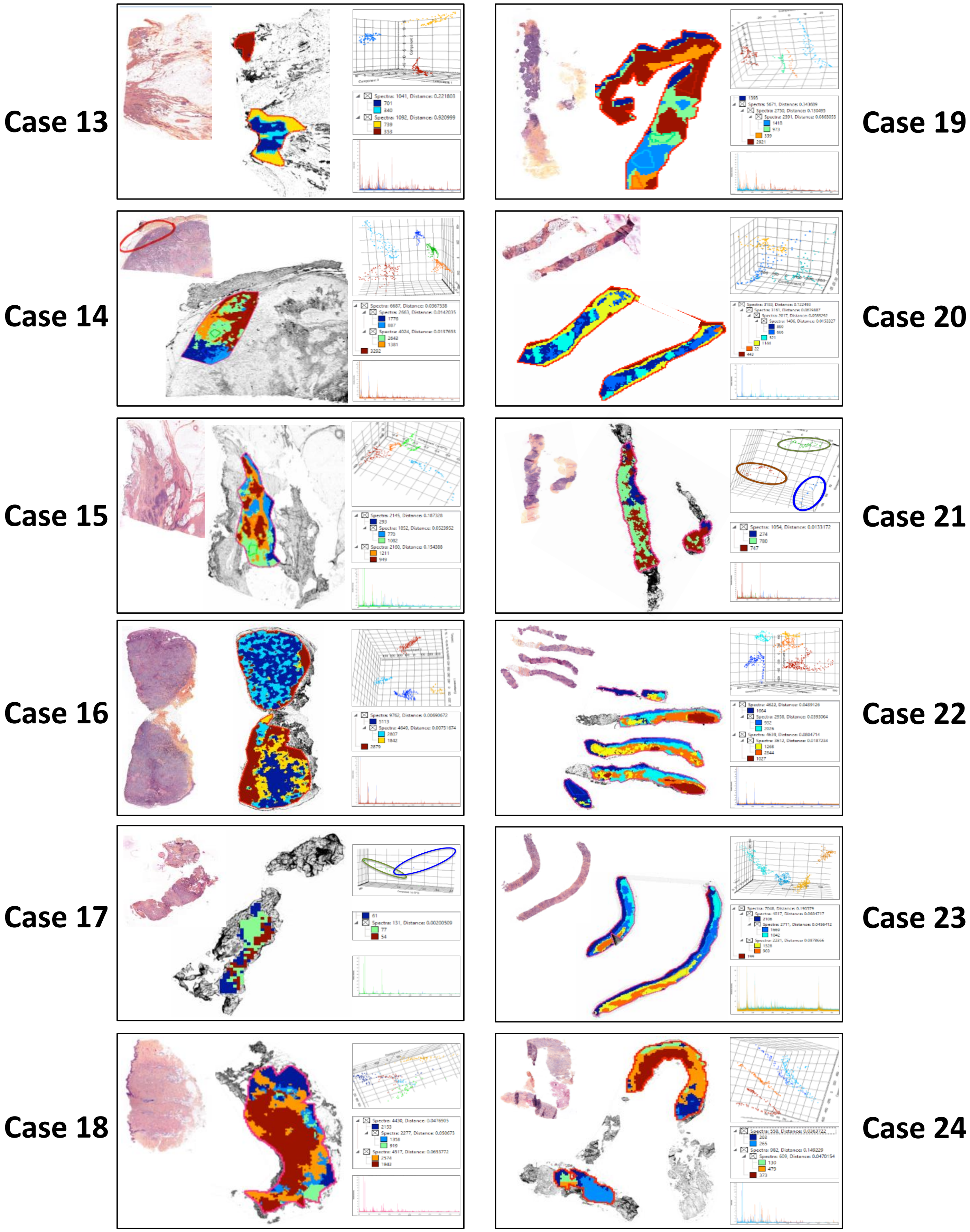
Case 13
Case 19
Case 14
Case 20
Case 15
Case 21
Case 16
Case 22
Case 17
Case 23
Case 18
Case 24

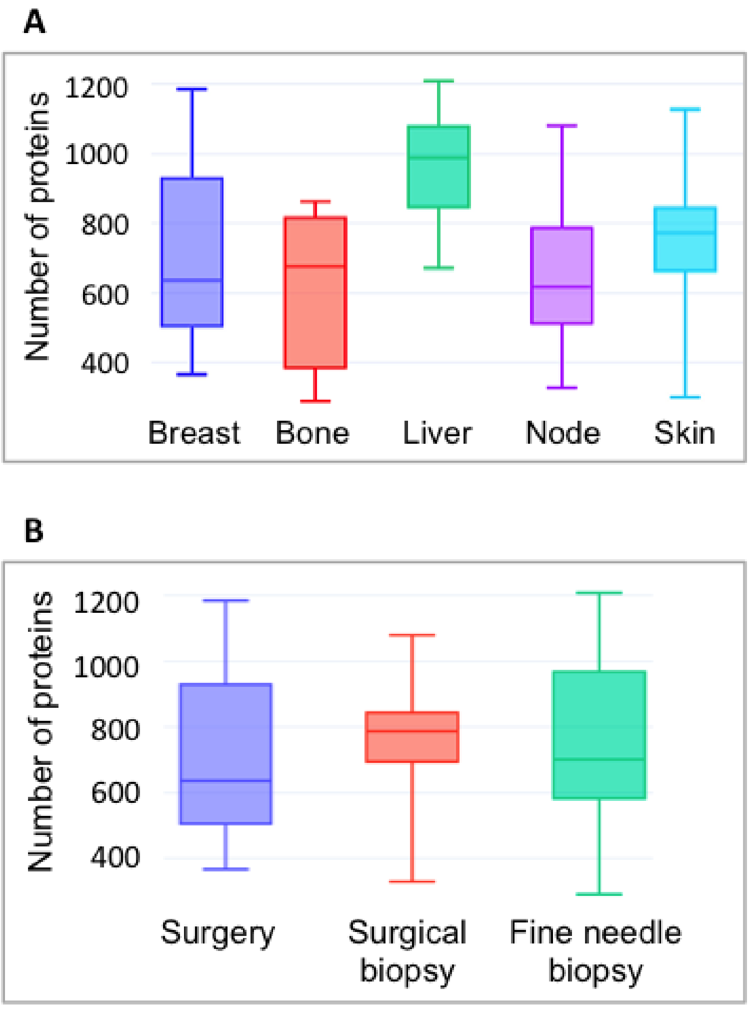
A
Number of proteins
1200
1000
800
600
400
Breast
Bone
Liver
Node
Skin
B
Number of proteins
1200
1000
800
600
400
Surgery
Surgical biopsy
Fine needle biopsy

Supplemental material 7

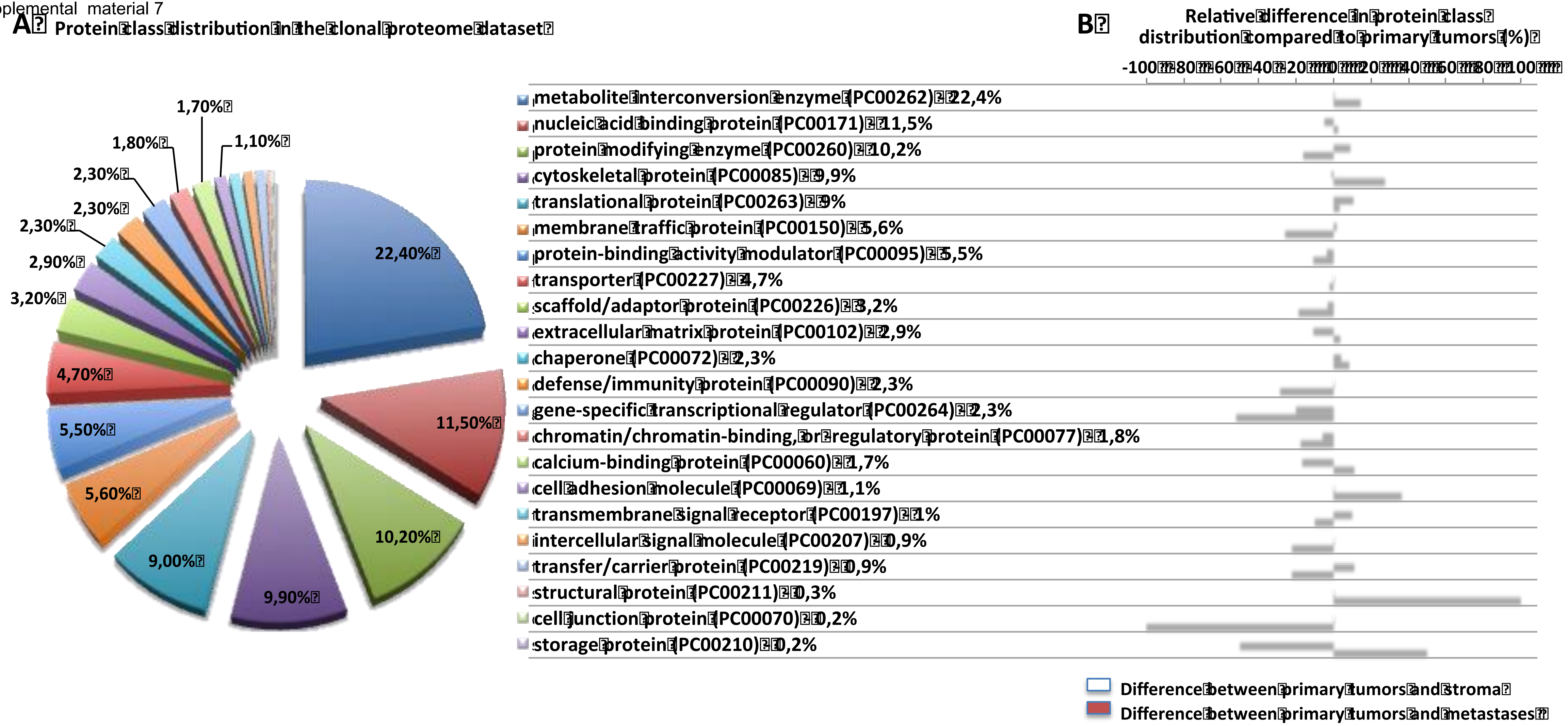

Supplemental material 8

| Gene name | Clonal proteome primary tumors | Clonal proteome metastases | TCGA EBC | TCGA metastases | BC360 | CDx |
|---|---|---|---|---|---|---|
| CDH1 | ■ | ■ | ■ | ■ | ■ | ■ |
| GATA3 | ■ | ■ | ■ | ■ | ■ | ■ |
| ERBB2 | ■ | ■ | ■ | ■ | ■ | ■ |
| PPP2R1A | ■ | ■ | ■ | ■ | ■ | ■ |
| SDHA | ■ | ■ | ■ | ■ | ■ | ■ |
| TFRC | ■ | ■ | ■ | ■ | ■ | |
| HLA-A | ■ | ■ | ■ | ■ | ■ | |
| HMGA1 | ■ | ■ | ■ | ■ | ■ | |
| SF3B1 | ■ | ■ | ■ | ■ | | ■ |
| SMARCA4 | ■ | ■ | ■ | ■ | | ■ |
| GNAS | ■ | ■ | ■ | ■ | | ■ |
| PRKAR1A | ■ | ■ | ■ | ■ | | ■ |
| FUBP1 | ■ | ■ | ■ | ■ | | ■ |
| IDH1 | ■ | ■ | ■ | ■ | | ■ |
| FH | ■ | ■ | ■ | ■ | | ■ |
| RAC1 | ■ | ■ | ■ | ■ | | ■ |
| NPM1 | ■ | ■ | ■ | ■ | | ■ |
| IDH2 | ■ | ■ | ■ | ■ | | ■ |
| RNF213 | ■ | ■ | ■ | ■ | | |
| MYH9 | ■ | ■ | ■ | ■ | | |
| MLLT4 | ■ | ■ | ■ | ■ | | |
| TPR | ■ | ■ | ■ | ■ | | |
| CHD4 | ■ | ■ | ■ | ■ | | |
| PDE4DIP | ■ | ■ | ■ | ■ | | |
| MYH11 | ■ | ■ | ■ | ■ | | |
| RANBP2 | ■ | ■ | ■ | ■ | | |
| PTPRC | ■ | ■ | ■ | ■ | | |
| NUMA1 | ■ | ■ | ■ | ■ | | |
| CLTC | ■ | ■ | ■ | ■ | | |
| EIF4A2 | ■ | ■ | ■ | ■ | | |
| KTN1 | ■ | ■ | ■ | ■ | | |
| DDX5 | ■ | ■ | ■ | ■ | | |
| COL1A1 | ■ | ■ | ■ | ■ | | |

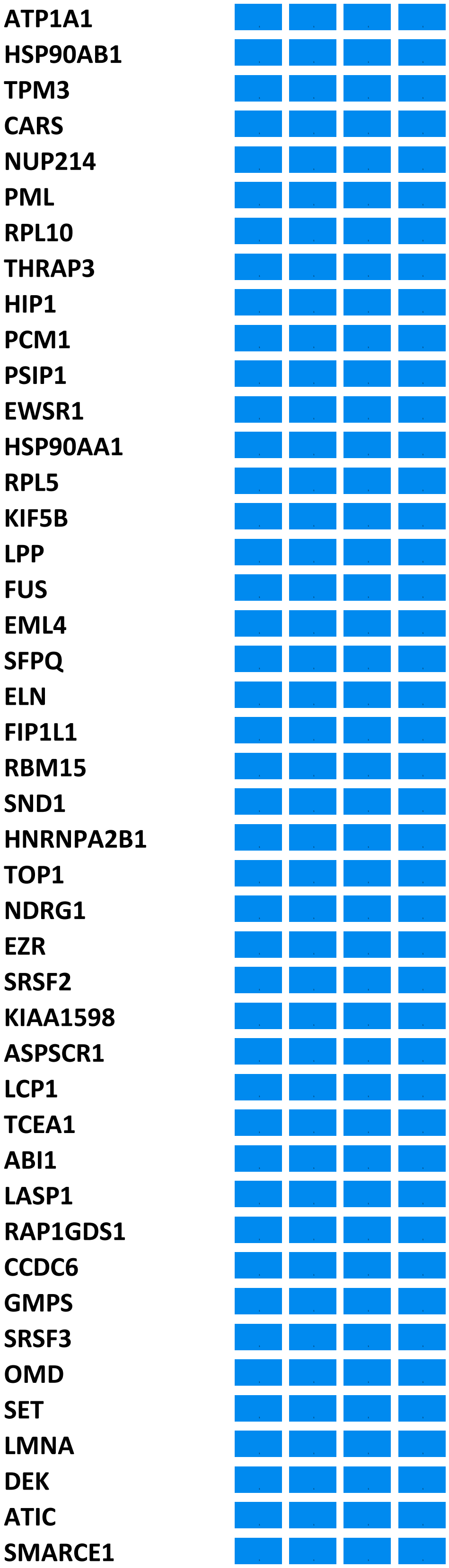
ATP1A1
HSP90AB1
TPM3
CARS
NUP214
PML
RPL10
THRAP3
HIP1
PCM1
PSIP1
EWSR1
HSP90AA1
RPL5
KIF5B
LPP
FUS
EML4
SFPQ
ELN
FIP1L1
RBM15
SND1
HNRNPA2B1
TOP1
NDRG1
EZR
SRSF2
KIAA1598
ASPSCR1
LCP1
TCEA1
ABI1
LASP1
RAP1GDS1
CCDC6
GMPS
SRSF3
OMD
SET
LMNA
DEK
ATIC
SMARCE1

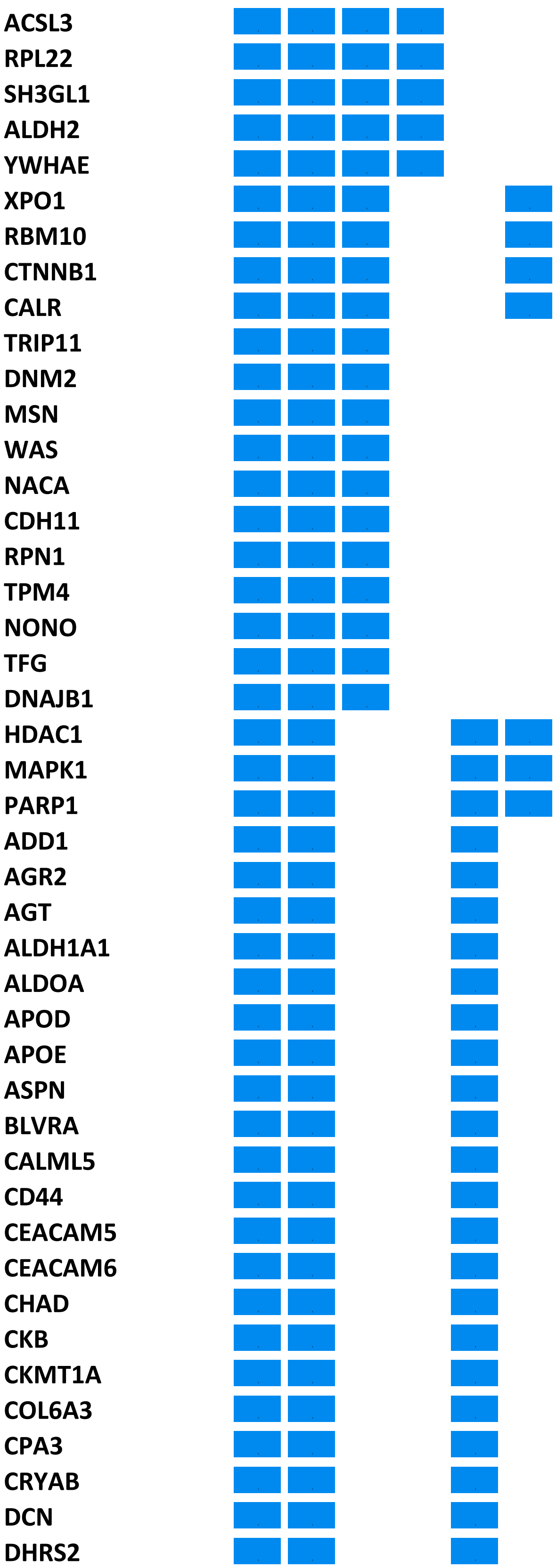
ACSL3
RPL22
SH3GL1
ALDH2
YWHAE
XPO1
RBM10
CTNNB1
CALR
TRIP11
DNM2
MSN
WAS
NACA
CDH11
RPN1
TPM4
NONO
TFG
DNAJB1
HDAC1
MAPK1
PARP1
ADD1
AGR2
AGT
ALDH1A1
ALDOA
APOD
APOE
ASPN
BLVRA
CALML5
CD44
CEACAM5
CEACAM6
CHAD
CKB
CKMT1A
COL6A3
CPA3
CRYAB
DCN
DHRS2

| |
|---|
| EIF3B |
| ENO1 |
| FBN1 |
| FHL1 |
| FLNC |
| GPX3 |
| HBB |
| HDAC2 |
| HIST1H1C |
| HLA-B |
| HLA-C |
| HLA-DRA |
| HLA-DRB1 |
| HSPA2 |
| ICAM1 |
| ITGA6 |
| ITGAV |
| ITGB1 |
| LAMB3 |
| LRP2 |
| MAPK3 |
| MAPT |
| MCM2 |
| MCM3 |
| MLPH |
| MMP14 |
| MMP9 |
| NASP |
| NAT1 |
| NPEPPS |
| OAS3 |
| OGN |
| PCNA |
| PFDN2 |
| PGK1 |
| PHGDH |
| PIP |
| PREP |
| PRKDC |
| PYCARD |
| RAC2 |
| RNASE2 |
| S100A14 |
| SERBP1 |

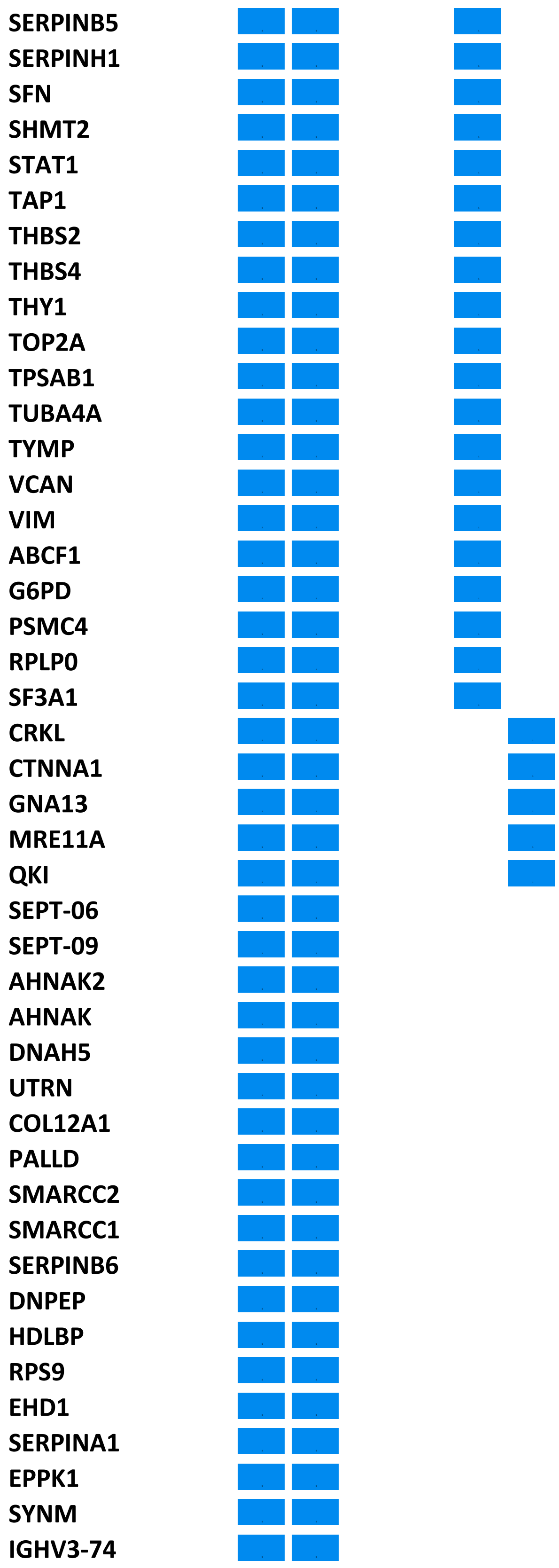

SERPINB5
SERPINH1
SFN
SHMT2
STAT1
TAP1
THBS2
THBS4
THY1
TOP2A
TPSAB1
TUBA4A
TYMP
VCAN
VIM
ABCF1
G6PD
PSMC4
RPLP0
SF3A1
CRKL
CTNNA1
GNA13
MRE11A
QKI
SEPT-06
SEPT-09
AHNAK2
AHNAK
DNAH5
UTRN
COL12A1
PALLD
SMARCC2
SMARCC1
SERPINB6
DNPEP
HDLBP
RPS9
EHD1
SERPINA1
EPPK1
SYNM
IGHV3-74

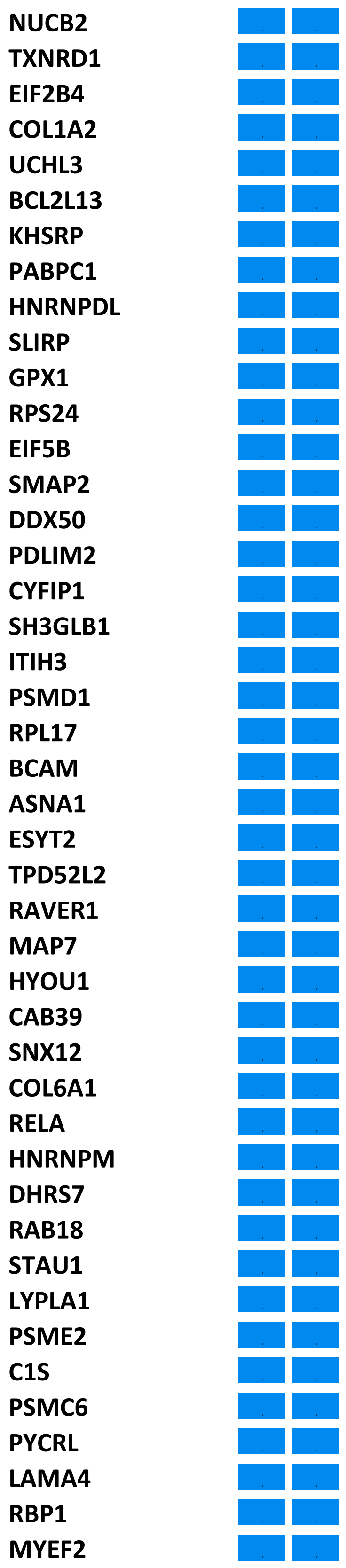
NUCB2
TXNRD1
EIF2B4
COL1A2
UCHL3
BCL2L13
KHSRP
PABPC1
HNRNPDL
SLIRP
GPX1
RPS24
EIF5B
SMAP2
DDX50
PDLIM2
CYFIP1
SH3GLB1
ITIH3
PSMD1
RPL17
BCAM
ASNA1
ESYT2
TPD52L2
RAVER1
MAP7
HYOU1
CAB39
SNX12
COL6A1
RELA
HNRNPM
DHRS7
RAB18
STAU1
LYPLA1
PSME2
C1S
PSMC6
PYCRL
LAMA4
RBP1
MYEF2

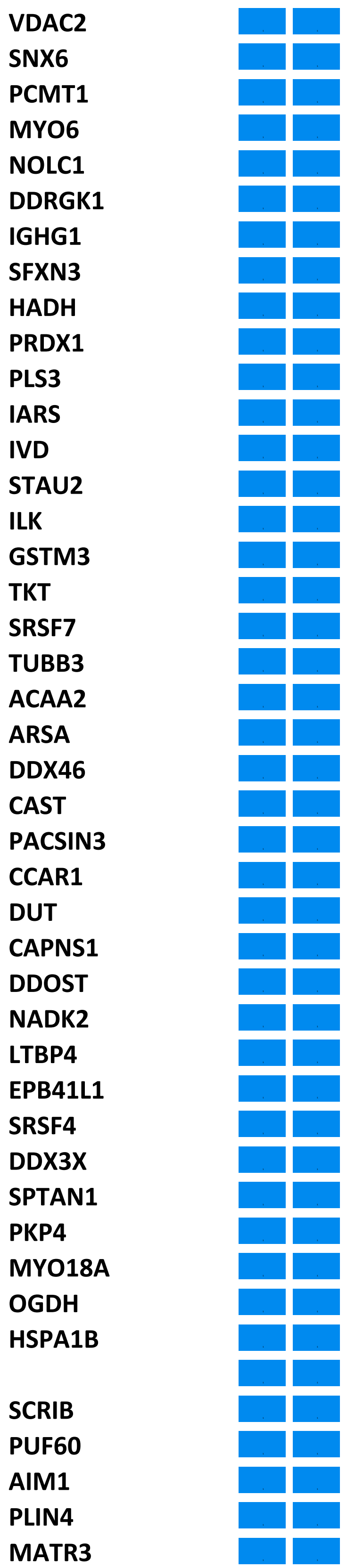
VDAC2
SNX6
PCMT1
MYO6
NOLC1
DDRGK1
IGHG1
SFXN3
HADH
PRDX1
PLS3
IARS
IVD
STAU2
ILK
GSTM3
TKT
SRSF7
TUBB3
ACAA2
ARSA
DDX46
CAST
PACSIN3
CCAR1
DUT
CAPNS1
DDOST
NADK2
LTBP4
EPB41L1
SRSF4
DDX3X
SPTAN1
PKP4
MYO18A
OGDH
HSPA1B
SCRIB
PUF60
AIM1
PLIN4
MATR3

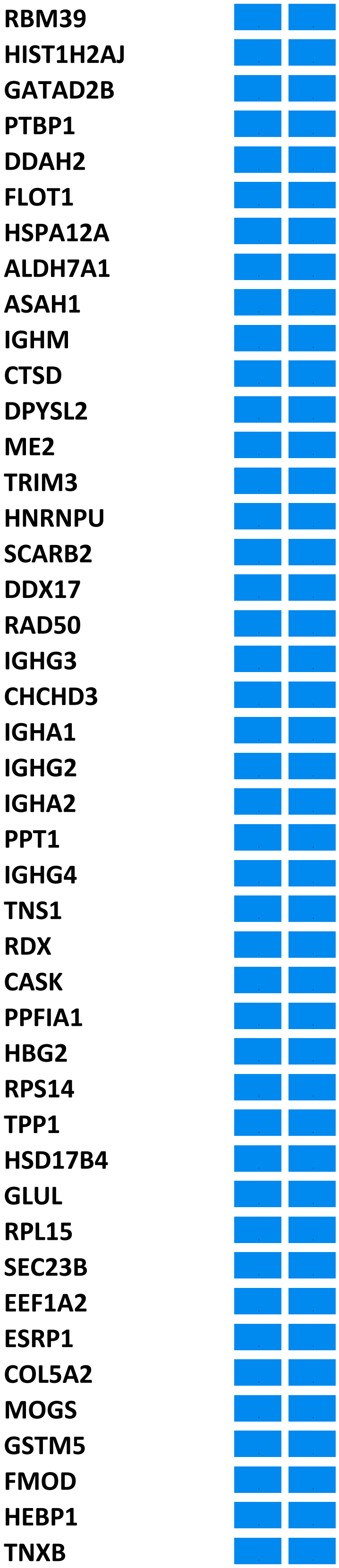
RBM39
HIST1H2AJ
GATAD2B
PTBP1
DDAH2
FLOT1
HSPA12A
ALDH7A1
ASAH1
IGHM
CTSD
DPYSL2
ME2
TRIM3
HNRNPU
SCARB2
DDX17
RAD50
IGHG3
CHCHD3
IGHA1
IGHG2
IGHA2
PPT1
IGHG4
TNS1
RDX
CASK
PPFIA1
HBG2
RPS14
TPP1
HSD17B4
GLUL
RPL15
SEC23B
EEF1A2
ESRP1
COL5A2
MOGS
GSTM5
FMOD
HEBP1
TNXB

SNTB1
IMPACT
PGM1
RAB2B
NHP2L1
EIF2B5
UBA6
ACO2
STMN1
RPS6
HN1L
USP14
RNPEP
TP53BP1
CRYZ
PPP2R4
HBE1
RPL23A
ALDOC
EIF3L
MB
UBE2I
CPNE1
ARCN1
MCM5
FBLN1
CAPZB
GNB1
MTR
PABPC4
HNRNPC
SEC14L2
CNN2
CS
EPS8L1
MRPL12
NQO1
OLFML3
STK24
ACTR3
FXR1
ILF2
CFB
CORO1C

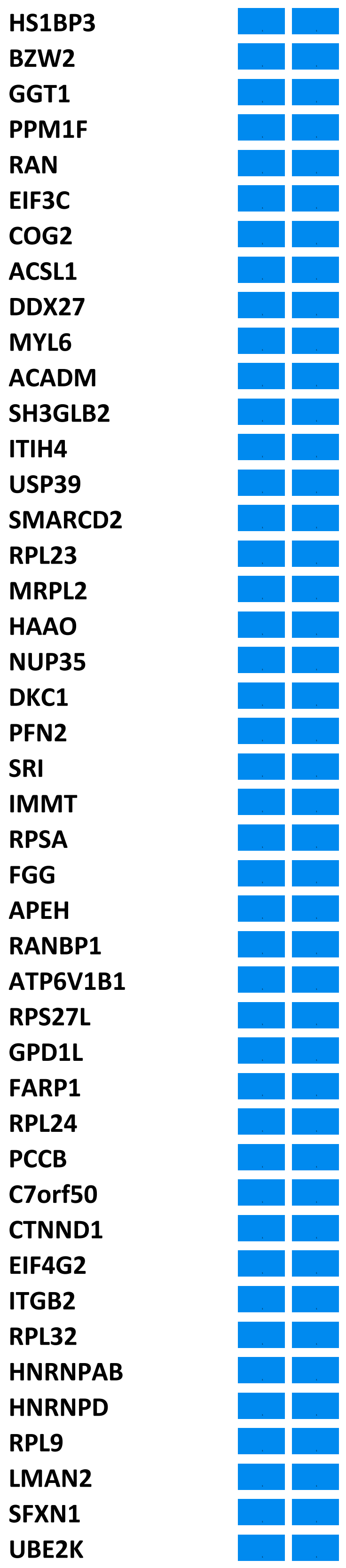
HS1BP3
BZW2
GGT1
PPM1F
RAN
EIF3C
COG2
ACSL1
DDX27
MYL6
ACADM
SH3GLB2
ITIH4
USP39
SMARCD2
RPL23
MRPL2
HAAO
NUP35
DKC1
PFN2
SRI
IMMT
RPSA
FGG
APEH
RANBP1
ATP6V1B1
RPS27L
GPD1L
FARP1
RPL24
PCCB
C7orf50
CTNND1
EIF4G2
ITGB2
RPL32
HNRNPAB
HNRNPD
RPL9
LMAN2
SFXN1
UBE2K

SEPT-11
SEC31A
GC
LARP7
CYB5B
ISOC1
ADH1B
EPHX2
TBCA
ERLIN2
MTDH
ANXA6
FLOT2
RAB1A
CCT5
RPL14
SEPT-07
NDUFV2
LTF
KIAA0196
ATP2B1
U2SURP
PLOD2
MARK2
SSFA2
EIF4G1
MAP4
HCLS1
CSTF2
EIF4B
RAB11FIP1
CALD1
PAICS
RBBP7
HSPA4L
MYO1B
DLAT
LRBA
DLD
TBL2
CP
MAP1A
SERPING1
COPS8

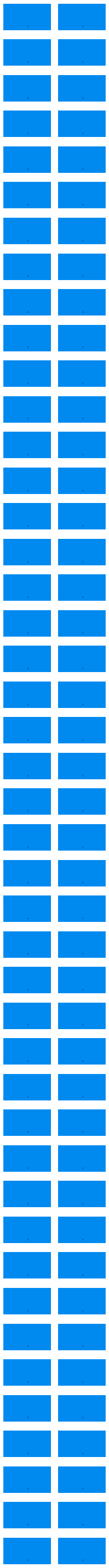

MCCC1
DYNC1LI1
LANCL1
F2
CLNS1A
UBTF
RPL27A
CSDE1
PSMC3
ARFGAP2
CSRP1
EXOSC4
SF3B2
FAU
SEC16A
PTGR1
CCT2
LAMTOR1
PSMD9
CHP1
PHB2
OTUB1
ATP6V0D1
SLC3A2
STARD10
MTHFD1
SEC23A
TSG101
ALDH16A1
NAP1L1
BCKDHA
MYO1C
IQGAP2
GNS
POLDIP3
CAD
C12orf10
UBE2N
SLC25A3
CSRP2
CNPY2
AK2
HNRNPA1
UBAP2L

GOLIM4
EIF2A
CPSF6
NDUFV1
CAPRIN1
DPP3
FTH1
RPL18
TCERG1
ERH
SNW1
APEX1
LTBP2
PNP
PSMA6
ZNF512
LAMB1
SLC12A2
PDE5A
SEC24C
HNRNPH1
DNM1L
IMPDH2
APMAP
DNPH1
SYNPO2
PRPF4B
RPS2
CRIP2
HADHA
CCDC86
SARNP
ETFA
IDH3A
ISLR
RMDN3
ANP32A
ANXA2
AKR7A2
RNPS1
RPS17
ABAT
HEXA
MACF1

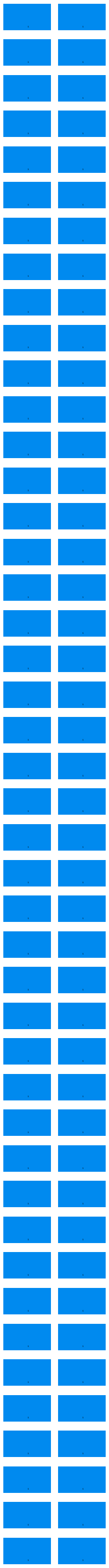

MMP2
HACD3
RBMX
PMM2
CIAPIN1
TPM1
PDIA3
ESD
PPP1R7
EBNA1BP2
RPL3
GIGYF2
NSF
HMOX2
MYBBP1A
PCYT2
CLUH
EIF5A
EIF4A3
LCMT1
VPS4A
TOR1AIP1
LUC7L3
FAM83H
OLA1
DCXR
MPRIP
ARHGDIA
SRSF1
MRPS23
PNPO
ALDH3A2
MYO1D
MYL12A
ZNF207
RSL1D1
BUB3
LONP1
FARSA
MRPL4
PPL
PRKCSH
EIF1
COPE

FBL
RPS11
ARHGEF1
ECH1
NAPA
CHMP2A
CLPP
PDLIM1
MYO1F
AIP
AP3B1
PSMD11
PSMD12
PGRMC1
CLIC1
EIF3F
QSOX1
WASL
AGRN
VWA5A
NOP56
MAN2B1
PDXK
PDCD5
SLC9A3R1
PSMA7
SCAMP3
OPLAH
IFIT3
HGS
PPP1R12A
SPTBN2
PFAS
ARPC1B
ARPC2
PGRMC2
LAMA5
SPTLC1
PPM1G
STX7
P4HA2
DHX15
PRPF4
DYNC1LI2

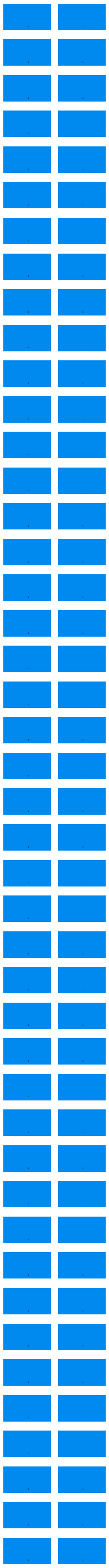

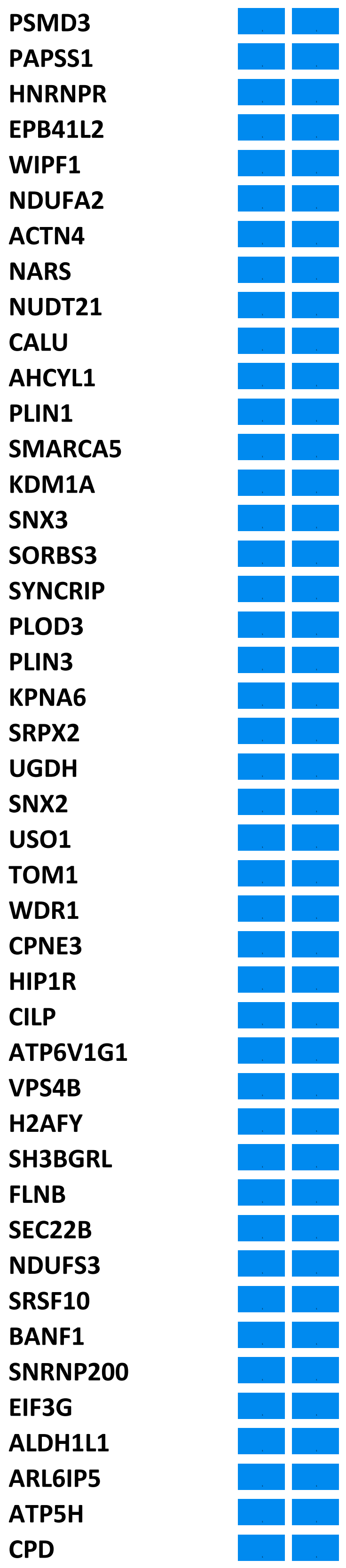

PSMD3
PAPSS1
HNRNPR
EPB41L2
WIPF1
NDUFA2
ACTN4
NARS
NUDT21
CALU
AHCYL1
PLIN1
SMARCA5
KDM1A
SNX3
SORBS3
SYNCRIP
PLOD3
PLIN3
KPNA6
SRPX2
UGDH
SNX2
USO1
TOM1
WDR1
CPNE3
HIP1R
CILP
ATP6V1G1
VPS4B
H2AFY
SH3BGRL
FLNB
SEC22B
NDUFS3
SRSF10
BANF1
SNRNP200
EIF3G
ALDH1L1
ARL6IP5
ATP5H
CPD

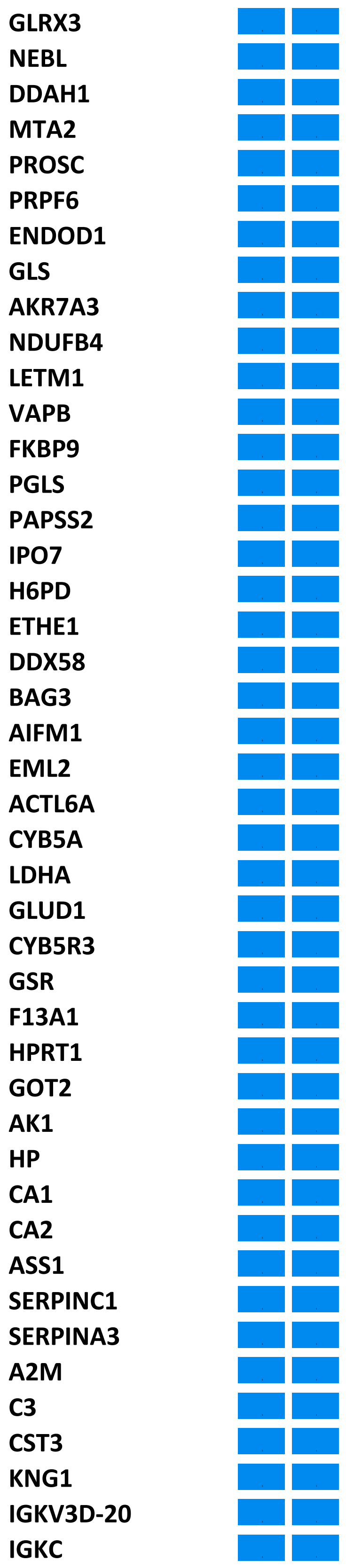
GLRX3
NEBL
DDAH1
MTA2
PROSC
PRPF6
ENDOD1
GLS
AKR7A3
NDUFB4
LETM1
VAPB
FKBP9
PGLS
PAPSS2
IPO7
H6PD
ETHE1
DDX58
BAG3
AIFM1
EML2
ACTL6A
CYB5A
LDHA
GLUD1
CYB5R3
GSR
F13A1
HPRT1
GOT2
AK1
HP
CA1
CA2
ASS1
SERPINC1
SERPINA3
A2M
C3
CST3
KNG1
IGKV3D-20
IGKC

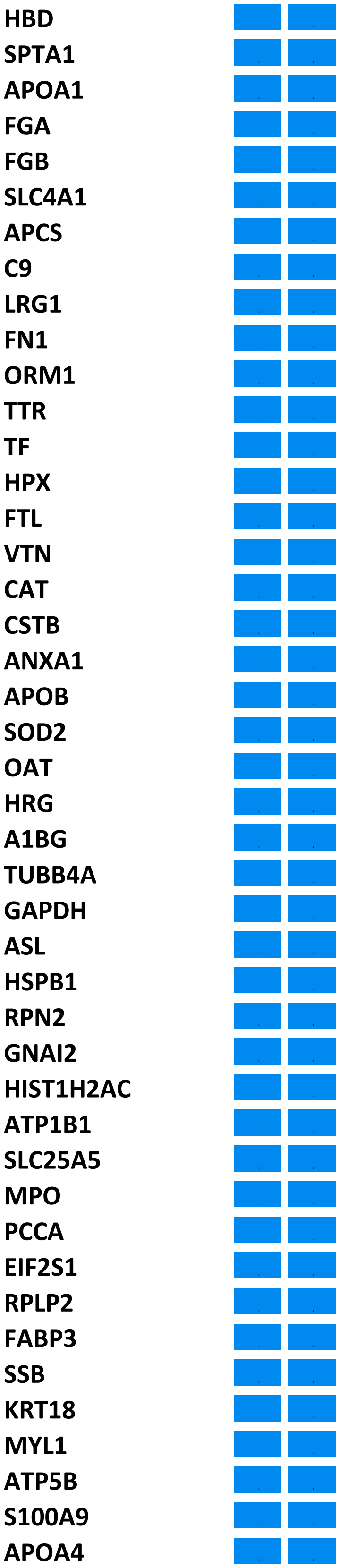
HBD
SPTA1
APOA1
FGA
FGB
SLC4A1
APCS
C9
LRG1
FN1
ORM1
TTR
TF
HPX
FTL
VTN
CAT
CSTB
ANXA1
APOB
SOD2
OAT
HRG
A1BG
TUBB4A
GAPDH
ASL
HSPB1
RPN2
GNAI2
HIST1H2AC
ATP1B1
SLC25A5
MPO
PCCA
EIF2S1
RPLP2
FABP3
SSB
KRT18
MYL1
ATP5B
S100A9
APOA4

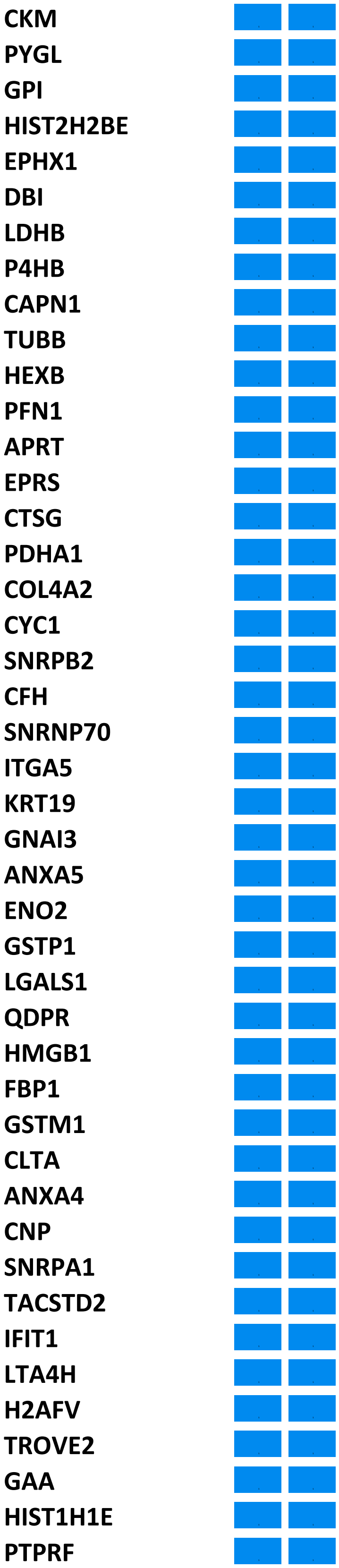
CKM
PYGL
GPI
HIST2H2BE
EPHX1
DBI
LDHB
P4HB
CAPN1
TUBB
HEXB
PFN1
APRT
EPRS
CTSG
PDHA1
COL4A2
CYC1
SNRPB2
CFH
SNRNP70
ITGA5
KRT19
GNAI3
ANXA5
ENO2
GSTP1
LGALS1
QDPR
HMGB1
FBP1
GSTM1
CLTA
ANXA4
CNP
SNRPA1
TACSTD2
IFIT1
LTA4H
H2AFV
TROVE2
GAA
HIST1H1E
PTPRF

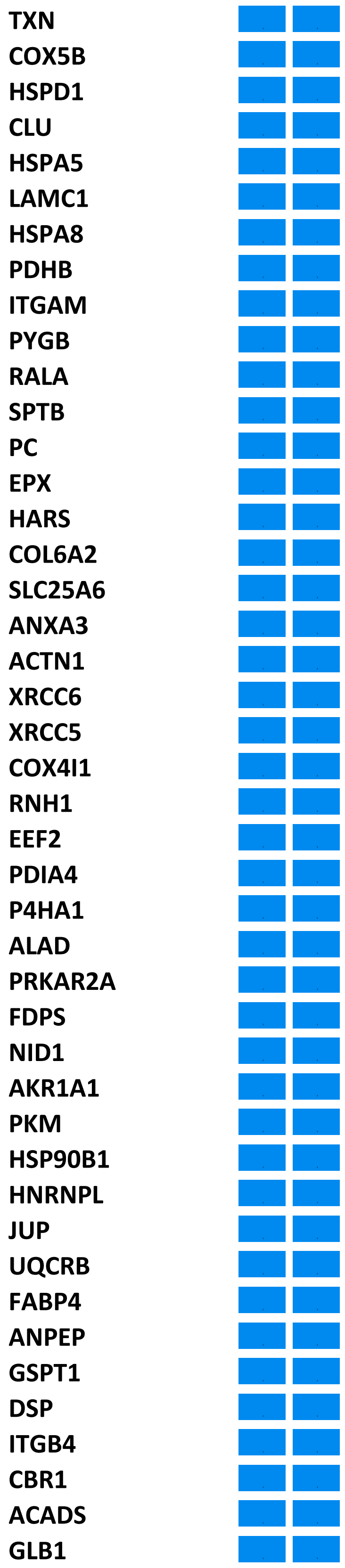
TXN
COX5B
HSPD1
CLU
HSPA5
LAMC1
HSPA8
PDHB
ITGAM
PYGB
RALA
SPTB
PC
EPX
HARS
COL6A2
SLC25A6
ANXA3
ACTN1
XRCC6
XRCC5
COX4I1
RNH1
EEF2
PDIA4
P4HA1
ALAD
PRKAR2A
FDPS
NID1
AKR1A1
PKM
HSP90B1
HNRNPL
JUP
UQCRB
FABP4
ANPEP
GSPT1
DSP
ITGB4
CBR1
ACADS
GLB1

HIST1H1B
HIST1H1D
POR
ATP2A2
FAH
YBX3
GOT1
CAPN2
DES
PFKL
LGALS3
TCP1
RPL35A
ITGB5
ARF4
RPL7
VCL
SON
PGAM1
RCC1
NCL
HK1
SRM
ORM2
ITIH1
NFKB1
EIF2S2
ANXA7
RAB5A
RAB6A
MX1
MX2
PSMB1
LMNB1
COL5A1
ATP6V1B2
FLNA
MAOA
ACO1
GPD1
VDAC1
BGN
TGM2
MUT

OSBP
GART
UBA1
ENPP1
FDXR
UQCRC2
CES1
PPIB
WARS
RPS3
AHCY
CFL1
EEF1B2
ATP5F1
ACP1
ACAT1
TNC
MYL9
MPZ
AZGP1
MPST
ATP5A1
PSMA1
U2AF2
RPL13
S100A4
HMGB2
TARS
VARS
EEF1G
STOM
PON1
MAOB
YWHAQ
RPA1
CANX
PSMA5
ABCD3
NDUFS1
LAP3
TPP2
IMPA1
CRABP2
LMOD1

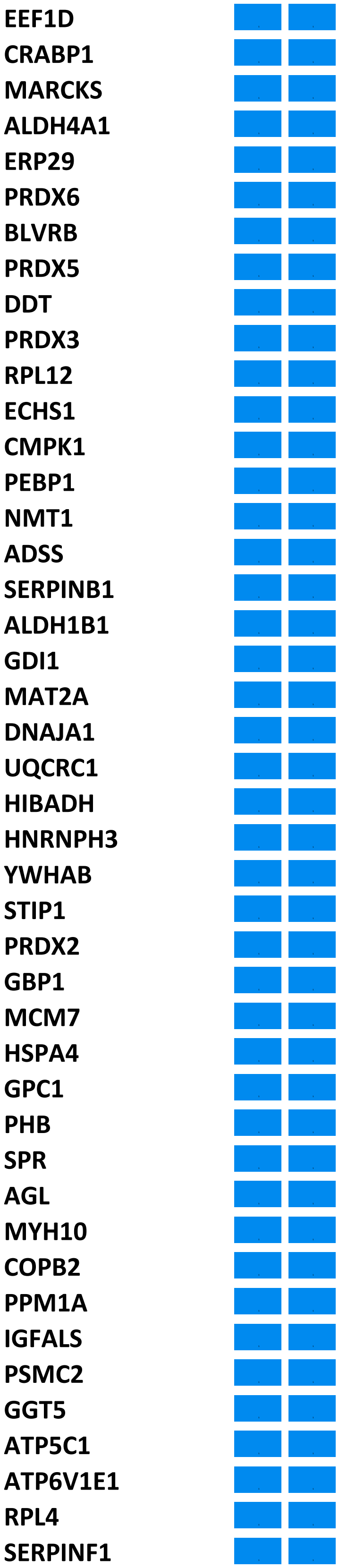
EEF1D
CRABP1
MARCKS
ALDH4A1
ERP29
PRDX6
BLVRB
PRDX5
DDT
PRDX3
RPL12
ECHS1
CMPK1
PEBP1
NMT1
ADSS
SERPINB1
ALDH1B1
GDI1
MAT2A
DNAJA1
UQCRC1
HIBADH
HNRNPH3
YWHAB
STIP1
PRDX2
GBP1
MCM7
HSPA4
GPC1
PHB
SPR
AGL
MYH10
COPB2
PPM1A
IGFALS
PSMC2
GGT5
ATP5C1
ATP6V1E1
RPL4
SERPINF1

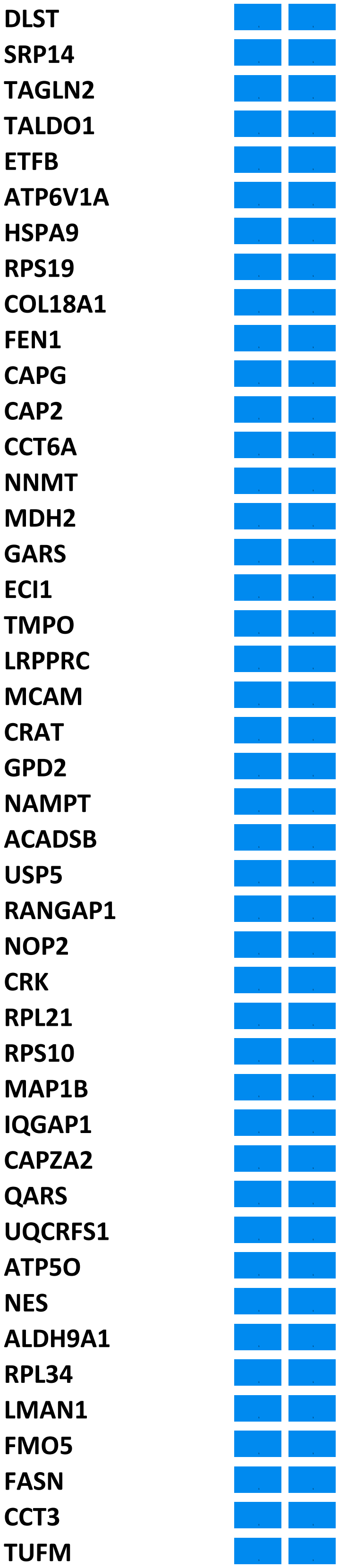
DLST
SRP14
TAGLN2
TALDO1
ETFB
ATP6V1A
HSPA9
RPS19
COL18A1
FEN1
CAPG
CAP2
CCT6A
NNMT
MDH2
GARS
ECI1
TMPO
LRPPRC
MCAM
CRAT
GPD2
NAMPT
ACADSB
USP5
RANGAP1
NOP2
CRK
RPL21
RPS10
MAP1B
IQGAP1
CAPZA2
QARS
UQCRFS1
ATP5O
NES
ALDH9A1
RPL34
LMAN1
FMO5
FASN
CCT3
TUFM

AARS
SARS
ACADVL
YLPM1
TMED10
RBM25
HINT1
NUP153
SEPHS1
LIG3
GDI2
EMD
SERPINB9
PDLIM4
ST13
VASP
CCT8
CCT4
ANXA11
RAB5C
RAB7A
GALK1
SSR4
MECP2
HCFC1
SGSH
HDGF
LUM
PRELP
CNN1
HNRNPA3
PGD
KPNA2
KPNA1
ARHGDIB
HNRNPF
HRSP12
CAPZA1
ARFIP1
ACLY
COPB1
COPA
SUB1
RARS

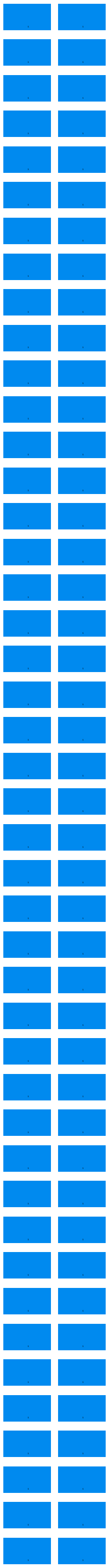

YARS
RAD23B
HMGCS2
ALDH18A1
EIF5
PSMD4
CSE1L
VCP
HADHB
ADAR
LAMB2
TPD52
HNRNPH2
MARS
ITGA1
EIF6
CTBP2
TPI1
EIF4A1
RAB2A
UBE2M
RAB14
ACTR2
ACTR1A
ARF1
RAP1B
RPS3A
MAGOH
RPL27
HSPE1
HNRNPK
YWHAG
RPS7
PPP1CA
PPP1CB
PSMC1
RPS8
RPS15A
RPS16
RPS18
RPS13
SNRPD2
SNRPD3
TMSB4X

RPL7A
ETF1
RPS4X
PPP2CA
ACTA2
HIST1H4A
RPS25
RPS28
RPL31
RPL10A
RPL11
RPL8
PPIA
FKBP1A
TRA2B
YWHAZ
RPL38
RPS21
GNB2L1
ACTG1
YBX1
ACTC1
UBE2L3
EEF1A1P5
TUBB4B
HBA1
GTF2I
GSTO1
BASP1
IGLV3-9
FBLN2
HSPG2
RBM3
FKBP3
SORD
SPTBN1
TIAL1
FABP5
CAP1
HMGCS1
PFKP
TAGLN
ALDH6A1
HIST1H1A

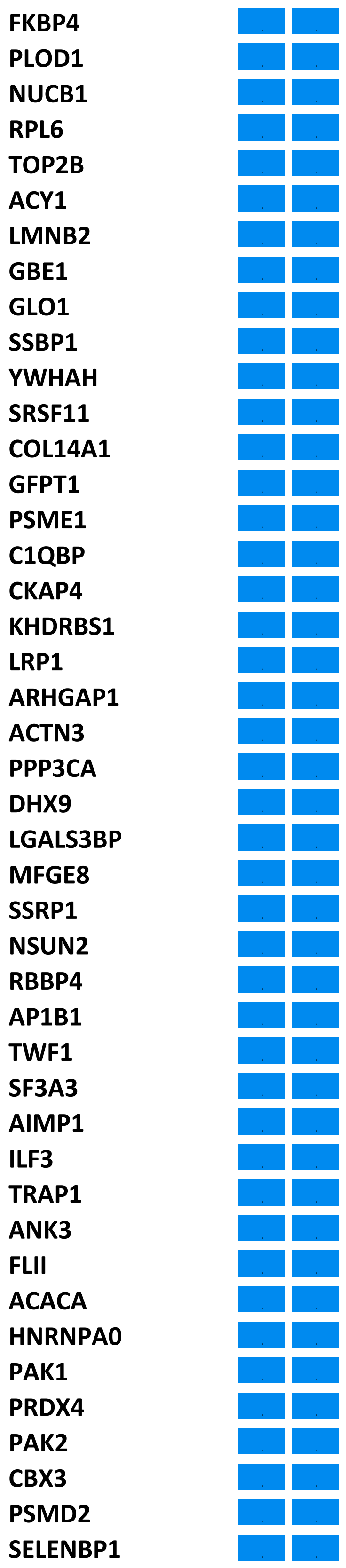
FKBP4
PLOD1
NUCB1
RPL6
TOP2B
ACY1
LMNB2
GBE1
GLO1
SSBP1
YWHAH
SRSF11
COL14A1
GFPT1
PSME1
C1QBP
CKAP4
KHDRBS1
LRP1
ARHGAP1
ACTN3
PPP3CA
DHX9
LGALS3BP
MFGE8
SSRP1
NSUN2
RBBP4
AP1B1
TWF1
SF3A3
AIMP1
ILF3
TRAP1
ANK3
FLII
ACACA
HNRNPA0
PAK1
PRDX4
PAK2
CBX3
PSMD2
SELENBP1

| |
|---|
| SRSF9 |
| SRSF5 |
| SRSF6 |
| TRIM28 |
| G3BP1 |
| PTK7 |
| EIF3I |
| CTBP1 |
| NNT |
| SNTB2 |
| PDAP1 |
| FKBP5 |
| PIN1 |
| DCTN2 |
| TRA2A |
| SNX1 |
| TSTA3 |
| TPBG |
| ALCAM |
| DDX39B |
| TUBB2A |
| CAPS |
| IL16 |
| CIRBP |
| NID2 |
| DSG2 |
| SAFB2 |
| EIF3A |
| TTLL12 |
| DPYSL3 |
| DYNC1H1 |
| CTTN |
| GALE |
| MCM6 |
| SMC1A |
| USP10 |
| GANAB |
| MVP |
| GOLGB1 |
| NFIX |
| KPNB1 |
| SEPT-02 |
| SART3 |
| EFTUD2 |

KARS
EIF4H
POSTN
EEA1
PDIA6
PAFAH1B3
PCOLCE
PMVK
PLEC
PPA1
RCN1
PCBP1
PCBP2
SF3B3
CNN3
SAFB
TGFBI
SLC9A3R2
SF1
MAPRE1
ELAVL1
MYLK
SLC1A5
TOMM34
UBE2V2
ZYX
PSMD5
ECM1
DBN1
FSCN1
DECR1
TST
PCK2
UGP2
AOC3
HNRNPUL2
INF2
TSR1
SMU1
LEPRE1
UAP1L1
DAK
ALDH1L2
TMEM119

FNDC1
FAM98B
PDCD4
PDLIM3
ACTBL2
HSP90AB2P
MAGED2
MIA3
PITRM1
VPS16
UCHL5
PSMF1
PHYHD1
HP1BP3
CHCHD2
IBA57
KIAA1217
C9orf64
NTPCR
DDI2
RPRD2
LYPLAL1
RBM17
KANK2
MAP1S
HIST2H2BD
TWF2
SLC25A24
PTRF
CC2D1A
PRPF8
PDXDC1
POTEE
CSPG4
NAPRT
CD109
IKBIP
TUBA1A
ZC3HAV1
NUP54
MYH14
MAVS
PODN
IRF2BP2

HUWE1
ACOT1
PABPN1
PHLDB1
FERMT3
ALYREF
CAND1
LSR
DDX42
AEBP1
LEPREL2
CARKD
FTSJ3
CCAR2
LRRC47
MRPL50
CPSF7
GOLM1
COLGALT1
TXNDC5
SCCPDH
LEMD2
APOA1BP
CHDH
NUP43
ABHD11
CMAS
GDAP1
AGR3
SSH3
NUP210
IPO4
DYNLRB1
LRRC15
PPP1R13L
NUP133
PDCD6IP
THEM6
SCFD1
UBLCP1
ATXN2L
PSPC1
DNAJC9
IRGQ

DDX1
H1FX
CCT6B
HSPH1
GCN1L1
PXDN
ANP32B
OSTF1
UFD1L
UPF1
GCDH
TNPO1
ATP2A3
TCEAL3
MAL2
IGSF8
ERGIC1
HIST1H2BA
LRRC59
FKBP10
EFHD2
GALM
DCPS
CCDC124
KCTD12
RCN3
PGM2
PDLIM5
ERO1L
PGAM5
FUBP3
SUCLG2
ABHD14B
CDK5RAP3
CLMN
FAM84B
CNDP2
ZFR
CLIC6
IPO9
RBM14
VPS35
GFM1
HMCN1

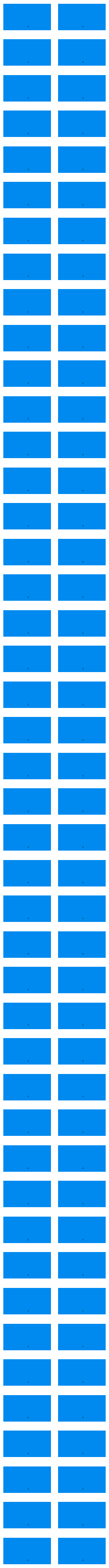

STRBP
CPXM1
FAM129B
CDC5L
PARK7
VAT1
HSD17B10
NAP1L4
CCT7
CORO1B
TXNDC17
ERP44
LXN
ESYT1
TMEM43
HGH1
TUBB6
C9orf142
HNRNPUL1
EFHD1
DDX23
GNL3
TPPP3
RBM4
SORBS1
CECR5
EMILIN2
FAM129A
API5
TNKS1BP1
XRN2
EHD4
PPA2
PNN
BOLA2
ACBD3
PTPN23
RPAP3
NO66
DCTPP1
GORASP2
MCCC2
PREB
ISYNA1

RTN4
RPRD1B
KIF13B
PDLIM7
DDX21
NANS
MXRA5
UBQLN4
PHPT1
FARSB
IARS2
ECHDC1
RBM12
FAM49B
SLTM
ADPRHL2
STAB1
BCLAF1
TMOD3
UGGT1
FAM120A
MAT2B
MYOF
EHD2
LMCD1
MRPL15
VAPA
H2AFY2
CAMSAP3
KIAA1522
RCC2
RRBP1
LARS
CGN
MRC2
GRHPR
DNAJB11
SUN2
LIMA1
TRPS1
DPP7
CYB5R1
AK3
NAGK

SLC25A13
DBNL
STOML2
APPL1
RALY
PRPF19
UBQLN1
NENF
STUB1
NOVA2
NSFL1C
SRRM2
PA2G4
BAIAP2
SMC3
RUVBL2
CLEC11A
RUVBL1
PADI2
CARHSP1
NOP58
LUC7L2
SF3B6
STRAP
RTCB
RPL36
SAMHD1
PKP3
TLN1
TLN2
AFG3L2
SUPT16H
CD2AP
RBM8A
SNX9
HEBP2
COPG1
CLIC4
NFS1
EMILIN1
SQRDL
SCIN
WASF2
SEC23IP

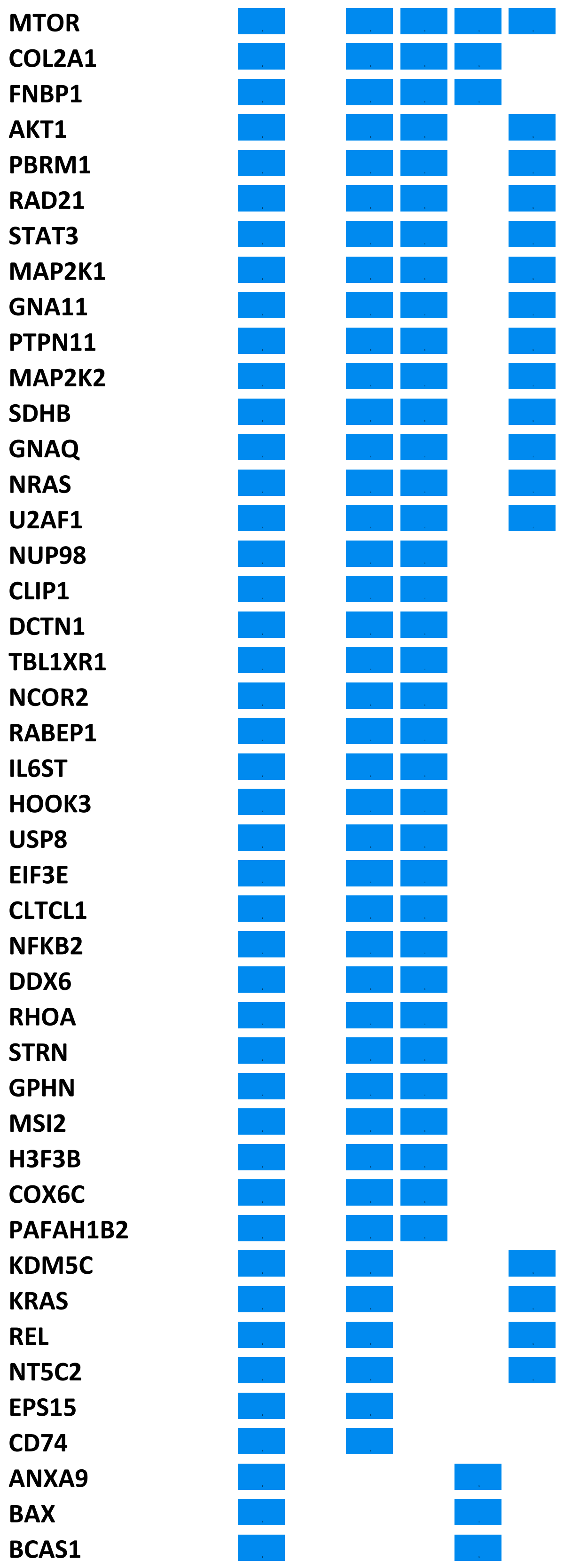
MTOR
COL2A1
FNBP1
AKT1
PBRM1
RAD21
STAT3
MAP2K1
GNA11
PTPN11
MAP2K2
SDHB
GNAQ
NRAS
U2AF1
NUP98
CLIP1
DCTN1
TBL1XR1
NCOR2
RABEP1
IL6ST
HOOK3
USP8
EIF3E
CLTCL1
NFKB2
DDX6
RHOA
STRN
GPHN
MSI2
H3F3B
COX6C
PAFAH1B2
KDM5C
KRAS
REL
NT5C2
EPS15
CD74
ANXA9
BAX
BCAS1

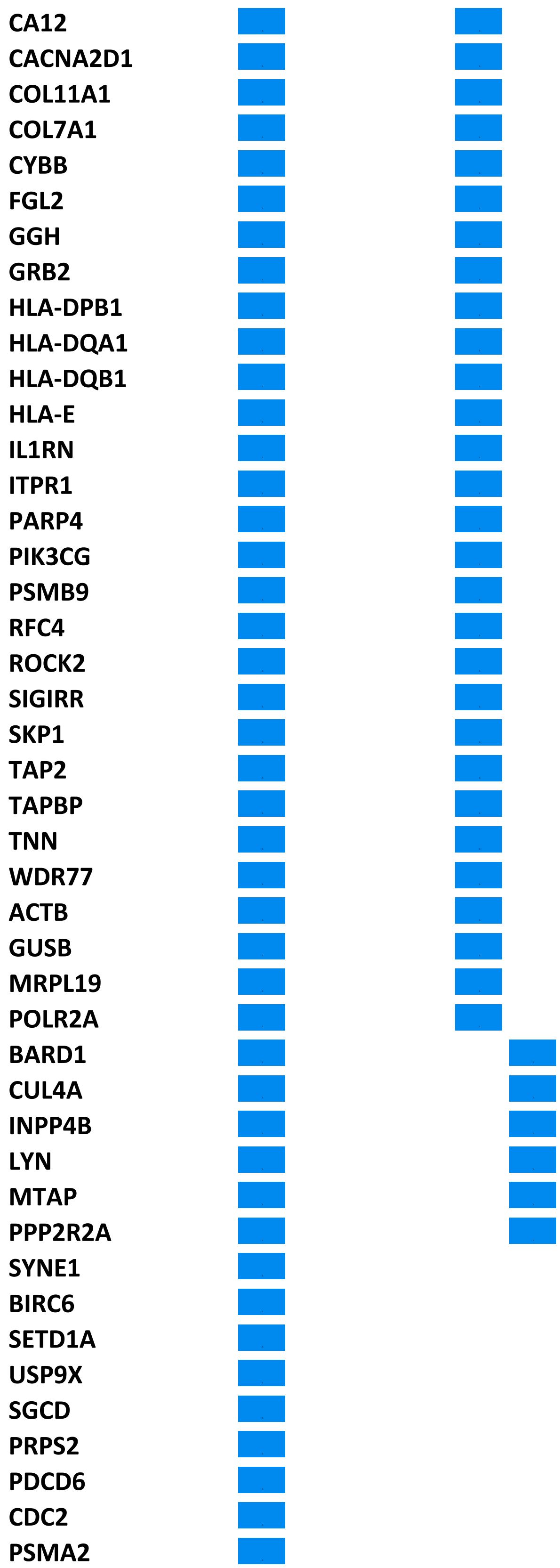
CA12
CACNA2D1
COL11A1
COL7A1
CYBB
FGL2
GGH
GRB2
HLA-DPB1
HLA-DQA1
HLA-DQB1
HLA-E
IL1RN
ITPR1
PARP4
PIK3CG
PSMB9
RFC4
ROCK2
SIGIRR
SKP1
TAP2
TAPBP
TNN
WDR77
ACTB
GUSB
MRPL19
POLR2A
BARD1
CUL4A
INPP4B
LYN
MTAP
PPP2R2A
SYNE1
BIRC6
SETD1A
USP9X
SGCD
PRPS2
PDCD6
CDC2
PSMA2

| |
|---|
| BAG6 |
| IGLV3-10 |
| IGKV2-40 |
| LTV1 |
| KIAA1468 |
| IGHV3OR15-7 |
| GPX4 |
| TMC4 |
| PYCR2 |
| UBXN1 |
| GMPPA |
| PSMB3 |
| PEG10 |
| SYNJ2BP-COX16 |
| PRODH |
| THOC1 |
| TARDBP |
| NDUFA10 |
| RMND1 |
| Em:AP000351.3 |
| AP2M1 |
| F11R |
| FAM21A |
| FBN2 |
| CCDC47 |
| EIF3H |
| IDH3B |
| COL15A1 |
| SEPT-08 |
| ARFIP2 |
| NDUFA5 |
| AP2B1 |
| PTGS1 |
| SPAG9 |
| C21orf33 |
| EPB41L3 |
| OGFR |
| SMARCA1 |
| IGKV3D-11 |
| MFF |
| ACE |
| AKR1C3 |
| NUDC |
| C1QB |

PITPNB
NELFE
ATP1A3
SCN5A
FKBP15
HYI
SULT1A1
CORO7-PAM16
ACSS3
ARHGAP4
TRPV1
RPS15
IGHV3OR16-9
CYP51A1
TM9SF4
DLG1
NELFA
ECI2
GIT1
CDK11B
TLE4
NBEA
PNKP
UBE3A
HLA-F
DIAPH1
PPP1R18
ABHD16A
HLA-DRB4
C4B
GBA
MUC2
HLA-G
ADI1
PANK4
CELF2
PON2
CLASP2
EFEMP1
PACSIN2
ELF1
SPATA20
ENAH
HLA-DRB3

CPT2
PXN
PLEKHA6
EPB41
UNC45A
POLR2B
NFIC
GOLGA2
CCDC9
CSNK2A1
POLR1C
SHROOM3
ADNP
PFKM
DOCK10
GCLC
AP1S1
SSR1
TCOF1
ECE1
IDE
NDUFA9
ANKS1A
EPS8L2
C1R
AGK
PARP14
MYO1G
AMDHD2
VPS13C
TRIM25
GLA
PRPF40A
RBM47
IGLC6
ILVBL
SH3PXD2B
MPG
MRPL28
VWA8
CNOT1
PGP
NUDT5
FBLL1

| |
|---|
| SMCHD1 |
| NCF1C |
| EIF1AY |
| RGPD3 |
| ENSA |
| COL6A6 |
| SHROOM1 |
| MANF |
| FMR1 |
| GNAT3 |
| SNRPG |
| SRRM1 |
| EIF3D |
| HMOX1 |
| STX12 |
| MRPL43 |
| CTTNBP2NL |
| SAE1 |
| BCAT2 |
| POLDIP2 |
| PACS1 |
| COPS2 |
| ANXA8L1 |
| METTL3 |
| PTK2 |
| STK26 |
| OS9 |
| SEPT-10 |
| MAPK14 |
| EML3 |
| HNRNPLL |
| RIC8B |
| HBS1L |
| RANBP3 |
| SLMAP |
| PTMA |
| TCEB2 |
| WASH3P |
| DISC1 |
| BZW1 |
| PCYT1A |
| BCAP31 |
| EEF1E1 |
| ARF5 |

BRE
CROT
MYADM
AARSD1
ALDH5A1
NHLRC3
JCHAIN
ATP1B3
TBC1D15
SP100
ACAA1
NSDHL
CYCS
MRPL39
GNB2
PSAP
MRPS34
ATP5J2-PTCD1
ALB
MTPN
STEAP4
TIA1
LBR
IGFBP5
SPTBN4
UBE2H
ABI3BP
CAMK2D
ABCE1
FBLIM1
COPS4
OCIAD1
IDUA
CD14
ERAP2
MRPS27
SETD7
RMDN1
TCEB1
SGK3
RPL30
RPL26
SWAP70
ITGA2

LSP1
TACC2
DYNC1I2
HMGB3
IK
ERMP1
MTA1
MRPL3
ARAP1
KLC1
CYFIP2
NIF3L1
NLN
MAN2B2
SRP9
PPIE
NUP155
GSTK1
PPP2R5D
TSN
NDUFB9
SUN1
GSTM2
ACP2
GSDMD
NEU1
TCP11L1
PARP10
PRMT1
NEDD8-MDP1
RRM1
RAB1B
CSTF3
TCIRG1
C11orf58
STIM1
PPFIBP2
ANP32E
RAE1
PFDN4
PARVA
CHMP4A
TOLLIP
RPL35

PITPNA
ADA
SPARC
REXO2
PAAF1
HMHA1
MLEC
UBC
TUBA1B
DHX8
DDX19A
TINAGL1
PDE12
GLOD4
RBM38
PIP4K2C
FKBP11
TNS2
COPZ1
PACS2
SMPDL3B
VPS29
PPP1CC
C17orf49
MMP19
UBA3
ELMO3
ARPC4-TTLL3
B3GAT3
CASP1
TJP1
HBA2
NUP107
ATP6V1D
OSGEP
DGKA
GMFB
FBLN5
PSMA3
PRMT5
MIA2
TRAM1
MRPS22
NELFCD

AMPD2
DPM1
EIF2B3
GOLGA4
NCSTN
CALM2
TLR9
IPO5
TTC1
POLB
PPP5C
RSF1
MSI1
YARS2
RPL28
CERS2
PSMA4
ANKRD17
ISG20
SEC11A
VPS13D
NOL3
ARPP19
IST1
FAM96B
COX5A
AMFR
BCKDK
CSNK2A2
RHOT2
HAPLN3
PAGR1
NOB1
GCA
ABCB6
DHX30
RHOT1
NUPL1
PPIL3
RETSAT
HGD
IAH1
TOM1L1
DPF2

ECM29
CCS
SUMF2
APOL2
CEP170B
ICT1
BLMH
RPL19
CHERP
SCRN2
SNRPN
VWA1
MRPS7
TUBG1
PSMD8
TMEM205
GIPC1
GLYR1
PSME3
CILP2
EIF3K
ICAM3
MFAP4
DUSP3
GHDC
DCAKD
SARS2
MYDGF
FCGRT
DTNA
GGCT
RPS5
TRIP10
EPS15L1
RPL18A
SNRPA
EMC10
AGPS
ACOT7
SNAP23
GTPBP1
LGALS9
STXBP3
SULT2B1

DFFA
SULT1C2
MATN2
PSMD14
SDCBP
PIR
ACACB
SCD
SLC27A2
GAK
PLXNB2
DCLK1
PFDN6
ZNF185
MAPK13
OGT
YKT6
ARPC5
NDUFS4
SART1
TXNL1
PI15
AP1G1
SGTA
GMDS
EXOC3
LPXN
HIST1H2BN
CCDC22
PQBP1
EDF1
KERA
DNAJC13
ADAMTS4
DNAJB6
NDUFS2
GBAS
VPS26A
PMPCB
ERLIN1
PRKRA
SLC25A12
PALM
GRAP2

EIF3J
CBR3
BCAS2
SASH3
WFS1
CLPX
SNCG
SRP72
STK10
TOMM70A
SEC24D
UFL1
SORBS2
UBXN7
AP2A2
HEXIM1
TJP3
NDUFB8
SNAPIN
PGM3
SVIL
AHSA1
SEC24A
ASMTL
OXSR1
AP2A1
SDPR
BPNT1
TXNDC12
SCGB1D2
NUDT3
MT-CO2
PLG
PIGR
RBP4
AMBP
FUCA1
VWF
SERPIND1
GSN
PR
H1F0
C8G
CTSB

SRPR
SOD3
GNAO1
RRAS
RALB
LAMP1
IGF2R
LAMP2
CEACAM1
MIF
DARS
CPB1
RPA2
H2AFX
PPP3CB
NAGA
HSPA6
GJA1
TAT
ENG
PTPRA
ATP5J
EIF2AK2
ITIH2
NCF2
RAB4A
ITGAL
PZP
AGA
SCP2
NME1-NME2
FECH
ATP2B4
DTYMK
CMA1
ADRBK1
CTNNA2
FKBP2
PSM8
PSMB4
PSMB5
LOX
PTPN6
SHC1

PPIF
LRPAP1
PRKAR2B
PYCR1
GBP2
GALNS
HSPA1L
RPA3
IRS1
TIMP3
GLRX
HMGCL
ARL3
GNL1
MMP12
RPL13A
MDH1
EIF2S3
MNDA
UBA7
NAA10
CSK
ACTR1B
SKIV2L2
SLC1A4
PAFAH1B1
TSFM
GNPDA1
YAP1
GYG1
STT3A
ME1
PIP4K2A
PRRC2A
GSS
CD97
MARCKSL1
MAPKAPK2
ARRB1
AMPH
PSMB2
CPT1A
METAP2
DAP3

SMARCA2
TPMT
PSMD7
SUOX
AGFG1
STAT2
THOP1
SUCLG1
SMTN
RAD23A
NAGLU
AIF1
GP2
SEC13
OXCT1
ATP5I
RPS20
S100A10
CDC42
RAB5B
PCBD1
VBP1
DCAF7
WDR5
PSMC5
RPS23
SNRPE
LSM3
SNRPD1
RAP1A
ACTG2
TMSB10
IGBP1
PIP4K2B
SRPK2
LCN2
IFI35
DCD
MRPS9
COG7
RHOG
EFNB1
TFAM
PURA

| |
|---|
| REEP5 |
| CTBS |
| DR1 |
| PLCB3 |
| AKAP12 |
| TGFBR3 |
| CSTF1 |
| PPAT |
| GOLGA3 |
| LOXL1 |
| SLFN5 |
| VAC14 |
| NCBP1 |
| HSPA14 |
| CGNL1 |
| GALNT2 |
| ASPH |
| DPYD |
| PPP1R8 |
| STK4 |
| COASY |
| PRKAA1 |
| DNAJC3 |
| MRPL49 |
| SQSTM1 |
| NAE1 |
| BYSL |
| CKAP5 |
| BOP1 |
| GAMT |
| PDIA5 |
| PLS1 |
| CLINT1 |
| MESDC2 |
| RFTN1 |
| CRYM |
| ZNF638 |
| PSMD6 |
| FAM175B |
| RRS1 |
| PGM5 |
| PPP2R5A |
| PVRL1 |
| RSU1 |

SKIV2L
TRADD
TRIP6
TBCC
STXBP2
ADIRF
RAB11B
PKN2
DDB1
CDC37
PTGIS
IFI16
MAN2A1
GUK1
TMEM132A
HLA-DRB5
LRRFIP1
TMEM30B
RABL6
HSP90AB4P
ZNF326
MOV10
RRP12
DLG3
MORF4L2
CRNKL1
MRPS18A
SNX5
PDZK1
HECTD3
UBR4
XPNPEP1
HIST2H3PS2
NDRG3
ARFGEF3
PRR14L
CROCC
BROX
GLIPR2
ECHDC3
TBC1D9B
ARHGAP17
FCRLB
ATL3

HIST2H2AA3
KIAA1841
HIBCH
EDC4
CASC4
PGM2L1
DARS2
FIGNL1
LARP1
SMARCD3
IGSF10
HSDL2
PPP1R21
THSD4
SH3BP1
TPRG1
XIRP1
UNC13D
CCDC80
SUPT6H
TRMT10C
GDPD3
EPM2AIP1
NUFIP2
HDDC2
HDGFRP2
MRPL55
TMED4
GALNT7
GLRX5
SETD3
FAM134C
RTTN
TMEM173
CARM1
NIT1
VRK2
GATAD2A
IRF2BP1
PLD3
HID1
MCAT
FAM114A1
DNAJC10

MRPL41
EXOC8
THNSL1
CYP4X1
NDUFAF2
NUP93
PROM2
CPXM2
MINK1
OXR1
IGSF1
FAM98A
KCNH5
NPEPL1
RBM15B
PHC3
APPL2
REPS2
PLBD2
GADD45GIP1
NPLOC4
MUC7
STT3B
PNPT1
PREX1
NEK9
PLEKHO2
DTX3L
CHD5
GNPDA2
MICAL1
ADAMTS16
WIPF2
COG1
MZB1
PPIL4
PRPF31
GBF1
AP3S1
SORL1
PVRL2
HTRA1
ARPC1A
GLG1

ARHGEF2
USP7
CUL5
ATP6V0A1
NCLN
CCDC102A
MRPL24
SYAP1
EXOC4
ISOC2
SLC25A46
PBXIP1
SH3KBP1
PTER
FAF2
SNRNP40
MRPL38
DNAJA3
AHCYL2
LRIG1
COG3
CHAMP1
USP47
DNAJC1
EXOC2
SNX27
DNHD1
SFXN2
PGLYRP2
GBP5
GBP4
NEDD4L
TRNT1
FYTTD1
SNX18
IL4I1
WRNIP1
RANBP9
SIN3A
PFDN5
RNF2
SORT1
S100A13
SEPHS2

NAPG
MIPEP
TM9SF2
CIB1
NIPSNAP1
ARPC5L
MACROD1
FYCO1
MXRA8
SDF4
WIBG
FAM213A
TACO1
HDHD3
LRRC1
PI4K2A
BDH2
PBDC1
UTP14A
DUSP23
TMED9
ACAT2
TAPBPL
HINT2
SRRT
MRPL13
NIFK
POTEKP
MRPL37
OSBPL8
DPY30
UBE2O
PITHD1
KLC2
MRPL18
IRF2BPL
NUCKS1
SLK
TAOK3
MRPL46
FN3K
POFUT1
ARMT1
LRRC40

ELP3
TBC1D17
CARS2
PPCS
SIAE
GRPEL1
SCPEP1
CACYBP
RDH14
MUC5B
SPON1
NCOA5
CELSR2
ATP13A1
MRPL47
ARFGAP3
LZTFL1
MRPL40
NIT2
SAR1A
CTPS2
AASDHPPT
ADARB2
TOMM22
ATG3
SACM1L
ABHD10
LIN7C
AGPAT5
ABCF3
ATAD3A
ARL8B
DDX18
ARMC1
NECAP2
HIF1AN
MRPL16
THUMPD1
P4HTM
CDKN2AIP
GAR1
CELSR1
ZNF226
TECR

ERAP1
THYN1
NDUFA13
RAI14
COL20A1
DIP2B
PTGFRN
MBD2
COPG2
UBA2
SEL1L
COPS7A
ADD3
DKFZp566H1924
SEC63
SUSD2
ARMCX3
SRP68
UBQLN2
PCYOX1
EVL
BAZ1B
HOOK1
GGA2
HN1
MRTO4
CDV3
MCTS1
MKL2
COL17A1
NFU1
G3BP2
COPS3
C14orf166
PIN4
PLAA
VDAC3
DRG1
COG6
WBP11
SUGT1
PPIL1
UFC1
TSC22D4

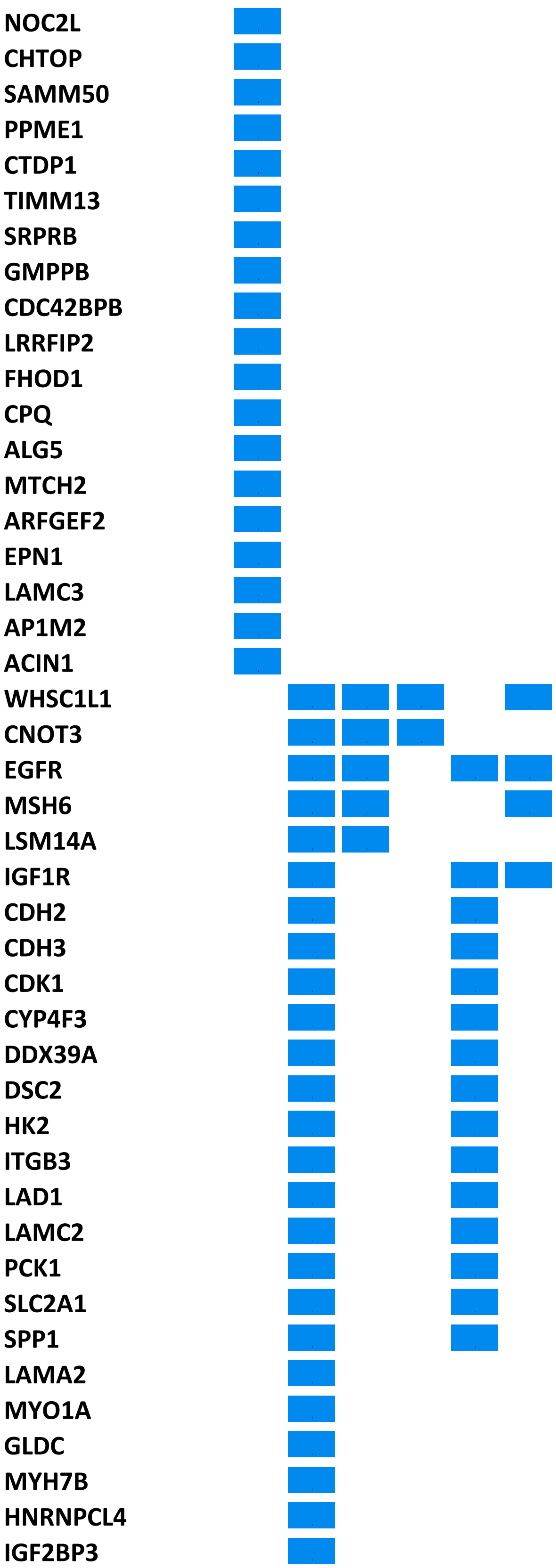
NOC2L
CHTOP
SAMM50
PPME1
CTDP1
TIMM13
SRPRB
GMPPB
CDC42BPB
LRRFIP2
FHOD1
CPQ
ALG5
MTCH2
ARFGEF2
EPN1
LAMC3
AP1M2
ACIN1
WHSC1L1
CNOT3
EGFR
MSH6
LSM14A
IGF1R
CDH2
CDH3
CDK1
CYP4F3
DDX39A
DSC2
HK2
ITGB3
LAD1
LAMC2
PCK1
SLC2A1
SPP1
LAMA2
MYO1A
GLDC
MYH7B
HNRNPCL4
IGF2BP3

CES2
HSPB6
HSD17B6
UBE2L6
ATP2A1
ALOX15B
NCAM2
DENR
SSSCA1
KLHL41
HIST1H2BL
LDB3
CRTAP
UGT2B15
MOCS3
FTCD
ADH1C
PAH
OTC
CSTA
IGHV3-20
MBP
PMP2
ALDOB
ARG1
CYP1A2
CYP2E1
UGT2B4
FABP1
NEFL
NEFM
ADH1A
TPM2
ABCB1
GSTA3
ADH4
HMBS
ITGA2B
MYL3
MMP10
C4A
UBA52
CBS
CYP2C8

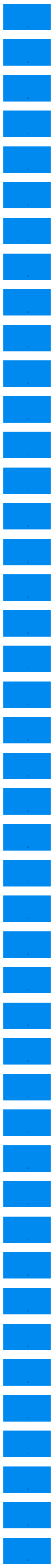

| |
|---|
| MYL2 |
| MYH3 |
| SLC2A2 |
| PYGM |
| CYP2A6 |
| CYP2C9 |
| NEFH |
| RNASE3 |
| MYL4 |
| MYH1 |
| MYH7 |
| PEPD |
| MYH6 |
| TNNT1 |
| GYS1 |
| ENO3 |
| MYL6B |
| SNRPB |
| ANK1 |
| UGT2B7 |
| ZNF30 |
| AKR1C4 |
| CKMT2 |
| CTPS1 |
| NEB |
| AGXT |
| NME2 |
| TUBG2 |
| GPT |
| PTX3 |
| AK4 |
| S100A2 |
| PBLD |
| PKLR |
| CPS1 |
| CASQ1 |
| FMO3 |
| CASP14 |
| CEACAM8 |
| HPD |
| DSG3 |
| CTH |
| MCM4 |
| SHMT1 |

ACTN2
CPOX
RPL13AP3
EIF2S3L
PRPH
TNNT3
LGALS7
PCYT1B
HARS2
MTHFS
KHK
SULT1A2
GATM
SERPINB8
FXR2
MYOM1
AKR1C2
MYOM2
MTTP
LGALS4
ARPC4
RPL26L1
RAB11A
GNB4
ACTA1
TUBA1C
CSNK2A3
HIST2H3A
CBX1
MAT1A
CDK3
MYBPC1
DSG1
CYP4A11
AKR1C1
PTPN12
AOX1
SULT2A1
EHHADH
GNL2
PKP1
DPYS
TRIM29
UBE4A

NEDD8
ETFDH
HIST2H2AC
CA9
COL28A1
FGD3
FNBP1L
ANXA8L2
MYOM3
KRT79
ACSM2B
MTHFD1L
OLFML1
TRIM72
EEPD1
HSD17B13
ARPIN
YTHDF3
AACS
MISP
RBM12B
CLYBL
CADM4
AKR7L
NT5C
TTN
EVPL
GLMN
BHMT
MYLPF
SCLY
PPP1R9B
TBCD
MRI1
TUBB2B
LSM14B
PRX
FUT8
SIGLEC1
C11orf68
CHMP4B
MYOZ1
MYOZ2
INCENP

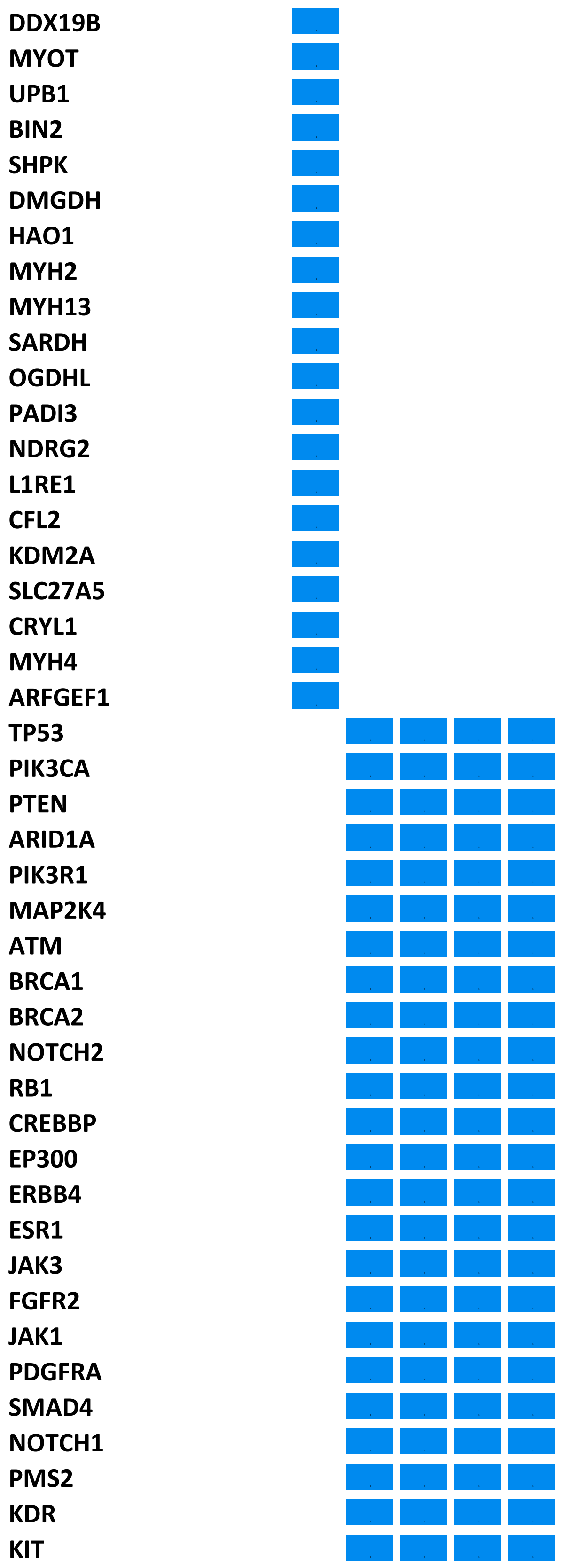
DDX19B
MYOT
UPB1
BIN2
SHPK
DMGDH
HAO1
MYH2
MYH13
SARDH
OGDHL
PADI3
NDRG2
L1RE1
CFL2
KDM2A
SLC27A5
CRYL1
MYH4
ARFGEF1
TP53
PIK3CA
PTEN
ARID1A
PIK3R1
MAP2K4
ATM
BRCA1
BRCA2
NOTCH2
RB1
CREBBP
EP300
ERBB4
ESR1
JAK3
FGFR2
JAK1
PDGFRA
SMAD4
NOTCH1
PMS2
KDR
KIT

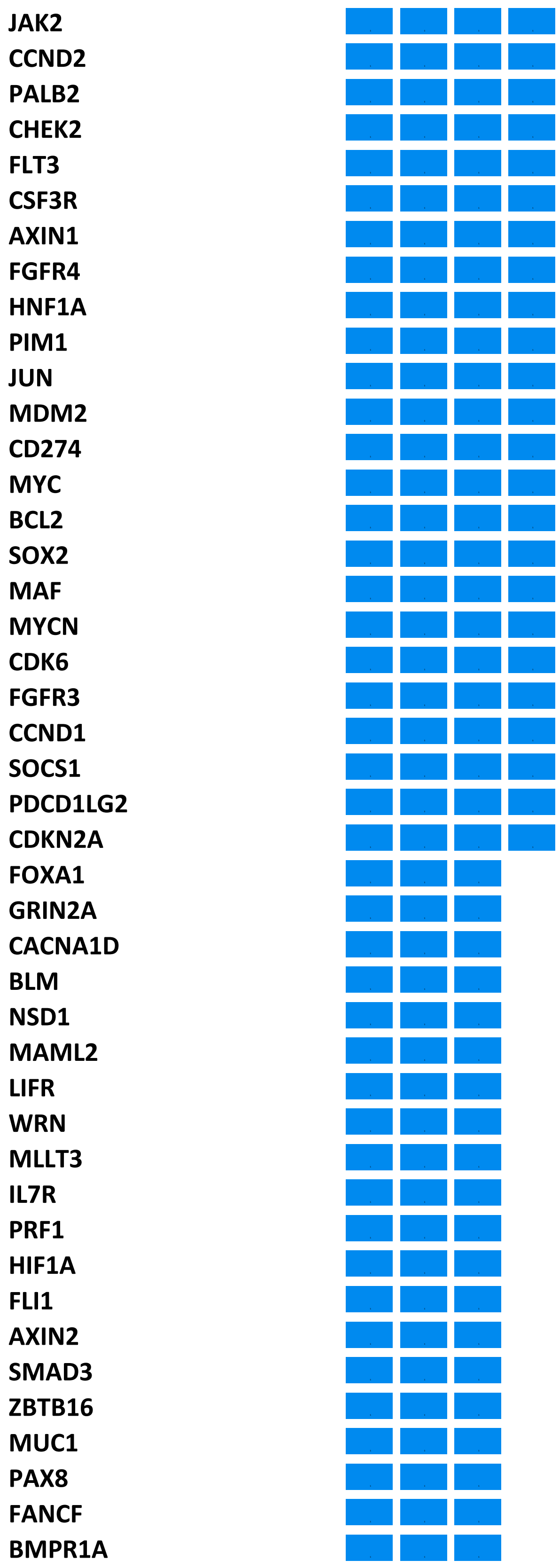
JAK2
CCND2
PALB2
CHEK2
FLT3
CSF3R
AXIN1
FGFR4
HNF1A
PIM1
JUN
MDM2
CD274
MYC
BCL2
SOX2
MAF
MYCN
CDK6
FGFR3
CCND1
SOCS1
PDCD1LG2
CDKN2A
FOXA1
GRIN2A
CACNA1D
BLM
NSD1
MAML2
LIFR
WRN
MLLT3
IL7R
PRF1
HIF1A
FLI1
AXIN2
SMAD3
ZBTB16
MUC1
PAX8
FANCF
BMPR1A

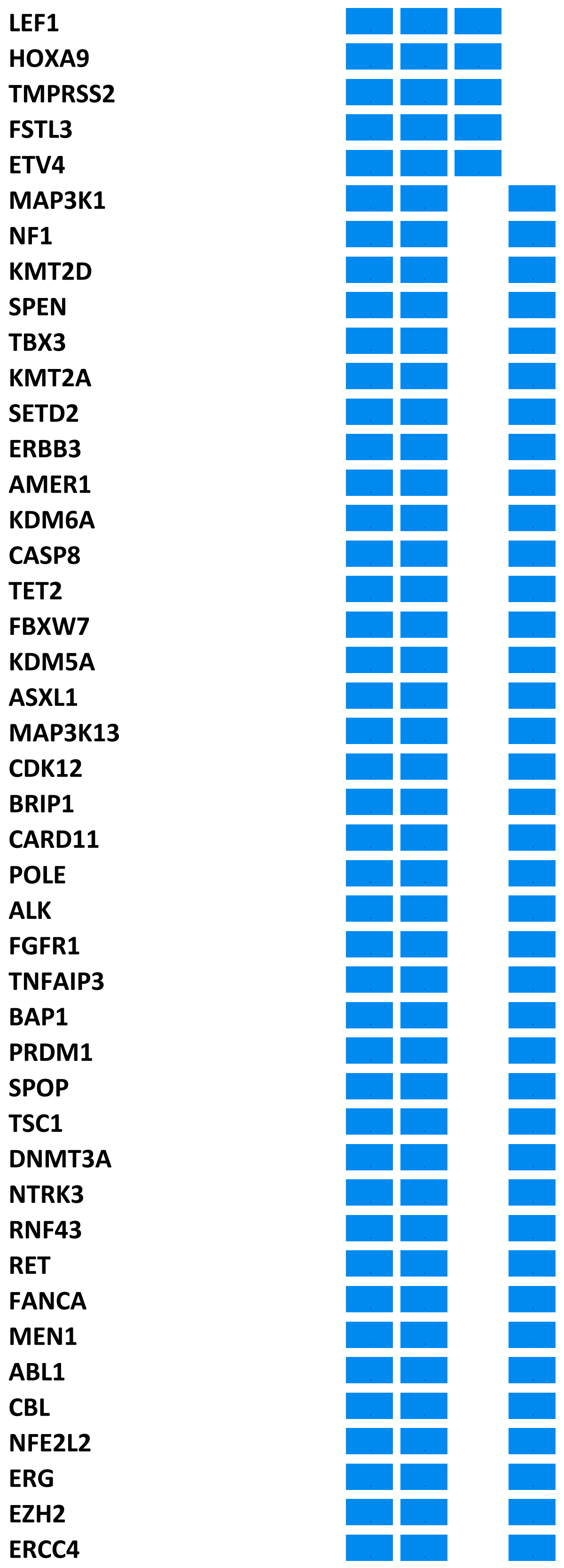
LEF1
HOXA9
TMPRSS2
FSTL3
ETV4
MAP3K1
NF1
KMT2D
SPEN
TBX3
KMT2A
SETD2
ERBB3
AMER1
KDM6A
CASP8
TET2
FBXW7
KDM5A
ASXL1
MAP3K13
CDK12
BRIP1
CARD11
POLE
ALK
FGFR1
TNFAIP3
BAP1
PRDM1
SPOP
TSC1
DNMT3A
NTRK3
RNF43
RET
FANCA
MEN1
ABL1
CBL
NFE2L2
ERG
EZH2
ERCC4

| | | | |
|---|---|---|---|
| BRAF | ■ | ■ | ■ |
| NF2 | ■ | ■ | ■ |
| NTRK1 | ■ | ■ | ■ |
| NBN | ■ | ■ | ■ |
| IKZF1 | ■ | ■ | ■ |
| TNFRSF14 | ■ | ■ | ■ |
| DAXX | ■ | ■ | ■ |
| RAD51B | ■ | ■ | ■ |
| CIC | ■ | ■ | ■ |
| MITF | ■ | ■ | ■ |
| RARA | ■ | ■ | ■ |
| MYCL | ■ | ■ | ■ |
| CD79B | ■ | ■ | ■ |
| HRAS | ■ | ■ | ■ |
| CCND3 | ■ | ■ | ■ |
| CXCR4 | ■ | ■ | ■ |
| BCL6 | ■ | ■ | ■ |
| SYK | ■ | ■ | ■ |
| SUFU | ■ | ■ | ■ |
| MDM4 | ■ | ■ | ■ |
| SDHC | ■ | ■ | ■ |
| H3F3A | ■ | ■ | ■ |
| FLCN | ■ | ■ | ■ |
| SMARCB1 | ■ | ■ | ■ |
| STK11 | ■ | ■ | ■ |
| IRF4 | ■ | ■ | ■ |
| MUTYH | ■ | ■ | ■ |
| MPL | ■ | ■ | ■ |
| FAS | ■ | ■ | ■ |
| NKX2-1 | ■ | ■ | ■ |
| FOXL2 | ■ | ■ | ■ |
| SDHD | ■ | ■ | ■ |
| CEBPA | ■ | ■ | ■ |
| KMT2C | ■ | ■ | |
| NCOR1 | ■ | ■ | |
| RUNX1 | ■ | ■ | |
| AKAP9 | ■ | ■ | |
| PTPRB | ■ | ■ | |
| UBR5 | ■ | ■ | |
| ARID1B | ■ | ■ | |
| RUNX1T1 | ■ | ■ | |
| TET1 | ■ | ■ | |
| TRRAP | ■ | ■ | |
| ZFHX3 | ■ | ■ | |

| |
|---|
| KAT6B |
| ATP2B3 |
| AFF3 |
| NIN |
| AFF1 |
| BCR |
| KAT6A |
| BCL9 |
| MYO5A |
| MECOM |
| ZMYM2 |
| PLCG1 |
| CUX1 |
| CBLB |
| CAMTA1 |
| PTPN13 |
| IKBKB |
| STAT6 |
| RANBP17 |
| CASC5 |
| TRIM33 |
| KIAA1549 |
| CHN1 |
| MLLT10 |
| ABL2 |
| ERCC5 |
| NCOA2 |
| ERCC3 |
| ZCCHC8 |
| ERC1 |
| NFATC2 |
| TRIM24 |
| FOXP1 |
| TCF12 |
| PCSK7 |
| TSHR |
| FOXO3 |
| ARHGEF12 |
| ERCC2 |
| USP6 |
| PER1 |
| PMS1 |
| PAX3 |
| MYB |

BCL11B
PRRX1
SPECC1
MKL1
RALGDS
FGFR1OP
ELL
CIITA
NRG1
EXT1
RECQL4
ETV1
ETV5
TCF7L2
HEY1
NFIB
TERT
NUTM1
SLC45A3
PICALM
CANT1
NCOA1
STAT5B
STIL
PRCC
LZTR1
SUZ12
LCK
HOXA11
ZNF384
ETNK1
GPC3
ARNT
PHOX2B
FLT4
BRD3
ECT2L
DDX10
KLK2
PRDM16
GAS7
PLAG1
FCGR2B
FAM131B

FCRL4
SMARCD1
POU5F1
ACKR3
HOXC13
MAX
MAFB
TCF3
NAB2
CCNB1IP1
ZNF331
PAX7
CREB3L2
NCKIPSD
BCL10
TTL
BUB1B
CREB1
PPP6C
BCL7A
NUTM2A
RSPO2
HOXA13
MLF1
WWTR1
RHOH
TFPT
MN1
SH2B3
ATF1
KCNJ5
TAL1
TNFRSF17
SDC4
HOXD13
PHF6
CHIC2
BIRC3
SS18L1
CRTC3
TFEB
LHFP
PATZ1
PBX1

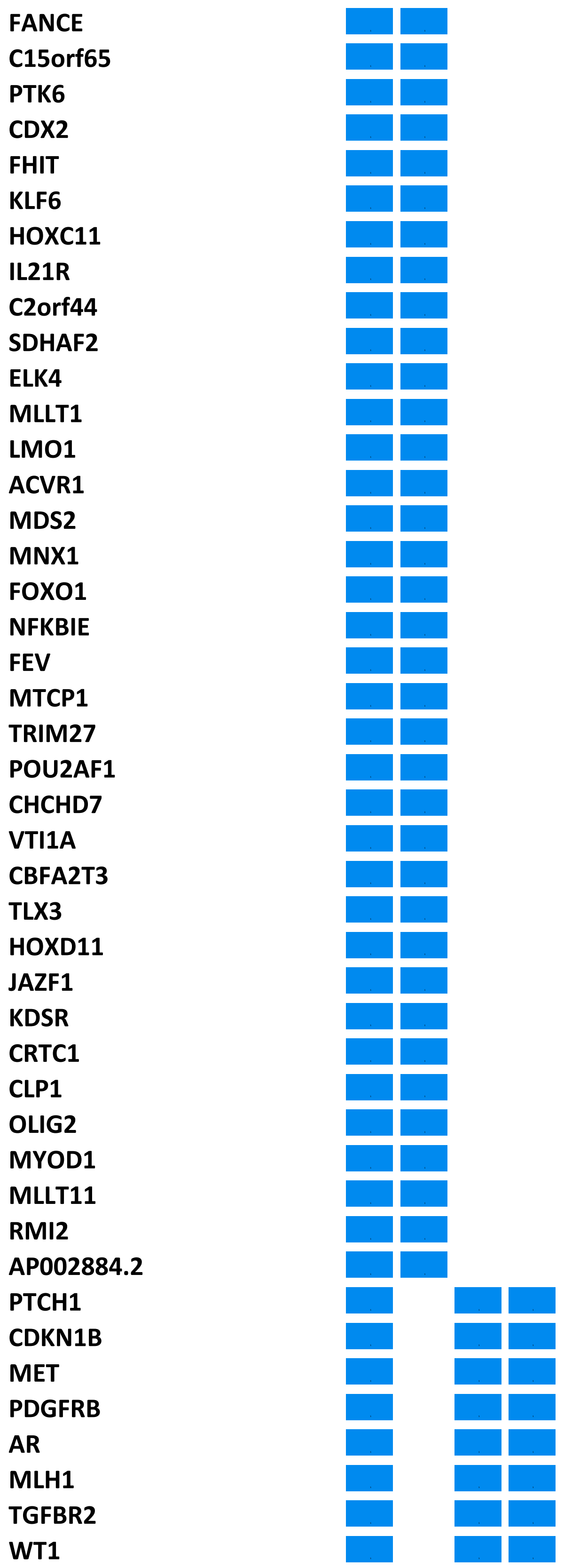
FANCE
C15orf65
PTK6
CDX2
FHIT
KLF6
HOXC11
IL21R
C2orf44
SDHAF2
ELK4
MLLT1
LMO1
ACVR1
MDS2
MNX1
FOXO1
NFKBIE
FEV
MTCP1
TRIM27
POU2AF1
CHCHD7
VTI1A
CBFA2T3
TLX3
HOXD11
JAZF1
KDSR
CRTC1
CLP1
OLIG2
MYOD1
MLLT11
RMI2
AP002884.2
PTCH1
CDKN1B
MET
PDGFRB
AR
MLH1
TGFBR2
WT1

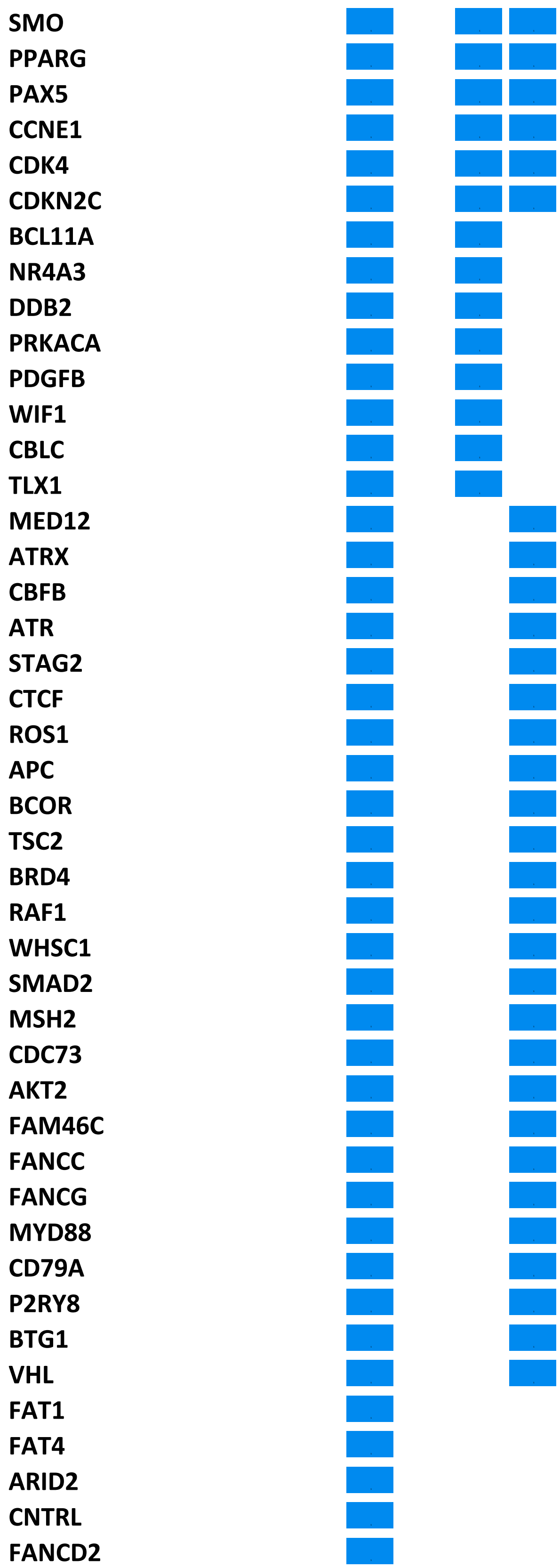
SMO
PPARG
PAX5
CCNE1
CDK4
CDKN2C
BCL11A
NR4A3
DDB2
PRKACA
PDGFB
WIF1
CBLC
TLX1
MED12
ATRX
CBFB
ATR
STAG2
CTCF
ROS1
APC
BCOR
TSC2
BRD4
RAF1
WHSC1
SMAD2
MSH2
CDC73
AKT2
FAM46C
FANCC
FANCG
MYD88
CD79A
P2RY8
BTG1
VHL
FAT1
FAT4
ARID2
CNTRL
FANCD2

ZNF521
DICER1
CYLD
AFF4
CEP89
SETBP1
ETV6
XPC
FOXO4
TFE3
ITK
EXT2
SS18
EBF1
PPFIBP1
ARHGAP26
LRIG3
SRGAP3
ELF4
PTPRK
SLC34A2
ACSL6
HLF
HMGA2
TCL6
MALT1
FBXO11
GATA2
TRAF7
SSX1
CREB3L1
KLF4
GOPC
PWWP2A
GOLGA5
POT1
BCL3
HERPUD1
SBDS
DDIT3
GATA1
TAL2
LYL1
LMO2

IL2
RSPO3
HMGN2P46
CNBP
SSX2
ZRSR2
NUTM2B
XPA
TCL1A
CRLF2
ACVR1B
AKT3
AURKA
AURKB
BCL2L1
BTG2
CDKN1A
CDKN2B
DDR2
FGF10
FGF12
GATA4
GSK3B
HGF
NOTCH3
NTRK2
PARP2
PDCD1
POLD1
RAD51
RAD51C
RAD52
RAD54L
SOX9
TEK
VEGFA
XRCC2
ABCA8
ACTR3B
ACVR1C
ACVRL1
ADAM12
ADCY9
ADM

AGTR1
ANGPT1
ANLN
APH1B
AREG
ARNT2
ASPM
ATAD2
ATP10B
B3GNT3
BAD
BAG1
BAIAP2L1
BAIAP3
BAMBI
BBC3
BBOX1
BCL2A1
BCL6B
BDNF
BIRC5
BMP2
BMP4
BMP5
BMP6
BMP7
BMP8A
BMPR1B
BMPR2
BNC2
BNIP3
BORCS7
C5orf38
CACNA1H
CACNA2D3
CACNG1
CACNG4
CACNG6
CAMK2B
CAV1
CCL2
CCL21
CCL3L1
CCL4

| |
|---|
| CCL5 |
| CCL7 |
| CCL8 |
| CCNA1 |
| CCNA2 |
| CCNB1 |
| CCNE2 |
| CCR1 |
| CCR2 |
| CCR5 |
| CD163 |
| CD19 |
| CD1E |
| CD24 |
| CD27 |
| CD276 |
| CD34 |
| CD36 |
| CD68 |
| CD84 |
| CD8A |
| CD8B |
| CDC14A |
| CDC14B |
| CDC20 |
| CDC25A |
| CDC25B |
| CDC25C |
| CDC6 |
| CDC7 |
| CDCA5 |
| CDCA7L |
| CDCA8 |
| CDH5 |
| CDKN1C |
| CDKN2D |
| CDKN3 |
| CENPF |
| CEP55 |
| CETN2 |
| CFD |
| CHI3L1 |
| CHIT1 |
| CHRNA5 |

| |
|---|
| CKS1B |
| CLDN1 |
| CLDN3 |
| CLDN4 |
| CLDN7 |
| CLEC14A |
| CLEC5A |
| CMKLR1 |
| CNTFR |
| COL27A1 |
| COL4A6 |
| COL9A3 |
| COLEC12 |
| COMP |
| CTSW |
| CXADR |
| CXCL10 |
| CXCL12 |
| CXCL13 |
| CXCL5 |
| CXCL8 |
| CXCL9 |
| CXCR6 |
| CXorf36 |
| CXXC5 |
| DEPDC1 |
| DKK1 |
| DKK2 |
| DLGAP5 |
| DLL1 |
| DLL3 |
| DLL4 |
| DNAJC12 |
| DPT |
| DTX1 |
| DTX3 |
| DTX4 |
| DUSP4 |
| DUSP6 |
| E2F1 |
| E2F5 |
| ECM2 |
| EDN1 |
| EDNRB |

| |
|---|
| EFNA3 |
| EFNA5 |
| EGF |
| EGLN2 |
| EGLN3 |
| EIF2AK3 |
| EIF4E2 |
| EIF4EBP1 |
| ELF3 |
| ELK3 |
| ELOVL2 |
| EMCN |
| ENPP2 |
| EPAS1 |
| ERCC1 |
| EREG |
| ESPL1 |
| ETV7 |
| EXO1 |
| EYA1 |
| EYA2 |
| EYA4 |
| F3 |
| FAM124B |
| FAM198B |
| FAM214A |
| FAM83D |
| FAP |
| FGF1 |
| FGF13 |
| FGF18 |
| FGF2 |
| FGF7 |
| FGF9 |
| FLRT3 |
| FOS |
| FOSL1 |
| FOXC1 |
| FOXC2 |
| FOXM1 |
| FOXP3 |
| FREM2 |
| FST |
| FSTL1 |

FUT3
FXYD3
FZD10
FZD7
FZD8
FZD9
GABRP
GADD45A
GADD45B
GADD45G
GAS1
GDF15
GDF5
GHR
GJB2
GLI3
GNG4
GNLY
GPC4
GPR160
GRB7
GREM1
GRIA3
GRIN1
GTF2H2
GZMA
GZMB
GZMH
GZMM
HAPLN1
HAS1
HDAC10
HDAC11
HDAC5
HDAC6
HDC
HEG1
HELLS
HEMK1
HES1
HIST1H2BH
HIST1H3H
HIST3H2BB
HLA-DMA

HLA-DMB
HLA-DOB
HLA-DPA1
HOXA5
HOXA7
HOXB13
HOXB3
IBSP
ID1
ID2
ID4
IDO1
IFT140
IGF1
IKZF3
IL10RA
IL11RA
IL12RB2
IL13RA1
IL1B
IL1R2
IL20RA
IL20RB
IL22RA2
IL24
IL2RA
IL2RB
IL3RA
IL4R
IL6
IL6R
INHBA
INHBB
IRF6
IRX1
ISG15
ISM1
ITGB6
JAG1
JAG2
JAM2
JCAD
KAT2B
KCNB1

| |
|---|
| KIAA0040 |
| KIF11 |
| KIF14 |
| KIF23 |
| KIF2C |
| KIFC1 |
| KLRK1 |
| KRT14 |
| KRT17 |
| KRT5 |
| KRT6B |
| KRT7 |
| LAG3 |
| LAMA3 |
| LEFTY2 |
| LEMD1 |
| LEP |
| LEPR |
| LFNG |
| LIF |
| LINC02381 |
| LPL |
| LRRC32 |
| LTB |
| LTBP1 |
| MAD2L1 |
| MAP3K12 |
| MAPK10 |
| MAPK8IP2 |
| MARCO |
| MED1 |
| MELK |
| MEOX2 |
| MFNG |
| MIA |
| MIS18A |
| MKI67 |
| MME |
| MMP11 |
| MMP3 |
| MMP7 |
| MMRN2 |
| MRE11 |
| MS4A2 |

MSR1
MT1G
MUS81
MYBL2
MYCT1
NCAM1
NCAPH2
NDC80
NDP
NEIL1
NEIL2
NEIL3
NEO1
NETO2
NFATC1
NFKBIZ
NGFR
NKG7
NOD2
NPR1
NR4A1
NRCAM
NRXN1
NRXN3
NSD3
NUDT1
NUF2
NUMBL
NUPR1
OCLN
OLFML2B
ORC6
PALMD
PBX3
PDE9A
PDK4
PECAM1
PGR
PIK3CD
PIK3R2
PIK3R3
PIK3R5
PKMYT1
PLA1A

PLA2G2A
PLA2G3
PLA2G4A
PLA2G4F
PLAT
PLCB1
PLCB4
PLCE1
PLD1
POLQ
POPDC3
PPARGC1A
PPP2CB
PPP2R2C
PRC1
PRKAA2
PRKACB
PRKCA
PRKCB
PRKX
PRLR
PROM1
PSAT1
PSMB10
PSMB7
PTGDS
PTGER3
PTGS2
PTTG1
RAC3
RARRES3
RASAL1
RASGRF1
RASGRF2
RASGRP1
RBL1
RBL2
RBX1
RELN
RNF103
ROBO4
ROCK1
RORA
RORB

| |
|---|
| RPS6KA5 |
| RPS6KB1 |
| RPS6KB2 |
| RRM2 |
| RUNX3 |
| S100A7 |
| S1PR1 |
| SCARA5 |
| SCUBE2 |
| SELE |
| SFRP1 |
| SFRP2 |
| SFRP4 |
| SHC2 |
| SHC4 |
| SHE |
| SIDT1 |
| SIX1 |
| SKA3 |
| SKP2 |
| SLC39A6 |
| SLC44A4 |
| SLPI |
| SMAD1 |
| SMAD5 |
| SMC1B |
| SMURF2 |
| SNAI1 |
| SNAI2 |
| SOCS2 |
| SOCS3 |
| SOX10 |
| SOX17 |
| SP1 |
| SPC25 |
| SPDEF |
| SPN |
| SPRY1 |
| SPRY2 |
| SPRY4 |
| SRPX |
| ST6GALNAC2 |
| STC1 |
| SUV39H2 |

| |
|---|
| SYTL4 |
| TBC1D9 |
| TBX1 |
| TCEAL1 |
| TCF4 |
| TCF7L1 |
| TFDP1 |
| TFF1 |
| TFF3 |
| TGFB1 |
| TGFB2 |
| TGFB3 |
| THBS1 |
| TIE1 |
| TIGIT |
| TIMP4 |
| TLE3 |
| TLR4 |
| TMEM45B |
| TMPRSS4 |
| TNF |
| TNFAIP6 |
| TNFSF10 |
| TNKS |
| TNKS2 |
| TRIP13 |
| TSPAN1 |
| TSPAN7 |
| TTK |
| TTYH1 |
| TWIST1 |
| TWIST2 |
| TYK2 |
| TYMS |
| UBE2C |
| UBE2T |
| VEGFD |
| VIT |
| WEE1 |
| WNT10A |
| WNT11 |
| WNT2 |
| WNT4 |
| WNT5A |

WNT5B
WNT6
WNT7B
XRCC3
ZEB1
ZEB2
ZFPM2
ZFYVE9
ZIC2
ZNF205
Internal Reference (
NRDE2
OAZ1
PUM1
STK11IP
TBC1D10B
TBP
UBB
ALOX12B
ARAF
ARFRP1
AXL
BCL2L2
BCORL1
BTK
C11orf30
CD22
CD70
CDK8
CHEK1
CSF1R
CUL3
CYP17A1
DDR1
DIS3
DOT1L
EED
EPHA3
EPHB1
EPHB4
ERRFI1
FANCL
FGF14
FGF19

| |
|---|
| FGF23 |
| FGF3 |
| FGF4 |
| FGF6 |
| FLT1 |
| GABRA6 |
| GATA6 |
| GID4 |
| GRM3 |
| HSD3B1 |
| ID3 |
| IKBKE |
| IRF2 |
| IRS2 |
| KEAP1 |
| KEL |
| KLHL6 |
| LTK |
| MCL1 |
| MEF2B |
| MERTK |
| MKNK1 |
| MSH3 |
| MST1R |
| NFKBIA |
| Page |
| PARK2 |
| PARP3 |
| PDK1 |
| PIK3C2B |
| PIK3C2G |
| PIK3CB |
| PRKCI |
| PTPRO |
| RAD51D |
| RAL-0003-03 |
| RICTOR |
| RPTOR |
| SGK1 |
| SNCAIP |
| SRC |
| TIPARP |
| TYRO3 |
| ZNF217 |

**ZNF703**

| Gene ID | Target name |
|---|---|
| EGFR | Epidermal growth factor receptor |
| MTOR | Serine/threonine-protein kinase mTOR |
| ERBB2 | Receptor tyrosine-protein kinase erbB-2 |
| EGFR | Epidermal growth factor receptor |
| ERBB2 | Receptor tyrosine-protein kinase erbB-2 |
| EGFR | Epidermal growth factor receptor |
| ERBB2 | Receptor tyrosine-protein kinase erbB-2 |
| MTOR | Serine/threonine-protein kinase mTOR |
| EGFR | Epidermal growth factor receptor |
| ERBB2 | Receptor tyrosine-protein kinase erbB-2 |
| EGFR | Epidermal growth factor receptor |
| EGFR | Epidermal growth factor receptor |
| ERBB2 | Receptor tyrosine-protein kinase erbB-2 |
| EGFR | Epidermal growth factor receptor |
| ERBB2 | Receptor tyrosine-protein kinase erbB-2 |
| ERBB2 | Receptor tyrosine-protein kinase erbB-2 |
| MTOR | Serine/threonine-protein kinase mTOR |
| EGFR | Epidermal growth factor receptor |
| EGFR | Epidermal growth factor receptor |
| ERBB2 | Receptor tyrosine-protein kinase erbB-2 |
| MTOR | Serine/threonine-protein kinase mTOR |
| EGFR | Epidermal growth factor receptor |
| ERBB2 | Receptor tyrosine-protein kinase erbB-2 |
| EGFR | Epidermal growth factor receptor |
| EGFR | Epidermal growth factor receptor |
| EGFR | Epidermal growth factor receptor |
| ERBB2 | Receptor tyrosine-protein kinase erbB-2 |
| EGFR | Epidermal growth factor receptor |
| ERBB2 | Receptor tyrosine-protein kinase erbB-2 |
| EGFR | Epidermal growth factor receptor |
| MTOR | Serine/threonine-protein kinase mTOR |
| EGFR | Epidermal growth factor receptor |
| ERBB2 | Receptor tyrosine-protein kinase erbB-2 |
| EGFR | Epidermal growth factor receptor |
| EGFR | Epidermal growth factor receptor |

| | |
|---|---|
| EGFR | Epidermal growth factor receptor |
| EGFR | Epidermal growth factor receptor |
| ERBB2 | Receptor tyrosine-protein kinase erbB-2 |
| ERBB2 | Receptor tyrosine-protein kinase erbB-2 |
| ERBB2 | Receptor tyrosine-protein kinase erbB-2 |
| EGFR | Epidermal growth factor receptor |
| EGFR | Epidermal growth factor receptor |
| EGFR | Epidermal growth factor receptor |
| EGFR | Epidermal growth factor receptor |
| RAC1 | Ras-related C3 botulinum toxin substrate 1 |
| MAP2K1 | Dual specificity mitogen-activated protein kinase kinase 1 |
| MAP2K1 | Dual specificity mitogen-activated protein kinase kinase; MEK1/2 |
| MAP2K2 | Dual specificity mitogen-activated protein kinase kinase 2 |
| AKT1 | RAC-alpha serine/threonine-protein kinase |
| MAP2K1 | Dual specificity mitogen-activated protein kinase kinase 1 |
| MAP2K2 | Dual specificity mitogen-activated protein kinase kinase 2 |
| MAP2K1 | Dual specificity mitogen-activated protein kinase kinase 1 |
| MAP2K2 | Dual specificity mitogen-activated protein kinase kinase 2 |
| MAP2K1 | Dual specificity mitogen-activated protein kinase kinase 1 |
| MAP2K2 | Dual specificity mitogen-activated protein kinase kinase 2 |
| MAP2K1 | Dual specificity mitogen-activated protein kinase kinase 1 |
| MAP2K2 | Dual specificity mitogen-activated protein kinase kinase 2 |
| MAP2K1 | Dual specificity mitogen-activated protein kinase kinase 1 |
| MAP2K2 | Dual specificity mitogen-activated protein kinase kinase 2 |
| MAP2K2 | Dual specificity mitogen-activated protein kinase kinase 2 |
| MAP2K1 | Dual specificity mitogen-activated protein kinase kinase 1 |
| MAP2K1 | Dual specificity mitogen-activated protein kinase kinase 1 |
| MAP2K2 | Dual specificity mitogen-activated protein kinase kinase 2 |
| MAP2K1 | Dual specificity mitogen-activated protein kinase kinase 1 |
| MAP2K2 | Dual specificity mitogen-activated protein kinase kinase 2 |
| TOP1 | DNA topoisomerase 1 |
| TOP1 | DNA topoisomerase 1 |
| TOP1 | DNA topoisomerase 1 |
| TOP1 | DNA topoisomerase 1 |
| HDAC1 | Histone deacetylase 1 |
| IGF1R | Insulin-like growth factor 1 receptor |
| MAPK1 | Mitogen-activated protein kinase 1 |
| IGF1R | Insulin-like growth factor 1 receptor |
| HDAC1 | Histone deacetylase |
| HDAC1 | Histone deacetylase 1 |
| HDAC1 | Histone deacetylase |
| HDAC1 | Histone deacetylase 1/3/5/8 |
| HDAC1 | Histone deacetylase 1 |
| HDAC1 | Histone deacetylase (HDAC1 and HDAC2) |

| | |
|---|---|
| IGF1R | Insulin-like growth factor 1 receptor |
| HDAC1 | Histone deacetylase 1 |
| IGF1R | Insulin-like growth factor 1 receptor |
| HDAC1 | Histone deacetylase 1 |
| IGF1R | Insulin-like growth factor 1 receptor |
| PARP1 | Poly [ADP-ribose] polymerase 1 |
| PARP1 | Poly [ADP-ribose] polymerase 1 |
| PARP1 | Poly [ADP-ribose] polymerase 1 |
| TOP2A | DNA topoisomerase 2-alpha |
| TOP2A | DNA topoisomerase II |
| HDAC2 | Histone deacetylase 2 |
| MMP9 | Matrix metalloproteinase-9 |
| MAPK3 | Mitogen-activated protein kinase 3 |
| MMP9 | Matrix metalloproteinase-9 |
| GRB2 | Growth factor receptor-bound protein 2 |
| TOP2A | DNA topoisomerase 2-alpha |
| TOP2A | DNA topoisomerase 2-alpha |
| TOP2A | DNA topoisomerase 2-alpha |
| TOP2A | DNA topoisomerase 2-alpha |
| CA12 | Carbonic anhydrase 12 |
| TOP2A | DNA topoisomerase 2-alpha |
| CA12 | Carbonic anhydrase 12 |
| MAPK3 | Mitogen-activated protein kinase 3 |
| PIK3CG | Phosphatidylinositol 4,5-bisphosphate 3-kinase catalytic subunit gamma isoform |
| ROCK2 | Rho-associated protein kinase 2 |
| TOP2A | DNA topoisomerase 2-alpha |
| TOP2A | DNA topoisomerase 2-alpha |
| HDAC2 | Histone deacetylase 2 |
| HDAC2 | Histone deacetylase 2 |
| ROCK2 | Rho-associated protein kinase 2 |
| ROCK2 | Rho-associated protein kinase 2 |
| ROCK2 | Rho-associated protein kinase 2 |
| ROCK2 | Rho-associated protein kinase 2 |
| MAPK3 | Mitogen-activated protein kinase 3 |
| HDAC2 | Histone deacetylase 2 |
| ROCK2 | Rho-associated protein kinase 2 |
| ROCK2 | Rho-associated protein kinase 2 |
| HDAC2 | Histone deacetylase 2 |
| PIK3CG | Phosphatidylinositol 4,5-bisphosphate 3-kinase catalytic subunit gamma isoform |
| PRKDC | DNA-dependent protein kinase catalytic subunit |
| IL1RN | Interleukin-1 receptor antagonist protein |
| PARP4 | Poly [ADP-ribose] polymerase 4 |
| LYN | Tyrosine-protein kinase Lyn |
| LYN | Tyrosine-protein kinase Lyn |

| | |
|---|---|
| LYN | Tyrosine-protein kinase Lyn |
| LYN | Tyrosine-protein kinase Lyn |
| LYN | Tyrosine-protein kinase Lyn |
| LYN | Tyrosine-protein kinase Lyn |
| LYN | Tyrosine-protein kinase Lyn |
| LYN | Tyrosine-protein kinase Lyn |
| LYN | Tyrosine-protein kinase Lyn |
| LYN | Tyrosine-protein kinase Lyn |
| LYN | Tyrosine-protein kinase Lyn |
| LYN | Tyrosine-protein kinase Lyn |
| LYN | Tyrosine-protein kinase Lyn |
| LYN | Tyrosine-protein kinase Lyn |
| LYN | Tyrosine-protein kinase Lyn |
| MMP2 | 72 kDa type IV collagenase |
| CYP1A2 | Cytochrome P450 1A2 |
| TOP2B | DNA topoisomerase 2-beta |
| TXNRD1 | Thioredoxin reductase 1, cytoplasmic |
| TPMT | Thiopurine S-methyltransferase |
| PSMA1 | Proteasome subunit alpha type-1 |
| F2 | Prothrombin |
| CTSG | Cathepsin G |
| CMA1 | Chymase |
| PSMB1 | Proteasome subunit beta type-1 |
| PSMB5 | Proteasome subunit beta type-5 |
| PSMB2 | Proteasome subunit beta type-2 |
| NFKB1 | Nuclear factor NF-kappa-B complex |
| PSMD1 | 26S proteasome non-ATPase regulatory subunit 1 |
| GSR | Glutathione reductase, mitochondrial |
| CTSD | Cathepsin D |
| RRM1 | Ribonucleoside-diphosphate reductase large subunit |
| POLB | DNA polymerase beta |
| GAK | Cyclin-G-associated kinase |
| STK10 | Serine/threonine-protein kinase 10 |
| CSK | Tyrosine-protein kinase CSK |
| SLK | STE20-like serine/threonine-protein kinase |
| STK4 | Serine/threonine-protein kinase 4 |
| CDC42BPB | Serine/threonine-protein kinase MRCK beta |
| VRK2 | Serine/threonine-protein kinase VRK2 |
| MAPK14 | Mitogen-activated protein kinase 14 |
| ABCB1 | Multidrug resistance protein 1 |
| MMP2 | 72 kDa type IV collagenase |
| TUBB | Tubulin beta |
| ABCB1 | Multidrug resistance protein 1 |
| CASP1 | Caspase-1 |

| | |
|---|---|
| ABCB1 | Multidrug resistance protein 1 |
| MMP2 | 72 kDa type IV collagenase |
| GAK | Cyclin-G-associated kinase |
| EIF2AK2 | Interferon-induced, double-stranded RNA-activated protein kinase |
| STK10 | Serine/threonine-protein kinase 10 |
| CSK | Tyrosine-protein kinase CSK |
| PIP4K2C | Phosphatidylinositol 5-phosphate 4-kinase type-2 gamma |
| CAMK2D | Calcium/calmodulin-dependent protein kinase type II subunit delta |
| SLK | STE20-like serine/threonine-protein kinase |
| ABCB1 | Multidrug resistance protein 1 |
| FKBP1A | Peptidyl-prolyl cis-trans isomerase FKBP1A |
| ABCB1 | Multidrug resistance protein 1 |
| CYP1A2 | Cytochrome P450 1A2 |
| EPHX2 | Bifunctional epoxide hydrolase 2 |
| GAK | Cyclin-G-associated kinase |
| STK10 | Serine/threonine-protein kinase 10 |
| PIP4K2C | Phosphatidylinositol 5-phosphate 4-kinase type-2 gamma |
| SLK | STE20-like serine/threonine-protein kinase |
| CMPK1 | UMP-CMP kinase |
| RRM1 | Ribonucleoside-diphosphate reductase large subunit |
| ABCB1 | Multidrug resistance protein 1 |
| CA2 | Carbonic anhydrase 2 |
| ABCB1 | Multidrug resistance protein 1 |
| GAK | Cyclin-G-associated kinase |
| PIP4K2C | Phosphatidylinositol 5-phosphate 4-kinase type-2 gamma |
| CA9 | Carbonic anhydrase 9 |
| CA1 | Carbonic anhydrase 1 |
| MAPK14 | Mitogen-activated protein kinase 14 |
| ABCB1 | Multidrug resistance protein 1 |
| TUBB | Tubulin beta |
| ABCB1 | Multidrug resistance protein 1 |
| STK10 | Serine/threonine-protein kinase 10 |
| SLK | STE20-like serine/threonine-protein kinase |
| CYP1A2 | Cytochrome P450 1A2 |
| MAOA | Amine oxidase [flavin-containing] A |
| ALOX15B | Arachidonate 15-lipoxygenase, type II |
| TTR | Transthyretin |
| PPAT | Amidophosphoribosyltransferase |
| PTGS1 | Prostaglandin G/H synthase 1 |
| TPMT | Thiopurine S-methyltransferase |
| PNP | Purine nucleoside phosphorylase |
| ABCB1 | Multidrug resistance protein 1 |
| ABCB1 | Multidrug resistance protein 1 |
| MPG | DNA-3-methyladenine glycosylase |

| | |
|---|---|
| IMPDH2 | Inosine-5'-monophosphate dehydrogenase 2 |
| CA2 | Carbonic anhydrase 2 |
| STK10 | Serine/threonine-protein kinase 10 |
| CSK | Tyrosine-protein kinase CSK |
| PIP4K2C | Phosphatidylinositol 5-phosphate 4-kinase type-2 gamma |
| CDC42BPB | Serine/threonine-protein kinase MRCK beta |
| CA9 | Carbonic anhydrase 9 |
| CA1 | Carbonic anhydrase 1 |
| MAPK14 | Mitogen-activated protein kinase 14 |
| TUBB | Tubulin beta |
| TUBB | Tubulin beta chain |
| ABCB1 | Multidrug resistance protein 1 |
| TUBB3 | Tubulin beta-3 chain |
| SHMT1 | Serine hydroxymethyltransferase, cytosolic |
| ADA | Adenosine deaminase |
| FKBP1A | Peptidyl-prolyl cis-trans isomerase FKBP1A |
| CYP2C9 | Cytochrome P450 2C9 |
| FKBP3 | Peptidyl-prolyl cis-trans isomerase FKBP3 |
| ABCB1 | Multidrug resistance protein 1 |
| PDCD4 | Programmed cell death protein 4 |
| FKBP5 | Peptidyl-prolyl cis-trans isomerase FKBP5 |
| STK10 | Serine/threonine-protein kinase 10 |
| CSK | Tyrosine-protein kinase CSK |
| SLK | STE20-like serine/threonine-protein kinase |
| EPHX2 | Bifunctional epoxide hydrolase 2 |
| MAPK13 | Mitogen-activated protein kinase 13 |
| MAPK14 | Mitogen-activated protein kinase 14 |
| DCLK1 | Serine/threonine-protein kinase DCLK1 |
| GAK | Cyclin-G-associated kinase |
| EIF2AK2 | Interferon-induced, double-stranded RNA-activated protein kinase |
| PRPF4 | U4/U6 small nuclear ribonucleoprotein Prp4 |
| OXSR1 | Serine/threonine-protein kinase OSR1 |
| SGK3 | Serine/threonine-protein kinase Sgk3 |
| STK10 | Serine/threonine-protein kinase 10 |
| MARK2 | Serine/threonine-protein kinase MARK2 |
| CSNK2A2 | Casein kinase II subunit alpha' |
| PKN2 | Serine/threonine-protein kinase N2 |
| MYLK | Myosin light chain kinase, smooth muscle |
| SLK | STE20-like serine/threonine-protein kinase |
| STK4 | Serine/threonine-protein kinase 4 |
| PTK2 | Focal adhesion kinase 1 |
| PRPF4B | Serine/threonine-protein kinase PRP4 homolog |
| STK24 | Serine/threonine-protein kinase 24 |
| PIP4K2B | Phosphatidylinositol 5-phosphate 4-kinase type-2 beta |

| | |
|---|---|
| SRPK2 | SRSF protein kinase 2 |
| CAMK2D | Calcium/calmodulin-dependent protein kinase type II subunit delta |
| PTGS1 | Prostaglandin G/H synthase 1 |
| PPP3CA | Serine/threonine-protein phosphatase 2B catalytic subunit alpha isoform |
| AOX1 | Aldehyde oxidase |
| ABCB1 | Multidrug resistance protein 1 |
| ABCB1 | Multidrug resistance protein 1 |
| PTGS1 | Prostaglandin G/H synthase 1 |
| ABCB1 | Multidrug resistance protein 1 |
| TUBB | Tubulin beta |
| ABCB1 | Multidrug resistance protein 1 |
| TUBB | Tubulin beta |
| ABCB1 | Multidrug resistance protein 1 |
| TUBB | Tubulin beta |
| TUBB | Tubulin beta |
| GAK | Cyclin-G-associated kinase |
| EIF2AK2 | Interferon-induced, double-stranded RNA-activated protein kinase |
| STK10 | Serine/threonine-protein kinase 10 |
| PIP4K2C | Phosphatidylinositol 5-phosphate 4-kinase type-2 gamma |
| SLK | STE20-like serine/threonine-protein kinase |
| TUBB | Tubulin beta |
| FKBP3 | Peptidyl-prolyl cis-trans isomerase FKBP3 |
| ABCB1 | Multidrug resistance protein 1 |
| FKBP1A | Peptidyl-prolyl cis-trans isomerase FKBP1A |
| TUBB | Tubulin beta |
| GAK | Cyclin-G-associated kinase |
| STK10 | Serine/threonine-protein kinase 10 |
| CSK | Tyrosine-protein kinase CSK |
| SLK | STE20-like serine/threonine-protein kinase |
| CDC42BPB | Serine/threonine-protein kinase MRCK beta |
| DCLK1 | Serine/threonine-protein kinase DCLK1 |
| EIF2AK2 | Interferon-induced, double-stranded RNA-activated protein kinase |
| STK10 | Serine/threonine-protein kinase 10 |
| SLK | STE20-like serine/threonine-protein kinase |
| STK4 | Serine/threonine-protein kinase 4 |
| NEK9 | Serine/threonine-protein kinase Nek9 |
| PTK2 | Focal adhesion kinase 1 |
| CASK | Peripheral plasma membrane protein CASK |
| DCLK1 | Serine/threonine-protein kinase DCLK1 |
| GAK | Cyclin-G-associated kinase |
| OXSR1 | Serine/threonine-protein kinase OSR1 |
| MARK2 | Serine/threonine-protein kinase MARK2 |
| CSNK2A2 | Casein kinase II subunit alpha' |
| MYLK | Myosin light chain kinase, smooth muscle |

| | |
|---|---|
| CASK | Peripheral plasma membrane protein CASK |
| VRK2 | Serine/threonine-protein kinase VRK2 |
| CAMK2D | Calcium/calmodulin-dependent protein kinase type II subunit delta |
| GAK | Cyclin-G-associated kinase |
| EIF2AK2 | Interferon-induced, double-stranded RNA-activated protein kinase |
| STK10 | Serine/threonine-protein kinase 10 |
| CSNK2A2 | Casein kinase II subunit alpha' |
| PKN2 | Serine/threonine-protein kinase N2 |
| CSK | Tyrosine-protein kinase CSK |
| SLK | STE20-like serine/threonine-protein kinase |
| STK4 | Serine/threonine-protein kinase 4 |
| CDC42BPB | Serine/threonine-protein kinase MRCK beta |
| GAK | Cyclin-G-associated kinase |
| STK10 | Serine/threonine-protein kinase 10 |
| SLK | STE20-like serine/threonine-protein kinase |
| MAPK14 | Mitogen-activated protein kinase 14 |
| GAK | Cyclin-G-associated kinase |
| STK10 | Serine/threonine-protein kinase 10 |
| PKN2 | Serine/threonine-protein kinase N2 |
| CSK | Tyrosine-protein kinase CSK |
| MYLK | Myosin light chain kinase, smooth muscle |
| SLK | STE20-like serine/threonine-protein kinase |
| PAK1 | Serine/threonine-protein kinase PAK 1 |
| STK4 | Serine/threonine-protein kinase 4 |
| PTK2 | Focal adhesion kinase 1 |
| CASK | Peripheral plasma membrane protein CASK |
| STK24 | Serine/threonine-protein kinase 24 |
| PAK2 | Serine/threonine-protein kinase PAK 2 |
| VRK2 | Serine/threonine-protein kinase VRK2 |
| PSMB5 | Proteasome subunit beta type-5 |
| CTSB | Cathepsin B |
| EPHX2 | Bifunctional epoxide hydrolase 2 |
| DCLK1 | Serine/threonine-protein kinase DCLK1 |
| PKN2 | Serine/threonine-protein kinase N2 |
| CAMK2D | Calcium/calmodulin-dependent protein kinase type II subunit delta |
| CSK | Tyrosine-protein kinase CSK |
| DCLK1 | Serine/threonine-protein kinase DCLK1 |
| OXSR1 | Serine/threonine-protein kinase OSR1 |
| SGK3 | Serine/threonine-protein kinase Sgk3 |
| STK10 | Serine/threonine-protein kinase 10 |
| MARK2 | Serine/threonine-protein kinase MARK2 |
| CSNK2A2 | Casein kinase II subunit alpha' |
| PKN2 | Serine/threonine-protein kinase N2 |
| MYLK | Myosin light chain kinase, smooth muscle |

| | |
|---|---|
| SLK | STE20-like serine/threonine-protein kinase |
| STK4 | Serine/threonine-protein kinase 4 |
| PTK2 | Focal adhesion kinase 1 |
| PRPF4B | Serine/threonine-protein kinase PRP4 homolog |
| STK24 | Serine/threonine-protein kinase 24 |
| PAK2 | Serine/threonine-protein kinase PAK 2 |
| PIP4K2B | Phosphatidylinositol 5-phosphate 4-kinase type-2 beta |
| SRPK2 | SRSF protein kinase 2 |
| ITGAL | Leukocyte adhesion glycoprotein LFA-1 alpha |
| TUBB | Tubulin beta |
| TUBB | Tubulin beta |
| PSMB5 | Proteasome subunit beta type-5 |
| PSMB2 | Proteasome subunit beta type-2 |
| PSMB1 | Proteasome subunit beta type-1 |
| PARP10 | Poly [ADP-ribose] polymerase 10 |
| PARP14 | Poly [ADP-ribose] polymerase 14 |
| PPARG | Peroxisome proliferator-activated receptor gamma |
| AR | Androgen receptor |
| FLT3 | Receptor-type tyrosine-protein kinase FLT3 |
| KDR | Vascular endothelial growth factor receptor 2 |
| FGFR3 | Fibroblast growth factor receptor 3 |
| PDGFRA | Platelet-derived growth factor receptor alpha |
| JAK2 | Tyrosine-protein kinase JAK2 |
| JAK3 | Tyrosine-protein kinase JAK3 |
| PDGFRB | Platelet-derived growth factor receptor beta |
| FGFR2 | Fibroblast growth factor receptor 2 |
| KIT | Mast/stem cell growth factor receptor Kit |
| ERBB4 | Receptor tyrosine-protein kinase erbB-4 |
| PIK3CA | Phosphatidylinositol 4,5-bisphosphate 3-kinase catalytic subunit alpha isoform |
| TGFBR2 | TGF-beta receptor type-2 |
| FLT3 | Receptor-type tyrosine-protein kinase FLT3 |
| KDR | Vascular endothelial growth factor receptor 2 |
| PDGFRA | Platelet-derived growth factor receptor alpha |
| MET | Hepatocyte growth factor receptor |
| JAK2 | Tyrosine-protein kinase JAK2 |
| JAK3 | Tyrosine-protein kinase JAK3 |
| PDGFRB | Platelet-derived growth factor receptor beta |
| FGFR2 | Fibroblast growth factor receptor 2 |
| KIT | Mast/stem cell growth factor receptor Kit |
| ERBB4 | Receptor tyrosine-protein kinase erbB-4 |
| AR | Androgen receptor |
| AR | Androgen receptor |
| ESR1 | Estrogen receptor |
| FLT3 | Receptor-type tyrosine-protein kinase FLT3 |

| | |
|---|---|
| KDR | Vascular endothelial growth factor receptor 2 |
| MET | Hepatocyte growth factor receptor |
| KIT | Mast/stem cell growth factor receptor Kit |
| ERBB4 | Receptor tyrosine-protein kinase erbB-4 |
| CHEK2 | Serine/threonine-protein kinase Chk2 |
| FGFR2 | Fibroblast growth factor receptor 2 |
| JAK2 | Tyrosine-protein kinase JAK2 |
| FLT3 | Receptor-type tyrosine-protein kinase FLT3 |
| KDR | Vascular endothelial growth factor receptor 2 |
| PDGFRA | Platelet-derived growth factor receptor alpha |
| PDGFRB | Platelet-derived growth factor receptor beta |
| KIT | Mast/stem cell growth factor receptor Kit |
| PDGFRA | Platelet-derived growth factor receptor |
| PDGFRB | Platelet-derived growth factor receptor beta |
| ERBB4 | Receptor tyrosine-protein kinase erbB-4 |
| AR | Androgen receptor |
| ESR1 | Estrogen receptor |
| PIM1 | Serine/threonine-protein kinase pim-1 |
| FGFR2 | Fibroblast growth factor receptor 2 |
| JAK2 | Tyrosine-protein kinase JAK2 |
| FLT3 | Receptor-type tyrosine-protein kinase FLT3 |
| PDGFRA | Platelet-derived growth factor receptor alpha |
| PDGFRB | Platelet-derived growth factor receptor beta |
| KIT | Mast/stem cell growth factor receptor Kit |
| AR | Androgen receptor |
| FLT3 | Receptor-type tyrosine-protein kinase FLT3 |
| KDR | Vascular endothelial growth factor receptor 2 |
| FGFR3 | Fibroblast growth factor receptor 3 |
| PDGFRA | Platelet-derived growth factor receptor alpha |
| JAK3 | Tyrosine-protein kinase JAK3 |
| PDGFRB | Platelet-derived growth factor receptor beta |
| FGFR2 | Fibroblast growth factor receptor 2 |
| KIT | Mast/stem cell growth factor receptor Kit |
| TGFBR2 | TGF-beta receptor type-2 |
| FLT3 | Receptor-type tyrosine-protein kinase FLT3 |
| KDR | Vascular endothelial growth factor receptor 2 |
| FGFR3 | Fibroblast growth factor receptor 3 |
| FGFR4 | Fibroblast growth factor receptor 4 |
| MET | Hepatocyte growth factor receptor |
| JAK2 | Tyrosine-protein kinase JAK2 |
| JAK1 | Tyrosine-protein kinase JAK1 |
| JAK3 | Tyrosine-protein kinase JAK3 |
| PDGFRB | Platelet-derived growth factor receptor beta |
| FGFR2 | Fibroblast growth factor receptor 2 |

| | |
|---|---|
| KIT | Mast/stem cell growth factor receptor Kit |
| PIK3CA | Phosphatidylinositol 4,5-bisphosphate 3-kinase catalytic subunit alpha isoform |
| MAP2K4 | Dual specificity mitogen-activated protein kinase kinase 4 |
| CDK4 | Cyclin-dependent kinase 4 |
| CHEK2 | Serine/threonine-protein kinase Chk2 |
| PDGFRA | Platelet-derived growth factor receptor alpha |
| ESR1 | Estrogen receptor |
| ESR1 | Estrogen receptor |
| ESR1 | Estrogen receptor |
| FLT3 | Receptor-type tyrosine-protein kinase FLT3 |
| KDR | Vascular endothelial growth factor receptor 2 |
| FGFR3 | Fibroblast growth factor receptor 3 |
| FGFR4 | Fibroblast growth factor receptor 4 |
| PDGFRA | Platelet-derived growth factor receptor alpha |
| MET | Hepatocyte growth factor receptor |
| JAK2 | Tyrosine-protein kinase JAK2 |
| JAK3 | Tyrosine-protein kinase JAK3 |
| PDGFRB | Platelet-derived growth factor receptor beta |
| FGFR2 | Fibroblast growth factor receptor 2 |
| KIT | Mast/stem cell growth factor receptor Kit |
| TGFBR2 | TGF-beta receptor type-2 |
| MAP2K4 | Dual specificity mitogen-activated protein kinase kinase 4 |
| FLT3 | Receptor-type tyrosine-protein kinase FLT3 |
| KDR | Vascular endothelial growth factor receptor 2 |
| FGFR3 | Fibroblast growth factor receptor 3 |
| FGFR4 | Fibroblast growth factor receptor 4 |
| PDGFRA | Platelet-derived growth factor receptor alpha |
| MET | Hepatocyte growth factor receptor |
| PDGFRB | Platelet-derived growth factor receptor beta |
| FGFR2 | Fibroblast growth factor receptor 2 |
| KIT | Mast/stem cell growth factor receptor Kit |
| ERBB4 | Receptor tyrosine-protein kinase erbB-4 |
| FLT3 | Receptor-type tyrosine-protein kinase FLT3 |
| FGFR3 | Fibroblast growth factor receptor 3 |
| MET | Hepatocyte growth factor receptor |
| JAK2 | Tyrosine-protein kinase JAK2 |
| JAK1 | Tyrosine-protein kinase JAK1 |
| JAK3 | Tyrosine-protein kinase JAK3 |
| KIT | Mast/stem cell growth factor receptor Kit |
| JAK2 | Tyrosine-protein kinase JAK2 |
| JAK2 | JAK2/TYK2 |
| JAK1 | JAK1/JAK2/TYK2 |
| JAK1 | JAK1/TYK2 |
| JAK1 | Tyrosine-protein kinase JAK1 |

| | |
|---|---|
| JAK1 | JAK3/JAK1 |
| JAK1 | JAK2/JAK1 |
| JAK3 | Tyrosine-protein kinase JAK3 |
| KIT | Mast/stem cell growth factor receptor Kit |
| MAP2K4 | Dual specificity mitogen-activated protein kinase kinase 4 |
| FLT3 | Receptor-type tyrosine-protein kinase FLT3 |
| KDR | Vascular endothelial growth factor receptor 2 |
| FGFR3 | Fibroblast growth factor receptor 3 |
| PDGFRA | Platelet-derived growth factor receptor alpha |
| MET | Hepatocyte growth factor receptor |
| JAK2 | Tyrosine-protein kinase JAK2 |
| JAK3 | Tyrosine-protein kinase JAK3 |
| PDGFRB | Platelet-derived growth factor receptor beta |
| FGFR2 | Fibroblast growth factor receptor 2 |
| KIT | Mast/stem cell growth factor receptor Kit |
| SMO | Smoothened homolog |
| FLT3 | Receptor-type tyrosine-protein kinase FLT3 |
| MET | Hepatocyte growth factor receptor |
| ERBB4 | Receptor tyrosine-protein kinase erbB-4 |
| FLT3 | Receptor-type tyrosine-protein kinase FLT3 |
| PDGFRA | Platelet-derived growth factor receptor alpha |
| MET | Hepatocyte growth factor receptor |
| JAK2 | Tyrosine-protein kinase JAK2 |
| JAK3 | Tyrosine-protein kinase JAK3 |
| PDGFRB | Platelet-derived growth factor receptor beta |
| KIT | Mast/stem cell growth factor receptor Kit |
| ERBB4 | Receptor tyrosine-protein kinase erbB-4 |
| MAP2K4 | Dual specificity mitogen-activated protein kinase kinase 4 |
| CHEK2 | Serine/threonine-protein kinase Chk2 |
| AR | Androgen receptor |
| KDR | Vascular endothelial growth factor receptor 2 |
| JAK3 | Tyrosine-protein kinase JAK3 |
| JAK2 | Tyrosine-protein kinase JAK2 |
| JAK2 | JAK2/TYK2 |
| JAK1 | JAK1/JAK2/TYK2 |
| JAK1 | JAK1/TYK2 |
| JAK1 | Tyrosine-protein kinase JAK1 |
| JAK1 | JAK2/JAK1 |
| JAK1 | JAK3/JAK1 |
| KDR | Vascular endothelial growth factor receptor 2 |
| MET | Hepatocyte growth factor receptor |
| FLT3 | Receptor-type tyrosine-protein kinase FLT3 |
| KIT | Mast/stem cell growth factor receptor Kit |
| KIT | Mast/stem cell growth factor receptor Kit |

| | |
|---|---|
| KDR | Vascular endothelial growth factor receptor 2 |
| PDGFRA | Platelet-derived growth factor receptor alpha |
| FLT3 | Receptor-type tyrosine-protein kinase FLT3 |
| FGFR2 | Fibroblast growth factor receptor 2 |
| ERBB4 | Receptor tyrosine-protein kinase erbB-4 |
| JAK3 | Tyrosine-protein kinase JAK3 |
| FLT3 | Receptor-type tyrosine-protein kinase FLT3 |
| KDR | Vascular endothelial growth factor receptor 2 |
| FGFR3 | Fibroblast growth factor receptor 3 |
| FGFR4 | Fibroblast growth factor receptor 4 |
| PDGFRA | Platelet-derived growth factor receptor alpha |
| MET | Hepatocyte growth factor receptor |
| JAK2 | Tyrosine-protein kinase JAK2 |
| JAK1 | Tyrosine-protein kinase JAK1 |
| JAK3 | Tyrosine-protein kinase JAK3 |
| FGFR2 | Fibroblast growth factor receptor 2 |
| KIT | Mast/stem cell growth factor receptor Kit |
| CDK4 | Cyclin-dependent kinase 4 |
| PIK3CA | Phosphatidylinositol 4,5-bisphosphate 3-kinase catalytic subunit alpha isoform |
| KDR | Vascular endothelial growth factor receptor 2 |
| PDGFRA | Platelet-derived growth factor receptor alpha |
| FLT3 | Receptor-type tyrosine-protein kinase FLT3 |
| FGFR3 | Fibroblast growth factor receptor 3 |
| FGFR4 | Fibroblast growth factor receptor 4 |
| MET | Hepatocyte growth factor receptor |
| JAK2 | Tyrosine-protein kinase JAK2 |
| JAK1 | Tyrosine-protein kinase JAK1 |
| JAK3 | Tyrosine-protein kinase JAK3 |
| PDGFRB | Platelet-derived growth factor receptor beta |
| FGFR2 | Fibroblast growth factor receptor 2 |
| KIT | Mast/stem cell growth factor receptor Kit |
| MAP2K4 | Dual specificity mitogen-activated protein kinase kinase 4 |
| CDK4 | Cyclin-dependent kinase 4 |
| KDR | Vascular endothelial growth factor receptor 2 |
| KDR | Vascular endothelial growth factor receptor 2 |
| CDK4 | Cyclin-dependent kinase 4 |
| CDK6 | Cyclin-dependent kinase 6 |
| FLT3 | Receptor-type tyrosine-protein kinase FLT3 |
| JAK3 | Tyrosine-protein kinase JAK3 |
| CCND1 | Cyclin-dependent kinase 4/cyclin D1 |
| CCND1 | CDK6/cyclin D1 |
| KDR | Vascular endothelial growth factor receptor 2 |
| PDGFRA | Platelet-derived growth factor receptor alpha |
| PDGFRB | Platelet-derived growth factor receptor beta |

| | |
|---|---|
| KIT | Mast/stem cell growth factor receptor Kit |
| CSF3R | Granulocyte colony-stimulating factor receptor |
| CSF3R | Granulocyte colony-stimulating factor receptor |
| CSF3R | Granulocyte colony-stimulating factor receptor |
| SMO | Smoothened homolog |
| BCL2 | Apoptosis regulator Bcl-2 |
| CD274 | Programmed cell death 1 ligand 1 |
| CSF3R | Granulocyte colony-stimulating factor receptor |
| PDGFRA | Platelet-derived growth factor receptor alpha |
| JAK1 | Tyrosine-protein kinase JAK1 |
| JAK2 | Tyrosine-protein kinase JAK2 |
| JAK3 | Tyrosine-protein kinase JAK3 |
| CDK4 | Cyclin-dependent kinase 4 |
| CDK6 | Cyclin-dependent kinase 6 |
| CD274 | Programmed cell death 1 ligand 1 |
| BMPR1A | Bone morphogenetic protein receptor type-1A |
| HIF1A | Hypoxia-inducible factor 1-alpha |
| DNMT3A | DNA (cytosine-5)-methyltransferase 3A |
| RARA | Retinoic acid receptor alpha |
| ABL1 | Tyrosine-protein kinase ABL1 |
| SYK | Tyrosine-protein kinase SYK |
| RAF1 | RAF proto-oncogene serine/threonine-protein kinase |
| MAP3K13 | Mitogen-activated protein kinase kinase kinase 13 |
| BRAF | Serine/threonine-protein kinase B-raf |
| FGFR1 | Fibroblast growth factor receptor 1 |
| ERBB3 | Receptor tyrosine-protein kinase erbB-3 |
| RET | Proto-oncogene tyrosine-protein kinase receptor Ret |
| ABL1 | Bcr/Abl fusion protein |
| DNMT3A | DNA (cytosine-5)-methyltransferase 3A |
| ABL1 | Tyrosine-protein kinase ABL1 |
| ERBB3 | Receptor tyrosine-protein kinase erbB-3 |
| RET | Proto-oncogene tyrosine-protein kinase receptor Ret |
| ALK | ALK tyrosine kinase receptor |
| ABL1 | Tyrosine-protein kinase ABL1 |
| ERBB3 | Receptor tyrosine-protein kinase erbB-3 |
| RET | Proto-oncogene tyrosine-protein kinase receptor Ret |
| FGFR1 | Fibroblast growth factor receptor 1 |
| ABL1 | Tyrosine-protein kinase ABL1 |
| SYK | Tyrosine-protein kinase SYK |
| RAF1 | RAF proto-oncogene serine/threonine-protein kinase |
| BRAF | Serine/threonine-protein kinase B-raf |
| ABL1 | Tyrosine-protein kinase ABL |
| ABL1 | Bcr/Abl fusion protein |
| ABL1 | Tyrosine-protein kinase ABL1 |

| | |
|---|---|
| ERBB3 | Receptor tyrosine-protein kinase erbB-3 |
| FGFR1 | Fibroblast growth factor receptor 1 |
| ABL1 | Tyrosine-protein kinase ABL1 |
| RAF1 | RAF proto-oncogene serine/threonine-protein kinase |
| BRAF | Serine/threonine-protein kinase B-raf |
| NTRK3 | NT-3 growth factor receptor |
| RET | Proto-oncogene tyrosine-protein kinase receptor Ret |
| ABL1 | Bcr/Abl fusion protein |
| ABL1 | Tyrosine-protein kinase ABL1 |
| RAF1 | RAF proto-oncogene serine/threonine-protein kinase |
| BRAF | Serine/threonine-protein kinase B-raf |
| NTRK1 | High affinity nerve growth factor receptor |
| FGFR1 | Fibroblast growth factor receptor 1 |
| NTRK3 | NT-3 growth factor receptor |
| RET | Proto-oncogene tyrosine-protein kinase receptor Ret |
| RET | Proto-oncogene tyrosine-protein kinase receptor Ret |
| ABL1 | Tyrosine-protein kinase ABL1 |
| MAP3K13 | Mitogen-activated protein kinase kinase kinase 13 |
| NTRK1 | High affinity nerve growth factor receptor |
| FGFR1 | Fibroblast growth factor receptor 1 |
| AKT2 | RAC-beta serine/threonine-protein kinase |
| NTRK3 | NT-3 growth factor receptor |
| STK11 | Serine/threonine-protein kinase STK11 |
| ALK | ALK tyrosine kinase receptor |
| ABL1 | Tyrosine-protein kinase ABL1 |
| SYK | Tyrosine-protein kinase SYK |
| RAF1 | RAF proto-oncogene serine/threonine-protein kinase |
| BRAF | Serine/threonine-protein kinase B-raf |
| FGFR1 | Fibroblast growth factor receptor 1 |
| ROS1 | Proto-oncogene tyrosine-protein kinase ROS |
| MAP3K1 | Mitogen-activated protein kinase kinase kinase 1 |
| RET | Proto-oncogene tyrosine-protein kinase receptor Ret |
| ALK | ALK tyrosine kinase receptor |
| ABL1 | Tyrosine-protein kinase ABL1 |
| FGFR1 | Fibroblast growth factor receptor 1 |
| ERBB3 | Receptor tyrosine-protein kinase erbB-3 |
| RET | Proto-oncogene tyrosine-protein kinase receptor Ret |
| ALK | ALK tyrosine kinase receptor |
| ABL1 | Tyrosine-protein kinase ABL1 |
| SYK | Tyrosine-protein kinase SYK |
| MAP3K13 | Mitogen-activated protein kinase kinase kinase 13 |
| NTRK1 | High affinity nerve growth factor receptor |
| ALK | NPM/ALK (Nucleophosmin/ALK tyrosine kinase receptor) |
| NTRK3 | NT-3 growth factor receptor |

| | |
|---|---|
| ROS1 | Proto-oncogene tyrosine-protein kinase ROS |
| MAP3K1 | Mitogen-activated protein kinase kinase kinase 1 |
| RET | Proto-oncogene tyrosine-protein kinase receptor Ret |
| ALK | ALK tyrosine kinase receptor |
| ABL1 | Tyrosine-protein kinase ABL1 |
| BRAF | Serine/threonine-protein kinase B-raf |
| NTRK1 | High affinity nerve growth factor receptor |
| NTRK3 | NT-3 growth factor receptor |
| MAP3K1 | Mitogen-activated protein kinase kinase kinase 1 |
| RET | Proto-oncogene tyrosine-protein kinase receptor Ret |
| ALK | ALK tyrosine kinase receptor |
| ABL1 | Tyrosine-protein kinase ABL1 |
| NTRK1 | High affinity nerve growth factor receptor |
| FGFR1 | Fibroblast growth factor receptor 1 |
| ROS1 | Proto-oncogene tyrosine-protein kinase ROS |
| RET | Proto-oncogene tyrosine-protein kinase receptor Ret |
| ABL1 | Tyrosine-protein kinase ABL1 |
| ERBB3 | Receptor tyrosine-protein kinase erbB-3 |
| ABL1 | Tyrosine-protein kinase ABL1 |
| SYK | Tyrosine-protein kinase SYK |
| MAP3K13 | Mitogen-activated protein kinase kinase kinase 13 |
| NTRK1 | High affinity nerve growth factor receptor |
| ALK | NPM/ALK (Nucleophosmin/ALK tyrosine kinase receptor) |
| NTRK3 | NT-3 growth factor receptor |
| ERBB3 | Receptor tyrosine-protein kinase erbB-3 |
| MAP3K1 | Mitogen-activated protein kinase kinase kinase 1 |
| ABL1 | Tyrosine-protein kinase ABL |
| RET | Proto-oncogene tyrosine-protein kinase receptor Ret |
| ALK | ALK tyrosine kinase receptor |
| ABL1 | Bcr/Abl fusion protein |
| CXCR4 | C-X-C chemokine receptor type 4 |
| BRAF | Serine/threonine-protein kinase B-raf |
| ABL1 | Tyrosine-protein kinase ABL1 |
| RET | Proto-oncogene tyrosine-protein kinase receptor Ret |
| RET | Proto-oncogene tyrosine-protein kinase receptor Ret |
| ABL1 | Tyrosine-protein kinase ABL1 |
| FGFR1 | Fibroblast growth factor receptor 1 |
| ABL1 | Bcr/Abl fusion protein |
| RET | Proto-oncogene tyrosine-protein kinase receptor Ret |
| BRAF | Serine/threonine-protein kinase B-raf |
| RAF1 | RAF proto-oncogene serine/threonine-protein kinase |
| RET | Proto-oncogene tyrosine-protein kinase receptor Ret |
| ALK | ALK tyrosine kinase receptor |
| ABL1 | Tyrosine-protein kinase ABL1 |

| | |
|---|---|
| SYK | Tyrosine-protein kinase SYK |
| NTRK1 | High affinity nerve growth factor receptor |
| ROS1 | Proto-oncogene tyrosine-protein kinase ROS |
| RET | Proto-oncogene tyrosine-protein kinase receptor Ret |
| FGFR1 | Fibroblast growth factor receptor 1 |
| ABL1 | Tyrosine-protein kinase ABL1 |
| MAP3K13 | Mitogen-activated protein kinase kinase kinase 13 |
| NTRK1 | High affinity nerve growth factor receptor |
| NTRK3 | NT-3 growth factor receptor |
| STK11 | Serine/threonine-protein kinase STK11 |
| ROS1 | Proto-oncogene tyrosine-protein kinase ROS |
| RET | Proto-oncogene tyrosine-protein kinase receptor Ret |
| ALK | ALK tyrosine kinase receptor |
| RET | Proto-oncogene tyrosine-protein kinase receptor Ret |
| ALK | ALK tyrosine kinase receptor |
| FGFR1 | Fibroblast growth factor receptor 1 |
| RET | Proto-oncogene tyrosine-protein kinase receptor Ret |
| BCR | Breakpoint cluster region protein |
| PTK6 | Protein-tyrosine kinase 6 |
| ACVR1 | Activin receptor type-1 |
| ABL2 | Abelson tyrosine-protein kinase 2 |
| LCK | Tyrosine-protein kinase Lck |
| LCK | Tyrosine-protein kinase Lck |
| TERT | Telomerase reverse transcriptase |
| ABL2 | Abelson tyrosine-protein kinase 2 |
| FLT4 | Vascular endothelial growth factor receptor 3 |
| LCK | Tyrosine-protein kinase Lck |
| ABL2 | Abelson tyrosine-protein kinase 2 |
| LCK | Tyrosine-protein kinase Lck |
| ABL2 | Abelson tyrosine-protein kinase 2 |
| LCK | Tyrosine-protein kinase Lck |
| ABL2 | Abelson tyrosine-protein kinase 2 |
| LCK | Tyrosine-protein kinase Lck |
| ABL2 | Abelson tyrosine-protein kinase 2 |
| FLT4 | Vascular endothelial growth factor receptor 3 |
| LCK | Tyrosine-protein kinase Lck |
| ITK | Tyrosine-protein kinase ITK/TSK |
| PTK6 | Protein-tyrosine kinase 6 |
| ABL2 | Abelson tyrosine-protein kinase 2 |
| FLT4 | Vascular endothelial growth factor receptor 3 |
| LCK | Tyrosine-protein kinase Lck |
| LCK | Tyrosine-protein kinase Lck |
| PTK6 | Protein-tyrosine kinase 6 |
| ABL2 | Abelson tyrosine-protein kinase 2 |

| | |
|---|---|
| FLT4 | Vascular endothelial growth factor receptor 3 |
| LCK | Tyrosine-protein kinase Lck |
| ITK | Tyrosine-protein kinase ITK/TSK |
| PTK6 | Protein-tyrosine kinase 6 |
| ACVR1 | Activin receptor type-1 |
| ABL2 | Abelson tyrosine-protein kinase 2 |
| FLT4 | Vascular endothelial growth factor receptor 3 |
| LCK | Tyrosine-protein kinase Lck |
| ITK | Tyrosine-protein kinase ITK/TSK |
| IKBKB | Inhibitor of nuclear factor kappa-B kinase subunit beta |
| ACVR1 | Activin receptor type-1 |
| ABL2 | Abelson tyrosine-protein kinase 2 |
| LCK | Tyrosine-protein kinase Lck |
| IKBKB | Inhibitor of nuclear factor kappa-B kinase subunit beta |
| ITK | Tyrosine-protein kinase ITK/TSK |
| ABL2 | Abelson tyrosine-protein kinase 2 |
| FLT4 | Vascular endothelial growth factor receptor 3 |
| LCK | Tyrosine-protein kinase Lck |
| LCK | Tyrosine-protein kinase Lck |
| ITK | Tyrosine-protein kinase ITK/TSK |
| ABL2 | Abelson tyrosine-protein kinase 2 |
| LCK | Tyrosine-protein kinase Lck |
| LCK | Tyrosine-protein kinase Lck |
| ACKR3 | Atypical chemokine receptor 3 |
| LCK | Tyrosine-protein kinase Lck |
| BCR | Breakpoint cluster region protein |
| LCK | Tyrosine-protein kinase Lck |
| ITK | Tyrosine-protein kinase ITK/TSK |
| ETV6 | Transcription factor ETV6 |
| LCK | Tyrosine-protein kinase Lck |
| FLT4 | Vascular endothelial growth factor receptor 3 |
| ITK | Tyrosine-protein kinase ITK/TSK |
| ACVR1 | Activin receptor type-1 |
| ABL2 | Abelson tyrosine-protein kinase 2 |
| LCK | Tyrosine-protein kinase Lck |
| FLT4 | Vascular endothelial growth factor receptor 3 |
| AURKA | Aurora kinase A |
| AURKB | Aurora kinase B |
| ACVR1B | Activin receptor type-1B |
| AURKA | Aurora kinase A |
| AURKA | Aurora kinase A |
| AURKB | Aurora kinase B |
| TEK | Angiopoietin-1 receptor |
| NTRK2 | BDNF/NT-3 growth factors receptor |

| | |
|---|---|
| AURKA | Aurora kinase A |
| AURKB | Aurora kinase B |
| TEK | Angiopoietin-1 receptor |
| NTRK2 | BDNF/NT-3 growth factors receptor |
| AURKA | Aurora kinase A |
| AURKB | Aurora kinase B |
| NTRK2 | BDNF/NT-3 growth factors receptor |
| AURKA | Aurora kinase A |
| TEK | Angiopoietin-1 receptor |
| TEK | Angiopoietin-1 receptor |
| AURKA | Aurora kinase A |
| AURKB | Aurora kinase B |
| TEK | Angiopoietin-1 receptor |
| ACVR1B | Activin receptor type-1B |
| NTRK2 | BDNF/NT-3 growth factors receptor |
| NTRK2 | BDNF/NT-3 growth factors receptor |
| AURKA | Aurora kinase A |
| AURKB | Aurora kinase B |
| TEK | Angiopoietin-1 receptor |
| TEK | Angiopoietin-1 receptor |
| NTRK2 | BDNF/NT-3 growth factors receptor |
| TEK | Angiopoietin-1 receptor |
| NTRK2 | BDNF/NT-3 growth factors receptor |
| AURKB | Aurora kinase B |
| TEK | Angiopoietin-1 receptor |
| ACVR1B | Activin receptor type-1B |
| NTRK2 | BDNF/NT-3 growth factors receptor |
| GSK3B | Glycogen synthase kinase-3 beta |
| PARP2 | Poly [ADP-ribose] polymerase 2 |
| VEGFA | Vascular endothelial growth factor A |
| PDCD1 | Programmed cell death protein 1 |
| PDCD1 | Programmed cell death protein 1 |
| VEGFA | Vascular endothelial growth factor A |
| BCL2L1 | Bcl-2-like protein 1 |
| PARP2 | Poly [ADP-ribose] polymerase 2 |
| PARP2 | Poly [ADP-ribose] polymerase 2 |
| TYMS | Thymidylate synthase |
| HDAC10 | Histone deacetylase 10 |
| HDAC6 | Histone deacetylase 6 |
| TYMS | Thymidylate synthase |
| RRM2 | Ribonucleoside-diphosphate reductase subunit M2 |
| WEE1 | Wee1-like protein kinase |
| PKMYT1 | Membrane-associated tyrosine- and threonine-specific cdc2-inhibitory kinase |
| BMPR1B | Bone morphogenetic protein receptor type-1B |

| | |
|---|---|
| TYK2 | Non-receptor tyrosine-protein kinase TYK2 |
| PIK3CD | Phosphatidylinositol 4,5-bisphosphate 3-kinase catalytic subunit delta isoform |
| ACVRL1 | Serine/threonine-protein kinase receptor R3 |
| MAPK10 | Mitogen-activated protein kinase 10 |
| TTK | Dual specificity protein kinase TTK |
| TIE1 | Tyrosine-protein kinase receptor Tie-1 |
| TYK2 | Non-receptor tyrosine-protein kinase TYK2 |
| TYMS | Thymidylate synthase |
| PGR | Progesterone receptor |
| MAPK10 | Mitogen-activated protein kinase 10 |
| TYMS | Thymidylate synthase |
| RRM2 | Ribonucleoside-diphosphate reductase subunit M2 |
| MAPK10 | Mitogen-activated protein kinase 10 |
| MELK | Maternal embryonic leucine zipper kinase |
| TYK2 | Non-receptor tyrosine-protein kinase TYK2 |
| TYMS | Thymidylate synthase |
| CDC25A | Dual specificity phosphatase Cdc25A |
| IDO1 | Indoleamine 2,3-dioxygenase 1 |
| MAPK10 | Mitogen-activated protein kinase 10 |
| TIE1 | Tyrosine-protein kinase receptor Tie-1 |
| TYMS | Thymidylate synthase |
| TYMS | Thymidylate synthase |
| TTK | Dual specificity protein kinase TTK |
| TIE1 | Tyrosine-protein kinase receptor Tie-1 |
| MAPK10 | Mitogen-activated protein kinase 10 |
| CAMK2B | Calcium/calmodulin-dependent protein kinase type II subunit beta |
| MELK | Maternal embryonic leucine zipper kinase |
| WEE1 | Wee1-like protein kinase |
| TTK | Dual specificity protein kinase TTK |
| RPS6KA5 | Ribosomal protein S6 kinase alpha-5 |
| ROCK1 | Rho-associated protein kinase 1 |
| BMPR1B | Bone morphogenetic protein receptor type-1B |
| TYK2 | Non-receptor tyrosine-protein kinase TYK2 |
| BMPR2 | Bone morphogenetic protein receptor type-2 |
| PIK3CD | Phosphatidylinositol 4,5-bisphosphate 3-kinase catalytic subunit delta isoform |
| TNKS2 | Tankyrase-2 |
| PRKAA2 | 5'-AMP-activated protein kinase catalytic subunit alpha-2 |
| TIE1 | Tyrosine-protein kinase receptor Tie-1 |
| MAP3K12 | Mitogen-activated protein kinase kinase kinase 12 |
| RPS6KB1 | Ribosomal protein S6 kinase beta-1 |
| PTGS2 | Prostaglandin G/H synthase 2 |
| PGR | Progesterone receptor |
| TNF | Tumor necrosis factor |
| PTGS2 | Prostaglandin G/H synthase 2 |

| | |
|---|---|
| NOD2 | Nucleotide-binding oligomerization domain-containing protein 2 |
| TNF | Tumor necrosis factor |
| MAPK10 | Mitogen-activated protein kinase 10 |
| TTK | Dual specificity protein kinase TTK |
| BMPR1B | Bone morphogenetic protein receptor type-1B |
| TYK2 | Non-receptor tyrosine-protein kinase TYK2 |
| TIE1 | Tyrosine-protein kinase receptor Tie-1 |
| HDAC6 | Histone deacetylase 6 |
| HDAC5 | Histone deacetylase 5 |
| HDAC10 | Histone deacetylase 10 |
| HDAC11 | Histone deacetylase 11 |
| HDAC6 | Histone deacetylase 6 |
| HDAC5 | Histone deacetylase 5 |
| S1PR1 | Sphingosine 1-phosphate receptor 1 |
| BMPR1B | Bone morphogenetic protein receptor type-1B |
| ACVRL1 | Serine/threonine-protein kinase receptor R3 |
| TIE1 | Tyrosine-protein kinase receptor Tie-1 |
| RPS6KB1 | Ribosomal protein S6 kinase beta-1 |
| ROCK1 | Rho-associated protein kinase 1 |
| BMPR1B | Bone morphogenetic protein receptor type-1B |
| TYK2 | Non-receptor tyrosine-protein kinase TYK2 |
| TIE1 | Tyrosine-protein kinase receptor Tie-1 |
| MAP3K12 | Mitogen-activated protein kinase kinase kinase 12 |
| RPS6KB1 | Ribosomal protein S6 kinase beta-1 |
| CAMK2B | Calcium/calmodulin-dependent protein kinase type II subunit beta |
| MELK | Maternal embryonic leucine zipper kinase |
| TTK | Dual specificity protein kinase TTK |
| RPS6KA5 | Ribosomal protein S6 kinase alpha-5 |
| ROCK1 | Rho-associated protein kinase 1 |
| BMPR1B | Bone morphogenetic protein receptor type-1B |
| TYK2 | Non-receptor tyrosine-protein kinase TYK2 |
| BMPR2 | Bone morphogenetic protein receptor type-2 |
| MAPK10 | Mitogen-activated protein kinase 10 |
| TTK | Dual specificity protein kinase TTK |
| BMPR1B | Bone morphogenetic protein receptor type-1B |
| TYK2 | Non-receptor tyrosine-protein kinase TYK2 |
| TIE1 | Tyrosine-protein kinase receptor Tie-1 |
| MAP3K12 | Mitogen-activated protein kinase kinase kinase 12 |
| MAPK10 | Mitogen-activated protein kinase 10 |
| WEE1 | Wee1-like protein kinase |
| ROCK1 | Rho-associated protein kinase 1 |
| PKMYT1 | Membrane-associated tyrosine- and threonine-specific cdc2-inhibitory kinase |
| TYK2 | Non-receptor tyrosine-protein kinase TYK2 |
| BMPR2 | Bone morphogenetic protein receptor type-2 |

| | |
|---|---|
| TIE1 | Tyrosine-protein kinase receptor Tie-1 |
| MAP3K12 | Mitogen-activated protein kinase kinase kinase 12 |
| RPS6KB1 | Ribosomal protein S6 kinase beta-1 |
| CCR2 | C-C chemokine receptor type 2 |
| CXCL12 | Stromal cell-derived factor 1 |
| CCR2 | C-C chemokine receptor type 2 |
| HDAC10 | Histone deacetylase 10 |
| HDAC11 | Histone deacetylase 11 |
| HDAC6 | Histone deacetylase 6 |
| HDAC5 | Histone deacetylase 5 |
| ROCK1 | Rho-associated protein kinase 1 |
| TYK2 | Non-receptor tyrosine-protein kinase TYK2 |
| HDAC10 | Histone deacetylase 10 |
| HDAC11 | Histone deacetylase 11 |
| HDAC6 | Histone deacetylase 6 |
| HDAC5 | Histone deacetylase 5 |
| PIK3CD | Phosphatidylinositol 4,5-bisphosphate 3-kinase catalytic subunit delta isoform |
| MAPK10 | Mitogen-activated protein kinase 10 |
| MELK | Maternal embryonic leucine zipper kinase |
| WEE1 | Wee1-like protein kinase |
| TTK | Dual specificity protein kinase TTK |
| RPS6KA5 | Ribosomal protein S6 kinase alpha-5 |
| TYK2 | Non-receptor tyrosine-protein kinase TYK2 |
| BMPR2 | Bone morphogenetic protein receptor type-2 |
| PRKAA2 | 5'-AMP-activated protein kinase catalytic subunit alpha-2 |
| ACVRL1 | Serine/threonine-protein kinase receptor R3 |
| TIE1 | Tyrosine-protein kinase receptor Tie-1 |
| MAP3K12 | Mitogen-activated protein kinase kinase kinase 12 |
| RPS6KB1 | Ribosomal protein S6 kinase beta-1 |
| TNF | Tumor necrosis factor |
| TNKS | Tankyrase-1 |
| CD19 | B-lymphocyte antigen CD19 |
| IL6R | Interleukin-6 receptor |
| IL2RA | Interleukin-2 receptor subunit alpha |
| CCNA2 | CDK2/Cyclin A |
| IL1B | Interleukin-1 beta |
| IL2RA | Interleukin-2 receptor |
| TNF | Tumor necrosis factor |
| TNF | Tumor necrosis factor |
| TNF | Tumor necrosis factor |
| IL6 | Interleukin-6 |
| TNF | Tumor necrosis factor |
| IL2RA | Interleukin-2 receptor |
| IL11RA | Interleukin-11 receptor |

| | |
|---|---|
| IL2RA | Interleukin-2 receptor |
| IL1B | Interleukin-1 beta |
| TYK2 | Non-receptor tyrosine-protein kinase TYK2 |
| TNKS | Tankyrase-1 |
| TNKS2 | Tankyrase-2 |
| FLT1 | Vascular endothelial growth factor receptor 1 |
| SRC | Proto-oncogene tyrosine-protein kinase Src |
| EPHB4 | Ephrin type-B receptor 4 |
| BTK | Tyrosine-protein kinase BTK |
| EPHA3 | Ephrin type-A receptor 3 |
| EPHB1 | Ephrin type-B receptor 1 |
| CSF1R | Macrophage colony-stimulating factor 1 receptor |
| DDR1 | Epithelial discoidin domain-containing receptor 1 |
| FLT1 | Vascular endothelial growth factor receptor 1 |
| SRC | Proto-oncogene tyrosine-protein kinase Src |
| EPHB4 | Ephrin type-B receptor 4 |
| MKNK1 | MAP kinase-interacting serine/threonine-protein kinase 1 |
| EPHA3 | Ephrin type-A receptor 3 |
| DDR1 | Epithelial discoidin domain-containing receptor 1 |
| TYRO3 | Tyrosine-protein kinase receptor TYRO3 |
| LTK | Leukocyte tyrosine kinase receptor |
| AXL | Tyrosine-protein kinase receptor UFO |
| MERTK | Tyrosine-protein kinase Mer |
| EPHB1 | Ephrin type-B receptor 1 |
| SRC | Proto-oncogene tyrosine-protein kinase Src |
| EPHB4 | Ephrin type-B receptor 4 |
| MKNK1 | MAP kinase-interacting serine/threonine-protein kinase 1 |
| EPHA3 | Ephrin type-A receptor 3 |
| EPHB1 | Ephrin type-B receptor 1 |
| LTK | Leukocyte tyrosine kinase receptor |
| AXL | Tyrosine-protein kinase receptor UFO |
| EPHB1 | Ephrin type-B receptor 1 |
| EPHB4 | Ephrin type-B receptor 4 |
| SRC | Proto-oncogene tyrosine-protein kinase Src |
| CSF1R | Macrophage colony-stimulating factor 1 receptor |
| DDR1 | Epithelial discoidin domain-containing receptor 1 |
| PIK3C2G | Phosphatidylinositol 4-phosphate 3-kinase C2 domain-containing subunit gamma |
| PIK3C2B | Phosphatidylinositol 4-phosphate 3-kinase C2 domain-containing subunit beta |
| BTK | Tyrosine-protein kinase BTK |
| SRC | Proto-oncogene tyrosine-protein kinase Src |
| EPHB4 | Ephrin type-B receptor 4 |
| EPHA3 | Ephrin type-A receptor 3 |
| EPHB1 | Ephrin type-B receptor 1 |
| CSF1R | Macrophage colony-stimulating factor 1 receptor |

| | |
|---|---|
| DDR1 | Epithelial discoidin domain-containing receptor 1 |
| CDK8 | Cyclin-dependent kinase 8 |
| FLT1 | Vascular endothelial growth factor receptor 1 |
| SRC | Proto-oncogene tyrosine-protein kinase Src |
| EPHB4 | Ephrin type-B receptor 4 |
| MKNK1 | MAP kinase-interacting serine/threonine-protein kinase 1 |
| EPHA3 | Ephrin type-A receptor 3 |
| EPHB1 | Ephrin type-B receptor 1 |
| CSF1R | Macrophage colony-stimulating factor 1 receptor |
| DDR1 | Epithelial discoidin domain-containing receptor 1 |
| AXL | Tyrosine-protein kinase receptor UFO |
| MERTK | Tyrosine-protein kinase Mer |
| SRC | Proto-oncogene tyrosine-protein kinase Src |
| BTK | Tyrosine-protein kinase BTK |
| MKNK1 | MAP kinase-interacting serine/threonine-protein kinase 1 |
| EPHA3 | Ephrin type-A receptor 3 |
| EPHB1 | Ephrin type-B receptor 1 |
| IKBKE | Inhibitor of nuclear factor kappa-B kinase subunit epsilon |
| CSF1R | Macrophage colony-stimulating factor 1 receptor |
| DDR1 | Epithelial discoidin domain-containing receptor 1 |
| TYRO3 | Tyrosine-protein kinase receptor TYRO3 |
| LTK | Leukocyte tyrosine kinase receptor |
| AXL | Tyrosine-protein kinase receptor UFO |
| CHEK1 | Serine/threonine-protein kinase Chk1 |
| MERTK | Tyrosine-protein kinase Mer |
| EPHB4 | Ephrin type-B receptor 4 |
| FLT1 | Vascular endothelial growth factor receptor 1 |
| FLT1 | Vascular endothelial growth factor receptor 1 |
| SRC | Proto-oncogene tyrosine-protein kinase Src |
| CSF1R | Macrophage colony-stimulating factor 1 receptor |
| DDR1 | Epithelial discoidin domain-containing receptor 1 |
| LTK | Leukocyte tyrosine kinase receptor |
| MERTK | Tyrosine-protein kinase Mer |
| FLT1 | Vascular endothelial growth factor receptor 1 |
| SRC | Proto-oncogene tyrosine-protein kinase Src |
| EPHB4 | Ephrin type-B receptor 4 |
| BTK | Tyrosine-protein kinase BTK |
| MKNK1 | MAP kinase-interacting serine/threonine-protein kinase 1 |
| EPHA3 | Ephrin type-A receptor 3 |
| MST1R | Macrophage-stimulating protein receptor |
| EPHB1 | Ephrin type-B receptor 1 |
| CSF1R | Macrophage colony-stimulating factor 1 receptor |
| DDR1 | Epithelial discoidin domain-containing receptor 1 |
| TYRO3 | Tyrosine-protein kinase receptor TYRO3 |

| | |
|---|---|
| LTK | Leukocyte tyrosine kinase receptor |
| AXL | Tyrosine-protein kinase receptor UFO |
| MERTK | Tyrosine-protein kinase Mer |
| FLT1 | Vascular endothelial growth factor receptor 1 |
| SRC | Proto-oncogene tyrosine-protein kinase Src |
| EPHB4 | Ephrin type-B receptor 4 |
| BTK | Tyrosine-protein kinase BTK |
| EPHA3 | Ephrin type-A receptor 3 |
| MST1R | Macrophage-stimulating protein receptor |
| EPHB1 | Ephrin type-B receptor 1 |
| IKBKE | Inhibitor of nuclear factor kappa-B kinase subunit epsilon |
| CSF1R | Macrophage colony-stimulating factor 1 receptor |
| DDR1 | Epithelial discoidin domain-containing receptor 1 |
| TYRO3 | Tyrosine-protein kinase receptor TYRO3 |
| LTK | Leukocyte tyrosine kinase receptor |
| AXL | Tyrosine-protein kinase receptor UFO |
| MERTK | Tyrosine-protein kinase Mer |
| EPHA3 | Ephrin type-A receptor 3 |
| IKBKE | Inhibitor of nuclear factor kappa-B kinase subunit epsilon |
| LTK | Leukocyte tyrosine kinase receptor |
| FLT1 | Vascular endothelial growth factor receptor 1 |
| CSF1R | Macrophage colony-stimulating factor 1 receptor |
| DDR1 | Epithelial discoidin domain-containing receptor 1 |
| AXL | Tyrosine-protein kinase receptor UFO |
| MERTK | Tyrosine-protein kinase Mer |
| SRC | Proto-oncogene tyrosine-protein kinase Src |
| MKNK1 | MAP kinase-interacting serine/threonine-protein kinase 1 |
| AXL | Tyrosine-protein kinase receptor UFO |
| SRC | Proto-oncogene tyrosine-protein kinase Src |
| EPHB4 | Ephrin type-B receptor 4 |
| BTK | Tyrosine-protein kinase BTK |
| EPHA3 | Ephrin type-A receptor 3 |
| MST1R | Macrophage-stimulating protein receptor |
| EPHB1 | Ephrin type-B receptor 1 |
| IKBKE | Inhibitor of nuclear factor kappa-B kinase subunit epsilon |
| CSF1R | Macrophage colony-stimulating factor 1 receptor |
| DDR1 | Epithelial discoidin domain-containing receptor 1 |
| TYRO3 | Tyrosine-protein kinase receptor TYRO3 |
| LTK | Leukocyte tyrosine kinase receptor |
| AXL | Tyrosine-protein kinase receptor UFO |
| CHEK1 | Serine/threonine-protein kinase Chk1 |
| MERTK | Tyrosine-protein kinase Mer |
| SRC | Proto-oncogene tyrosine-protein kinase Src |
| MST1R | Macrophage-stimulating protein receptor |

| | |
|---|---|
| SRC | Proto-oncogene tyrosine-protein kinase Src |
| CDK8 | Cyclin-dependent kinase 8 |
| KEAP1 | Kelch-like ECH-associated protein 1 |
| ARAF | Serine/threonine-protein kinase A-Raf |
| BTK | Tyrosine-protein kinase BTK |
| SRC | Proto-oncogene tyrosine-protein kinase Src |
| SRC | Proto-oncogene tyrosine-protein kinase Src |
| BTK | Tyrosine-protein kinase BTK |
| PIK3CB | Phosphatidylinositol 4,5-bisphosphate 3-kinase catalytic subunit beta isoform |
| FLT1 | Vascular endothelial growth factor receptor 1 |
| SRC | Proto-oncogene tyrosine-protein kinase Src |
| EPHB4 | Ephrin type-B receptor 4 |
| BTK | Tyrosine-protein kinase BTK |
| EPHA3 | Ephrin type-A receptor 3 |
| EPHB1 | Ephrin type-B receptor 1 |
| IKBKE | Inhibitor of nuclear factor kappa-B kinase subunit epsilon |
| CSF1R | Macrophage colony-stimulating factor 1 receptor |
| DDR1 | Epithelial discoidin domain-containing receptor 1 |
| TYRO3 | Tyrosine-protein kinase receptor TYRO3 |
| LTK | Leukocyte tyrosine kinase receptor |
| AXL | Tyrosine-protein kinase receptor UFO |
| CHEK1 | Serine/threonine-protein kinase Chk1 |
| MERTK | Tyrosine-protein kinase Mer |
| PARP3 | Poly [ADP-ribose] polymerase 3 |
| FLT1 | Vascular endothelial growth factor receptor 1 |
| BCL2L2 | Apoptosis regulator Bcl-W |
| PARP3 | Poly [ADP-ribose] polymerase 3 |

| Target class | Accession | Drug name |
| --- | --- | --- |
| Kinase | P00533 | dasatinib |
| Kinase | P42345 | dasatinib |
| Kinase | P04626 | dasatinib |
| Kinase | P00533 | docetaxel |
| Kinase | P04626 | doxorubicin |
| Kinase | P00533 | erlotinib |
| Kinase | P04626 | erlotinib |
| Kinase | P42345 | everolimus |
| Kinase | P00533 | gefitinib |
| Kinase | P04626 | gefitinib |
| Kinase | P00533 | imatinib |
| Kinase | P00533 | lapatinib |
| Kinase | P04626 | lapatinib |
| Kinase | P00533 | mitoxantrone |
| Kinase | P04626 | mitoxantrone |
| Kinase | P04626 | paclitaxel |
| Kinase | P42345 | sirolimus |
| Kinase | P00533 | sunitinib |
| Kinase | P00533 | tamoxifen |
| Kinase | P04626 | tamoxifen |
| Kinase | P42345 | temsirolimus |
| Kinase | P00533 | vandetanib |
| Kinase | P04626 | vandetanib |
| Kinase | P00533 | crizotinib |
| Kinase | P00533 | axitinib |
| Kinase | P00533 | afatinib |
| Kinase | P04626 | afatinib |
| Kinase | P00533 | bosutinib |
| Kinase | P04626 | bosutinib |
| Kinase | P00533 | cisplatin |
| Kinase | P42345 | ridaforolimus |
| Kinase | P00533 | ibrutinib |
| Kinase | P04626 | ibrutinib |
| Kinase | P00533 | ceritinib |
| Kinase | P00533 | lenvatinib |

| | | |
|---|---|---|
| Kinase | P00533 | cetuximab |
| Kinase | P00533 | panitumumab |
| Kinase | P04626 | pertuzumab |
| Kinase | P04626 | trastuzumab |
| Kinase | P04626 | trastuzumab emtansine |
| Kinase | P00533 | osimertinib |
| Kinase | P00533 | necitumumab |
| Kinase | P00533 | icotinib |
| Kinase | P00533 | olmutinib |
| Unclassified | P63000 | azathioprine |
| Kinase | Q02750 | dasatinib |
| Kinase | P36507 | dasatinib |
| Kinase | P36507 | dasatinib |
| Kinase | P31749 | miltefosine |
| Kinase | Q02750 | sunitinib |
| Kinase | P36507 | sunitinib |
| Kinase | Q02750 | vandetanib |
| Kinase | P36507 | vandetanib |
| Kinase | Q02750 | ruxolitinib |
| Kinase | P36507 | ruxolitinib |
| Kinase | Q02750 | axitinib |
| Kinase | P36507 | axitinib |
| Kinase | Q02750 | bosutinib |
| Kinase | P36507 | bosutinib |
| Kinase | P36507 | trametinib |
| Kinase | Q02750 | trametinib |
| Kinase | Q02750 | nintedanib |
| Kinase | P36507 | nintedanib |
| Kinase | Q02750 | cobimetinib |
| Kinase | P36507 | cobimetinib |
| Enzyme | P11387 | amsacrine |
| Enzyme | P11387 | doxorubicin |
| Enzyme | P11387 | irinotecan |
| Enzyme | P11387 | topotecan |
| Enzyme | Q13547 | bendamustine |
| Kinase | P08069 | masoprocol |
| Kinase | P28482 | sorafenib |
| Kinase | P08069 | sunitinib |
| Enzyme | O15379 | romidepsin |
| Enzyme | Q13547 | romidepsin |
| Enzyme | O15379 | vorinostat |
| Enzyme | O15379 | vorinostat |
| Enzyme | Q13547 | vorinostat |
| Enzyme | Q13547 | vorinostat |

| | | |
|---|---|---|
| Kinase | P08069 | crizotinib |
| Enzyme | Q13547 | panobinostat |
| Kinase | P08069 | ceritinib |
| Enzyme | Q13547 | belinostat |
| Kinase | P08069 | nintedanib |
| Enzyme | P09874 | olaparib |
| Enzyme | P09874 | rucaparib |
| Enzyme | P09874 | niraparib |
| Enzyme | P11388 | amsacrine |
| Enzyme | P11388 | amsacrine |
| Enzyme | Q92769 | bendamustine |
| Enzyme | P14780 | busulfan |
| Kinase | P27361 | carmofur |
| Enzyme | P14780 | dacarbazine |
| Cytosolic other | P62993 | dactinomycin |
| Enzyme | P11388 | daunorubicin |
| Enzyme | P11388 | doxorubicin |
| Enzyme | P11388 | etoposide |
| Enzyme | P11388 | idarubicin |
| Enzyme | O43570 | imatinib |
| Enzyme | P11388 | mitoxantrone |
| Enzyme | O43570 | nilotinib |
| Kinase | P27361 | sorafenib |
| Kinase | P48736 | sunitinib |
| Kinase | O75116 | sunitinib |
| Enzyme | P11388 | teniposide |
| Enzyme | P11388 | valrubicin |
| Enzyme | Q92769 | romidepsin |
| Enzyme | Q92769 | vorinostat |
| Kinase | O75116 | vandetanib |
| Kinase | O75116 | crizotinib |
| Kinase | O75116 | ruxolitinib |
| Kinase | O75116 | bosutinib |
| Kinase | P27361 | cisplatin |
| Enzyme | Q92769 | panobinostat |
| Kinase | O75116 | tofacitinib |
| Kinase | O75116 | ceritinib |
| Enzyme | Q92769 | belinostat |
| Kinase | P48736 | idelalisib |
| Kinase | P78527 | idelalisib |
| Unclassified | P18510 | rilonacept |
| Enzyme | Q9UKK3 | rucaparib |
| Kinase | P07948 | dasatinib |
| Kinase | P07948 | erlotinib |

| | | |
|---|---|---|
| Kinase | P07948 | gefitinib |
| Kinase | P07948 | imatinib |
| Kinase | P07948 | nilotinib |
| Kinase | P07948 | sorafenib |
| Kinase | P07948 | sunitinib |
| Kinase | P07948 | pazopanib |
| Kinase | P07948 | vandetanib |
| Kinase | P07948 | crizotinib |
| Kinase | P07948 | bosutinib |
| Kinase | P07948 | ponatinib |
| Kinase | P07948 | ibrutinib |
| Kinase | P07948 | ceritinib |
| Kinase | P07948 | nintedanib |
| Enzyme | P08253 | aclarubicin |
| Enzyme | P05177 | altretamine |
| Enzyme | Q02880 | amsacrine |
| Enzyme | Q16881 | arsenic trioxide |
| Enzyme | P51580 | azathioprine |
| Enzyme | P25786 | bortezomib |
| Enzyme | P00734 | bortezomib |
| Enzyme | P08311 | bortezomib |
| Enzyme | P23946 | bortezomib |
| Enzyme | P20618 | bortezomib |
| Enzyme | P28074 | bortezomib |
| Enzyme | P49721 | bortezomib |
| Unclassified | P19838 | bortezomib |
| Cytosolic other | Q99460 | bortezomib |
| Enzyme | P00390 | carmustine |
| Enzyme | P07339 | chlorambucil |
| Enzyme | P23921 | clofarabine |
| Enzyme | P06746 | cytarabine |
| Kinase | O14976 | dasatinib |
| Kinase | O94804 | dasatinib |
| Kinase | P41240 | dasatinib |
| Kinase | Q9H2G2 | dasatinib |
| Kinase | Q13043 | dasatinib |
| Kinase | Q9Y5S2 | dasatinib |
| Kinase | Q86Y07 | dasatinib |
| Kinase | Q16539 | dasatinib |
| Transporter | P08183 | daunorubicin |
| Enzyme | P08253 | daunorubicin |
| Structural | P04350 | docetaxel |
| Transporter | P08183 | docetaxel |
| Enzyme | P29466 | doxorubicin |

| | | |
|---|---|---|
| Transporter | P08183 | doxorubicin |
| Enzyme | P08253 | doxorubicin |
| Kinase | O14976 | erlotinib |
| Kinase | P19525 | erlotinib |
| Kinase | O94804 | erlotinib |
| Kinase | P41240 | erlotinib |
| Kinase | Q8TBX8 | erlotinib |
| Kinase | Q13557 | erlotinib |
| Kinase | Q9H2G2 | erlotinib |
| Transporter | P08183 | etoposide |
| Enzyme | P62942 | everolimus |
| Transporter | P08183 | everolimus |
| Enzyme | P05177 | flutamide |
| Enzyme | P34913 | fulvestrant |
| Kinase | O14976 | gefitinib |
| Kinase | O94804 | gefitinib |
| Kinase | Q8TBX8 | gefitinib |
| Kinase | Q9H2G2 | gefitinib |
| Kinase | P30085 | gemcitabine |
| Enzyme | P23921 | gemcitabine |
| Transporter | P08183 | idarubicin |
| Enzyme | P00918 | imatinib |
| Transporter | P08183 | imatinib |
| Kinase | O14976 | imatinib |
| Kinase | Q8TBX8 | imatinib |
| Enzyme | Q16790 | imatinib |
| Enzyme | P00915 | imatinib |
| Kinase | Q16539 | imatinib |
| Transporter | P08183 | irinotecan |
| Structural | P04350 | ixabepilone |
| Transporter | P08183 | lapatinib |
| Kinase | O94804 | lapatinib |
| Kinase | Q9H2G2 | lapatinib |
| Enzyme | P05177 | leflunomide |
| Enzyme | P21397 | leflunomide |
| Enzyme | O15296 | masoprocol |
| Secreted | P02766 | masoprocol |
| Enzyme | Q06203 | mercaptopurine |
| Enzyme | P23219 | mercaptopurine |
| Enzyme | P51580 | mercaptopurine |
| Enzyme | P00491 | mercaptopurine |
| Transporter | P08183 | methotrexate |
| Transporter | P08183 | mitoxantrone |
| Enzyme | P29372 | mitoxantrone |

| | | |
|---|---|---|
| Enzyme | P12268 | mycophenolic acid |
| Enzyme | P00918 | nilotinib |
| Kinase | O94804 | nilotinib |
| Kinase | P41240 | nilotinib |
| Kinase | Q8TBX8 | nilotinib |
| Kinase | Q9Y5S2 | nilotinib |
| Enzyme | Q16790 | nilotinib |
| Enzyme | P00915 | nilotinib |
| Kinase | Q16539 | nilotinib |
| Structural | P04350 | paclitaxel |
| Structural | P07437 | paclitaxel |
| Transporter | P08183 | paclitaxel |
| Structural | Q13509 | paclitaxel |
| Enzyme | P34896 | pemetrexed |
| Enzyme | P00813 | pentostatin |
| Enzyme | P62942 | sirolimus |
| Enzyme | P11712 | sirolimus |
| Enzyme | Q00688 | sirolimus |
| Transporter | P08183 | sirolimus |
| Unclassified | Q53EL6 | sirolimus |
| Enzyme | Q13451 | sirolimus |
| Kinase | O94804 | sorafenib |
| Kinase | P41240 | sorafenib |
| Kinase | Q9H2G2 | sorafenib |
| Enzyme | P34913 | sorafenib |
| Kinase | O15264 | sorafenib |
| Kinase | Q16539 | sorafenib |
| Kinase | O15075 | sunitinib |
| Kinase | O14976 | sunitinib |
| Kinase | P19525 | sunitinib |
| Unclassified | O43172 | sunitinib |
| Kinase | O95747 | sunitinib |
| Kinase | Q96BR1 | sunitinib |
| Kinase | O94804 | sunitinib |
| Kinase | Q7KZI7 | sunitinib |
| Kinase | P19784 | sunitinib |
| Kinase | Q16513 | sunitinib |
| Kinase | Q15746 | sunitinib |
| Kinase | Q9H2G2 | sunitinib |
| Kinase | Q13043 | sunitinib |
| Kinase | Q05397 | sunitinib |
| Kinase | Q13523 | sunitinib |
| Kinase | Q9Y6E0 | sunitinib |
| Kinase | P78356 | sunitinib |

| | | |
|---|---|---|
| Kinase | P78362 | sunitinib |
| Kinase | Q13557 | sunitinib |
| Enzyme | P23219 | tamoxifen |
| Enzyme | Q08209 | tamoxifen |
| Enzyme | Q06278 | tamoxifen |
| Transporter | P08183 | tamoxifen |
| Transporter | P08183 | teniposide |
| Enzyme | P23219 | thalidomide |
| Transporter | P08183 | topotecan |
| Structural | P04350 | vinblastine |
| Transporter | P08183 | vinblastine |
| Structural | P04350 | vincristine |
| Transporter | P08183 | vincristine |
| Structural | P04350 | vinorelbine |
| Structural | P04350 | vinflunine |
| Kinase | O14976 | pazopanib |
| Kinase | P19525 | pazopanib |
| Kinase | O94804 | pazopanib |
| Kinase | Q8TBX8 | pazopanib |
| Kinase | Q9H2G2 | pazopanib |
| Structural | P04350 | cabazitaxel |
| Enzyme | Q00688 | temsirolimus |
| Transporter | P08183 | temsirolimus |
| Enzyme | P62942 | temsirolimus |
| Structural | P04350 | eribulin |
| Kinase | O14976 | vandetanib |
| Kinase | O94804 | vandetanib |
| Kinase | P41240 | vandetanib |
| Kinase | Q9H2G2 | vandetanib |
| Kinase | Q9Y5S2 | vandetanib |
| Kinase | O15075 | crizotinib |
| Kinase | P19525 | crizotinib |
| Kinase | O94804 | crizotinib |
| Kinase | Q9H2G2 | crizotinib |
| Kinase | Q13043 | crizotinib |
| Kinase | Q8TD19 | crizotinib |
| Kinase | Q05397 | crizotinib |
| Kinase | O14936 | crizotinib |
| Kinase | O15075 | ruxolitinib |
| Kinase | O14976 | ruxolitinib |
| Kinase | O95747 | ruxolitinib |
| Kinase | Q7KZI7 | ruxolitinib |
| Kinase | P19784 | ruxolitinib |
| Kinase | Q15746 | ruxolitinib |

| | | |
|---|---|---|
| Kinase | O14936 | ruxolitinib |
| Kinase | Q86Y07 | ruxolitinib |
| Kinase | Q13557 | ruxolitinib |
| Kinase | O14976 | axitinib |
| Kinase | P19525 | axitinib |
| Kinase | O94804 | axitinib |
| Kinase | P19784 | axitinib |
| Kinase | Q16513 | axitinib |
| Kinase | P41240 | axitinib |
| Kinase | Q9H2G2 | axitinib |
| Kinase | Q13043 | axitinib |
| Kinase | Q9Y5S2 | axitinib |
| Kinase | O14976 | afatinib |
| Kinase | O94804 | afatinib |
| Kinase | Q9H2G2 | afatinib |
| Kinase | Q16539 | afatinib |
| Kinase | O14976 | bosutinib |
| Kinase | O94804 | bosutinib |
| Kinase | Q16513 | bosutinib |
| Kinase | P41240 | bosutinib |
| Kinase | Q15746 | bosutinib |
| Kinase | Q9H2G2 | bosutinib |
| Kinase | Q13153 | bosutinib |
| Kinase | Q13043 | bosutinib |
| Kinase | Q05397 | bosutinib |
| Kinase | O14936 | bosutinib |
| Kinase | Q9Y6E0 | bosutinib |
| Kinase | Q13177 | bosutinib |
| Kinase | Q86Y07 | bosutinib |
| Enzyme | P28074 | carfilzomib |
| Enzyme | P07858 | carfilzomib |
| Enzyme | P34913 | regorafenib |
| Kinase | O15075 | tofacitinib |
| Kinase | Q16513 | tofacitinib |
| Kinase | Q13557 | tofacitinib |
| Kinase | P41240 | ponatinib |
| Kinase | O15075 | nintedanib |
| Kinase | O95747 | nintedanib |
| Kinase | Q96BR1 | nintedanib |
| Kinase | O94804 | nintedanib |
| Kinase | Q7KZI7 | nintedanib |
| Kinase | P19784 | nintedanib |
| Kinase | Q16513 | nintedanib |
| Kinase | Q15746 | nintedanib |

| | | |
|---|---|---|
| Kinase | Q9H2G2 | nintedanib |
| Kinase | Q13043 | nintedanib |
| Kinase | Q05397 | nintedanib |
| Kinase | Q13523 | nintedanib |
| Kinase | Q9Y6E0 | nintedanib |
| Kinase | Q13177 | nintedanib |
| Kinase | P78356 | nintedanib |
| Kinase | P78362 | nintedanib |
| Adhesion | P20701 | efalizumab |
| Structural | P04350 | brentuximab vedotin |
| Structural | P04350 | trastuzumab emtansine |
| Enzyme | P28074 | ixazomib |
| Enzyme | P49721 | ixazomib |
| Enzyme | P20618 | ixazomib |
| Enzyme | Q53GL7 | rucaparib |
| Enzyme | Q460N5 | rucaparib |
| Nuclear hormone receptor | P37231 | bexarotene |
| Nuclear hormone receptor | P10275 | bicalutamide |
| Kinase | P36888 | dasatinib |
| Kinase | P35968 | dasatinib |
| Kinase | P22607 | dasatinib |
| Kinase | P16234 | dasatinib |
| Kinase | O60674 | dasatinib |
| Kinase | P52333 | dasatinib |
| Kinase | P09619 | dasatinib |
| Kinase | P21802 | dasatinib |
| Kinase | P10721 | dasatinib |
| Kinase | Q15303 | dasatinib |
| Kinase | P42336 | dasatinib |
| Kinase | P37173 | dasatinib |
| Kinase | P36888 | erlotinib |
| Kinase | P35968 | erlotinib |
| Kinase | P16234 | erlotinib |
| Kinase | P08581 | erlotinib |
| Kinase | O60674 | erlotinib |
| Kinase | P52333 | erlotinib |
| Kinase | P09619 | erlotinib |
| Kinase | P21802 | erlotinib |
| Kinase | P10721 | erlotinib |
| Kinase | Q15303 | erlotinib |
| Nuclear hormone receptor | P10275 | exemestane |
| Nuclear hormone receptor | P10275 | flutamide |
| Nuclear hormone receptor | P03372 | fulvestrant |
| Kinase | P36888 | gefitinib |

| | | |
|---|---|---|
| Kinase | P35968 | gefitinib |
| Kinase | P08581 | gefitinib |
| Kinase | P10721 | gefitinib |
| Kinase | Q15303 | gefitinib |
| Kinase | O96017 | gefitinib |
| Kinase | P21802 | imatinib |
| Kinase | O60674 | imatinib |
| Kinase | P36888 | imatinib |
| Kinase | P35968 | imatinib |
| Kinase | P16234 | imatinib |
| Kinase | P09619 | imatinib |
| Kinase | P10721 | imatinib |
| Kinase | P09619 | imatinib |
| Kinase | P09619 | lapatinib |
| Kinase | Q15303 | lapatinib |
| Nuclear hormone receptor | P10275 | masoprocol |
| Nuclear hormone receptor | P03372 | masoprocol |
| Kinase | P11309 | mitoxantrone |
| Kinase | P21802 | nilotinib |
| Kinase | O60674 | nilotinib |
| Kinase | P36888 | nilotinib |
| Kinase | P16234 | nilotinib |
| Kinase | P09619 | nilotinib |
| Kinase | P10721 | nilotinib |
| Nuclear hormone receptor | P10275 | nilutamide |
| Kinase | P36888 | sorafenib |
| Kinase | P35968 | sorafenib |
| Kinase | P22607 | sorafenib |
| Kinase | P16234 | sorafenib |
| Kinase | P52333 | sorafenib |
| Kinase | P09619 | sorafenib |
| Kinase | P21802 | sorafenib |
| Kinase | P10721 | sorafenib |
| Kinase | P37173 | sorafenib |
| Kinase | P36888 | sunitinib |
| Kinase | P35968 | sunitinib |
| Kinase | P22607 | sunitinib |
| Kinase | P22455 | sunitinib |
| Kinase | P08581 | sunitinib |
| Kinase | O60674 | sunitinib |
| Kinase | P23458 | sunitinib |
| Kinase | P52333 | sunitinib |
| Kinase | P09619 | sunitinib |
| Kinase | P21802 | sunitinib |

| | | |
|---|---|---|
| Kinase | P10721 | sunitinib |
| Kinase | P42336 | sunitinib |
| Kinase | P45985 | sunitinib |
| Kinase | P11802 | sunitinib |
| Kinase | O96017 | sunitinib |
| Kinase | P16234 | sunitinib |
| Nuclear hormone receptor | P03372 | tamoxifen |
| Nuclear hormone receptor | P03372 | toremifene |
| Nuclear hormone receptor | P03372 | polyestradiol phosphate |
| Kinase | P36888 | pazopanib |
| Kinase | P35968 | pazopanib |
| Kinase | P22607 | pazopanib |
| Kinase | P22455 | pazopanib |
| Kinase | P16234 | pazopanib |
| Kinase | P08581 | pazopanib |
| Kinase | O60674 | pazopanib |
| Kinase | P52333 | pazopanib |
| Kinase | P09619 | pazopanib |
| Kinase | P21802 | pazopanib |
| Kinase | P10721 | pazopanib |
| Kinase | P37173 | pazopanib |
| Kinase | P45985 | pazopanib |
| Kinase | P36888 | vandetanib |
| Kinase | P35968 | vandetanib |
| Kinase | P22607 | vandetanib |
| Kinase | P22455 | vandetanib |
| Kinase | P16234 | vandetanib |
| Kinase | P08581 | vandetanib |
| Kinase | P09619 | vandetanib |
| Kinase | P21802 | vandetanib |
| Kinase | P10721 | vandetanib |
| Kinase | Q15303 | vandetanib |
| Kinase | P36888 | crizotinib |
| Kinase | P22607 | crizotinib |
| Kinase | P08581 | crizotinib |
| Kinase | O60674 | crizotinib |
| Kinase | P23458 | crizotinib |
| Kinase | P52333 | crizotinib |
| Kinase | P10721 | crizotinib |
| Kinase | O60674 | ruxolitinib |
| Kinase | O60674 | ruxolitinib |
| Kinase | O60674 | ruxolitinib |
| Kinase | P23458 | ruxolitinib |
| Kinase | P23458 | ruxolitinib |

| | | |
|---|---|---|
| Kinase | P23458 | ruxolitinib |
| Kinase | O60674 | ruxolitinib |
| Kinase | P52333 | ruxolitinib |
| Kinase | P10721 | ruxolitinib |
| Kinase | P45985 | ruxolitinib |
| Kinase | P36888 | axitinib |
| Kinase | P35968 | axitinib |
| Kinase | P22607 | axitinib |
| Kinase | P16234 | axitinib |
| Kinase | P08581 | axitinib |
| Kinase | O60674 | axitinib |
| Kinase | P52333 | axitinib |
| Kinase | P09619 | axitinib |
| Kinase | P21802 | axitinib |
| Kinase | P10721 | axitinib |
| GPCR | Q99835 | vismodegib |
| Kinase | P36888 | afatinib |
| Kinase | P08581 | afatinib |
| Kinase | Q15303 | afatinib |
| Kinase | P36888 | bosutinib |
| Kinase | P16234 | bosutinib |
| Kinase | P08581 | bosutinib |
| Kinase | O60674 | bosutinib |
| Kinase | P52333 | bosutinib |
| Kinase | P09619 | bosutinib |
| Kinase | P10721 | bosutinib |
| Kinase | Q15303 | bosutinib |
| Kinase | P45985 | bosutinib |
| Kinase | O96017 | bosutinib |
| Nuclear hormone receptor | P10275 | enzalutamide |
| Kinase | P35968 | regorafenib |
| Kinase | P52333 | tofacitinib |
| Kinase | O60674 | tofacitinib |
| Kinase | O60674 | tofacitinib |
| Kinase | O60674 | tofacitinib |
| Kinase | P23458 | tofacitinib |
| Kinase | P23458 | tofacitinib |
| Kinase | O60674 | tofacitinib |
| Kinase | P23458 | tofacitinib |
| Kinase | P35968 | cabozantinib |
| Kinase | P08581 | cabozantinib |
| Kinase | P36888 | cabozantinib |
| Kinase | P10721 | cabozantinib |
| Kinase | P10721 | ponatinib |

| | | |
|---|---|---|
| Kinase | P35968 | ponatinib |
| Kinase | P16234 | ponatinib |
| Kinase | P36888 | ponatinib |
| Kinase | P21802 | ibrutinib |
| Kinase | Q15303 | ibrutinib |
| Kinase | P52333 | ibrutinib |
| Kinase | P36888 | ceritinib |
| Kinase | P35968 | ceritinib |
| Kinase | P22607 | ceritinib |
| Kinase | P22455 | ceritinib |
| Kinase | P16234 | ceritinib |
| Kinase | P08581 | ceritinib |
| Kinase | O60674 | ceritinib |
| Kinase | P23458 | ceritinib |
| Kinase | P52333 | ceritinib |
| Kinase | P21802 | ceritinib |
| Kinase | P10721 | ceritinib |
| Kinase | P11802 | ceritinib |
| Kinase | P42336 | idelalisib |
| Kinase | P35968 | nintedanib |
| Kinase | P16234 | nintedanib |
| Kinase | P36888 | nintedanib |
| Kinase | P22607 | nintedanib |
| Kinase | P22455 | nintedanib |
| Kinase | P08581 | nintedanib |
| Kinase | O60674 | nintedanib |
| Kinase | P23458 | nintedanib |
| Kinase | P52333 | nintedanib |
| Kinase | P09619 | nintedanib |
| Kinase | P21802 | nintedanib |
| Kinase | P10721 | nintedanib |
| Kinase | P45985 | nintedanib |
| Kinase | P11802 | nintedanib |
| Kinase | P35968 | ramucirumab |
| Kinase | P35968 | alectinib |
| Kinase | P11802 | palbociclib |
| Kinase | Q00534 | palbociclib |
| Kinase | P36888 | palbociclib |
| Kinase | P52333 | palbociclib |
| Kinase | P11802 | palbociclib |
| Kinase | P24385 | palbociclib |
| Kinase | P35968 | lenvatinib |
| Kinase | P16234 | lenvatinib |
| Kinase | P09619 | lenvatinib |

| | | |
|---|---|---|
| Kinase | P10721 | lenvatinib |
| Membrane receptor | Q99062 | pegfilgrastim |
| Membrane receptor | Q99062 | filgrastim |
| Membrane receptor | Q99062 | lipegfilgrastim |
| GPCR | Q99835 | sonidegib |
| Cytosolic other | P10415 | venetoclax |
| CD molecules | Q9NZQ7 | atezolizumab |
| Membrane receptor | Q99062 | lenograstim |
| Kinase | P16234 | olaratumab |
| Kinase | P23458 | baricitinib |
| Kinase | O60674 | baricitinib |
| Kinase | P52333 | baricitinib |
| Kinase | P11802 | ribociclib |
| Kinase | Q00534 | ribociclib |
| CD molecules | Q9NZQ7 | avelumab |
| Kinase | P36894 | dasatinib |
| Transcription factor | Q16665 | topotecan |
| Enzyme | Q9Y6K1 | azacitidine |
| Nuclear hormone receptor | P10276 | bexarotene |
| Kinase | P00519 | dasatinib |
| Kinase | P43405 | dasatinib |
| Kinase | P04049 | dasatinib |
| Kinase | O43283 | dasatinib |
| Kinase | P15056 | dasatinib |
| Kinase | P11362 | dasatinib |
| Kinase | P21860 | dasatinib |
| Kinase | P07949 | dasatinib |
| Kinase | P00519 | dasatinib |
| Enzyme | Q9Y6K1 | decitabine |
| Kinase | P00519 | erlotinib |
| Kinase | P21860 | erlotinib |
| Kinase | P07949 | erlotinib |
| Kinase | Q9UM73 | erlotinib |
| Kinase | P00519 | gefitinib |
| Kinase | P21860 | gefitinib |
| Kinase | P07949 | gefitinib |
| Kinase | P11362 | imatinib |
| Kinase | P00519 | imatinib |
| Kinase | P43405 | imatinib |
| Kinase | P04049 | imatinib |
| Kinase | P15056 | imatinib |
| Kinase | P00519 | imatinib |
| Kinase | P00519 | imatinib |
| Kinase | P00519 | lapatinib |

| | | |
|---|---|---|
| Kinase | P21860 | lapatinib |
| Kinase | P11362 | nilotinib |
| Kinase | P00519 | nilotinib |
| Kinase | P04049 | nilotinib |
| Kinase | P15056 | nilotinib |
| Kinase | Q16288 | nilotinib |
| Kinase | P07949 | nilotinib |
| Kinase | P00519 | nilotinib |
| Kinase | P00519 | sorafenib |
| Kinase | P04049 | sorafenib |
| Kinase | P15056 | sorafenib |
| Kinase | P04629 | sorafenib |
| Kinase | P11362 | sorafenib |
| Kinase | Q16288 | sorafenib |
| Kinase | P07949 | sorafenib |
| Kinase | P07949 | sunitinib |
| Kinase | P00519 | sunitinib |
| Kinase | O43283 | sunitinib |
| Kinase | P04629 | sunitinib |
| Kinase | P11362 | sunitinib |
| Kinase | P31751 | sunitinib |
| Kinase | Q16288 | sunitinib |
| Kinase | Q15831 | sunitinib |
| Kinase | Q9UM73 | sunitinib |
| Kinase | P00519 | pazopanib |
| Kinase | P43405 | pazopanib |
| Kinase | P04049 | pazopanib |
| Kinase | P15056 | pazopanib |
| Kinase | P11362 | pazopanib |
| Kinase | P08922 | pazopanib |
| Kinase | Q13233 | pazopanib |
| Kinase | P07949 | pazopanib |
| Kinase | Q9UM73 | pazopanib |
| Kinase | P00519 | vandetanib |
| Kinase | P11362 | vandetanib |
| Kinase | P21860 | vandetanib |
| Kinase | P07949 | vandetanib |
| Kinase | Q9UM73 | vandetanib |
| Kinase | P00519 | crizotinib |
| Kinase | P43405 | crizotinib |
| Kinase | O43283 | crizotinib |
| Kinase | P04629 | crizotinib |
| Kinase | P06748 | crizotinib |
| Kinase | Q16288 | crizotinib |

| | | |
|---|---|---|
| Kinase | P08922 | crizotinib |
| Kinase | Q13233 | crizotinib |
| Kinase | P07949 | crizotinib |
| Kinase | Q9UM73 | crizotinib |
| Kinase | P00519 | ruxolitinib |
| Kinase | P15056 | ruxolitinib |
| Kinase | P04629 | ruxolitinib |
| Kinase | Q16288 | ruxolitinib |
| Kinase | Q13233 | ruxolitinib |
| Kinase | P07949 | ruxolitinib |
| Kinase | Q9UM73 | ruxolitinib |
| Kinase | P00519 | axitinib |
| Kinase | P04629 | axitinib |
| Kinase | P11362 | axitinib |
| Kinase | P08922 | axitinib |
| Kinase | P07949 | axitinib |
| Kinase | P00519 | afatinib |
| Kinase | P21860 | afatinib |
| Kinase | P00519 | bosutinib |
| Kinase | P43405 | bosutinib |
| Kinase | O43283 | bosutinib |
| Kinase | P04629 | bosutinib |
| Kinase | P06748 | bosutinib |
| Kinase | Q16288 | bosutinib |
| Kinase | P21860 | bosutinib |
| Kinase | Q13233 | bosutinib |
| Kinase | P00519 | bosutinib |
| Kinase | P07949 | bosutinib |
| Kinase | Q9UM73 | bosutinib |
| Kinase | P00519 | bosutinib |
| GPCR | P61073 | plerixafor |
| Kinase | P15056 | regorafenib |
| Kinase | P00519 | tofacitinib |
| Kinase | P07949 | tofacitinib |
| Kinase | P07949 | cabozantinib |
| Kinase | P00519 | ponatinib |
| Kinase | P11362 | ponatinib |
| Kinase | P00519 | ponatinib |
| Kinase | P07949 | ponatinib |
| Kinase | P15056 | dabrafenib |
| Kinase | P04049 | dabrafenib |
| Kinase | P07949 | ibrutinib |
| Kinase | Q9UM73 | ceritinib |
| Kinase | P00519 | ceritinib |

| | | |
|---|---|---|
| Kinase | P43405 | ceritinib |
| Kinase | P04629 | ceritinib |
| Kinase | P08922 | ceritinib |
| Kinase | P07949 | ceritinib |
| Kinase | P11362 | nintedanib |
| Kinase | P00519 | nintedanib |
| Kinase | O43283 | nintedanib |
| Kinase | P04629 | nintedanib |
| Kinase | Q16288 | nintedanib |
| Kinase | Q15831 | nintedanib |
| Kinase | P08922 | nintedanib |
| Kinase | P07949 | nintedanib |
| Kinase | Q9UM73 | nintedanib |
| Kinase | P07949 | alectinib |
| Kinase | Q9UM73 | alectinib |
| Kinase | P11362 | lenvatinib |
| Kinase | P07949 | lenvatinib |
| Kinase | P11274 | dasatinib |
| Kinase | Q13882 | dasatinib |
| Kinase | Q04771 | dasatinib |
| Kinase | P42684 | dasatinib |
| Kinase | P06239 | dasatinib |
| Kinase | P06239 | doxorubicin |
| Enzyme | O14746 | doxorubicin |
| Kinase | P42684 | erlotinib |
| Kinase | P35916 | erlotinib |
| Kinase | P06239 | erlotinib |
| Kinase | P42684 | gefitinib |
| Kinase | P06239 | gefitinib |
| Kinase | P42684 | imatinib |
| Kinase | P06239 | imatinib |
| Kinase | P42684 | nilotinib |
| Kinase | P06239 | nilotinib |
| Kinase | P42684 | sorafenib |
| Kinase | P35916 | sorafenib |
| Kinase | P06239 | sorafenib |
| Kinase | Q08881 | sunitinib |
| Kinase | Q13882 | sunitinib |
| Kinase | P42684 | sunitinib |
| Kinase | P35916 | sunitinib |
| Kinase | P06239 | sunitinib |
| Kinase | P06239 | tamoxifen |
| Kinase | Q13882 | pazopanib |
| Kinase | P42684 | pazopanib |

| | | |
|---|---|---|
| Kinase | P35916 | pazopanib |
| Kinase | P06239 | pazopanib |
| Kinase | Q08881 | pazopanib |
| Kinase | Q13882 | vandetanib |
| Kinase | Q04771 | vandetanib |
| Kinase | P42684 | vandetanib |
| Kinase | P35916 | vandetanib |
| Kinase | P06239 | vandetanib |
| Kinase | Q08881 | crizotinib |
| Kinase | O14920 | crizotinib |
| Kinase | Q04771 | crizotinib |
| Kinase | P42684 | crizotinib |
| Kinase | P06239 | crizotinib |
| Kinase | O14920 | ruxolitinib |
| Kinase | Q08881 | axitinib |
| Kinase | P42684 | axitinib |
| Kinase | P35916 | axitinib |
| Kinase | P06239 | axitinib |
| Kinase | P06239 | afatinib |
| Kinase | Q08881 | bosutinib |
| Kinase | P42684 | bosutinib |
| Kinase | P06239 | bosutinib |
| Kinase | P06239 | cisplatin |
| GPCR | P25106 | plerixafor |
| Kinase | P06239 | tofacitinib |
| Kinase | P11274 | ponatinib |
| Kinase | P06239 | ibrutinib |
| Kinase | Q08881 | ibrutinib |
| Transcription factor | P41212 | ceritinib |
| Kinase | P06239 | ceritinib |
| Kinase | P35916 | nintedanib |
| Kinase | Q08881 | nintedanib |
| Kinase | Q04771 | nintedanib |
| Kinase | P42684 | nintedanib |
| Kinase | P06239 | nintedanib |
| Kinase | P35916 | lenvatinib |
| Kinase | O14965 | dasatinib |
| Kinase | Q96GD4 | dasatinib |
| Kinase | P36896 | dasatinib |
| Kinase | O14965 | doxorubicin |
| Kinase | O14965 | erlotinib |
| Kinase | Q96GD4 | erlotinib |
| Kinase | Q02763 | nilotinib |
| Kinase | Q16620 | nilotinib |

| | | |
|---|---|---|
| Kinase | O14965 | sorafenib |
| Kinase | Q96GD4 | sorafenib |
| Kinase | Q02763 | sorafenib |
| Kinase | Q16620 | sorafenib |
| Kinase | O14965 | sunitinib |
| Kinase | Q96GD4 | sunitinib |
| Kinase | Q16620 | sunitinib |
| Kinase | O14965 | pazopanib |
| Kinase | Q02763 | pazopanib |
| Kinase | Q02763 | vandetanib |
| Kinase | O14965 | crizotinib |
| Kinase | Q96GD4 | crizotinib |
| Kinase | Q02763 | crizotinib |
| Kinase | P36896 | crizotinib |
| Kinase | Q16620 | crizotinib |
| Kinase | Q16620 | ruxolitinib |
| Kinase | O14965 | axitinib |
| Kinase | Q96GD4 | axitinib |
| Kinase | Q02763 | axitinib |
| Kinase | Q02763 | bosutinib |
| Kinase | Q16620 | bosutinib |
| Kinase | Q02763 | cabozantinib |
| Kinase | Q16620 | ceritinib |
| Kinase | Q96GD4 | nintedanib |
| Kinase | Q02763 | nintedanib |
| Kinase | P36896 | nintedanib |
| Kinase | Q16620 | nintedanib |
| Kinase | P49841 | nintedanib |
| Enzyme | Q9UGN5 | olaparib |
| Secreted | P15692 | aflibercept |
| Membrane receptor | Q15116 | nivolumab |
| Membrane receptor | Q15116 | pembrolizumab |
| Secreted | P15692 | bevacizumab |
| Cytosolic other | Q07817 | venetoclax |
| Enzyme | Q9UGN5 | rucaparib |
| Enzyme | Q9UGN5 | niraparib |
| Enzyme | P04818 | fluorouracil |
| Enzyme | Q969S8 | bendamustine |
| Enzyme | Q9UBN7 | bendamustine |
| Enzyme | P04818 | capecitabine |
| Enzyme | P31350 | clofarabine |
| Kinase | P30291 | dasatinib |
| Kinase | Q99640 | dasatinib |
| Kinase | O00238 | dasatinib |

| | | |
|---|---|---|
| Kinase | P29597 | dasatinib |
| Kinase | O00329 | dasatinib |
| Kinase | P37023 | dasatinib |
| Kinase | P53779 | erlotinib |
| Kinase | P33981 | erlotinib |
| Kinase | P35590 | erlotinib |
| Kinase | P29597 | erlotinib |
| Enzyme | P04818 | floxuridine |
| Nuclear hormone receptor | P06401 | fulvestrant |
| Kinase | P53779 | gefitinib |
| Enzyme | P04818 | gemcitabine |
| Enzyme | P31350 | gemcitabine |
| Kinase | P53779 | imatinib |
| Kinase | Q14680 | imatinib |
| Kinase | P29597 | imatinib |
| Enzyme | P04818 | methotrexate |
| Enzyme | P30304 | miltefosine |
| Enzyme | P14902 | mitomycin |
| Kinase | P53779 | nilotinib |
| Kinase | P35590 | nilotinib |
| Enzyme | P04818 | pemetrexed |
| Enzyme | P04818 | raltitrexed |
| Kinase | P33981 | sorafenib |
| Kinase | P35590 | sorafenib |
| Kinase | P53779 | sunitinib |
| Kinase | Q13554 | sunitinib |
| Kinase | Q14680 | sunitinib |
| Kinase | P30291 | sunitinib |
| Kinase | P33981 | sunitinib |
| Kinase | O75582 | sunitinib |
| Kinase | Q13464 | sunitinib |
| Kinase | O00238 | sunitinib |
| Kinase | P29597 | sunitinib |
| Kinase | Q13873 | sunitinib |
| Kinase | O00329 | sunitinib |
| Enzyme | Q9H2K2 | sunitinib |
| Kinase | P54646 | sunitinib |
| Kinase | P35590 | sunitinib |
| Kinase | Q12852 | sunitinib |
| Kinase | P23443 | sunitinib |
| Enzyme | P35354 | tamoxifen |
| Nuclear hormone receptor | P06401 | tamoxifen |
| Cytokine | P01375 | thalidomide |
| Enzyme | P35354 | thalidomide |

| | | |
|---|---|---|
| Cytosolic other | Q9HC29 | mifamurtide |
| Cytokine | P01375 | lenalidomide |
| Kinase | P53779 | pazopanib |
| Kinase | P33981 | pazopanib |
| Kinase | O00238 | pazopanib |
| Kinase | P29597 | pazopanib |
| Kinase | P35590 | pazopanib |
| Enzyme | Q9UBN7 | romidepsin |
| Enzyme | Q9UQL6 | romidepsin |
| Enzyme | Q969S8 | vorinostat |
| Enzyme | Q96DB2 | vorinostat |
| Enzyme | Q9UBN7 | vorinostat |
| Enzyme | Q9UQL6 | vorinostat |
| GPCR | P21453 | fingolimod |
| Kinase | O00238 | vandetanib |
| Kinase | P37023 | vandetanib |
| Kinase | P35590 | vandetanib |
| Kinase | P23443 | vandetanib |
| Kinase | Q13464 | crizotinib |
| Kinase | O00238 | crizotinib |
| Kinase | P29597 | crizotinib |
| Kinase | P35590 | crizotinib |
| Kinase | Q12852 | crizotinib |
| Kinase | P23443 | crizotinib |
| Kinase | Q13554 | ruxolitinib |
| Kinase | Q14680 | ruxolitinib |
| Kinase | P33981 | ruxolitinib |
| Kinase | O75582 | ruxolitinib |
| Kinase | Q13464 | ruxolitinib |
| Kinase | O00238 | ruxolitinib |
| Kinase | P29597 | ruxolitinib |
| Kinase | Q13873 | ruxolitinib |
| Kinase | P53779 | axitinib |
| Kinase | P33981 | axitinib |
| Kinase | O00238 | axitinib |
| Kinase | P29597 | axitinib |
| Kinase | P35590 | axitinib |
| Kinase | Q12852 | axitinib |
| Kinase | P53779 | afatinib |
| Kinase | P30291 | bosutinib |
| Kinase | Q13464 | bosutinib |
| Kinase | Q99640 | bosutinib |
| Kinase | P29597 | bosutinib |
| Kinase | Q13873 | bosutinib |

| | | |
|---|---|---|
| Kinase | P35590 | bosutinib |
| Kinase | Q12852 | bosutinib |
| Kinase | P23443 | bosutinib |
| GPCR | P41597 | cisplatin |
| Cytokine | P48061 | plerixafor |
| GPCR | P41597 | plerixafor |
| Enzyme | Q969S8 | panobinostat |
| Enzyme | Q96DB2 | panobinostat |
| Enzyme | Q9UBN7 | panobinostat |
| Enzyme | Q9UQL6 | panobinostat |
| Kinase | Q13464 | tofacitinib |
| Kinase | P29597 | tofacitinib |
| Enzyme | Q969S8 | belinostat |
| Enzyme | Q96DB2 | belinostat |
| Enzyme | Q9UBN7 | belinostat |
| Enzyme | Q9UQL6 | belinostat |
| Kinase | O00329 | idelalisib |
| Kinase | P53779 | nintedanib |
| Kinase | Q14680 | nintedanib |
| Kinase | P30291 | nintedanib |
| Kinase | P33981 | nintedanib |
| Kinase | O75582 | nintedanib |
| Kinase | P29597 | nintedanib |
| Kinase | Q13873 | nintedanib |
| Kinase | P54646 | nintedanib |
| Kinase | P37023 | nintedanib |
| Kinase | P35590 | nintedanib |
| Kinase | Q12852 | nintedanib |
| Kinase | P23443 | nintedanib |
| Cytokine | P01375 | adalimumab |
| Enzyme | O95271 | olaparib |
| Surface antigen | P15391 | blinatumomab |
| Membrane receptor | P08887 | tocilizumab |
| Membrane receptor | P01589 | basiliximab |
| Kinase | P20248 | palbociclib |
| Cytokine | P01584 | canakinumab |
| Membrane receptor | P01589 | daclizumab |
| Cytokine | P01375 | golimumab |
| Cytokine | P01375 | certolizumab pegol |
| Cytokine | P01375 | infliximab |
| Cytokine | P05231 | siltuximab |
| Cytokine | P01375 | etanercept |
| Membrane receptor | P01589 | aldesleukin |
| Membrane receptor | P40189 | oprelvekin |

| | | |
|---|---|---|
| Membrane receptor | P01589 | denileukin diftitox |
| Cytokine | P01584 | rilonacept |
| Kinase | P29597 | baricitinib |
| Enzyme | O95271 | rucaparib |
| Enzyme | Q9H2K2 | rucaparib |
| Kinase | P17948 | dasatinib |
| Kinase | P12931 | dasatinib |
| Kinase | P54760 | dasatinib |
| Kinase | Q06187 | dasatinib |
| Kinase | P29320 | dasatinib |
| Kinase | P54762 | dasatinib |
| Kinase | P07333 | dasatinib |
| Kinase | Q08345 | dasatinib |
| Kinase | P17948 | erlotinib |
| Kinase | P12931 | erlotinib |
| Kinase | P54760 | erlotinib |
| Kinase | Q9BUB5 | erlotinib |
| Kinase | P29320 | erlotinib |
| Kinase | Q08345 | erlotinib |
| Kinase | Q06418 | erlotinib |
| Kinase | P29376 | erlotinib |
| Kinase | P30530 | erlotinib |
| Kinase | Q12866 | erlotinib |
| Kinase | P54762 | erlotinib |
| Kinase | P12931 | gefitinib |
| Kinase | P54760 | gefitinib |
| Kinase | Q9BUB5 | gefitinib |
| Kinase | P29320 | gefitinib |
| Kinase | P54762 | gefitinib |
| Kinase | P29376 | gefitinib |
| Kinase | P30530 | gefitinib |
| Kinase | P54762 | imatinib |
| Kinase | P54760 | imatinib |
| Kinase | P12931 | imatinib |
| Kinase | P07333 | imatinib |
| Kinase | Q08345 | imatinib |
| Kinase | O75747 | lapatinib |
| Kinase | O00750 | lapatinib |
| Kinase | Q06187 | mitoxantrone |
| Kinase | P12931 | nilotinib |
| Kinase | P54760 | nilotinib |
| Kinase | P29320 | nilotinib |
| Kinase | P54762 | nilotinib |
| Kinase | P07333 | nilotinib |

| | | |
|---|---|---|
| Kinase | Q08345 | nilotinib |
| Kinase | P49336 | sorafenib |
| Kinase | P17948 | sorafenib |
| Kinase | P12931 | sorafenib |
| Kinase | P54760 | sorafenib |
| Kinase | Q9BUB5 | sorafenib |
| Kinase | P29320 | sorafenib |
| Kinase | P54762 | sorafenib |
| Kinase | P07333 | sorafenib |
| Kinase | Q08345 | sorafenib |
| Kinase | P30530 | sorafenib |
| Kinase | Q12866 | sorafenib |
| Kinase | P12931 | sunitinib |
| Kinase | Q06187 | sunitinib |
| Kinase | Q9BUB5 | sunitinib |
| Kinase | P29320 | sunitinib |
| Kinase | P54762 | sunitinib |
| Kinase | Q14164 | sunitinib |
| Kinase | P07333 | sunitinib |
| Kinase | Q08345 | sunitinib |
| Kinase | Q06418 | sunitinib |
| Kinase | P29376 | sunitinib |
| Kinase | P30530 | sunitinib |
| Kinase | O14757 | sunitinib |
| Kinase | Q12866 | sunitinib |
| Kinase | P54760 | sunitinib |
| Kinase | P17948 | sunitinib |
| Kinase | P17948 | pazopanib |
| Kinase | P12931 | pazopanib |
| Kinase | P07333 | pazopanib |
| Kinase | Q08345 | pazopanib |
| Kinase | P29376 | pazopanib |
| Kinase | Q12866 | pazopanib |
| Kinase | P17948 | vandetanib |
| Kinase | P12931 | vandetanib |
| Kinase | P54760 | vandetanib |
| Kinase | Q06187 | vandetanib |
| Kinase | Q9BUB5 | vandetanib |
| Kinase | P29320 | vandetanib |
| Kinase | Q04912 | vandetanib |
| Kinase | P54762 | vandetanib |
| Kinase | P07333 | vandetanib |
| Kinase | Q08345 | vandetanib |
| Kinase | Q06418 | vandetanib |

| | | |
|---|---|---|
| Kinase | P29376 | vandetanib |
| Kinase | P30530 | vandetanib |
| Kinase | Q12866 | vandetanib |
| Kinase | P17948 | crizotinib |
| Kinase | P12931 | crizotinib |
| Kinase | P54760 | crizotinib |
| Kinase | Q06187 | crizotinib |
| Kinase | P29320 | crizotinib |
| Kinase | Q04912 | crizotinib |
| Kinase | P54762 | crizotinib |
| Kinase | Q14164 | crizotinib |
| Kinase | P07333 | crizotinib |
| Kinase | Q08345 | crizotinib |
| Kinase | Q06418 | crizotinib |
| Kinase | P29376 | crizotinib |
| Kinase | P30530 | crizotinib |
| Kinase | Q12866 | crizotinib |
| Kinase | P29320 | ruxolitinib |
| Kinase | Q14164 | ruxolitinib |
| Kinase | P29376 | ruxolitinib |
| Kinase | P17948 | axitinib |
| Kinase | P07333 | axitinib |
| Kinase | Q08345 | axitinib |
| Kinase | P30530 | axitinib |
| Kinase | Q12866 | axitinib |
| Kinase | P12931 | afatinib |
| Kinase | Q9BUB5 | afatinib |
| Kinase | P30530 | afatinib |
| Kinase | P12931 | bosutinib |
| Kinase | P54760 | bosutinib |
| Kinase | Q06187 | bosutinib |
| Kinase | P29320 | bosutinib |
| Kinase | Q04912 | bosutinib |
| Kinase | P54762 | bosutinib |
| Kinase | Q14164 | bosutinib |
| Kinase | P07333 | bosutinib |
| Kinase | Q08345 | bosutinib |
| Kinase | Q06418 | bosutinib |
| Kinase | P29376 | bosutinib |
| Kinase | P30530 | bosutinib |
| Kinase | O14757 | bosutinib |
| Kinase | Q12866 | bosutinib |
| Kinase | P12931 | trabectedin |
| Kinase | Q04912 | cabozantinib |

| | | |
|---|---|---|
| Kinase | P12931 | ponatinib |
| Kinase | P49336 | ponatinib |
| Cytosolic other | Q14145 | dimethyl fumarate |
| Kinase | P10398 | dabrafenib |
| Kinase | Q06187 | ibrutinib |
| Kinase | P12931 | ibrutinib |
| Kinase | P12931 | ceritinib |
| Kinase | Q06187 | ceritinib |
| Kinase | P42338 | idelalisib |
| Kinase | P17948 | nintedanib |
| Kinase | P12931 | nintedanib |
| Kinase | P54760 | nintedanib |
| Kinase | Q06187 | nintedanib |
| Kinase | P29320 | nintedanib |
| Kinase | P54762 | nintedanib |
| Kinase | Q14164 | nintedanib |
| Kinase | P07333 | nintedanib |
| Kinase | Q08345 | nintedanib |
| Kinase | Q06418 | nintedanib |
| Kinase | P29376 | nintedanib |
| Kinase | P30530 | nintedanib |
| Kinase | O14757 | nintedanib |
| Kinase | Q12866 | nintedanib |
| Enzyme | Q9Y6F1 | olaparib |
| Kinase | P17948 | lenvatinib |
| Cytosolic other | Q92843 | venetoclax |
| Enzyme | Q9Y6F1 | rucaparib |

| Drug family | ATC category | Druggability | Present in Clonal proteome | Present in TCGA |
|---|---|---|---|---|
| Protein kinase inhibitors | Antineoplastic agents | TCLIN | | |
| Protein kinase inhibitors | Antineoplastic agents | TCLIN | | |
| Protein kinase inhibitors | Antineoplastic agents | TCLIN | | |
| Taxanes | Antineoplastic agents | TCLIN | | |
| Anthracyclines and related substances | Antineoplastic agents | TCLIN | | |
| Protein kinase inhibitors | Antineoplastic agents | TCLIN | | |
| Protein kinase inhibitors | Antineoplastic agents | TCLIN | | |
| Selective immunosuppressants | Immunosuppressants | TCLIN | | |
| Protein kinase inhibitors | Antineoplastic agents | TCLIN | | |
| Protein kinase inhibitors | Antineoplastic agents | TCLIN | | |
| Protein kinase inhibitors | Antineoplastic agents | TCLIN | | |
| Protein kinase inhibitors | Antineoplastic agents | TCLIN | | |
| Protein kinase inhibitors | Antineoplastic agents | TCLIN | | |
| Anthracyclines and related substances | Antineoplastic agents | TCLIN | | |
| Anthracyclines and related substances | Antineoplastic agents | TCLIN | | |
| Taxanes | Antineoplastic agents | TCLIN | | |
| Selective immunosuppressants | Immunosuppressants | TCLIN | | |
| Protein kinase inhibitors | Antineoplastic agents | TCLIN | | |
| Anti-estrogens | Endocrine therapy | TCLIN | | |
| Anti-estrogens | Endocrine therapy | TCLIN | | |
| Protein kinase inhibitors | Antineoplastic agents | TCLIN | | |
| Protein kinase inhibitors | Antineoplastic agents | TCLIN | | |
| Protein kinase inhibitors | Antineoplastic agents | TCLIN | | |
| Protein kinase inhibitors | Antineoplastic agents | TCLIN | | |
| Protein kinase inhibitors | Antineoplastic agents | TCLIN | | |
| Protein kinase inhibitors | Antineoplastic agents | TCLIN | | |
| Protein kinase inhibitors | Antineoplastic agents | TCLIN | | |
| Protein kinase inhibitors | Antineoplastic agents | TCLIN | | |
| Protein kinase inhibitors | Antineoplastic agents | TCLIN | | |
| Platinum compounds | Antineoplastic agents | TCLIN | | |
| Protein kinase inhibitors | Antineoplastic agents | TCLIN | | |
| Protein kinase inhibitors | Antineoplastic agents | TCLIN | | |
| Protein kinase inhibitors | Antineoplastic agents | TCLIN | | |
| Protein kinase inhibitors | Antineoplastic agents | TCLIN | | |
| Protein kinase inhibitors | Antineoplastic agents | TCLIN | | |

| | | |
|---|---|---|
| Monoclonal antibodies | Antineoplastic agents | TCLIN |
| Monoclonal antibodies | Antineoplastic agents | TCLIN |
| Monoclonal antibodies | Antineoplastic agents | TCLIN |
| Monoclonal antibodies | Antineoplastic agents | TCLIN |
| Monoclonal antibodies | Antineoplastic agents | TCLIN |
| Protein kinase inhibitors | Antineoplastic agents | TCLIN |
| Monoclonal antibodies | Antineoplastic agents | TCLIN |
| Protein kinase inhibitors | Antineoplastic agents | TCLIN |
| Protein kinase inhibitors | Antineoplastic agents | TCLIN |
| Other immunosuppressants | Immunosuppressants | TBIO |
| Protein kinase inhibitors | Antineoplastic agents | TCLIN |
| Protein kinase inhibitors | Antineoplastic agents | TCLIN |
| Protein kinase inhibitors | Antineoplastic agents | TCLIN |
| Other antineoplastic agents | Antineoplastic agents | TCHEM |
| Protein kinase inhibitors | Antineoplastic agents | TCLIN |
| Protein kinase inhibitors | Antineoplastic agents | TCLIN |
| Protein kinase inhibitors | Antineoplastic agents | TCLIN |
| Protein kinase inhibitors | Antineoplastic agents | TCLIN |
| Protein kinase inhibitors | Antineoplastic agents | TCLIN |
| Protein kinase inhibitors | Antineoplastic agents | TCLIN |
| Protein kinase inhibitors | Antineoplastic agents | TCLIN |
| Protein kinase inhibitors | Antineoplastic agents | TCLIN |
| Protein kinase inhibitors | Antineoplastic agents | TCLIN |
| Protein kinase inhibitors | Antineoplastic agents | TCLIN |
| Protein kinase inhibitors | Antineoplastic agents | TCLIN |
| Protein kinase inhibitors | Antineoplastic agents | TCLIN |
| Protein kinase inhibitors | Antineoplastic agents | TCLIN |
| Protein kinase inhibitors | Antineoplastic agents | TCLIN |
| Protein kinase inhibitors | Antineoplastic agents | TCLIN |
| Protein kinase inhibitors | Antineoplastic agents | TCLIN |
| Other antineoplastic agents | Antineoplastic agents | TCLIN |
| Anthracyclines and related substances | Antineoplastic agents | TCLIN |
| Other antineoplastic agents | Antineoplastic agents | TCLIN |
| Other antineoplastic agents | Antineoplastic agents | TCLIN |
| Nitrogen mustard analogues | Antineoplastic agents | TCLIN |
| Other antineoplastic agents | Antineoplastic agents | TCLIN |
| Protein kinase inhibitors | Antineoplastic agents | TCHEM |
| Protein kinase inhibitors | Antineoplastic agents | TCLIN |
| Other antineoplastic agents | Antineoplastic agents | TCLIN |
| Other antineoplastic agents | Antineoplastic agents | TCLIN |
| Other antineoplastic agents | Antineoplastic agents | TCLIN |
| Other antineoplastic agents | Antineoplastic agents | TCLIN |
| Other antineoplastic agents | Antineoplastic agents | TCLIN |
| Other antineoplastic agents | Antineoplastic agents | TCLIN |

| | | |
|---|---|---|
| Protein kinase inhibitors | Antineoplastic agents | TCLIN |
| Other antineoplastic agents | Antineoplastic agents | TCLIN |
| Protein kinase inhibitors | Antineoplastic agents | TCLIN |
| Other antineoplastic agents | Antineoplastic agents | TCLIN |
| Protein kinase inhibitors | Antineoplastic agents | TCLIN |
| Other antineoplastic agents | Antineoplastic agents | TCLIN |
| Other antineoplastic agents | Antineoplastic agents | TCLIN |
| Other antineoplastic agents | Antineoplastic agents | TCLIN |
| Other antineoplastic agents | Antineoplastic agents | TCLIN |
| Other antineoplastic agents | Antineoplastic agents | TCHEM |
| Nitrogen mustard analogues | Antineoplastic agents | TCLIN |
| Alkyl sulfonates | Antineoplastic agents | TCHEM |
| Pyrimidine analogues | Antineoplastic agents | TCHEM |
| Other alkylating agents | Antineoplastic agents | TCHEM |
| Actinomycines | Antineoplastic agents | TCHEM |
| Combinations of antineoplastic agents | Antineoplastic agents | TCLIN |
| Anthracyclines and related substances | Antineoplastic agents | TCLIN |
| Podophyllotoxin derivatives | Antineoplastic agents | TCLIN |
| Anthracyclines and related substances | Antineoplastic agents | TCLIN |
| Protein kinase inhibitors | Antineoplastic agents | TCLIN |
| Anthracyclines and related substances | Antineoplastic agents | TCLIN |
| Protein kinase inhibitors | Antineoplastic agents | TCLIN |
| Protein kinase inhibitors | Antineoplastic agents | TCHEM |
| Protein kinase inhibitors | Antineoplastic agents | TCHEM |
| Protein kinase inhibitors | Antineoplastic agents | TCLIN |
| Podophyllotoxin derivatives | Antineoplastic agents | TCLIN |
| Anthracyclines and related substances | Antineoplastic agents | TCLIN |
| Other antineoplastic agents | Antineoplastic agents | TCLIN |
| Other antineoplastic agents | Antineoplastic agents | TCLIN |
| Protein kinase inhibitors | Antineoplastic agents | TCLIN |
| Protein kinase inhibitors | Antineoplastic agents | TCLIN |
| Protein kinase inhibitors | Antineoplastic agents | TCLIN |
| Protein kinase inhibitors | Antineoplastic agents | TCLIN |
| Platinum compounds | Antineoplastic agents | TCHEM |
| Other antineoplastic agents | Antineoplastic agents | TCLIN |
| Selective immunosuppressants | Immunosuppressants | TCLIN |
| Protein kinase inhibitors | Antineoplastic agents | TCLIN |
| Other antineoplastic agents | Antineoplastic agents | TCLIN |
| Other antineoplastic agents | Antineoplastic agents | TCHEM |
| Other antineoplastic agents | Antineoplastic agents | TCHEM |
| Interleukin inhibitors | Immunosuppressants | TCHEM |
| Other antineoplastic agents | Antineoplastic agents | |
| Protein kinase inhibitors | Antineoplastic agents | TCLIN |
| Protein kinase inhibitors | Antineoplastic agents | TCLIN |

| | | |
|---|---|---|
| Protein kinase inhibitors | Antineoplastic agents | TCLIN |
| Protein kinase inhibitors | Antineoplastic agents | TCLIN |
| Protein kinase inhibitors | Antineoplastic agents | TCLIN |
| Protein kinase inhibitors | Antineoplastic agents | TCLIN |
| Protein kinase inhibitors | Antineoplastic agents | TCLIN |
| Protein kinase inhibitors | Antineoplastic agents | TCLIN |
| Protein kinase inhibitors | Antineoplastic agents | TCLIN |
| Protein kinase inhibitors | Antineoplastic agents | TCLIN |
| Protein kinase inhibitors | Antineoplastic agents | TCLIN |
| Protein kinase inhibitors | Antineoplastic agents | TCLIN |
| Protein kinase inhibitors | Antineoplastic agents | TCLIN |
| Protein kinase inhibitors | Antineoplastic agents | TCLIN |
| Protein kinase inhibitors | Antineoplastic agents | TCLIN |
| Anthracyclines and related substances | Antineoplastic agents | TCHEM |
| Other antineoplastic agents | Antineoplastic agents | TCHEM |
| Other antineoplastic agents | Antineoplastic agents | TCHEM |
| Other antineoplastic agents | Antineoplastic agents | TCLIN |
| Other immunosuppressants | Immunosuppressants | TCHEM |
| Other antineoplastic agents | Antineoplastic agents | TCLIN |
| Other antineoplastic agents | Antineoplastic agents | TCLIN |
| Other antineoplastic agents | Antineoplastic agents | TCHEM |
| Other antineoplastic agents | Antineoplastic agents | TCHEM |
| Other antineoplastic agents | Antineoplastic agents | TCLIN |
| Other antineoplastic agents | Antineoplastic agents | TCLIN |
| Other antineoplastic agents | Antineoplastic agents | TCLIN |
| Other antineoplastic agents | Antineoplastic agents | TCHEM |
| Other antineoplastic agents | Antineoplastic agents | TBIO |
| Nitrosoureas | Antineoplastic agents | TCLIN |
| Nitrogen mustard analogues | Antineoplastic agents | TCHEM |
| Purine analogues | Antineoplastic agents | TCLIN |
| Combinations of antineoplastic agents | Antineoplastic agents | TCHEM |
| Protein kinase inhibitors | Antineoplastic agents | TCHEM |
| Protein kinase inhibitors | Antineoplastic agents | TCHEM |
| Protein kinase inhibitors | Antineoplastic agents | TCHEM |
| Protein kinase inhibitors | Antineoplastic agents | TCHEM |
| Protein kinase inhibitors | Antineoplastic agents | TCHEM |
| Protein kinase inhibitors | Antineoplastic agents | TBIO |
| Protein kinase inhibitors | Antineoplastic agents | TBIO |
| Protein kinase inhibitors | Antineoplastic agents | TCHEM |
| Combinations of antineoplastic agents | Antineoplastic agents | TCHEM |
| Combinations of antineoplastic agents | Antineoplastic agents | TCHEM |
| Taxanes | Antineoplastic agents | TCLIN |
| Taxanes | Antineoplastic agents | TCHEM |
| Anthracyclines and related substances | Antineoplastic agents | TCHEM |

| | | |
|---|---|---|
| Anthracyclines and related substances | Antineoplastic agents | TCHEM |
| Anthracyclines and related substances | Antineoplastic agents | TCHEM |
| Protein kinase inhibitors | Antineoplastic agents | TCHEM |
| Protein kinase inhibitors | Antineoplastic agents | TBIO |
| Protein kinase inhibitors | Antineoplastic agents | TCHEM |
| Protein kinase inhibitors | Antineoplastic agents | TCHEM |
| Protein kinase inhibitors | Antineoplastic agents | TBIO |
| Protein kinase inhibitors | Antineoplastic agents | TCHEM |
| Protein kinase inhibitors | Antineoplastic agents | TCHEM |
| Podophyllotoxin derivatives | Antineoplastic agents | TCHEM |
| Selective immunosuppressants | Immunosuppressants | TCLIN |
| Selective immunosuppressants | Immunosuppressants | TCHEM |
| Anti-androgens | Endocrine therapy | TCHEM |
| Anti-estrogens | Endocrine therapy | TCHEM |
| Protein kinase inhibitors | Antineoplastic agents | TCHEM |
| Protein kinase inhibitors | Antineoplastic agents | TCHEM |
| Protein kinase inhibitors | Antineoplastic agents | TBIO |
| Protein kinase inhibitors | Antineoplastic agents | TCHEM |
| Pyrimidine analogues | Antineoplastic agents | TBIO |
| Pyrimidine analogues | Antineoplastic agents | TCLIN |
| Anthracyclines and related substances | Antineoplastic agents | TCHEM |
| Protein kinase inhibitors | Antineoplastic agents | TCLIN |
| Protein kinase inhibitors | Antineoplastic agents | TCHEM |
| Protein kinase inhibitors | Antineoplastic agents | TCHEM |
| Protein kinase inhibitors | Antineoplastic agents | TBIO |
| Protein kinase inhibitors | Antineoplastic agents | TCLIN |
| Protein kinase inhibitors | Antineoplastic agents | TCLIN |
| Protein kinase inhibitors | Antineoplastic agents | TCHEM |
| Other antineoplastic agents | Antineoplastic agents | TCHEM |
| Other cytotoxic antibiotics | Antineoplastic agents | TCLIN |
| Protein kinase inhibitors | Antineoplastic agents | TCHEM |
| Protein kinase inhibitors | Antineoplastic agents | TCHEM |
| Protein kinase inhibitors | Antineoplastic agents | TCHEM |
| Selective immunosuppressants | Immunosuppressants | TCHEM |
| Selective immunosuppressants | Immunosuppressants | TCLIN |
| Other antineoplastic agents | Antineoplastic agents | TCHEM |
| Other antineoplastic agents | Antineoplastic agents | TCLIN |
| Purine analogues | Antineoplastic agents | TCLIN |
| Purine analogues | Antineoplastic agents | TCLIN |
| Purine analogues | Antineoplastic agents | TCHEM |
| Purine analogues | Antineoplastic agents | TCLIN |
| Other immunosuppressants | Immunosuppressants | TCHEM |
| Anthracyclines and related substances | Antineoplastic agents | TCHEM |
| Anthracyclines and related substances | Antineoplastic agents | TCHEM |

| | | |
|---|---|---|
| Selective immunosuppressants | Immunosuppressants | TCLIN |
| Protein kinase inhibitors | Antineoplastic agents | TCLIN |
| Protein kinase inhibitors | Antineoplastic agents | TCHEM |
| Protein kinase inhibitors | Antineoplastic agents | TCHEM |
| Protein kinase inhibitors | Antineoplastic agents | TBIO |
| Protein kinase inhibitors | Antineoplastic agents | TBIO |
| Protein kinase inhibitors | Antineoplastic agents | TCLIN |
| Protein kinase inhibitors | Antineoplastic agents | TCLIN |
| Protein kinase inhibitors | Antineoplastic agents | TCHEM |
| Taxanes | Antineoplastic agents | TCLIN |
| Taxanes | Antineoplastic agents | TCLIN |
| Taxanes | Antineoplastic agents | TCHEM |
| Taxanes | Antineoplastic agents | TCLIN |
| Folic acid analogues | Antineoplastic agents | TBIO |
| Other antineoplastic agents | Antineoplastic agents | TCLIN |
| Selective immunosuppressants | Immunosuppressants | TCLIN |
| Selective immunosuppressants | Immunosuppressants | TCHEM |
| Selective immunosuppressants | Immunosuppressants | TBIO |
| Selective immunosuppressants | Immunosuppressants | TCHEM |
| Selective immunosuppressants | Immunosuppressants | TCHEM |
| Selective immunosuppressants | Immunosuppressants | TCHEM |
| Protein kinase inhibitors | Antineoplastic agents | TCHEM |
| Protein kinase inhibitors | Antineoplastic agents | TCHEM |
| Protein kinase inhibitors | Antineoplastic agents | TCHEM |
| Protein kinase inhibitors | Antineoplastic agents | TCHEM |
| Protein kinase inhibitors | Antineoplastic agents | TCHEM |
| Protein kinase inhibitors | Antineoplastic agents | TCHEM |
| Protein kinase inhibitors | Antineoplastic agents | TCHEM |
| Protein kinase inhibitors | Antineoplastic agents | TCHEM |
| Protein kinase inhibitors | Antineoplastic agents | TBIO |
| Protein kinase inhibitors | Antineoplastic agents | TCHEM |
| Protein kinase inhibitors | Antineoplastic agents | TBIO |
| Protein kinase inhibitors | Antineoplastic agents | TBIO |
| Protein kinase inhibitors | Antineoplastic agents | TCHEM |
| Protein kinase inhibitors | Antineoplastic agents | TCHEM |
| Protein kinase inhibitors | Antineoplastic agents | TCHEM |
| Protein kinase inhibitors | Antineoplastic agents | TCHEM |
| Protein kinase inhibitors | Antineoplastic agents | TCHEM |
| Protein kinase inhibitors | Antineoplastic agents | TCHEM |
| Protein kinase inhibitors | Antineoplastic agents | TCHEM |
| Protein kinase inhibitors | Antineoplastic agents | TCHEM |
| Protein kinase inhibitors | Antineoplastic agents | TCHEM |
| Protein kinase inhibitors | Antineoplastic agents | TCHEM |
| Protein kinase inhibitors | Antineoplastic agents | TCHEM |

| | | |
|---|---|---|
| Protein kinase inhibitors | Antineoplastic agents | TBIO |
| Protein kinase inhibitors | Antineoplastic agents | TCHEM |
| Anti-estrogens | Endocrine therapy | TCLIN |
| Anti-estrogens | Endocrine therapy | TCHEM |
| Anti-estrogens | Endocrine therapy | TBIO |
| Anti-estrogens | Endocrine therapy | TCHEM |
| Podophyllotoxin derivatives | Antineoplastic agents | TCHEM |
| Other immunosuppressants | Immunosuppressants | TCLIN |
| Other antineoplastic agents | Antineoplastic agents | TCHEM |
| Vinca alkaloids and analogues | Antineoplastic agents | TCLIN |
| Vinca alkaloids and analogues | Antineoplastic agents | TCHEM |
| Vinca alkaloids and analogues | Antineoplastic agents | TCLIN |
| Vinca alkaloids and analogues | Antineoplastic agents | TCHEM |
| Vinca alkaloids and analogues | Antineoplastic agents | TCLIN |
| Vinca alkaloids and analogues | Antineoplastic agents | TCLIN |
| Protein kinase inhibitors | Antineoplastic agents | TCHEM |
| Protein kinase inhibitors | Antineoplastic agents | TBIO |
| Protein kinase inhibitors | Antineoplastic agents | TCHEM |
| Protein kinase inhibitors | Antineoplastic agents | TBIO |
| Protein kinase inhibitors | Antineoplastic agents | TCHEM |
| Taxanes | Antineoplastic agents | TCLIN |
| Protein kinase inhibitors | Antineoplastic agents | TBIO |
| Protein kinase inhibitors | Antineoplastic agents | TCHEM |
| Protein kinase inhibitors | Antineoplastic agents | TCLIN |
| Other antineoplastic agents | Antineoplastic agents | TCLIN |
| Protein kinase inhibitors | Antineoplastic agents | TCHEM |
| Protein kinase inhibitors | Antineoplastic agents | TCHEM |
| Protein kinase inhibitors | Antineoplastic agents | TCHEM |
| Protein kinase inhibitors | Antineoplastic agents | TCHEM |
| Protein kinase inhibitors | Antineoplastic agents | TBIO |
| Protein kinase inhibitors | Antineoplastic agents | TCHEM |
| Protein kinase inhibitors | Antineoplastic agents | TBIO |
| Protein kinase inhibitors | Antineoplastic agents | TCHEM |
| Protein kinase inhibitors | Antineoplastic agents | TCHEM |
| Protein kinase inhibitors | Antineoplastic agents | TCHEM |
| Protein kinase inhibitors | Antineoplastic agents | TBIO |
| Protein kinase inhibitors | Antineoplastic agents | TCHEM |
| Protein kinase inhibitors | Antineoplastic agents | TBIO |
| Protein kinase inhibitors | Antineoplastic agents | TCHEM |
| Protein kinase inhibitors | Antineoplastic agents | TCHEM |
| Protein kinase inhibitors | Antineoplastic agents | TBIO |
| Protein kinase inhibitors | Antineoplastic agents | TCHEM |
| Protein kinase inhibitors | Antineoplastic agents | TCHEM |
| Protein kinase inhibitors | Antineoplastic agents | TCHEM |

| | | |
|---|---|---|
| Protein kinase inhibitors | Antineoplastic agents | TBIO |
| Protein kinase inhibitors | Antineoplastic agents | TBIO |
| Protein kinase inhibitors | Antineoplastic agents | TCHEM |
| Protein kinase inhibitors | Antineoplastic agents | TCHEM |
| Protein kinase inhibitors | Antineoplastic agents | TBIO |
| Protein kinase inhibitors | Antineoplastic agents | TCHEM |
| Protein kinase inhibitors | Antineoplastic agents | TCHEM |
| Protein kinase inhibitors | Antineoplastic agents | TCHEM |
| Protein kinase inhibitors | Antineoplastic agents | TCHEM |
| Protein kinase inhibitors | Antineoplastic agents | TCHEM |
| Protein kinase inhibitors | Antineoplastic agents | TCHEM |
| Protein kinase inhibitors | Antineoplastic agents | TBIO |
| Protein kinase inhibitors | Antineoplastic agents | TCHEM |
| Protein kinase inhibitors | Antineoplastic agents | TCHEM |
| Protein kinase inhibitors | Antineoplastic agents | TCHEM |
| Protein kinase inhibitors | Antineoplastic agents | TCHEM |
| Protein kinase inhibitors | Antineoplastic agents | TCHEM |
| Protein kinase inhibitors | Antineoplastic agents | TCHEM |
| Protein kinase inhibitors | Antineoplastic agents | TCHEM |
| Protein kinase inhibitors | Antineoplastic agents | TCHEM |
| Protein kinase inhibitors | Antineoplastic agents | TCHEM |
| Protein kinase inhibitors | Antineoplastic agents | TCHEM |
| Protein kinase inhibitors | Antineoplastic agents | TCHEM |
| Protein kinase inhibitors | Antineoplastic agents | TCHEM |
| Protein kinase inhibitors | Antineoplastic agents | TCHEM |
| Protein kinase inhibitors | Antineoplastic agents | TBIO |
| Protein kinase inhibitors | Antineoplastic agents | TCHEM |
| Protein kinase inhibitors | Antineoplastic agents | TCHEM |
| Protein kinase inhibitors | Antineoplastic agents | TBIO |
| Other antineoplastic agents | Antineoplastic agents | TCLIN |
| Other antineoplastic agents | Antineoplastic agents | TCHEM |
| Protein kinase inhibitors | Antineoplastic agents | TCHEM |
| Selective immunosuppressants | Immunosuppressants | TCHEM |
| Selective immunosuppressants | Immunosuppressants | TCHEM |
| Selective immunosuppressants | Immunosuppressants | TCHEM |
| Protein kinase inhibitors | Antineoplastic agents | TCHEM |
| Protein kinase inhibitors | Antineoplastic agents | TCHEM |
| Protein kinase inhibitors | Antineoplastic agents | TBIO |
| Protein kinase inhibitors | Antineoplastic agents | TBIO |
| Protein kinase inhibitors | Antineoplastic agents | TCHEM |
| Protein kinase inhibitors | Antineoplastic agents | TCHEM |
| Protein kinase inhibitors | Antineoplastic agents | TCHEM |
| Protein kinase inhibitors | Antineoplastic agents | TCHEM |
| Protein kinase inhibitors | Antineoplastic agents | TCHEM |

| | | |
|---|---|---|
| Protein kinase inhibitors | Antineoplastic agents | TCHEM |
| Protein kinase inhibitors | Antineoplastic agents | TCHEM |
| Protein kinase inhibitors | Antineoplastic agents | TCHEM |
| Protein kinase inhibitors | Antineoplastic agents | TCHEM |
| Protein kinase inhibitors | Antineoplastic agents | TCHEM |
| Protein kinase inhibitors | Antineoplastic agents | TCHEM |
| Protein kinase inhibitors | Antineoplastic agents | TCHEM |
| Protein kinase inhibitors | Antineoplastic agents | TBIO |
| Selective immunosuppressants | Immunosuppressants | TCLIN |
| Monoclonal antibodies | Antineoplastic agents | TCLIN |
| Monoclonal antibodies | Antineoplastic agents | TCLIN |
| Other antineoplastic agents | Antineoplastic agents | TCLIN |
| Other antineoplastic agents | Antineoplastic agents | TCLIN |
| Other antineoplastic agents | Antineoplastic agents | TCLIN |
| Other antineoplastic agents | Antineoplastic agents | |
| Other antineoplastic agents | Antineoplastic agents | |
| Other antineoplastic agents | Antineoplastic agents | TCLIN |
| Anti-androgens | Endocrine therapy | TCLIN |
| Protein kinase inhibitors | Antineoplastic agents | TCLIN |
| Protein kinase inhibitors | Antineoplastic agents | TCLIN |
| Protein kinase inhibitors | Antineoplastic agents | TCLIN |
| Protein kinase inhibitors | Antineoplastic agents | TCLIN |
| Protein kinase inhibitors | Antineoplastic agents | TCLIN |
| Protein kinase inhibitors | Antineoplastic agents | TCLIN |
| Protein kinase inhibitors | Antineoplastic agents | TCLIN |
| Protein kinase inhibitors | Antineoplastic agents | TCLIN |
| Protein kinase inhibitors | Antineoplastic agents | TCLIN |
| Protein kinase inhibitors | Antineoplastic agents | TCLIN |
| Protein kinase inhibitors | Antineoplastic agents | TCHEM |
| Protein kinase inhibitors | Antineoplastic agents | TCHEM |
| Protein kinase inhibitors | Antineoplastic agents | TCLIN |
| Protein kinase inhibitors | Antineoplastic agents | TCLIN |
| Protein kinase inhibitors | Antineoplastic agents | TCLIN |
| Protein kinase inhibitors | Antineoplastic agents | TCLIN |
| Protein kinase inhibitors | Antineoplastic agents | TCLIN |
| Protein kinase inhibitors | Antineoplastic agents | TCLIN |
| Protein kinase inhibitors | Antineoplastic agents | TCLIN |
| Protein kinase inhibitors | Antineoplastic agents | TCLIN |
| Protein kinase inhibitors | Antineoplastic agents | TCLIN |
| Protein kinase inhibitors | Antineoplastic agents | TCLIN |
| Aromatase inhibitors | Endocrine therapy | TCLIN |
| Anti-androgens | Endocrine therapy | TCLIN |
| Anti-estrogens | Endocrine therapy | TCLIN |
| Protein kinase inhibitors | Antineoplastic agents | TCLIN |

| | | |
|---|---|---|
| Protein kinase inhibitors | Antineoplastic agents | TCLIN |
| Protein kinase inhibitors | Antineoplastic agents | TCLIN |
| Protein kinase inhibitors | Antineoplastic agents | TCLIN |
| Protein kinase inhibitors | Antineoplastic agents | TCLIN |
| Protein kinase inhibitors | Antineoplastic agents | TCHEM |
| Protein kinase inhibitors | Antineoplastic agents | TCLIN |
| Protein kinase inhibitors | Antineoplastic agents | TCLIN |
| Protein kinase inhibitors | Antineoplastic agents | TCLIN |
| Protein kinase inhibitors | Antineoplastic agents | TCLIN |
| Protein kinase inhibitors | Antineoplastic agents | TCLIN |
| Protein kinase inhibitors | Antineoplastic agents | TCLIN |
| Protein kinase inhibitors | Antineoplastic agents | TCLIN |
| Protein kinase inhibitors | Antineoplastic agents | TCLIN |
| Protein kinase inhibitors | Antineoplastic agents | TCLIN |
| Protein kinase inhibitors | Antineoplastic agents | TCLIN |
| Other antineoplastic agents | Antineoplastic agents | TCLIN |
| Other antineoplastic agents | Antineoplastic agents | TCLIN |
| Anthracyclines and related substances | Antineoplastic agents | TCHEM |
| Protein kinase inhibitors | Antineoplastic agents | TCLIN |
| Protein kinase inhibitors | Antineoplastic agents | TCLIN |
| Protein kinase inhibitors | Antineoplastic agents | TCLIN |
| Protein kinase inhibitors | Antineoplastic agents | TCLIN |
| Protein kinase inhibitors | Antineoplastic agents | TCLIN |
| Protein kinase inhibitors | Antineoplastic agents | TCLIN |
| Anti-androgens | Endocrine therapy | TCLIN |
| Protein kinase inhibitors | Antineoplastic agents | TCLIN |
| Protein kinase inhibitors | Antineoplastic agents | TCLIN |
| Protein kinase inhibitors | Antineoplastic agents | TCLIN |
| Protein kinase inhibitors | Antineoplastic agents | TCLIN |
| Protein kinase inhibitors | Antineoplastic agents | TCLIN |
| Protein kinase inhibitors | Antineoplastic agents | TCLIN |
| Protein kinase inhibitors | Antineoplastic agents | TCLIN |
| Protein kinase inhibitors | Antineoplastic agents | TCLIN |
| Protein kinase inhibitors | Antineoplastic agents | TCHEM |
| Protein kinase inhibitors | Antineoplastic agents | TCLIN |
| Protein kinase inhibitors | Antineoplastic agents | TCLIN |
| Protein kinase inhibitors | Antineoplastic agents | TCLIN |
| Protein kinase inhibitors | Antineoplastic agents | TCLIN |
| Protein kinase inhibitors | Antineoplastic agents | TCLIN |
| Protein kinase inhibitors | Antineoplastic agents | TCLIN |
| Protein kinase inhibitors | Antineoplastic agents | TCLIN |
| Protein kinase inhibitors | Antineoplastic agents | TCLIN |
| Protein kinase inhibitors | Antineoplastic agents | TCLIN |
| Protein kinase inhibitors | Antineoplastic agents | TCLIN |

| | | |
|---|---|---|
| Protein kinase inhibitors | Antineoplastic agents | TCLIN |
| Protein kinase inhibitors | Antineoplastic agents | TCHEM |
| Protein kinase inhibitors | Antineoplastic agents | TCHEM |
| Protein kinase inhibitors | Antineoplastic agents | TCLIN |
| Protein kinase inhibitors | Antineoplastic agents | TCHEM |
| Protein kinase inhibitors | Antineoplastic agents | TCLIN |
| Anti-estrogens | Endocrine therapy | TCLIN |
| Anti-estrogens | Endocrine therapy | TCLIN |
| Estrogens | Endocrine therapy | TCLIN |
| Protein kinase inhibitors | Antineoplastic agents | TCLIN |
| Protein kinase inhibitors | Antineoplastic agents | TCLIN |
| Protein kinase inhibitors | Antineoplastic agents | TCLIN |
| Protein kinase inhibitors | Antineoplastic agents | TCLIN |
| Protein kinase inhibitors | Antineoplastic agents | TCLIN |
| Protein kinase inhibitors | Antineoplastic agents | TCLIN |
| Protein kinase inhibitors | Antineoplastic agents | TCLIN |
| Protein kinase inhibitors | Antineoplastic agents | TCLIN |
| Protein kinase inhibitors | Antineoplastic agents | TCLIN |
| Protein kinase inhibitors | Antineoplastic agents | TCLIN |
| Protein kinase inhibitors | Antineoplastic agents | TCLIN |
| Protein kinase inhibitors | Antineoplastic agents | TCHEM |
| Protein kinase inhibitors | Antineoplastic agents | TCHEM |
| Protein kinase inhibitors | Antineoplastic agents | TCLIN |
| Protein kinase inhibitors | Antineoplastic agents | TCLIN |
| Protein kinase inhibitors | Antineoplastic agents | TCLIN |
| Protein kinase inhibitors | Antineoplastic agents | TCLIN |
| Protein kinase inhibitors | Antineoplastic agents | TCLIN |
| Protein kinase inhibitors | Antineoplastic agents | TCLIN |
| Protein kinase inhibitors | Antineoplastic agents | TCLIN |
| Protein kinase inhibitors | Antineoplastic agents | TCLIN |
| Protein kinase inhibitors | Antineoplastic agents | TCLIN |
| Protein kinase inhibitors | Antineoplastic agents | TCLIN |
| Protein kinase inhibitors | Antineoplastic agents | TCLIN |
| Protein kinase inhibitors | Antineoplastic agents | TCLIN |
| Protein kinase inhibitors | Antineoplastic agents | TCLIN |
| Protein kinase inhibitors | Antineoplastic agents | TCLIN |
| Protein kinase inhibitors | Antineoplastic agents | TCLIN |
| Protein kinase inhibitors | Antineoplastic agents | TCLIN |
| Protein kinase inhibitors | Antineoplastic agents | TCLIN |
| Protein kinase inhibitors | Antineoplastic agents | TCLIN |
| Protein kinase inhibitors | Antineoplastic agents | TCLIN |
| Protein kinase inhibitors | Antineoplastic agents | TCLIN |
| Protein kinase inhibitors | Antineoplastic agents | TCLIN |
| Protein kinase inhibitors | Antineoplastic agents | TCLIN |

| | | |
|---|---|---|
| Protein kinase inhibitors | Antineoplastic agents | TCLIN |
| Protein kinase inhibitors | Antineoplastic agents | TCLIN |
| Protein kinase inhibitors | Antineoplastic agents | TCLIN |
| Protein kinase inhibitors | Antineoplastic agents | TCLIN |
| Protein kinase inhibitors | Antineoplastic agents | TCHEM |
| Protein kinase inhibitors | Antineoplastic agents | TCLIN |
| Protein kinase inhibitors | Antineoplastic agents | TCLIN |
| Protein kinase inhibitors | Antineoplastic agents | TCLIN |
| Protein kinase inhibitors | Antineoplastic agents | TCLIN |
| Protein kinase inhibitors | Antineoplastic agents | TCLIN |
| Protein kinase inhibitors | Antineoplastic agents | TCLIN |
| Protein kinase inhibitors | Antineoplastic agents | TCLIN |
| Protein kinase inhibitors | Antineoplastic agents | TCLIN |
| Protein kinase inhibitors | Antineoplastic agents | TCLIN |
| Protein kinase inhibitors | Antineoplastic agents | TCLIN |
| Other antineoplastic agents | Antineoplastic agents | TCLIN |
| Protein kinase inhibitors | Antineoplastic agents | TCLIN |
| Protein kinase inhibitors | Antineoplastic agents | TCLIN |
| Protein kinase inhibitors | Antineoplastic agents | TCLIN |
| Protein kinase inhibitors | Antineoplastic agents | TCLIN |
| Protein kinase inhibitors | Antineoplastic agents | TCLIN |
| Protein kinase inhibitors | Antineoplastic agents | TCLIN |
| Protein kinase inhibitors | Antineoplastic agents | TCLIN |
| Protein kinase inhibitors | Antineoplastic agents | TCLIN |
| Protein kinase inhibitors | Antineoplastic agents | TCLIN |
| Protein kinase inhibitors | Antineoplastic agents | TCLIN |
| Protein kinase inhibitors | Antineoplastic agents | TCLIN |
| Protein kinase inhibitors | Antineoplastic agents | TCHEM |
| Protein kinase inhibitors | Antineoplastic agents | TCHEM |
| Anti-androgens | Endocrine therapy | TCLIN |
| Protein kinase inhibitors | Antineoplastic agents | TCLIN |
| Selective immunosuppressants | Immunosuppressants | TCLIN |
| Selective immunosuppressants | Immunosuppressants | TCLIN |
| Selective immunosuppressants | Immunosuppressants | TCLIN |
| Selective immunosuppressants | Immunosuppressants | TCLIN |
| Selective immunosuppressants | Immunosuppressants | TCLIN |
| Selective immunosuppressants | Immunosuppressants | TCLIN |
| Selective immunosuppressants | Immunosuppressants | TCLIN |
| Selective immunosuppressants | Immunosuppressants | TCLIN |
| Protein kinase inhibitors | Antineoplastic agents | TCLIN |
| Protein kinase inhibitors | Antineoplastic agents | TCLIN |
| Protein kinase inhibitors | Antineoplastic agents | TCLIN |
| Protein kinase inhibitors | Antineoplastic agents | TCLIN |
| Protein kinase inhibitors | Antineoplastic agents | TCLIN |

| | | |
|---|---|---|
| Protein kinase inhibitors | Antineoplastic agents | TCLIN |
| Protein kinase inhibitors | Antineoplastic agents | TCLIN |
| Protein kinase inhibitors | Antineoplastic agents | TCLIN |
| Protein kinase inhibitors | Antineoplastic agents | TCLIN |
| Protein kinase inhibitors | Antineoplastic agents | TCLIN |
| Protein kinase inhibitors | Antineoplastic agents | TCLIN |
| Protein kinase inhibitors | Antineoplastic agents | TCLIN |
| Protein kinase inhibitors | Antineoplastic agents | TCLIN |
| Protein kinase inhibitors | Antineoplastic agents | TCLIN |
| Protein kinase inhibitors | Antineoplastic agents | TCLIN |
| Protein kinase inhibitors | Antineoplastic agents | TCLIN |
| Protein kinase inhibitors | Antineoplastic agents | TCLIN |
| Protein kinase inhibitors | Antineoplastic agents | TCLIN |
| Protein kinase inhibitors | Antineoplastic agents | TCLIN |
| Protein kinase inhibitors | Antineoplastic agents | TCLIN |
| Protein kinase inhibitors | Antineoplastic agents | TCLIN |
| Protein kinase inhibitors | Antineoplastic agents | TCLIN |
| Protein kinase inhibitors | Antineoplastic agents | TCLIN |
| Other antineoplastic agents | Antineoplastic agents | TCHEM |
| Protein kinase inhibitors | Antineoplastic agents | TCLIN |
| Protein kinase inhibitors | Antineoplastic agents | TCLIN |
| Protein kinase inhibitors | Antineoplastic agents | TCLIN |
| Protein kinase inhibitors | Antineoplastic agents | TCLIN |
| Protein kinase inhibitors | Antineoplastic agents | TCLIN |
| Protein kinase inhibitors | Antineoplastic agents | TCLIN |
| Protein kinase inhibitors | Antineoplastic agents | TCLIN |
| Protein kinase inhibitors | Antineoplastic agents | TCLIN |
| Protein kinase inhibitors | Antineoplastic agents | TCLIN |
| Protein kinase inhibitors | Antineoplastic agents | TCLIN |
| Protein kinase inhibitors | Antineoplastic agents | TCLIN |
| Protein kinase inhibitors | Antineoplastic agents | TCLIN |
| Protein kinase inhibitors | Antineoplastic agents | TCHEM |
| Protein kinase inhibitors | Antineoplastic agents | TCLIN |
| Monoclonal antibodies | Antineoplastic agents | TCLIN |
| Protein kinase inhibitors | Antineoplastic agents | TCLIN |
| Protein kinase inhibitors | Antineoplastic agents | TCLIN |
| Protein kinase inhibitors | Antineoplastic agents | TCLIN |
| Protein kinase inhibitors | Antineoplastic agents | TCLIN |
| Protein kinase inhibitors | Antineoplastic agents | TCLIN |
| Protein kinase inhibitors | Antineoplastic agents | TCHEM |
| Protein kinase inhibitors | Antineoplastic agents | TCHEM |
| Protein kinase inhibitors | Antineoplastic agents | TCLIN |
| Protein kinase inhibitors | Antineoplastic agents | TCLIN |
| Protein kinase inhibitors | Antineoplastic agents | TCLIN |

| | | |
|---|---|---|
| Protein kinase inhibitors | Antineoplastic agents | TCLIN |
| Colony stimulating factors | Immunostimulants | TCLIN |
| Colony stimulating factors | Immunostimulants | TCLIN |
| Colony stimulating factors | Immunostimulants | TCLIN |
| Other antineoplastic agents | Antineoplastic agents | TCLIN |
| Other antineoplastic agents | Antineoplastic agents | TCLIN |
| Monoclonal antibodies | Antineoplastic agents | TCLIN |
| Colony stimulating factors | Immunostimulants | TCLIN |
| Monoclonal antibodies | Antineoplastic agents | TCLIN |
| Selective immunosuppressants | Immunosuppressants | TCLIN |
| Selective immunosuppressants | Immunosuppressants | TCLIN |
| Selective immunosuppressants | Immunosuppressants | TCLIN |
| Protein kinase inhibitors | Antineoplastic agents | TCLIN |
| Protein kinase inhibitors | Antineoplastic agents | TCLIN |
| Monoclonal antibodies | Antineoplastic agents | TCLIN |
| Protein kinase inhibitors | Antineoplastic agents | TCHEM |
| Other antineoplastic agents | Antineoplastic agents | TCHEM |
| Pyrimidine analogues | Antineoplastic agents | TCLIN |
| Other antineoplastic agents | Antineoplastic agents | TCLIN |
| Protein kinase inhibitors | Antineoplastic agents | TCLIN |
| Protein kinase inhibitors | Antineoplastic agents | TCHEM |
| Protein kinase inhibitors | Antineoplastic agents | TCLIN |
| Protein kinase inhibitors | Antineoplastic agents | TBIO |
| Protein kinase inhibitors | Antineoplastic agents | TCLIN |
| Protein kinase inhibitors | Antineoplastic agents | TCLIN |
| Protein kinase inhibitors | Antineoplastic agents | TCHEM |
| Protein kinase inhibitors | Antineoplastic agents | TCLIN |
| Protein kinase inhibitors | Antineoplastic agents | TCLIN |
| Pyrimidine analogues | Antineoplastic agents | TCLIN |
| Protein kinase inhibitors | Antineoplastic agents | TCLIN |
| Protein kinase inhibitors | Antineoplastic agents | TCHEM |
| Protein kinase inhibitors | Antineoplastic agents | TCLIN |
| Protein kinase inhibitors | Antineoplastic agents | TCLIN |
| Protein kinase inhibitors | Antineoplastic agents | TCLIN |
| Protein kinase inhibitors | Antineoplastic agents | TCHEM |
| Protein kinase inhibitors | Antineoplastic agents | TCLIN |
| Protein kinase inhibitors | Antineoplastic agents | TCLIN |
| Protein kinase inhibitors | Antineoplastic agents | TCLIN |
| Protein kinase inhibitors | Antineoplastic agents | TCHEM |
| Protein kinase inhibitors | Antineoplastic agents | TCLIN |
| Protein kinase inhibitors | Antineoplastic agents | TCLIN |
| Protein kinase inhibitors | Antineoplastic agents | TCHEM |
| Protein kinase inhibitors | Antineoplastic agents | TCLIN |
| Protein kinase inhibitors | Antineoplastic agents | TCLIN |

| | | |
|---|---|---|
| Protein kinase inhibitors | Antineoplastic agents | TCHEM |
| Protein kinase inhibitors | Antineoplastic agents | TCLIN |
| Protein kinase inhibitors | Antineoplastic agents | TCLIN |
| Protein kinase inhibitors | Antineoplastic agents | TCLIN |
| Protein kinase inhibitors | Antineoplastic agents | TCLIN |
| Protein kinase inhibitors | Antineoplastic agents | TCHEM |
| Protein kinase inhibitors | Antineoplastic agents | TCLIN |
| Protein kinase inhibitors | Antineoplastic agents | TCLIN |
| Protein kinase inhibitors | Antineoplastic agents | TCLIN |
| Protein kinase inhibitors | Antineoplastic agents | TCLIN |
| Protein kinase inhibitors | Antineoplastic agents | TCLIN |
| Protein kinase inhibitors | Antineoplastic agents | TCHEM |
| Protein kinase inhibitors | Antineoplastic agents | TCLIN |
| Protein kinase inhibitors | Antineoplastic agents | TCHEM |
| Protein kinase inhibitors | Antineoplastic agents | TCLIN |
| Protein kinase inhibitors | Antineoplastic agents | TCLIN |
| Protein kinase inhibitors | Antineoplastic agents | TCLIN |
| Protein kinase inhibitors | Antineoplastic agents | TBIO |
| Protein kinase inhibitors | Antineoplastic agents | TCHEM |
| Protein kinase inhibitors | Antineoplastic agents | TCLIN |
| Protein kinase inhibitors | Antineoplastic agents | TCHEM |
| Protein kinase inhibitors | Antineoplastic agents | TCHEM |
| Protein kinase inhibitors | Antineoplastic agents | TCHEM |
| Protein kinase inhibitors | Antineoplastic agents | TCLIN |
| Protein kinase inhibitors | Antineoplastic agents | TCLIN |
| Protein kinase inhibitors | Antineoplastic agents | TCHEM |
| Protein kinase inhibitors | Antineoplastic agents | TCLIN |
| Protein kinase inhibitors | Antineoplastic agents | TCLIN |
| Protein kinase inhibitors | Antineoplastic agents | TCLIN |
| Protein kinase inhibitors | Antineoplastic agents | TCHEM |
| Protein kinase inhibitors | Antineoplastic agents | TBIO |
| Protein kinase inhibitors | Antineoplastic agents | TCLIN |
| Protein kinase inhibitors | Antineoplastic agents | TCLIN |
| Protein kinase inhibitors | Antineoplastic agents | TCLIN |
| Protein kinase inhibitors | Antineoplastic agents | TCLIN |
| Protein kinase inhibitors | Antineoplastic agents | TCHEM |
| Protein kinase inhibitors | Antineoplastic agents | TCLIN |
| Protein kinase inhibitors | Antineoplastic agents | TCLIN |
| Protein kinase inhibitors | Antineoplastic agents | TCLIN |
| Protein kinase inhibitors | Antineoplastic agents | TCHEM |
| Protein kinase inhibitors | Antineoplastic agents | TBIO |
| Protein kinase inhibitors | Antineoplastic agents | TCHEM |
| Protein kinase inhibitors | Antineoplastic agents | TBIO |
| Protein kinase inhibitors | Antineoplastic agents | TCHEM |

| | | |
|---|---|---|
| Protein kinase inhibitors | Antineoplastic agents | TCHEM |
| Protein kinase inhibitors | Antineoplastic agents | TBIO |
| Protein kinase inhibitors | Antineoplastic agents | TCLIN |
| Protein kinase inhibitors | Antineoplastic agents | TCLIN |
| Protein kinase inhibitors | Antineoplastic agents | TCLIN |
| Protein kinase inhibitors | Antineoplastic agents | TCLIN |
| Protein kinase inhibitors | Antineoplastic agents | TCHEM |
| Protein kinase inhibitors | Antineoplastic agents | TCHEM |
| Protein kinase inhibitors | Antineoplastic agents | TBIO |
| Protein kinase inhibitors | Antineoplastic agents | TCLIN |
| Protein kinase inhibitors | Antineoplastic agents | TCLIN |
| Protein kinase inhibitors | Antineoplastic agents | TCLIN |
| Protein kinase inhibitors | Antineoplastic agents | TCHEM |
| Protein kinase inhibitors | Antineoplastic agents | TCLIN |
| Protein kinase inhibitors | Antineoplastic agents | TCHEM |
| Protein kinase inhibitors | Antineoplastic agents | TCLIN |
| Protein kinase inhibitors | Antineoplastic agents | TCLIN |
| Protein kinase inhibitors | Antineoplastic agents | TCHEM |
| Protein kinase inhibitors | Antineoplastic agents | TCLIN |
| Protein kinase inhibitors | Antineoplastic agents | TCHEM |
| Protein kinase inhibitors | Antineoplastic agents | TBIO |
| Protein kinase inhibitors | Antineoplastic agents | TCHEM |
| Protein kinase inhibitors | Antineoplastic agents | TBIO |
| Protein kinase inhibitors | Antineoplastic agents | TCHEM |
| Protein kinase inhibitors | Antineoplastic agents | TCHEM |
| Protein kinase inhibitors | Antineoplastic agents | TBIO |
| Protein kinase inhibitors | Antineoplastic agents | TCHEM |
| Protein kinase inhibitors | Antineoplastic agents | TCLIN |
| Protein kinase inhibitors | Antineoplastic agents | TCLIN |
| Protein kinase inhibitors | Antineoplastic agents | TCLIN |
| Other immunostimulants | Immunostimulants | TCLIN |
| Protein kinase inhibitors | Antineoplastic agents | TCLIN |
| Selective immunosuppressants | Immunosuppressants | TCLIN |
| Selective immunosuppressants | Immunosuppressants | TCLIN |
| Protein kinase inhibitors | Antineoplastic agents | TCLIN |
| Protein kinase inhibitors | Antineoplastic agents | TCLIN |
| Protein kinase inhibitors | Antineoplastic agents | TCLIN |
| Protein kinase inhibitors | Antineoplastic agents | TCLIN |
| Protein kinase inhibitors | Antineoplastic agents | TCLIN |
| Protein kinase inhibitors | Antineoplastic agents | TCLIN |
| Protein kinase inhibitors | Antineoplastic agents | TCLIN |
| Protein kinase inhibitors | Antineoplastic agents | TCLIN |
| Protein kinase inhibitors | Antineoplastic agents | TCLIN |
| Protein kinase inhibitors | Antineoplastic agents | TCLIN |

| | | |
|---|---|---|
| Protein kinase inhibitors | Antineoplastic agents | TCHEM |
| Protein kinase inhibitors | Antineoplastic agents | TCHEM |
| Protein kinase inhibitors | Antineoplastic agents | TCHEM |
| Protein kinase inhibitors | Antineoplastic agents | TCLIN |
| Protein kinase inhibitors | Antineoplastic agents | TCLIN |
| Protein kinase inhibitors | Antineoplastic agents | TCLIN |
| Protein kinase inhibitors | Antineoplastic agents | TBIO |
| Protein kinase inhibitors | Antineoplastic agents | TCHEM |
| Protein kinase inhibitors | Antineoplastic agents | TCHEM |
| Protein kinase inhibitors | Antineoplastic agents | TCHEM |
| Protein kinase inhibitors | Antineoplastic agents | TCHEM |
| Protein kinase inhibitors | Antineoplastic agents | TCLIN |
| Protein kinase inhibitors | Antineoplastic agents | TCLIN |
| Protein kinase inhibitors | Antineoplastic agents | TCLIN |
| Protein kinase inhibitors | Antineoplastic agents | TCLIN |
| Protein kinase inhibitors | Antineoplastic agents | TCLIN |
| Protein kinase inhibitors | Antineoplastic agents | TCLIN |
| Protein kinase inhibitors | Antineoplastic agents | TCLIN |
| Protein kinase inhibitors | Antineoplastic agents | TCHEM |
| Protein kinase inhibitors | Antineoplastic agents | TCHEM |
| Protein kinase inhibitors | Antineoplastic agents | TCHEM |
| Protein kinase inhibitors | Antineoplastic agents | TCLIN |
| Anthracyclines and related substances | Antineoplastic agents | TCLIN |
| Anthracyclines and related substances | Antineoplastic agents | TCHEM |
| Protein kinase inhibitors | Antineoplastic agents | TCHEM |
| Protein kinase inhibitors | Antineoplastic agents | TCLIN |
| Protein kinase inhibitors | Antineoplastic agents | TCLIN |
| Protein kinase inhibitors | Antineoplastic agents | TCHEM |
| Protein kinase inhibitors | Antineoplastic agents | TCLIN |
| Protein kinase inhibitors | Antineoplastic agents | TCHEM |
| Protein kinase inhibitors | Antineoplastic agents | TCLIN |
| Protein kinase inhibitors | Antineoplastic agents | TCHEM |
| Protein kinase inhibitors | Antineoplastic agents | TCLIN |
| Protein kinase inhibitors | Antineoplastic agents | TCHEM |
| Protein kinase inhibitors | Antineoplastic agents | TCLIN |
| Protein kinase inhibitors | Antineoplastic agents | TCLIN |
| Protein kinase inhibitors | Antineoplastic agents | TCLIN |
| Protein kinase inhibitors | Antineoplastic agents | TCHEM |
| Protein kinase inhibitors | Antineoplastic agents | TCHEM |
| Protein kinase inhibitors | Antineoplastic agents | TCLIN |
| Protein kinase inhibitors | Antineoplastic agents | TCLIN |
| Anti-estrogens | Endocrine therapy | TCLIN |
| Protein kinase inhibitors | Antineoplastic agents | TCHEM |
| Protein kinase inhibitors | Antineoplastic agents | TCHEM |

| | | |
|---|---|---|
| Protein kinase inhibitors | Antineoplastic agents | TCLIN |
| Protein kinase inhibitors | Antineoplastic agents | TCLIN |
| Protein kinase inhibitors | Antineoplastic agents | TCLIN |
| Protein kinase inhibitors | Antineoplastic agents | TCHEM |
| Protein kinase inhibitors | Antineoplastic agents | TCHEM |
| Protein kinase inhibitors | Antineoplastic agents | TCHEM |
| Protein kinase inhibitors | Antineoplastic agents | TCLIN |
| Protein kinase inhibitors | Antineoplastic agents | TCLIN |
| Protein kinase inhibitors | Antineoplastic agents | TCLIN |
| Protein kinase inhibitors | Antineoplastic agents | TCHEM |
| Protein kinase inhibitors | Antineoplastic agents | TCHEM |
| Protein kinase inhibitors | Antineoplastic agents | TCHEM |
| Protein kinase inhibitors | Antineoplastic agents | TCLIN |
| Protein kinase inhibitors | Antineoplastic agents | TCHEM |
| Protein kinase inhibitors | Antineoplastic agents | TCLIN |
| Protein kinase inhibitors | Antineoplastic agents | TCHEM |
| Protein kinase inhibitors | Antineoplastic agents | TCLIN |
| Protein kinase inhibitors | Antineoplastic agents | TCLIN |
| Protein kinase inhibitors | Antineoplastic agents | TCLIN |
| Protein kinase inhibitors | Antineoplastic agents | TCLIN |
| Protein kinase inhibitors | Antineoplastic agents | TCHEM |
| Protein kinase inhibitors | Antineoplastic agents | TCLIN |
| Platinum compounds | Antineoplastic agents | TCLIN |
| Other immunostimulants | Immunostimulants | TCHEM |
| Selective immunosuppressants | Immunosuppressants | TCLIN |
| Protein kinase inhibitors | Antineoplastic agents | TCLIN |
| Protein kinase inhibitors | Antineoplastic agents | TCLIN |
| Protein kinase inhibitors | Antineoplastic agents | TCLIN |
| Protein kinase inhibitors | Antineoplastic agents | TBIO |
| Protein kinase inhibitors | Antineoplastic agents | TCLIN |
| Protein kinase inhibitors | Antineoplastic agents | TCLIN |
| Protein kinase inhibitors | Antineoplastic agents | TCLIN |
| Protein kinase inhibitors | Antineoplastic agents | TCHEM |
| Protein kinase inhibitors | Antineoplastic agents | TCHEM |
| Protein kinase inhibitors | Antineoplastic agents | TCLIN |
| Protein kinase inhibitors | Antineoplastic agents | TCLIN |
| Protein kinase inhibitors | Antineoplastic agents | TCHEM |
| Protein kinase inhibitors | Antineoplastic agents | TCHEM |
| Protein kinase inhibitors | Antineoplastic agents | TCHEM |
| Anthracyclines and related substances | Antineoplastic agents | TCHEM |
| Protein kinase inhibitors | Antineoplastic agents | TCHEM |
| Protein kinase inhibitors | Antineoplastic agents | TCHEM |
| Protein kinase inhibitors | Antineoplastic agents | TCLIN |
| Protein kinase inhibitors | Antineoplastic agents | TCHEM |

| | | |
|---|---|---|
| Protein kinase inhibitors | Antineoplastic agents | TCHEM |
| Protein kinase inhibitors | Antineoplastic agents | TCHEM |
| Protein kinase inhibitors | Antineoplastic agents | TCLIN |
| Protein kinase inhibitors | Antineoplastic agents | TCHEM |
| Protein kinase inhibitors | Antineoplastic agents | TCHEM |
| Protein kinase inhibitors | Antineoplastic agents | TCHEM |
| Protein kinase inhibitors | Antineoplastic agents | TCHEM |
| Protein kinase inhibitors | Antineoplastic agents | TCHEM |
| Protein kinase inhibitors | Antineoplastic agents | TCLIN |
| Protein kinase inhibitors | Antineoplastic agents | TCLIN |
| Protein kinase inhibitors | Antineoplastic agents | TCHEM |
| Protein kinase inhibitors | Antineoplastic agents | TCHEM |
| Protein kinase inhibitors | Antineoplastic agents | TCLIN |
| Protein kinase inhibitors | Antineoplastic agents | TCHEM |
| Protein kinase inhibitors | Antineoplastic agents | TCHEM |
| Protein kinase inhibitors | Antineoplastic agents | TCHEM |
| Protein kinase inhibitors | Antineoplastic agents | TCHEM |
| Protein kinase inhibitors | Antineoplastic agents | TCHEM |
| Protein kinase inhibitors | Antineoplastic agents | TCLIN |
| Protein kinase inhibitors | Antineoplastic agents | TCLIN |
| Protein kinase inhibitors | Antineoplastic agents | TCHEM |
| Protein kinase inhibitors | Antineoplastic agents | TCLIN |
| Protein kinase inhibitors | Antineoplastic agents | TCHEM |
| Protein kinase inhibitors | Antineoplastic agents | TCHEM |
| Protein kinase inhibitors | Antineoplastic agents | TCLIN |
| Protein kinase inhibitors | Antineoplastic agents | TCHEM |
| Protein kinase inhibitors | Antineoplastic agents | TCHEM |
| Protein kinase inhibitors | Antineoplastic agents | TCLIN |
| Other antineoplastic agents | Antineoplastic agents | TCLIN |
| Other antineoplastic agents | Antineoplastic agents | TCLIN |
| Monoclonal antibodies | Antineoplastic agents | TCLIN |
| Monoclonal antibodies | Antineoplastic agents | TCLIN |
| Monoclonal antibodies | Antineoplastic agents | TCLIN |
| Other antineoplastic agents | Antineoplastic agents | TCHEM |
| Other antineoplastic agents | Antineoplastic agents | TCLIN |
| Other antineoplastic agents | Antineoplastic agents | TCLIN |
| Pyrimidine analogues | Antineoplastic agents | TCLIN |
| Nitrogen mustard analogues | Antineoplastic agents | TCLIN |
| Nitrogen mustard analogues | Antineoplastic agents | TCLIN |
| Pyrimidine analogues | Antineoplastic agents | TCLIN |
| Purine analogues | Antineoplastic agents | TCLIN |
| Protein kinase inhibitors | Antineoplastic agents | TCHEM |
| Protein kinase inhibitors | Antineoplastic agents | TCHEM |
| Protein kinase inhibitors | Antineoplastic agents | TBIO |

| | | |
|---|---|---|
| Protein kinase inhibitors | Antineoplastic agents | TCLIN |
| Protein kinase inhibitors | Antineoplastic agents | TCLIN |
| Protein kinase inhibitors | Antineoplastic agents | TCHEM |
| Protein kinase inhibitors | Antineoplastic agents | TCHEM |
| Protein kinase inhibitors | Antineoplastic agents | TCHEM |
| Protein kinase inhibitors | Antineoplastic agents | TCHEM |
| Protein kinase inhibitors | Antineoplastic agents | TCLIN |
| Pyrimidine analogues | Antineoplastic agents | TCLIN |
| Anti-estrogens | Endocrine therapy | TCLIN |
| Protein kinase inhibitors | Antineoplastic agents | TCHEM |
| Pyrimidine analogues | Antineoplastic agents | TCLIN |
| Pyrimidine analogues | Antineoplastic agents | TCLIN |
| Protein kinase inhibitors | Antineoplastic agents | TCHEM |
| Protein kinase inhibitors | Antineoplastic agents | TCHEM |
| Protein kinase inhibitors | Antineoplastic agents | TCLIN |
| Other immunosuppressants | Immunosuppressants | TCLIN |
| Other antineoplastic agents | Antineoplastic agents | TCHEM |
| Other cytotoxic antibiotics | Antineoplastic agents | TCHEM |
| Protein kinase inhibitors | Antineoplastic agents | TCHEM |
| Protein kinase inhibitors | Antineoplastic agents | TCHEM |
| Folic acid analogues | Antineoplastic agents | TCLIN |
| Folic acid analogues | Antineoplastic agents | TCLIN |
| Protein kinase inhibitors | Antineoplastic agents | TCHEM |
| Protein kinase inhibitors | Antineoplastic agents | TCHEM |
| Protein kinase inhibitors | Antineoplastic agents | TCHEM |
| Protein kinase inhibitors | Antineoplastic agents | TCHEM |
| Protein kinase inhibitors | Antineoplastic agents | TCHEM |
| Protein kinase inhibitors | Antineoplastic agents | TCHEM |
| Protein kinase inhibitors | Antineoplastic agents | TCHEM |
| Protein kinase inhibitors | Antineoplastic agents | TCHEM |
| Protein kinase inhibitors | Antineoplastic agents | TCLIN |
| Protein kinase inhibitors | Antineoplastic agents | TBIO |
| Protein kinase inhibitors | Antineoplastic agents | TCLIN |
| Protein kinase inhibitors | Antineoplastic agents | TCHEM |
| Protein kinase inhibitors | Antineoplastic agents | TCLIN |
| Protein kinase inhibitors | Antineoplastic agents | TCHEM |
| Protein kinase inhibitors | Antineoplastic agents | TCHEM |
| Protein kinase inhibitors | Antineoplastic agents | TCHEM |
| Protein kinase inhibitors | Antineoplastic agents | TCHEM |
| Protein kinase inhibitors | Antineoplastic agents | TCHEM |
| Anti-estrogens | Endocrine therapy | TCLIN |
| Anti-estrogens | Endocrine therapy | TCLIN |
| Other immunosuppressants | Immunosuppressants | TCLIN |
| Other immunosuppressants | Immunosuppressants | TCLIN |

| | | |
|---|---|---|
| Other immunostimulants | Immunostimulants | TCLIN |
| Other immunosuppressants | Immunosuppressants | TCLIN |
| Protein kinase inhibitors | Antineoplastic agents | TCHEM |
| Protein kinase inhibitors | Antineoplastic agents | TCHEM |
| Protein kinase inhibitors | Antineoplastic agents | TBIO |
| Protein kinase inhibitors | Antineoplastic agents | TCLIN |
| Protein kinase inhibitors | Antineoplastic agents | TCHEM |
| Other antineoplastic agents | Antineoplastic agents | TCLIN |
| Other antineoplastic agents | Antineoplastic agents | TCLIN |
| Other antineoplastic agents | Antineoplastic agents | TCLIN |
| Other antineoplastic agents | Antineoplastic agents | TCLIN |
| Other antineoplastic agents | Antineoplastic agents | TCLIN |
| Other antineoplastic agents | Antineoplastic agents | TCLIN |
| Selective immunosuppressants | Immunosuppressants | TCLIN |
| Protein kinase inhibitors | Antineoplastic agents | TBIO |
| Protein kinase inhibitors | Antineoplastic agents | TCHEM |
| Protein kinase inhibitors | Antineoplastic agents | TCHEM |
| Protein kinase inhibitors | Antineoplastic agents | TCHEM |
| Protein kinase inhibitors | Antineoplastic agents | TCLIN |
| Protein kinase inhibitors | Antineoplastic agents | TBIO |
| Protein kinase inhibitors | Antineoplastic agents | TCLIN |
| Protein kinase inhibitors | Antineoplastic agents | TCHEM |
| Protein kinase inhibitors | Antineoplastic agents | TCHEM |
| Protein kinase inhibitors | Antineoplastic agents | TCHEM |
| Protein kinase inhibitors | Antineoplastic agents | TCHEM |
| Protein kinase inhibitors | Antineoplastic agents | TCHEM |
| Protein kinase inhibitors | Antineoplastic agents | TCHEM |
| Protein kinase inhibitors | Antineoplastic agents | TCHEM |
| Protein kinase inhibitors | Antineoplastic agents | TCLIN |
| Protein kinase inhibitors | Antineoplastic agents | TBIO |
| Protein kinase inhibitors | Antineoplastic agents | TCLIN |
| Protein kinase inhibitors | Antineoplastic agents | TCHEM |
| Protein kinase inhibitors | Antineoplastic agents | TCHEM |
| Protein kinase inhibitors | Antineoplastic agents | TCHEM |
| Protein kinase inhibitors | Antineoplastic agents | TBIO |
| Protein kinase inhibitors | Antineoplastic agents | TCLIN |
| Protein kinase inhibitors | Antineoplastic agents | TCHEM |
| Protein kinase inhibitors | Antineoplastic agents | TCHEM |
| Protein kinase inhibitors | Antineoplastic agents | TCHEM |
| Protein kinase inhibitors | Antineoplastic agents | TCHEM |
| Protein kinase inhibitors | Antineoplastic agents | TCLIN |
| Protein kinase inhibitors | Antineoplastic agents | TCHEM |
| Protein kinase inhibitors | Antineoplastic agents | TCLIN |
| Protein kinase inhibitors | Antineoplastic agents | TCHEM |

| | | |
|---|---|---|
| Protein kinase inhibitors | Antineoplastic agents | TCHEM |
| Protein kinase inhibitors | Antineoplastic agents | TCHEM |
| Protein kinase inhibitors | Antineoplastic agents | TCHEM |
| Platinum compounds | Antineoplastic agents | TCHEM |
| Other immunostimulants | Immunostimulants | TBIO |
| Other immunostimulants | Immunostimulants | TCHEM |
| Other antineoplastic agents | Antineoplastic agents | TCLIN |
| Other antineoplastic agents | Antineoplastic agents | TCLIN |
| Other antineoplastic agents | Antineoplastic agents | TCLIN |
| Other antineoplastic agents | Antineoplastic agents | TCLIN |
| Selective immunosuppressants | Immunosuppressants | TCLIN |
| Selective immunosuppressants | Immunosuppressants | TCLIN |
| Other antineoplastic agents | Antineoplastic agents | TCLIN |
| Other antineoplastic agents | Antineoplastic agents | TCLIN |
| Other antineoplastic agents | Antineoplastic agents | TCLIN |
| Other antineoplastic agents | Antineoplastic agents | TCLIN |
| Other antineoplastic agents | Antineoplastic agents | TCLIN |
| Protein kinase inhibitors | Antineoplastic agents | TCHEM |
| Protein kinase inhibitors | Antineoplastic agents | TCHEM |
| Protein kinase inhibitors | Antineoplastic agents | TCHEM |
| Protein kinase inhibitors | Antineoplastic agents | TCHEM |
| Protein kinase inhibitors | Antineoplastic agents | TCHEM |
| Protein kinase inhibitors | Antineoplastic agents | TCLIN |
| Protein kinase inhibitors | Antineoplastic agents | TCHEM |
| Protein kinase inhibitors | Antineoplastic agents | TCHEM |
| Protein kinase inhibitors | Antineoplastic agents | TCHEM |
| Protein kinase inhibitors | Antineoplastic agents | TCHEM |
| Protein kinase inhibitors | Antineoplastic agents | TCHEM |
| Protein kinase inhibitors | Antineoplastic agents | TCHEM |
| Tumor necrosis factor alpha (tnf-α) inhibitors | Immunosuppressants | TCLIN |
| Other antineoplastic agents | Antineoplastic agents | TCHEM |
| Monoclonal antibodies | Antineoplastic agents | TCLIN |
| Interleukin inhibitors | Immunosuppressants | TCLIN |
| Interleukin inhibitors | Immunosuppressants | TCLIN |
| Protein kinase inhibitors | Antineoplastic agents | TCHEM |
| Interleukin inhibitors | Immunosuppressants | TCLIN |
| Interleukin inhibitors | Immunosuppressants | TCLIN |
| Tumor necrosis factor alpha (tnf-α) inhibitors | Immunosuppressants | TCLIN |
| Tumor necrosis factor alpha (tnf-α) inhibitors | Immunosuppressants | TCLIN |
| Tumor necrosis factor alpha (tnf-α) inhibitors | Immunosuppressants | TCLIN |
| Interleukin inhibitors | Immunosuppressants | TCLIN |
| Tumor necrosis factor alpha (tnf-α) inhibitors | Immunosuppressants | TCLIN |
| Interleukins | Immunostimulants | TCLIN |
| Interleukins | Immunostimulants | TCLIN |

| | | |
|---|---|---|
| Other antineoplastic agents | Antineoplastic agents | TCLIN |
| Interleukin inhibitors | Immunosuppressants | TCLIN |
| Selective immunosuppressants | Immunosuppressants | TCLIN |
| Other antineoplastic agents | Antineoplastic agents | TCHEM |
| Other antineoplastic agents | Antineoplastic agents | TCHEM |
| Protein kinase inhibitors | Antineoplastic agents | TCLIN |
| Protein kinase inhibitors | Antineoplastic agents | TCLIN |
| Protein kinase inhibitors | Antineoplastic agents | TCHEM |
| Protein kinase inhibitors | Antineoplastic agents | TCLIN |
| Protein kinase inhibitors | Antineoplastic agents | TCHEM |
| Protein kinase inhibitors | Antineoplastic agents | TCHEM |
| Protein kinase inhibitors | Antineoplastic agents | TCLIN |
| Protein kinase inhibitors | Antineoplastic agents | TCHEM |
| Protein kinase inhibitors | Antineoplastic agents | TCLIN |
| Protein kinase inhibitors | Antineoplastic agents | TCLIN |
| Protein kinase inhibitors | Antineoplastic agents | TCHEM |
| Protein kinase inhibitors | Antineoplastic agents | TCHEM |
| Protein kinase inhibitors | Antineoplastic agents | TCHEM |
| Protein kinase inhibitors | Antineoplastic agents | TCHEM |
| Protein kinase inhibitors | Antineoplastic agents | TCHEM |
| Protein kinase inhibitors | Antineoplastic agents | TCHEM |
| Protein kinase inhibitors | Antineoplastic agents | TCHEM |
| Protein kinase inhibitors | Antineoplastic agents | TCHEM |
| Protein kinase inhibitors | Antineoplastic agents | TCHEM |
| Protein kinase inhibitors | Antineoplastic agents | TCLIN |
| Protein kinase inhibitors | Antineoplastic agents | TCHEM |
| Protein kinase inhibitors | Antineoplastic agents | TCHEM |
| Protein kinase inhibitors | Antineoplastic agents | TCHEM |
| Protein kinase inhibitors | Antineoplastic agents | TCHEM |
| Protein kinase inhibitors | Antineoplastic agents | TCHEM |
| Protein kinase inhibitors | Antineoplastic agents | TCHEM |
| Protein kinase inhibitors | Antineoplastic agents | TCHEM |
| Protein kinase inhibitors | Antineoplastic agents | TCHEM |
| Protein kinase inhibitors | Antineoplastic agents | TCLIN |
| Protein kinase inhibitors | Antineoplastic agents | TCLIN |
| Protein kinase inhibitors | Antineoplastic agents | TCHEM |
| Protein kinase inhibitors | Antineoplastic agents | TBIO |
| Protein kinase inhibitors | Antineoplastic agents | TCHEM |
| Anthracyclines and related substances | Antineoplastic agents | TCLIN |
| Protein kinase inhibitors | Antineoplastic agents | TCLIN |
| Protein kinase inhibitors | Antineoplastic agents | TCHEM |
| Protein kinase inhibitors | Antineoplastic agents | TCHEM |
| Protein kinase inhibitors | Antineoplastic agents | TCHEM |
| Protein kinase inhibitors | Antineoplastic agents | TCLIN |

| | | |
|---|---|---|
| Protein kinase inhibitors | Antineoplastic agents | TCHEM |
| Protein kinase inhibitors | Antineoplastic agents | TCHEM |
| Protein kinase inhibitors | Antineoplastic agents | TCLIN |
| Protein kinase inhibitors | Antineoplastic agents | TCLIN |
| Protein kinase inhibitors | Antineoplastic agents | TCHEM |
| Protein kinase inhibitors | Antineoplastic agents | TCHEM |
| Protein kinase inhibitors | Antineoplastic agents | TCHEM |
| Protein kinase inhibitors | Antineoplastic agents | TCHEM |
| Protein kinase inhibitors | Antineoplastic agents | TCLIN |
| Protein kinase inhibitors | Antineoplastic agents | TCHEM |
| Protein kinase inhibitors | Antineoplastic agents | TCHEM |
| Protein kinase inhibitors | Antineoplastic agents | TCHEM |
| Protein kinase inhibitors | Antineoplastic agents | TCLIN |
| Protein kinase inhibitors | Antineoplastic agents | TCLIN |
| Protein kinase inhibitors | Antineoplastic agents | TCHEM |
| Protein kinase inhibitors | Antineoplastic agents | TCHEM |
| Protein kinase inhibitors | Antineoplastic agents | TCHEM |
| Protein kinase inhibitors | Antineoplastic agents | TCHEM |
| Protein kinase inhibitors | Antineoplastic agents | TCLIN |
| Protein kinase inhibitors | Antineoplastic agents | TCHEM |
| Protein kinase inhibitors | Antineoplastic agents | TCHEM |
| Protein kinase inhibitors | Antineoplastic agents | TCHEM |
| Protein kinase inhibitors | Antineoplastic agents | TCHEM |
| Protein kinase inhibitors | Antineoplastic agents | TCHEM |
| Protein kinase inhibitors | Antineoplastic agents | TCHEM |
| Protein kinase inhibitors | Antineoplastic agents | TCHEM |
| Protein kinase inhibitors | Antineoplastic agents | TCLIN |
| Protein kinase inhibitors | Antineoplastic agents | TCLIN |
| Protein kinase inhibitors | Antineoplastic agents | TCLIN |
| Protein kinase inhibitors | Antineoplastic agents | TCLIN |
| Protein kinase inhibitors | Antineoplastic agents | TCHEM |
| Protein kinase inhibitors | Antineoplastic agents | TCHEM |
| Protein kinase inhibitors | Antineoplastic agents | TCHEM |
| Protein kinase inhibitors | Antineoplastic agents | TCLIN |
| Protein kinase inhibitors | Antineoplastic agents | TCLIN |
| Protein kinase inhibitors | Antineoplastic agents | TCHEM |
| Protein kinase inhibitors | Antineoplastic agents | TCLIN |
| Protein kinase inhibitors | Antineoplastic agents | TCHEM |
| Protein kinase inhibitors | Antineoplastic agents | TCHEM |
| Protein kinase inhibitors | Antineoplastic agents | TCHEM |
| Protein kinase inhibitors | Antineoplastic agents | TCHEM |
| Protein kinase inhibitors | Antineoplastic agents | TCLIN |
| Protein kinase inhibitors | Antineoplastic agents | TCHEM |
| Protein kinase inhibitors | Antineoplastic agents | TCHEM |

| | | |
|---|---|---|
| Protein kinase inhibitors | Antineoplastic agents | TCHEM |
| Protein kinase inhibitors | Antineoplastic agents | TCHEM |
| Protein kinase inhibitors | Antineoplastic agents | TCHEM |
| Protein kinase inhibitors | Antineoplastic agents | TCLIN |
| Protein kinase inhibitors | Antineoplastic agents | TCLIN |
| Protein kinase inhibitors | Antineoplastic agents | TCHEM |
| Protein kinase inhibitors | Antineoplastic agents | TCLIN |
| Protein kinase inhibitors | Antineoplastic agents | TCHEM |
| Protein kinase inhibitors | Antineoplastic agents | TCHEM |
| Protein kinase inhibitors | Antineoplastic agents | TCHEM |
| Protein kinase inhibitors | Antineoplastic agents | TCHEM |
| Protein kinase inhibitors | Antineoplastic agents | TCLIN |
| Protein kinase inhibitors | Antineoplastic agents | TCHEM |
| Protein kinase inhibitors | Antineoplastic agents | TCHEM |
| Protein kinase inhibitors | Antineoplastic agents | TCHEM |
| Protein kinase inhibitors | Antineoplastic agents | TCHEM |
| Protein kinase inhibitors | Antineoplastic agents | TCHEM |
| Protein kinase inhibitors | Antineoplastic agents | TCHEM |
| Protein kinase inhibitors | Antineoplastic agents | TCHEM |
| Protein kinase inhibitors | Antineoplastic agents | TCHEM |
| Protein kinase inhibitors | Antineoplastic agents | TCLIN |
| Protein kinase inhibitors | Antineoplastic agents | TCLIN |
| Protein kinase inhibitors | Antineoplastic agents | TCHEM |
| Protein kinase inhibitors | Antineoplastic agents | TCHEM |
| Protein kinase inhibitors | Antineoplastic agents | TCHEM |
| Protein kinase inhibitors | Antineoplastic agents | TCLIN |
| Protein kinase inhibitors | Antineoplastic agents | TCHEM |
| Protein kinase inhibitors | Antineoplastic agents | TCHEM |
| Protein kinase inhibitors | Antineoplastic agents | TCLIN |
| Protein kinase inhibitors | Antineoplastic agents | TCHEM |
| Protein kinase inhibitors | Antineoplastic agents | TCLIN |
| Protein kinase inhibitors | Antineoplastic agents | TCHEM |
| Protein kinase inhibitors | Antineoplastic agents | TCHEM |
| Protein kinase inhibitors | Antineoplastic agents | TCHEM |
| Protein kinase inhibitors | Antineoplastic agents | TCHEM |
| Protein kinase inhibitors | Antineoplastic agents | TCLIN |
| Protein kinase inhibitors | Antineoplastic agents | TCHEM |
| Protein kinase inhibitors | Antineoplastic agents | TCHEM |
| Protein kinase inhibitors | Antineoplastic agents | TCHEM |
| Protein kinase inhibitors | Antineoplastic agents | TCHEM |
| Protein kinase inhibitors | Antineoplastic agents | TCHEM |
| Protein kinase inhibitors | Antineoplastic agents | TCHEM |
| Other plant alkaloids and natural products | Antineoplastic agents | TCLIN |
| Protein kinase inhibitors | Antineoplastic agents | TCHEM |

| | | |
|---|---|---|
| Protein kinase inhibitors | Antineoplastic agents | TCLIN |
| Protein kinase inhibitors | Antineoplastic agents | TCHEM |
| Other immunosuppressants | Immunosuppressants | TCLIN |
| Protein kinase inhibitors | Antineoplastic agents | |
| Protein kinase inhibitors | Antineoplastic agents | TCLIN |
| Protein kinase inhibitors | Antineoplastic agents | TCLIN |
| Protein kinase inhibitors | Antineoplastic agents | TCLIN |
| Protein kinase inhibitors | Antineoplastic agents | TCLIN |
| Other antineoplastic agents | Antineoplastic agents | TCHEM |
| Protein kinase inhibitors | Antineoplastic agents | TCLIN |
| Protein kinase inhibitors | Antineoplastic agents | TCLIN |
| Protein kinase inhibitors | Antineoplastic agents | TCHEM |
| Protein kinase inhibitors | Antineoplastic agents | TCLIN |
| Protein kinase inhibitors | Antineoplastic agents | TCHEM |
| Protein kinase inhibitors | Antineoplastic agents | TCHEM |
| Protein kinase inhibitors | Antineoplastic agents | TCHEM |
| Protein kinase inhibitors | Antineoplastic agents | TCLIN |
| Protein kinase inhibitors | Antineoplastic agents | TCHEM |
| Protein kinase inhibitors | Antineoplastic agents | TCHEM |
| Protein kinase inhibitors | Antineoplastic agents | TCHEM |
| Protein kinase inhibitors | Antineoplastic agents | TCHEM |
| Protein kinase inhibitors | Antineoplastic agents | TCHEM |
| Protein kinase inhibitors | Antineoplastic agents | TCHEM |
| Other antineoplastic agents | Antineoplastic agents | TCLIN |
| Protein kinase inhibitors | Antineoplastic agents | TCLIN |
| Other antineoplastic agents | Antineoplastic agents | TCHEM |
| Other antineoplastic agents | Antineoplastic agents | TCLIN |

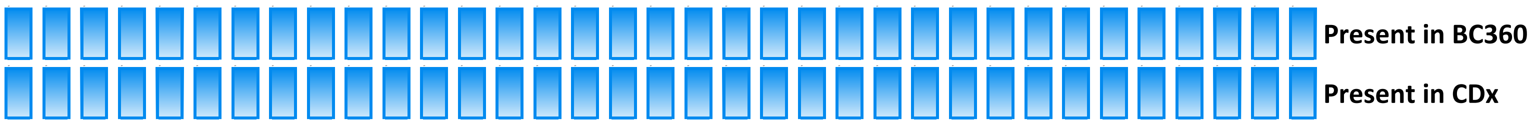
Present in BC360
Present in CDx

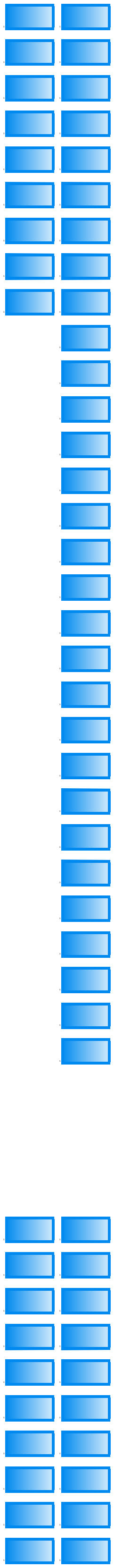

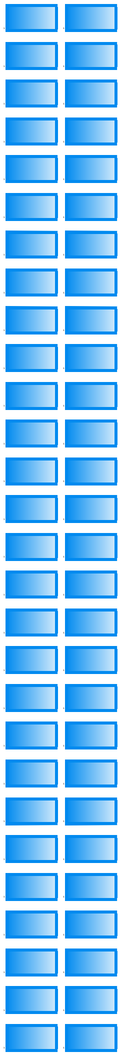

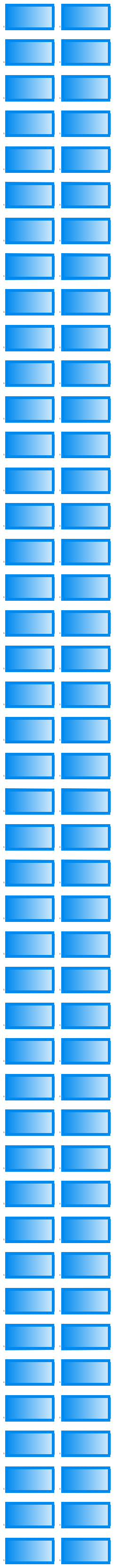

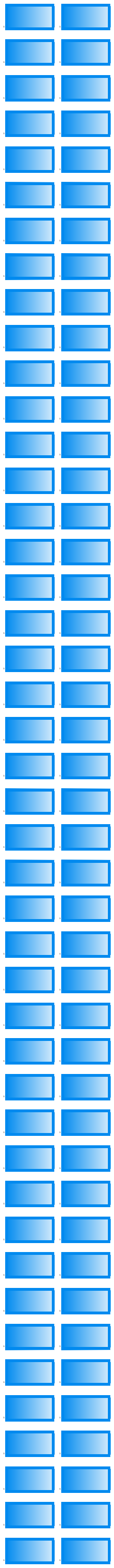

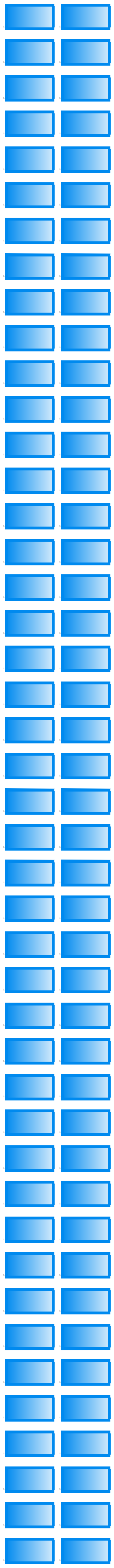

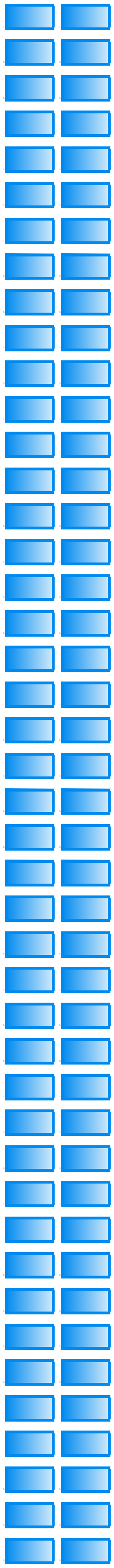

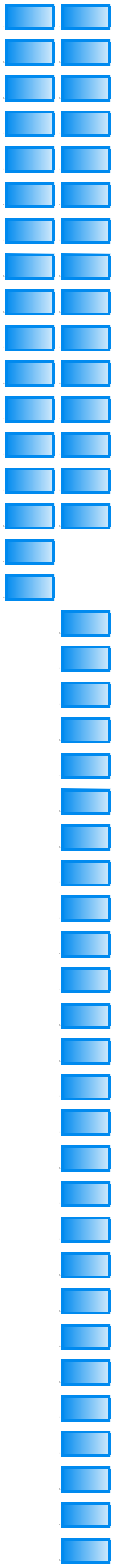

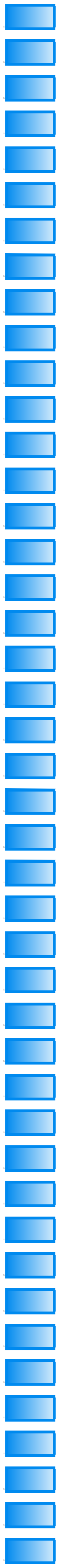

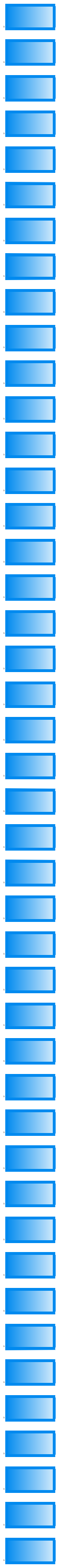

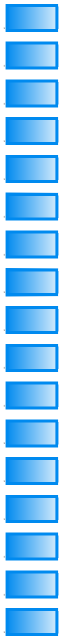

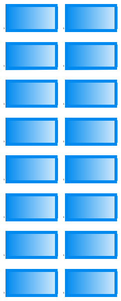

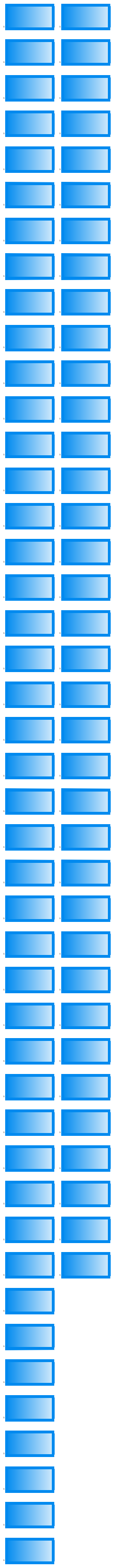

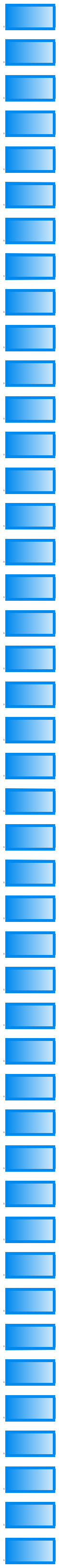

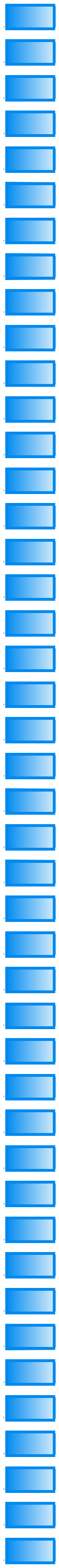

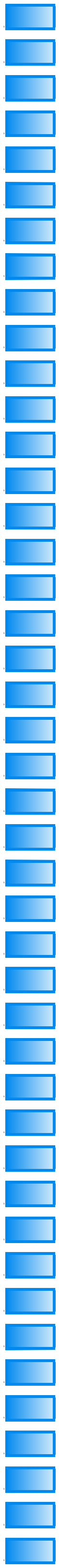

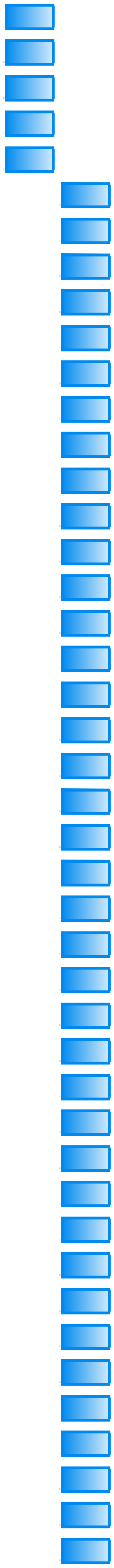

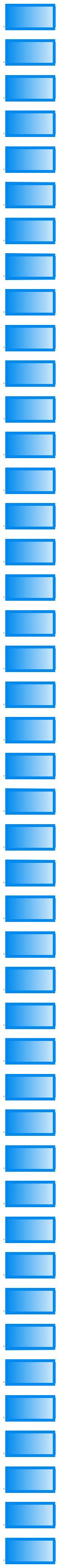

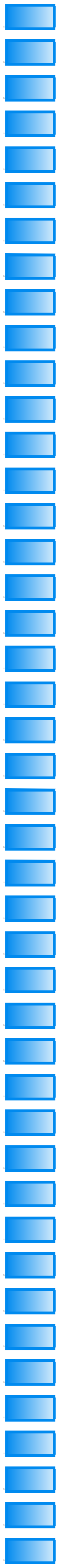

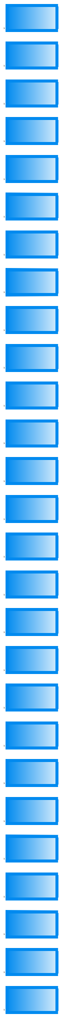

Supplemental material 10

### Cluster 1

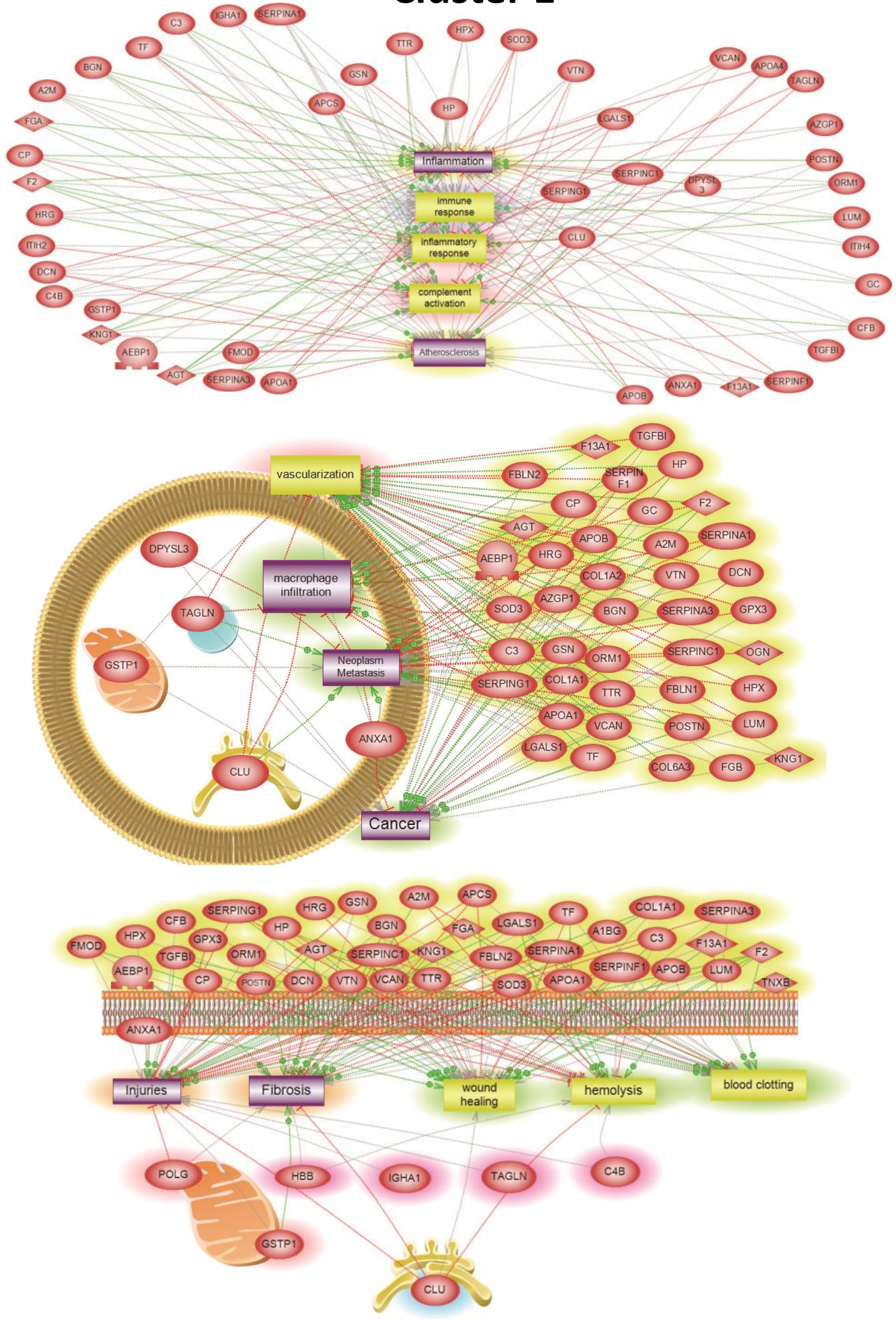

### Cluster 2

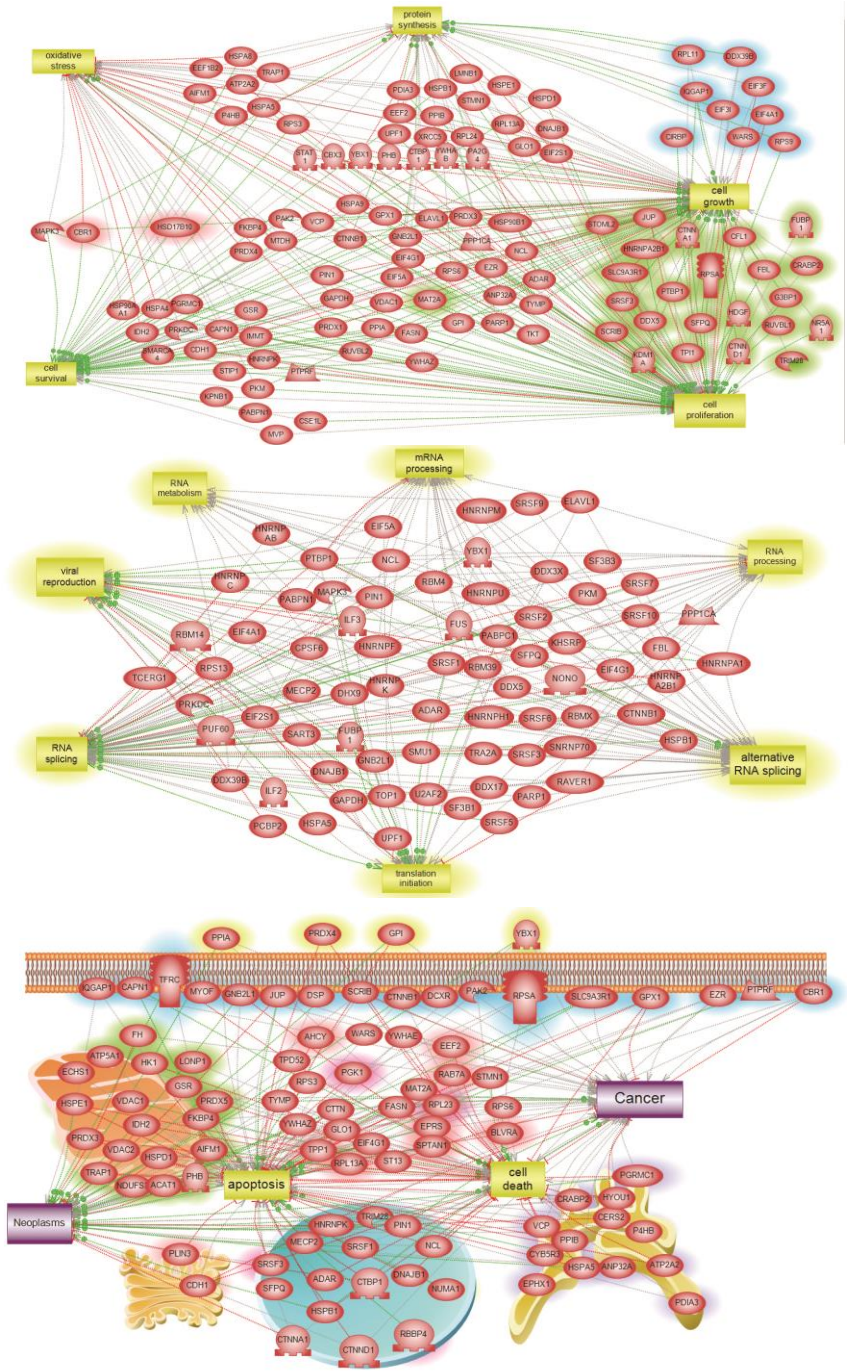
